# TRAINING WITH A NON-INVASIVE BRAIN-MACHINE INTERFACE, COMBINED WITH VIRTUAL REALITY AND ASSISTED ROBOTIC LOCOMOTION, INDUCES SIGNIFICANT MOTOR RECOVERY AND PARTIAL REVERSAL OF WIDESPREAD CORTICAL ATROPHY IN ASIA A PARAPLEGICS: A RANDOMIZED, CONTROLLED, SINGLE-CENTER TRIAL

**DOI:** 10.64898/2025.11.28.25340891

**Authors:** Guo-Guang Zhao, Eduardo J. L. Alho, Peng-Hu Wei, Yi Tang, Lin Liu, Lin Zhu, Yan-Feng Yang, Yuan-Yuan Zhang, Yi Shan, Jing Li, Chen-Xi Sun, Gui-Xiang Shan, Ping Wu, Jie Lu, Biao Chen, Wei-Qun Song, Hang Wu, Adriana R. J. Ferreira, Miguel A.L. Nicolelis

## Abstract

Twelve years ago, the Walk Again Project (WAP) introduced a multi-stage neurorehabilitation protocol, the Walk Again Neurorehabilitation Protocol (WANR), which combines virtual reality training with robotic gait systems, both controlled by an EEG-based, non-invasive brain-machine interface (BMI). Training with the WANR led to significant partial neurological and functional recovery in spinal cord injury (SCI) patients. Yet, the neural mechanisms underlying such a recovery remain unknown. Here, we report on the results obtained with an adapted version of the WANR protocol in a larger, randomized, controlled, assessor-blinded, two group clinical trial aimed at evaluating the concurrent morphological and functional brain changes that take place during motor recovery in SCI. A total of 19 ASIA A, SCI patients (age 21 to 59; lesion time 13 months to 25 years) were initially allocated in two groups: a control classical neurorehabilitation (NR, n=9), and an experimental group (WANR, n=10). Later on, four patients that had finished the NR protocol joined the WANR group. Altogether, 14 patients were trained with the WANR (12 completed 9 months of training and two completed 5 months). When compared to the NR group, the WANR group exhibited a significant motor recovery, measured by the Lower Extremity Motor Score (LEMS) and the Walking Scale for Spinal Cord Injury (WISCI), which was detected at 5 months (mean±SD LEMS= 1.83±1.19 and WISCI= 5.92±2.78), but peaked at 9 months (mean±SD LEMS= 2.75±1.54 and WISCI= 9.75±2.14). Overall, 50% of these WANR-trained patients made a transition from ASIA A to ASIA C during this period. Longitudinal brain imaging analysis revealed that chronic SCI induced a widespread reduction in cortical thickness, including a substantial bilateral atrophy (between 15-22%, or 0.4-0.8mm) in many cortical regions in the insula, temporal, frontal, parietal, and even the occipital lobes. These findings are consistent with a variety of cognitive impairments observed in 40-64% of SCI patients. Moreover, they reinforce the hypothesis that SCIs trigger an acceleration of the normal processes of brain aging, which could explain the higher risk that SCI patients have of developing neurodegenerative disorders. Training with the WANR induced a very significant process of cortical plasticity, represented by a significant reversal of cortical atrophy (up to 1mm in the temporal lobe and insula) and an increase in both functional connectivity strength in cerebellar regions and in eigenvector centrality in executive and integrative cortical regions, like the angular gyrus, precuneus, and middle frontal gyrus. Meanwhile, sensorimotor hubs experienced a centrality reduction. This suggests a functional shift from damaged motor loops towards potential compensatory cortical networks. Accordingly, we propose a new mechanism for BMI-induced clinical improvement and raise the hypothesis that the WANR could induce neurologic recovery in other neurological conditions that produce cortical atrophy. Our findings also suggest that BMI training, coupled with virtual reality, and vigorous exercise or robotic-assisted walking, may also help mitigate or delay the normal process of brain aging in elderly patients. Our findings also categorically indicate that there is neither clinical, nor ethical justification for employing highly invasive cortical implants to treat paraplegics, given that non-invasive protocols, like the WANR, may suffice to induce significant neurological and functional gains in those patients with a few months of non-invasive neurorehabilitation.

## INTRODUCTION

A recent global epidemiological study has indicated that in 2021 close to 3.4 billion people worldwide, or 43% of the world’s population, suffered from one of the 37 most prevalent neurological disorders [1]. In China alone, the estimated prevalence of neurological disorders in 2021 reached 468.29 million people [2]. This corresponded to 32.91% of the entire Chinese population and 34.5% of the total prevalence of all diseases in China [2]. Globally, around 15 million patients (0.44%) were found to live with the devastating clinical consequences of a spinal cord injury [1], which impacted profoundly their quality of life and longevity [3].

Exactly 25 years ago, the possibility of directly linking animal brains to robotic devices, through a direct brain-machine interface (BMI), was first demonstrated in rats [4] and monkeys [5–7], and soon after in humans [8, 9]. Although this paradigm emerged due to the introduction of new technologies for chronic and simultaneous, multielectrode neuronal recordings obtained via brain implants, and was originally aimed as a new approach to investigate how large populations of individual neurons interact to generate motor behaviors, the enormous clinical potential of BMIs was recognized early on [10–12]. Proof of this is that, merely 5 years after the publication of the original BMI description in rats, 11 Parkinsonian patients learned to operate, in a few minutes, a brain-machine interface introduced to them during a deep brain stimulator implant surgery [9]. This study demonstrated for the first time that human subjects, even when severely impaired, could seamlessly operate a closed-loop BMI with very little training, based on recordings from many single neurons. This work was soon reproduced by several groups [8, 13–21], raising the hypothesis that BMIs could revolutionize the neurorehabilitation of patients suffering from severe body paralysis, such as that caused by a devastating spinal cord injury (SCI).

Eleven years ago, this latter hypothesis began to be tested in a group of seven ASIA A and one ASIA B paraplegics by the Walk Again Project (WAP), a non-profit international consortium, through the design and implementation of a BMI-based neurorehabilitation protocol, named the Walk Again Neurorehabilitation (WANR) [14, 18–20]. The WANR was the first protocol to combine immersive virtual reality, realistic visual-tactile feedback, and robotic-assisted locomotion controlled by an EEG-based, non-invasive BMI, especially designed to restore autonomous locomotion in SCI patients.

The initial application of the WANR indicated that long-term training with a BMI could not only assist individuals with SCIs in regaining mobility, but it could also trigger a very significant partial neurological recovery, including major improvements in voluntary motor control of lower limbs, recovery of lower body tactile and proprioceptive perception, and reestablishment of major visceral functions, such as bladder control [14]. Such an important neurological recovery became detectable within a few months of training and peaked after 28 months of continuous training. Overall, it meant that 7 out of 8 ASIA A/B complete SCI patients (one patient dropped out of the study at 12 months) evolved from complete to partial paraplegia, reaching an ASIA C classification by the end of this period [20].

Upon the publication of the WANR findings, other groups entertained the possibility of using invasive BMIs, implemented through a variety of brain implant approaches [17, 21], to allow paraplegic patients to regain mobility. Most of these studies involved primarily ASIA C, i.e., SCI patients with partially preserved motor capabilities. As such, this made the interpretation of their clinical results somewhat difficult or, in some cases, even misleading. For instance, an approach that had been pioneered originally in rodents [22], known as brain-spine interface, was reported to improve the locomotion of one ASIA C patient who had been subjected to extensive prior training with an invasive BMI [17]. Clearly, larger clinical trials are needed to validate this single-case report claim.

More recently, several startup companies have aggressively pushed the claim that only invasive BMIs, based on chronic brain implants of still unknown or undocumented longevity, would be appropriate for the rehabilitation of both paraplegic and quadriplegic patients. A famous example is the Neuralink’s PRIME study (clinical trial ID NCT06429735 https://clinicaltrials.gov/study/NCT06429735?term=neuralink&rank=1). Such claims for the use of invasive BMIs, based primarily on single patient reports or short-term evaluation of a small number of subjects, tend to minimize or even ignore the well-known current limitations in both long-term performance and biocompatibility of chronic cortical implants, which have been documented in both experimental animals [23, 24] and human clinical studies [8]; for a recent review see [25]. Therefore, to this day, it remains undecided whether invasive versus non-invasive BMIs offer the best approach to induce significant neurological recovery in chronic SCI patients, and which of these solutions would offer the safest, most efficacious, most affordable, and most scalable neurorehabilitation solutions to offer tens of millions of chronic SCI patients worldwide. Parallel to this debate, the neurophysiological mechanisms that allow patients to express significant functional recovery after long-term training with BMI-based protocols remain totally unknown.

Here, we addressed some of these fundamental questions by using a slightly revised version of the WANR (i.e. absence of haptic feedback; see Methods) in a randomized, controlled, assessor-blind, single-center trial that involved a total of 19 ASIA A, SCI patients (age 21 to 59; lesion time 13 months to 25 years) that were initially allocated in two groups: a control classical neurorehabilitation (NR, n=9), and an experimental group (WANR, n=10). Later on, four patients that had finished the NR protocol joined the WANR group. Altogether, 14 patients were trained with the WANR (12 completed 9 months of training and two completed 5 months). Starting at 5 months of WANR training, we detected significant motor recovery in these SCI patients, which peaked at 9 months. No significant motor recovery was observed in any of the patients included in the NR group. This meant that after 9 months of training, 50% of the SCI patients had already transitioned from a complete to a partial paraplegia status (from Asia A to ASIA C). Moreover, even a patient with a 20-year-old SCI demonstrated significant clinical recovery, becoming capable of walking with crutches following the training. After 9 months of training, we observed statistically significant levels of motor recovery, measured by Lower Extremity Motor Score (LEMS) and Walking Index for Spinal Cord Injury (WISCI) evaluation and electromyography (EMG) recordings.

To examine what neurobiological mechanisms could be mediating such a remarkable motor recovery, we applied multiple brain imaging techniques (structural and functional MRI), as well as measured visual evoked response potentials, at the onset, middle point (5 months) and 9 months after training onset. Brain imaging revealed that SCI induced a widespread reduction in cortical thickness that spread well beyond the motor and somatosensory cortex to include frontal, temporal, parietal, and even visual cortical areas located throughout the occipital lobe. Training with the WANR induced a significant reversal of such widespread cortical atrophy, a finding that was further confirmed by functional MRI analysis, which included changes in multiple cortico-cortical and subcortical circuits, including cerebellar regions. In addition, training with the WANR also reversed the profound changes in ERP magnitude and latency observed in our chronic SCI patients. Altogether, brain imaging and ERP analysis suggests that training with a BMI induces a significant and widespread process of cortical plasticity.

## MATERIAL AND METHODS

### 2.1 Experiment Design

Our study was classified as a randomized, controlled, assessor-blind, two-group, single-center trial. The primary and secondary outcome measures (Table 2) were performed three times: before the beginning of training (pre), at the end of 18 weeks (mid or 5 months), and after the end of the 36-week intervention phase (final or 9 months). A diagram representing the entire study can be seen in Figure 1.

**Figure 1.**
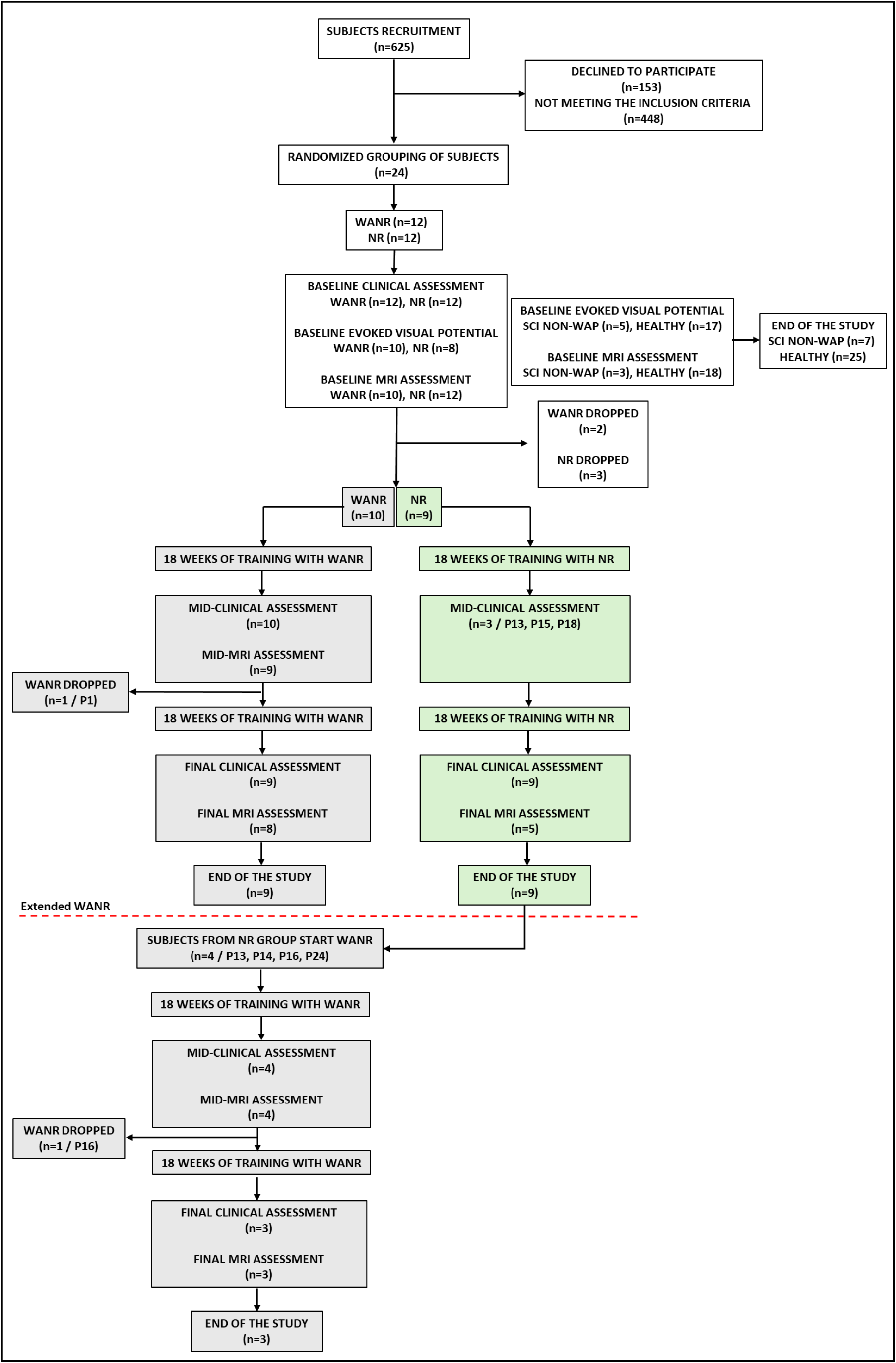
Diagram representation of the clinical trial flow. White boxes indicate actions performed for patients before the initiation of the WANR (experimental) and NR (control) training protocols. Grey boxes represent procedures specific to the WANR protocol, while green boxes denote those related to the NR protocol. The red dashed line marks the point at which patients who completed the NR protocol began the WANR protocol.

### 2.2 Participants and Randomization

The study was conducted at the AASDAP-Xuanwu Neurorehabilitation Laboratory located in the Xuanwu Hospital of the Capital Medical University in Beijing, China, from June 2019 to April 2024. Our recruitment team accepted volunteer enrollment applications through various means, such as outpatient clinics, paper applications, and emails. The hospital team conducted in-depth communications with volunteers through face-to-face interviews, home visits, emails, and telephone contacts to understand their general condition, willingness to enroll, and compliance with research criteria.

#### Inclusion criteria

Our inclusion criteria were the following: (1) discomplete spinal cord injury; (2) no traumatic head injury lesion; (3) lesion level at thoracic vertebrae 9 to 12 (T9-T12); (4) lesion duration > 12 months; (5) ability to perform daily activities; (6) stable emotional state.

#### Exclusion criteria

(1) age below 18 years or above 60 years; (2) inability to guarantee the time required for participation in this project; (3) participation in other research projects; (7) presence of acute or chronic decompensated diseases.

As shown in Figure 1, during the enrollment period, a total of 625 volunteers were contacted, among which 24 volunteers met the inclusion and exclusion criteria and, as such, were enrolled in the clinical trial. An independent on-line randomization routine concealed treatment allocation from investigators and blinded assessors prior to randomization of subjects. The 24 subjects were randomly assigned to either the Walk Again Neurorehabilitation protocol (WANR) group or the classical neurorehabilitation protocol (NR) group. During the experiment, due to factors such as COVID-19, three subjects from both the WANR and the NR groups dropped out, leaving nine subjects in each group completing the experiment. The patients’ demographic data are presented in Table 1.

**Table 1.**
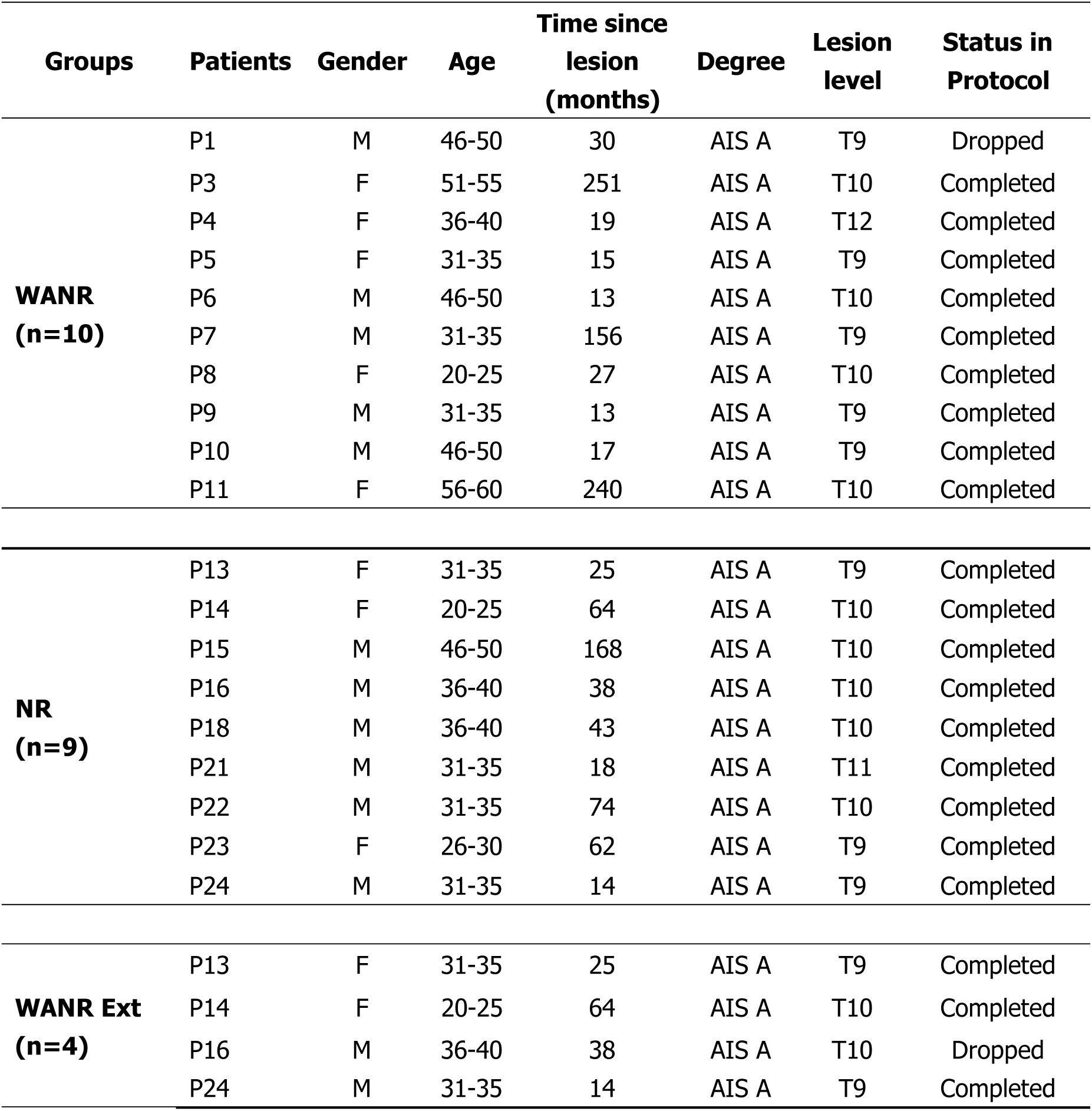
Patient’s demographic for WANR and NR groups. WANR Ext patients are those who migrated to WANR group after completing the NR protocol. Patients were categorized by study status as “Completed” (those who completed the 36 weeks of WANR/NR training) or “Dropped” (P1 and P16 withdrew from the study after mid-assessment of WANR and WANR Ext, respectively). Further details are provided in Figure 1.

| Groups | Patients | Gender | Age | Time since<br>lesion<br>(months) | Degree | Lesion<br>level | Status in<br>Protocol |
| --- | --- | --- | --- | --- | --- | --- | --- |
| **WANR**<br>**(n=10)** | P1 | M | 46-50 | 30 | AIS A | T9 | Dropped |
|  | P3 | F | 51-55 | 251 | AIS A | T10 | Completed |
|  | P4 | F | 36-40 | 19 | AIS A | T12 | Completed |
|  | P5 | F | 31-35 | 15 | AIS A | T9 | Completed |
|  | P6 | M | 46-50 | 13 | AIS A | T10 | Completed |
|  | P7 | M | 31-35 | 156 | AIS A | T9 | Completed |
|  | P8 | F | 20-25 | 27 | AIS A | T10 | Completed |
|  | P9 | M | 31-35 | 13 | AIS A | T9 | Completed |
|  | P10 | M | 46-50 | 17 | AIS A | T9 | Completed |
|  | P11 | F | 56-60 | 240 | AIS A | T10 | Completed |
| **NR**<br>**(n=9)** | P13 | F | 31-35 | 25 | AIS A | T9 | Completed |
|  | P14 | F | 20-25 | 64 | AIS A | T10 | Completed |
|  | P15 | M | 46-50 | 168 | AIS A | T10 | Completed |
|  | P16 | M | 36-40 | 38 | AIS A | T10 | Completed |
|  | P18 | M | 36-40 | 43 | AIS A | T10 | Completed |
|  | P21 | M | 31-35 | 18 | AIS A | T11 | Completed |
|  | P22 | M | 31-35 | 74 | AIS A | T10 | Completed |
|  | P23 | F | 26-30 | 62 | AIS A | T9 | Completed |
|  | P24 | M | 31-35 | 14 | AIS A | T9 | Completed |
| **WANR Ext**<br>**(n=4)** | P13 | F | 31-35 | 25 | AIS A | T9 | Completed |
|  | P14 | F | 20-25 | 64 | AIS A | T10 | Completed |
|  | P16 | M | 36-40 | 38 | AIS A | T10 | Dropped |
|  | P24 | M | 31-35 | 14 | AIS A | T9 | Completed |

### 2.3 Training protocol

Both the WANR and the NR groups underwent specific rehabilitation training programs, four times a week, for 36 weeks (Fig. 2A).

**Figure 2.**
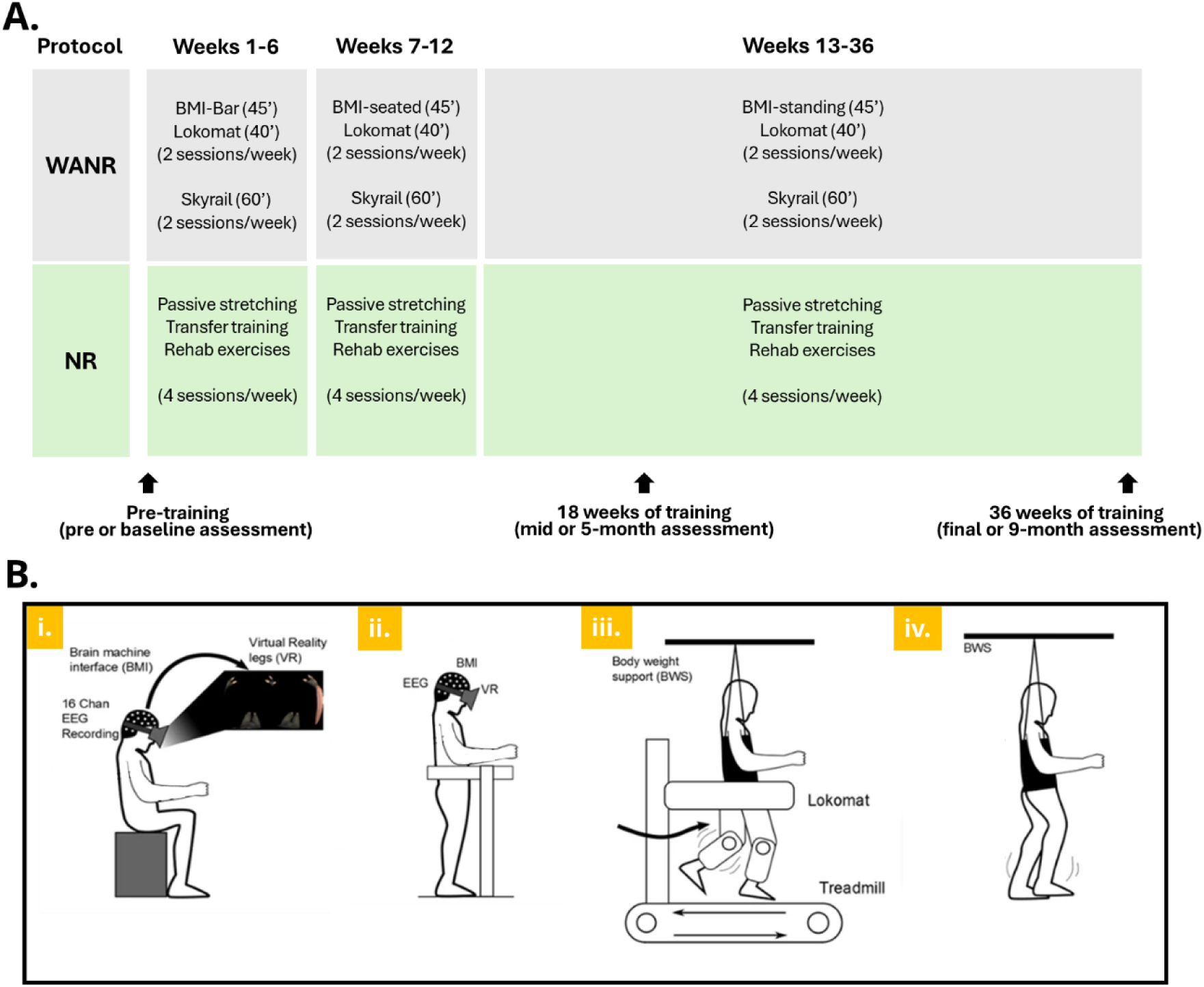
Experimental protocol. (**A**) Training protocol for the WANR group (grey) and NR group (green). Both groups underwent a training routine four times a week, for 36 weeks. Clinical assessments (indicated by arrows) were performed three times: Pre-training (pre or baseline assessment), at the end of 18 weeks of training (mid or 5-month assessment), and at the end of 36 weeks of training (final or 9-month assessment). (**B**) Experimental protocol for WANR group (figure adapted from Donati, Shokur et al, 2016). BMI training was performed in three different situations: (**i**) EEG-BAR training and BMI+VR seated training; and (**ii**) BMI+VR standing training. Assisted locomotion training was performed with (**iii**) Lokomat, a robotic gait system in a treadmill, and with (**iv**) Skytrack, a BWS gait system fixed on an underground track. On a given week, patients went through two sessions of BMI, followed by locomotion training with Lokomat, and two sessions of locomotion training with Skyrail. BMI + Lokomat training and Skyrail sessions were performed on alternate days.

#### WANR group

Patients in this group followed a routine that included two sessions of non-invasive Brain-Machine Interface (BMI) training with a 16-channel EEG system (Brain Products) and an immersive virtual reality head mounted displayer (Oculus Rift, Oculus VR), followed by locomotion training with a robotic gait system on a treadmill (Lokomat, Hocoma AG), and two sessions of locomotion training with a robotic body weight support (BWS) system fixed on an overground track (SkyTrack 800, Humaneotec) (Fig. 2B). The BMI training content for the WANR group was implemented in three phases: EEG-BAR training for weeks 1-6; BMI+VR seated training for weeks 7-12; and BMI+VR standing training for weeks 13-36. The BMI training consisted of alternating left and right leg motor imagery to trigger the corresponding stepping of a 3D avatar projected in the VR head-mounted display. Each BMI training session lasted 45 minutes. BMI setups and methods used in this study are the same as described in previous WAP studies [14, 18, 20]. However, an important difference in the present study methodology was the absence of haptic feedback, which was not applied to the subjects during the BMI sessions. For this reason, the WANR utilized in this investigation is regarded as a modified form of the WANR.

After the BMI training, there was a rest period of 10-20 minutes, followed by 40-minute Lokomat training. The Skytrack training and BMI training were conducted on alternate days each week, with each Skytrack training session lasting approximately 60 minutes. During the Skytrack training, patients were required to wear weight-bearing harnesses and KAFO (Knee Ankle Foot Orthoses) and use walkers for walking. With the Skytrack system supporting up to 70% of the patient’s body weight, therapists assisted the patients in standing and walking exercises.

#### NR group

Patients in this group followed a routine that included passive stretching, assisted standing, transfers, and other rehabilitation exercises based on their individual needs (Fig. 2A). When patients first entered the study, researchers demonstrated to both the patients and their families, through practical demonstrations, how to perform traditional home-based rehabilitation training. After the patients returned home, researchers oversaw and guided the patients and their families in completing the home-based rehabilitation training remotely (via telephone, WeChat, etc.).

After completing the 36-week controlled observation period, three patients in the NR group underwent the WANR rehabilitation training program following the same training procedures as the WANR group (Fig. 1). Thereafter, these three patients completed the 36-week rehabilitation training.

### 2.4 Clinical Assessments

To evaluate the effect of training on patients, we performed clinical assessments in three different periods (Fig. 2A): before the beginning of the training (pre-training assessment), at the completion of 18 weeks of training (mid or 5-month assessment), and after 36 weeks of training (final or 9-month assessment). The three assessments were performed by independent assessors. The assessment outcome measures and items are presented in Table 2.

**Table 2.** Assessment items and outcome measures.

| Outcome Measures | Items | Assessments |
| --- | --- | --- |
| Primary | Lower limbs motor function | ASIA motor score (LEMS) |
| Secondary | Nociception improvement | ASIA pin-prick sensory score |
|  | Walking function | Walking index for SCI (WISCI) |

#### 2.4.1 ISNCSCI assessments

Lower Extremity Motor Score (LEMS), Pin-Prick (PP) sensory score, Light-Touch (LT) sensory score, ASIA Degree, and ASIA injury Level were assessed. All clinical assessments were conducted in accordance with the standards of the International Standards for Neurological Classification of Spinal Cord Injury (ISNCSCI; Revised 2011).

**LEMS** was the primary outcome measure of the study. It evaluated motor function on a scale of 0 (no motor function) to 5 (full motor function), for five lower extremity muscle groups, with a 50-point maximum (25 per side). **Pin-Prick** was the secondary outcome measure of the study, which evaluated nociception improvement. It involved testing a key point in each of the 28 dermatomes (from C2 to S4-5) on the right and left sides of the body that can be readily located in relation to bony anatomical landmarks. Appreciation of pin prick sensation at each of the key points is separately scored on a three-point scale, with comparison to the sensation on the patients’ cheek as a normal frame of reference: 0 = absent; 1= altered; 2 = normal or intact; NT = not testable. The maximum scale was 112.

#### 2.4.2 Walking index for SCI (WISCI)

WISCI was the secondary outcome measure employed in this study to evaluate the walking ability of SCI patients. WISCI level is recorded on a scale of 0 (inability to keep a standing position) to 20 (ambulation with no devices, no braces and no physical assistance). According to the WISCI requirements, we provided patients with testing equipment, such as parallel bars, braces, walkers, crutches, cane and a 10-meter testing site. Depending on the patients’ functional level, 1-2 therapists provided necessary safety protection and guidance during the test.

EMG recordings from the quadriceps muscles of both legs were obtained from some patients from the WANR group.

### 2.5 Statistical analysis

Statistical analysis of primary and secondary measurements was conducted using SPSS 20.0 statistical software. Based on the characteristics of the data, ANOVA, paired T-test, independent sample T-test, and rank sum test methods were used for statistical calculations on the data. Repeated-measures two-way ANOVA was performed to assess the effects of **Group** (WANR vs. NR) and **Time** (Pre vs. Post) on motor outcomes (LEMS and WISCI). Post hoc paired *t*-tests within each group were conducted when significant interactions were detected. Results are expressed as **mean ± SD**, with individual data points plotted. Statistical significance is indicated with asterisks (*p* < 0.05, *p* < 0.01, *p* < 0.001), and non-significant comparisons are labeled as ns.

### 2.6 Study approval

This experiment has been approved by the Ethics Committee of Xuanwu Hospital and has been registered on ClinicalTrials.gov (https://clinicaltrials.gov/study/NCT03992690) with the registration number NCT03992690 and a registration date of August 1, 2019.

### 2.7. Magnetic Resonance Imaging (MRI)

To evaluate the possible effects of training on the structure of patients’ brains, we performed structural and functional MRI with diffusion tension imaging (DTI) imaging at the onset, middle point (5 months) and 9 months after training onset. Not all patients enrolled were able to undergo such imaging exams due to pandemic restrictions and particular clinical conditions (e.g., metal implants to stabilize the spine).

For the purpose of brain imaging analysis, we used the MRI data from the subjects recruited for this study, and also from two other sources - an open database of healthy subject MRIs (OpenfMRI database, Washington University) and our previous WANR study in Brazil. These subjects were distributed and analyzed as follows:

#### 2.7.1. Subjects Control groups

##### Healthy individuals

20 T1 weighted MRIs of healthy individuals from the OpenfMRI database (Washington University 120) with average age of 31.45 years, balanced sex ratio (10 males, 10 females), with no spinal cord injury (SCI) or paraplegia.

##### SCI group

23 patients with SCI, prior to submission to any rehabilitation protocols.

#### Experimental groups

##### WANR-BR

Six patients from a previous WAP study in Brazil, who participated in the WANR protocol, showed improvements in sensorimotor, visceral and psychological signs.

##### WANR-CN

Eleven patients of the present study, conducted in China, participated in the same WANR protocol. Of these 11 patients, two started the WANR protocol later, after finishing participation in the NR group. These two patients underwent MRI scans in five timepoints, including 5 months after starting training and 9 months.

Of the 23 patients of the SCI group who underwent MRI at baseline, five dropped from the protocol at the beginning of the study and were therefore not included in the subsequent MRIs. Fifteen participants underwent the final MRI (9 months): 10 from the WANR group and five from the NR group.

The inclusion criteria for SCI patients were as follows: (i) thoracic TSCI lasting more than one year; (ii) grade A according to the ASIA disability classification standard of the American Spinal Injury Association; (iii) no head or brain injury related to the trauma that caused the injury, and no psychiatric illness; (iv) no abnormalities found on routine brain MRI; (v) the subject’s handedness was evaluated using the Edinburgh Handedness Inventory, which determined that the subject was right-handed; and (vi) no contraindications to MRI.

The inclusion criteria for healthy volunteers, matched by age and gender, which were included as control subjects are as follows: (i) no history of head or brain injury; (ii) no medical or psychiatric conditions; (iii) no contraindications to MRI; and (iv) right-handedness.

#### 2.7.2. Image acquisition

All subjects recruited in this study were scanned using a GE 750 3.0 Tesla MRI scanner equipped with an 8-channel head coil. The scanning sequence mainly included Sagittal 3D T1-weighted, axial DTI and fMRI scans. The detailed sequences were collected from the brain as follows:

Sagittal 3D T1-weighted images were acquired with the following specific scanning parameters: repetition time (TR)=7700 ms, echo time (TE)=2900 ms, flip angle=12°, field of view (FOV)=224×224 mm2, matrix size = 224×224, isotropic voxel size=1×1×1 mm3, 166 slices, and slice thickness =1 mm. The total acquisition time of T1WI for each subject was approximately 3 minutes and 39 seconds.

Axial DTI: TR = 9232 ms, TE = 972 ms, acquisition matrix=112×112, FOV = 224×224mm, isotropic voxel=2×2×2mm3, slice thickness = 3.5mm, number of slices = 40, diffusion sensitive gradients at b = 1000 s/mm² were applied in 64 non-collinear directions, b0=10. The total time required for the DTI acquisition was 11 minutes and 32 seconds.

The resting-state functional data were obtained using a gradient-echo-planar imaging (EPI) pulse sequence as follows: TR = 2000 ms, TE = 30 ms, FOV = 224 mm × 224 mm2, slice thickness = 3.5 mm, matrix size = 64 × 64 and number of slices = 33. The total fMRI acquisition time was up to 8 minutes. During resting-state fMRI acquisition, subjects need to close their eyes to relax themselves and stay awake, without performing any thoughts or tasks.

DTI data were saved for another separate publication.

#### 2.7.3. Methods for Data Analysis

Since we have multiple data from different sources and a limited number of patients with heterogeneous clinical features, traditional statistics can underestimate the changes occurring at an individual level. Therefore, in addition to the traditional approach, a multilayered method of analysis was developed to understand the effects of BMI on brain structure, physiology, function, and correlation with clinical improvements.

##### First Layer: Structural brain MRI

This analysis gave us access to cortical characteristics of our subjects, mainly their cortical thickness and volume. Structural brain MRI data were analyzed using T1-weighted scans of different groups, detailed in section 2.7.1. The classical Voxel Based Morphometry [26] and statistical analysis was performed in CAT12.9 (Computational Anatomy Toolbox for SPM12): A toolbox of SPM12, which runs in Matlab R2023b, was utilized for advanced cortical thickness analysis, including pre-processing steps such as bias correction, spatial normalization, and tissue segmentation. Cortical thickness and central surface of the left- and right-side hemispheres were calculated following the projection-based thickness protocol (PBT) method [27]. A longitudinal analysis was performed for each individual who had at least two timepoints. Two-sample t tests, voxel p<0.001, cluster p<0.05, GRF correction were performed between groups (training x non-training, healthy x SCI, for example). The DK40 atlas was used for cortical parcellation and region-of-interest (ROI) analysis. It refers to the Desikan-Killiany atlas [28], a widely used anatomical brain atlas for cortical parcellation. It segments the cerebral cortex into 68 regions of interest (ROIs), with 34 regions per hemisphere. These regions correspond to well-defined anatomical landmarks based on gyral and sulcal structures observed in human brain anatomy. The DK40 atlas is commonly applied in structural brain MRI studies to measure cortical thickness, surface area, and volumes of specific cortical regions. It is especially useful for understanding regional brain morphology and its relationship to clinical or experimental conditions. For comparison with non-paraplegic individuals, a public database of 20 healthy individuals was also processed and compared to our SCI patients (see section 2.7.1.). Hedges’s g test was used for testing the significance of the differences in cortical thickness between the control and experimental groups, and between the two subgroups of the experimental group (i.e. patients that moved from ASIA A to C, and patients that remained as ASIA A after 9 months of training).

Hedges’ g represents how many pooled standard deviations apart two group means are, with a small-sample bias correction. Its magnitude conveys the effect size (∼0.2 small, ∼0.5 medium, ∼0.8 large, ≥1.2 very large). It is a unit-free scale, and it is widely used, especially in psychology, education, biomedicine, and meta-analysis [29].

##### Second Layer: Functional brain MRI

While the first layer accounts for changes especially in grey matter, the second layer, functional MRI (fMRI), was obtained to investigate functional connectivity and activity patterns in resting state.

All fMRI data were processed in the Conn functional connectivity toolbox (22.a), a MATLAB-based functional connectivity software used for pre-processing and statistical analysis of fMRI data. The pre-processing pipeline includes motion correction, normalization, and spatial smoothing. Data were aligned to a standard brain atlas to ensure cross-participant comparison. Resting-state functional connectivity was calculated using seed-to-voxel and ROI-to-ROI analysis. The 2^nd^ level analysis was run with standard settings for cluster-based inferences (cluster threshold *p < 0.05*, cluster-size p-FDR corrected, voxel threshold *p< 0.001* p-uncorrected). Results were statistically analyzed to determine functional network differences, particularly in SCI patients compared to controls and pre-post training differences. Analysis of eight parameters (global efficiency, local efficiency, betweenness centrality, closeness centrality, cost, average path length, clustering coefficient and degree) of 110 networks and 132 ROIs were computed and extracted for each patient and control.

For comparison with non-paraplegic individuals, a public database of 20 healthy individuals’ resting state fMRI with the eyes closed was also processed and compared to our SCI patients, with matching age and sex, obtained from the OpenNeuro documentation website, from the Human Connectome Project.

##### Third Layer: Evoked Visual Potentials

In this analysis layer, the physiological response to evoked visual stimuli was evaluated both in SCI and control patients. Of those, a total of 45 participants were recruited from Xuanwu Hospital. A total of seven participants were removed due to severe pain or because of a variety of circumstances associated with the COVID pandemic. Thus, the final sample consisted of 38 individuals, including 17 SCI patients and 21 healthy subjects. There was no difference in terms of gender and age between these two groups. All participants exhibited normal or corrected-to-normal vision. The Ethical Committee for Clinical Research of the Xuanwu Hospital approved the experimental protocol, and all participants provided informed consent.

All the participants completed a paired comparison task which included facial and photographs of buildings. Eighty natural, neutral grayscale facial pictures were obtained from the Chinese Standard Emotional FacePicture System. Half of them were male and half female. At the same time, eighty face-building pictures were collected from the Internet.

All the photographs maintained the same luminance and contrast values. The image resolution was 260×300 (width × height). Each stimulus appeared in the center of the 17-inch LCD computer screen (resolution 1024×768 pixels, refresh rate 75Hz).

A total of 140 face trials and 140 face-building trials were presented in Random order, and 18 practice trials were used to ensure that all the participants understood the experiment and were excluded from formal analysis. The stimuli were presented by E-Prime 2.0.

EEG data were recorded from a 64 Ag/AgCl electrode cap (Yunshen, Inc.) according to the extended International 10–20 system. The data were referenced offline to a common average reference (the algebraic mean of all electrodes) and filtered offline with a bandpass filter of 0.1–48Hz (24 db/oct) The scalp interelectrode impedances of each subject were guaranteed to drop below 5kΩ.

###### ERP data analysis

Baseline (V0) measurements were performed on all control and SCI patients. Long-term follow-up was conducted in seven individuals, and ERP results were collected twice (V1 V2) to evaluate the intervention effect. Based on previous research, six posterior electrode sites (O1, O2, PO3, PO4, PO7 and PO8) were considered for the analysis of the P1 from 80ms to 120ms. Eight electrodes between140-200ms (O1, O2, PO3, PO4, PO7, PO8, P7 and P8) from the parietal and parieto-occipital regions were used for the analysis of the N170. The P2 and N2 was scored between 140-200ms and 200-300ms over occipital frontal clusters (F3, FZ, F4, FC3, CZ, FC4, C3, CZ and C4).

For comparison purposes between healthy controls and SCI patients, the following electrode locations were used to measure the P1, N1, P2, and N2.

P1 : O1, O2, PO3, PO4, PO7, PO8

N1 : O1, O2, PO3, PO4, PO7, PO8, P7, P8

P2 : F3, FZ, F4, FC3, FCZ, FC4, C3, CZ, C4

N2 : F3, FZ, F4, FC3, FCZ, FC4, C3, CZ, C4

##### Fourth Layer: Clinical measurements

Clinical evaluation of the patients submitted to the WANR and NR protocols was performed systematically, as described in Section 2.4, using the ASIA standard assessment [30]. The motor score described the level of voluntary strength below the SCI level for each patient, ranging from 0 (absence of contraction) to 5 (for normal contraction, produced against gravity and strong opposing force). The lower extremity motor score (LEMS) was obtained by summing all key muscles (score for a healthy subject is 50, i.e., five key muscles, with a maximal score of 5, bilaterally). The Walking Index Spinal Cord Injury II (WISCI II) [31] was also an important tool to evaluate our patients, since it is a direct functional measurement that could be correlated to other measurements. Very important to this analysis were the time elapsed since the lesion occurred and patient’s age.

###### Multilayered analysis through principal components

Due to the high number of variables (clinical, imaging and neurophysiology) derived from a modest and heterogeneous cohort, the possibility of not identifying a pattern or finding spurious correlations always exists. To overcome this challenge, we opted to compare data grouping using principal components analysis (PCA). PCA is a widely used statistical technique for dimensionality reduction, data visualization, and feature extraction. It transforms a dataset with potentially many correlated variables into a smaller set of uncorrelated variables, known as principal components, while retaining as much of the variability/variance (information) in the dataset as possible. The first principal component (PC1) captures the largest variance in the data. The second principal component (PC2) is orthogonal (uncorrelated) to PC1 and captures the next largest variance, and so on. Data were standardized (mean = 0, standard deviation = 1) to ensure that all variables are on the same scale and equally contribute to the analysis. The covariance matrix of the datasets was computed to identify how variables are correlated. Data were projected onto the new axes (principal components) in a tridimensional PCA space, creating a transformed dataset with reduced dimensions.

PCA scores were calculated for cortical thickness, for all groups at all times, ERPs for all patients and all times, and fMRI measurements for all networks and ROIs. Pearson linear correlation analysis was performed in Matlab to evaluate possible correlations between PCAs of different sources (structural, functional and ERPs) with clinical measurements (Age, WISCI II, Lesion time). The results can be displayed in an *n-*dimensional space, with *n* equal to the number of variables that correlate with each other. In this way, we could simplify the complexity of evolution of structural, functional and physiological changes that occur in the brain during the natural course of SCI, its recovery with BMI and what factors influence for better and worse outcomes after SCI and BMI training.

## RESULTS

### 3.1. BMI Performance Analysis

As in our previous studies involving the WANR (e.g. [14, 18, 20], we initially evaluated the performance of our patients using the EEG-based BMI that serves as the core of our neurorehabilitation protocol. The same procedure was applied in the present study. Figure 3 depicts the performance of some of our SCI patients, in terms of classifier score (in percentage of trials correctly classified), showing that they were able to consistently perform this task above chance level. This allowed us to proceed with the regular training protocol as in previous studies, without any alteration in the training procedure.

**Figure 3.**
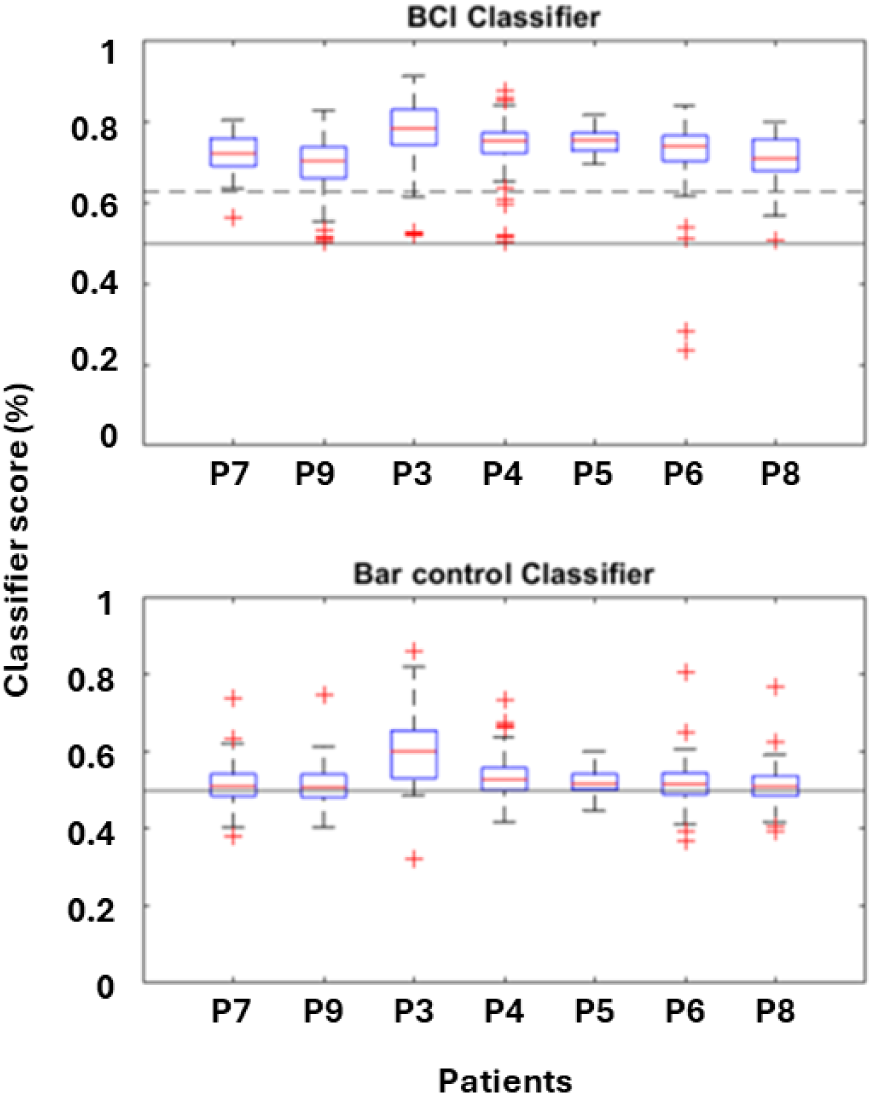
Classification accuracy of patients during BMI training. In both panels, the solid line between 0.4 and 0.6 denotes the boundary of chance level. Under the BCI classifier condition (top), with the exception of a few patients (e.g., P6), most patients achieved median accuracies of approximately 65%–80%, with concentrated distributions and performance significantly above chance, indicating stable and reproducible task execution. The Bar control condition served as a control to assess baseline performance in the absence of intentional control. Its classification accuracies were generally close to chance level, suggesting that the BCI classification results possess high specificity and are not substantially affected by external, non-specific factors.

### 3.2. Clinical Findings

In this study, a total of 14 ASIA SCI patients were trained with a modified version of the WANR protocol, which precluded the use of tactile feedback, but combined a non-invasive BMI, virtual reality and robotic walking. This included the original nine patients enrolled in the WANR group, plus three more patients that were transferred to the WANR group after being part of the original NR group for 9 months (see Figure 1 with Trial Flow in Methods). In addition, two more patients that did not complete the entire 9 months of WANR training were included in our analysis.

The proposed primary outcome of this clinical study was to measure changes in lower limb motor function induced by 9 months of training with the WANR protocol. This outcome was primarily assessed through the evolution of the ASIA motor score (LEMS) from the onset, mid (5 months), and end point (9 months) of training. In addition, other indices of motor performance, such as the Walking Index for Spinal Cord Injury (WISCI), and surface EMGs of multiple lower limb muscles, were used as secondary outcome measures of motor performance. Table 3 depicts the LEMS values obtained for the 12 SCI patients subjected to the entire duration of the WANR protocol, as well as the NR group at the onset, 5 months, and 9 months of the study. Meanwhile, Figures 4A and B show the evolution of LEMS for the WANR and NR group. Inspection of these graphs revealed that, while for the NR group there was no significant change in LEMS (mean±SD Pre= 0.56±0.88; Post= 0.78±1.20), a clear increase was documented for the WANR group, starting at 5 months, and peaking at 9 months of training (mean±SD Pre= 0.67±1.30; Mid= 1.83±1.19; Post= 2.75±1.54).

**Figure 4.**
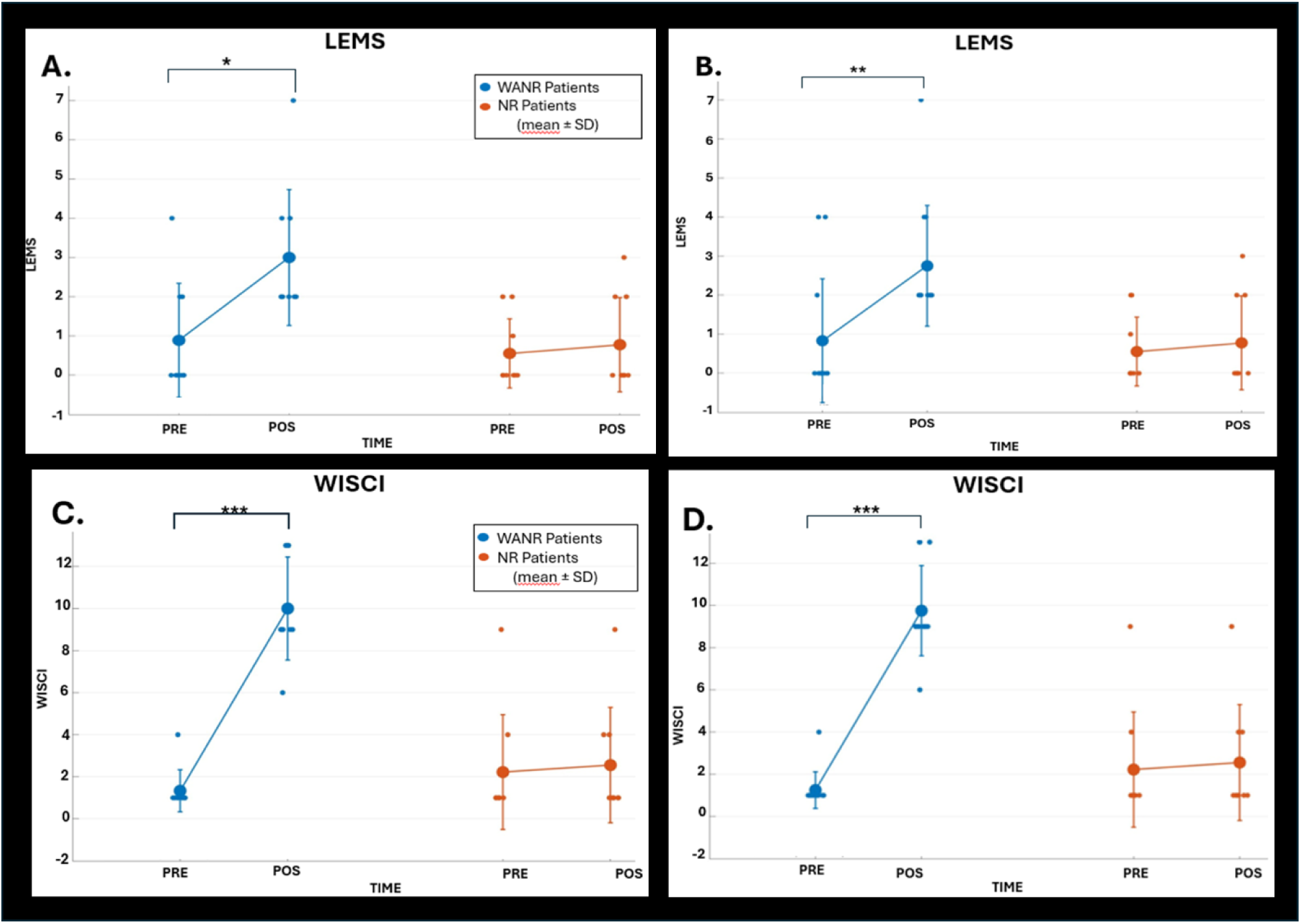
Repeated measures two-way ANOVA were performed to assess the effects of **Group** (WANR vs. NR) and **Time** (Pre vs. Post) on motor outcomes LEMS (**A** and **B**) and WISCI (**C** and **D**). Post hoc paired *t*-tests within each group were conducted when significant interactions were detected. Results are expressed as **mean ± SD**, with individual data points plotted. Statistical significance is indicated with asterisks (*p* < 0.05, **p** < 0.01, ***p*** < 0.001). Analysis revealed a significant **Time × Group interaction** for both LEMS and WISCI scores. In the WANR group, Post values were significantly higher compared to Pre (paired *t*-test, *p* < 0.05 to *p* < 0.001 depending on outcome), whereas no significant Pre–Post differences were observed in the NR group. Error bars represent SD, and figures display both individual subject values and group means**. (A)** LEMS mean ± SD for nine WANR and nine NR patients. WANR and NR Pre times (0.89±1.45; 0.56±0.88) and Post times (3.00±1.73; 0.78±1.20); t (paired, WANR)=-3.12, p=0.0142; t (paired, NR)=-0.80, p=0.447; ANOVA for time vs group interaction p= 0.0122. **(B)** LEMS mean ± SD for 12 WANR patients (9 original WANR plus 3 WANR extended) and nine NR patients. WANR and NR Pre times (0.67±1.30; 0.56±0.88) and Post times (2.75±1.54; 0.78±1.20); t (paired, WANR)=-3.62, p=0.004; t (paired, NR)=-0.80, p=0.447; ANOVA for time vs group interaction p= 0.015. **(C)** WISCI mean ± SD for nine WANR and nine NR patients. WANR and NR Pre times (1.33±1.00; 2.22±2.73) and Post times (10.00±2.45; 2.56±2.74); t (paired, WANR)=-11.93, p=2.24e-06/ t (paired, NR)=-1.00, p=0.347 / ANOVA for time vs group interaction p= 0.00054. **(D)** WISCI mean ± SD for 12 WANR patients (9 original WANR plus 3 WANR extended) and nine NR patients. WANR and NR Pre times (1.25±0.87; 2.22±2.73) and Post times (9.75±2.14; 2.56±2.74); t (paired, WANR)=-15.64, p=7.35e-09/ t (paired, NR)=-1.00, p=0.347; ANOVA for time vs group interaction p= 0.000132.

**Table 3.**
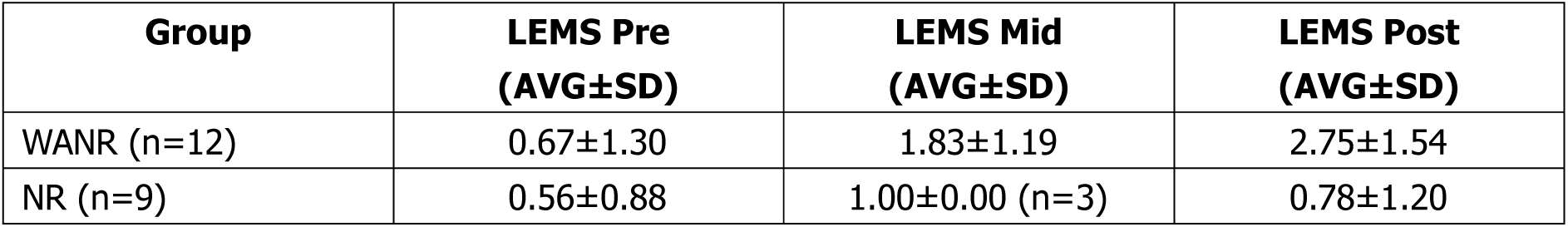
LEMS values (AVG±SD) obtained for 12 WANR and nine NR patients at Pre, Mid (5-month), and Post (9-month) assessments. Due to COVID restrictions, only three patients from the NR group went through Mid assessment; therefore, LEMS Mid statistics were not calculated (see Fig. 4).

| Group | LEMS Pre<br>(AVG±SD) | LEMS Mid<br>(AVG±SD) | LEMS Post<br>(AVG±SD) |
| --- | --- | --- | --- |
| WANR (n=12) | 0.67±1.30 | 1.83±1.19 | 2.75±1.54 |
| NR (n=9) | 0.56±0.88 | 1.00±0.00 (n=3) | 0.78±1.20 |

Inspection of Figure 5 reveals the evolution of WISCI for all 14 patients who participated in the WANR protocol (12 for 9 months, and two - P1 and P16 - for a shorter period of training). For all 14 patients, a considerable improvement in WISCI was already observed with 5 months of training, while the highest values were achieved for 12 patients with 9 months of training. The highest WISCI value reached was 13 for three patients, while eight patients reached a WISCI of 9, one reached 6, and two reached 4. P1 and P16 did not complete the full 9 months of training with the WANR protocol, therefore do not have WISCI measures for this period. Overall, the average WISCI for the WANR group that completed the protocol (n=12; mean±SD Pre=1.25±0.87; Mid=5.92±2.78; Post=9.75±2.14) was significantly higher than the control group (n=9; mean±SD Pre=2.22±2.73; Post=2.56±2.74) in which no change in WISCI was detected. Yet again, these WISCI values were very similar to those obtained in a previous study in which eight patients trained for 12 months with the same protocol (see Figure 3B in [14].

**Figure 5.**
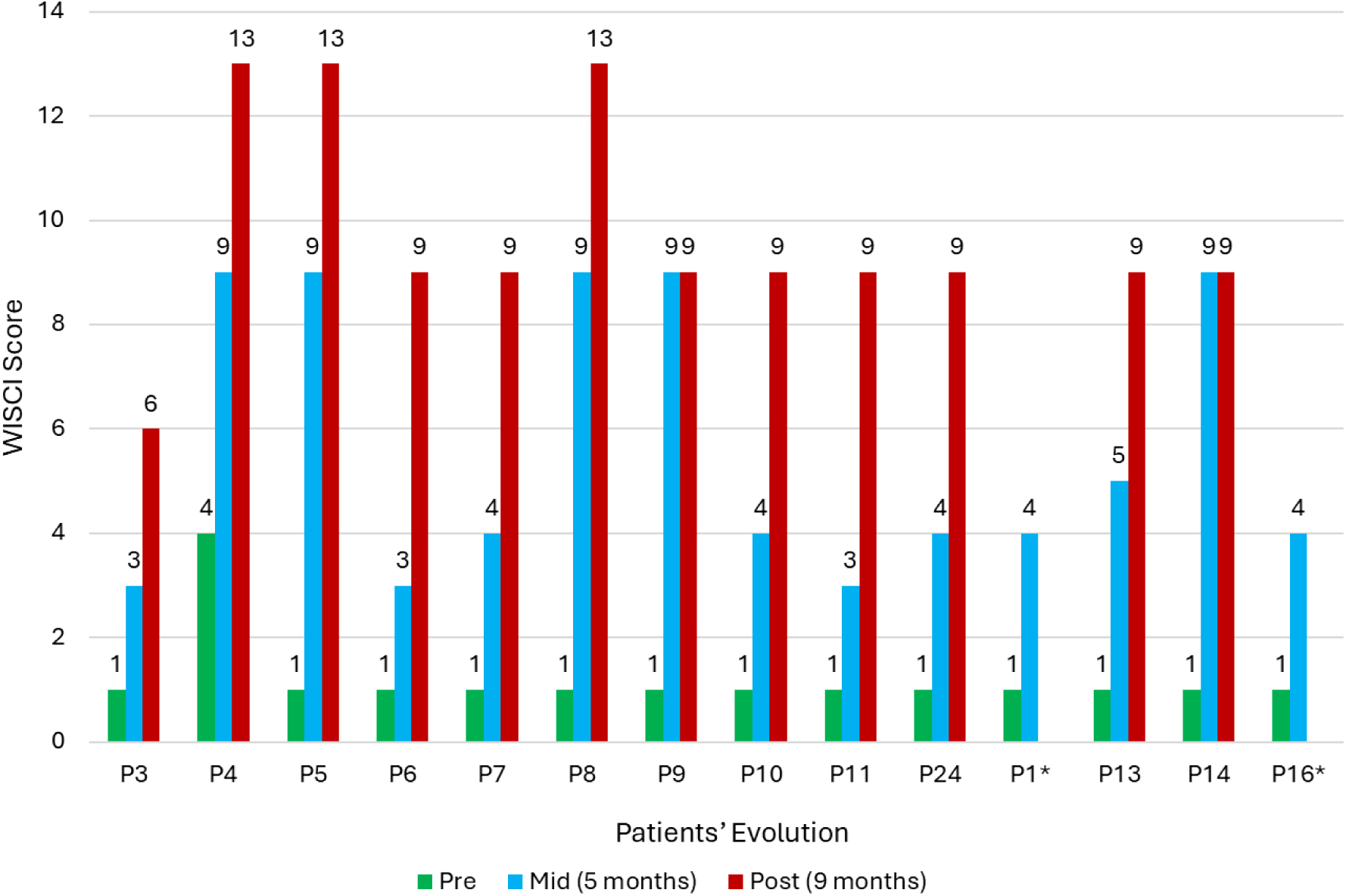
Evolution of WISCI for all the 14 patients of the present study subjected to the WANR protocol for at least 5 months and a maximum of 9 months. For all 14 patients, significant improvement in WISCI was observed as early as 5 months of training, while the highest values were achieved with 9 months of training for 12 patients. The highest WISCI value reached was 13 for three patients, while eight patients reached a WISCI of 9, one reached 6, and two reached 4 (P1 and P16 trained only for 5 months).

To better depict what such progress in voluntary muscle contractions and locomotion meant in terms of regaining voluntary control of lower limb musculature, Figure 6 illustrates the evolution of surface EMG recordings from the quadriceps muscles of both legs, obtained from patient P8, over a period of 36 weeks of training. This patient evolved from a LEMS of 0 to 5 in 9 months, while her WISCI went from 1 to 13 during the same period. As a result, the patient was upgraded from ASIA A to C after 9 months of training. As shown in Figure 6A, there was no, or only minimal EMG activity obtained on both the right and left legs of this patient at the onset of training. By week 14 (Figure 6B), some voluntary evoked muscle contractions could be identified primarily in the right leg. However, by week 36 (Figure 6C), clear voluntary muscle activity was recorded in both legs of the patient.

**Figure 6.**
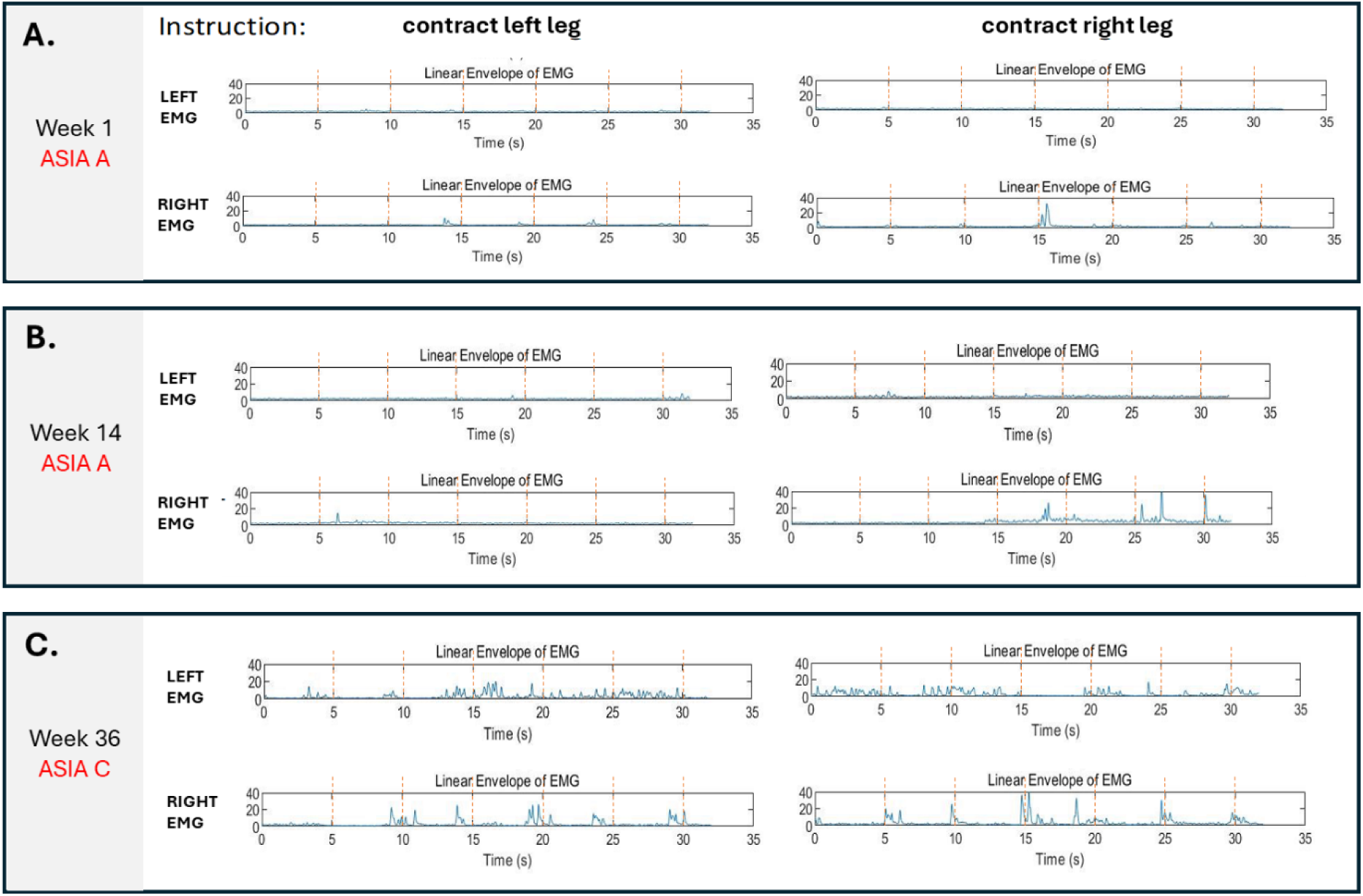
Evolution of surface EMG recordings obtained from the quadriceps muscles in both legs of patient P8 over a period of 36 weeks of training. For this patient, LEMS evolved from 0 to 5 in 9 months, and WISCI went from 1 to 13 in the same period. As a result, ASIA classification for this patient was upgraded from ASIA A to C after 9 months of training. **(A)** There was none or only minimal EMG activity obtained from the quadriceps of both the right and left legs of this patient at the onset of training. **(B)** By week 14, some voluntary evoked muscle contractions could be identified primarily in the right leg. **(C)** However, by week 36, clear voluntary muscle activity can be detected in both legs of the patient.

The overall ASIA evolution for all 14 patients who underwent the WANR for the entire duration of the protocol (n=12) or only partially (n=2) is shown in Figure 7. Overall, seven of the patients (50%) progressed from the original ASIA A status – complete paraplegia – to the ASIA C – or partial paraplegia. For six of these patients (P4, P11, P1, P13, P14, P16), the ASIA A to C transition required only 5 months of training, while for one remaining patient (P8) longer training, up to 9 months was needed for this transition to take place.

**Figure 7.**
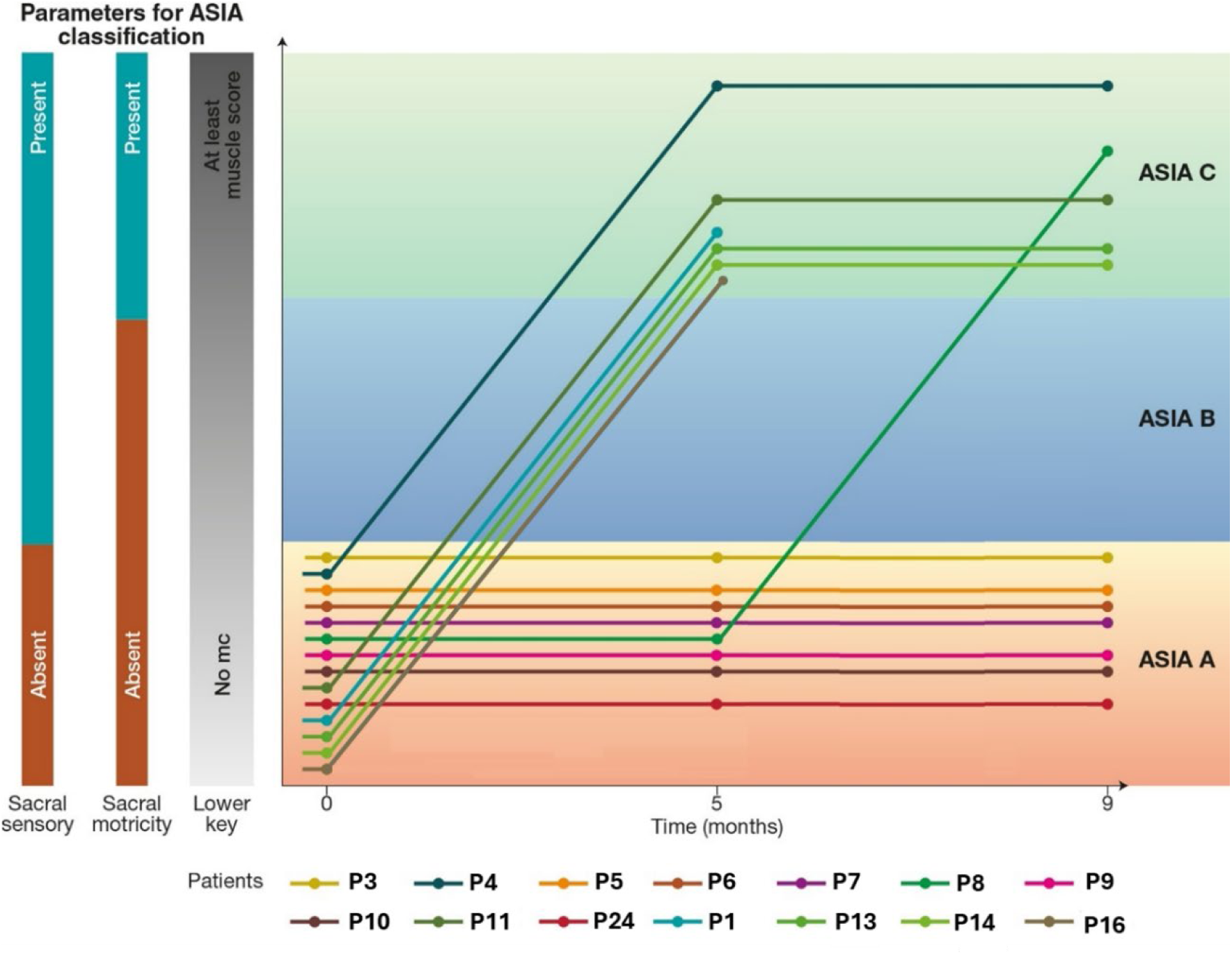
Evolution of ASIA classification for all 14 patients submitted to WANR. Overall, seven of the patients (50%) progressed from the original ASIA A status – complete paraplegia – to the ASIA C – partial paraplegia. For six of these patients (P4, P11, P1, P13, P14 and P16), the ASIA A-C transition required 5 months of training, while for one patient (P8), it was necessary 9 months for this transition to occur. Notice that P1 and P16 did not complete the full 9 months of training with the WANR protocol.

As mentioned above, the best locomotion recovery was observed in patient P4 who sustained a lower thoracic (T12) spinal cord lesion, 19 months before the onset of training. At the onset of training, this patient exhibited a WISCI score equal to 4. After 9 months of training, P4 displayed a remarkable partial functional recovery, reaching a final WISCI score of 13, and switching from an ASIA A to ASIA C classification. This patient did not need to use any body support system or even the non-invasive BMI system to walk autonomously. Thanks to her extraordinary functional recovery, she only required a regular walker for negotiating a variety of terrains and walks across multiple outside locations in the city of Beijing. We were able to follow this patient for several extra months after the conclusion of the 9-month training period and verified that her functional recovery remained stable following the end of the protocol.

Surprisingly, even a patient with 20 years of complete paraplegia (P11), who spent a great deal of this period bedridden, was able to recover a considerable degree of autonomous mobility (Initial WISCI=1; Final WISCI=9), using crutches to walk, and transition from ASIA A to C in 5 months.

Overall, we did not notice any major differences in tactile sensitivity, including fine and crude touch and pin prick sensation, between the experimental and the control group. As discussed below, this null result may be a direct consequence of the lack of haptic feedback in the modified WANR protocol employed during the present study.

### 3.3. Brain Imaging Analysis

Having demonstrated a significant level of patient motor recovery after 9 months of training, we next investigated the impact of the original SCI, as well as training with the WANR protocol, on our patients’ brain structure and function. This was done with structural and functional MRI.

#### 3.3.1. Structural MRI Results

##### 3.3.1.1. Impact of SCI on cortical thickness

As a representative sample of this analysis, Figure 8 illustrates the impact of spinal cord lesions, ranging from 15 to 251 months prior to onset of training, in a subset of six of our patients, as measured by structural MRI. In these plots, one can identify a widespread reduction in cortical thickness, represented by the different tones of dark blue, that extends well beyond the primary motor and somatosensory cortical areas in each patient, independent of the lesion time. In fact, even in the case with the shortest lesion time of our sample (P5, 15 months), one could already identify an important reduction in cortical thickness taking place not only in frontal and temporal cortical areas, but also, in a quite surprising way, spreading throughout multiple visual cortical regions in the occipital lobe and in the occipital-parietal-temporal borders. Similar patterns, with different spatial extents, were observed in patients who suffered lesions at 19 (P4), 27 (P8), 64 (P14), 156 (P7) and 251 months (P3) prior to the onset of training. In each of these images, different tones of blue indicate a reduction in cortical thickness, while colors ranging from yellow to orange and red represent increases in cortical thickness. For comparison purposes, the normal distribution of cortical thickness across a healthy human brain is also depicted at the top of the figure.

**Figure 8.**
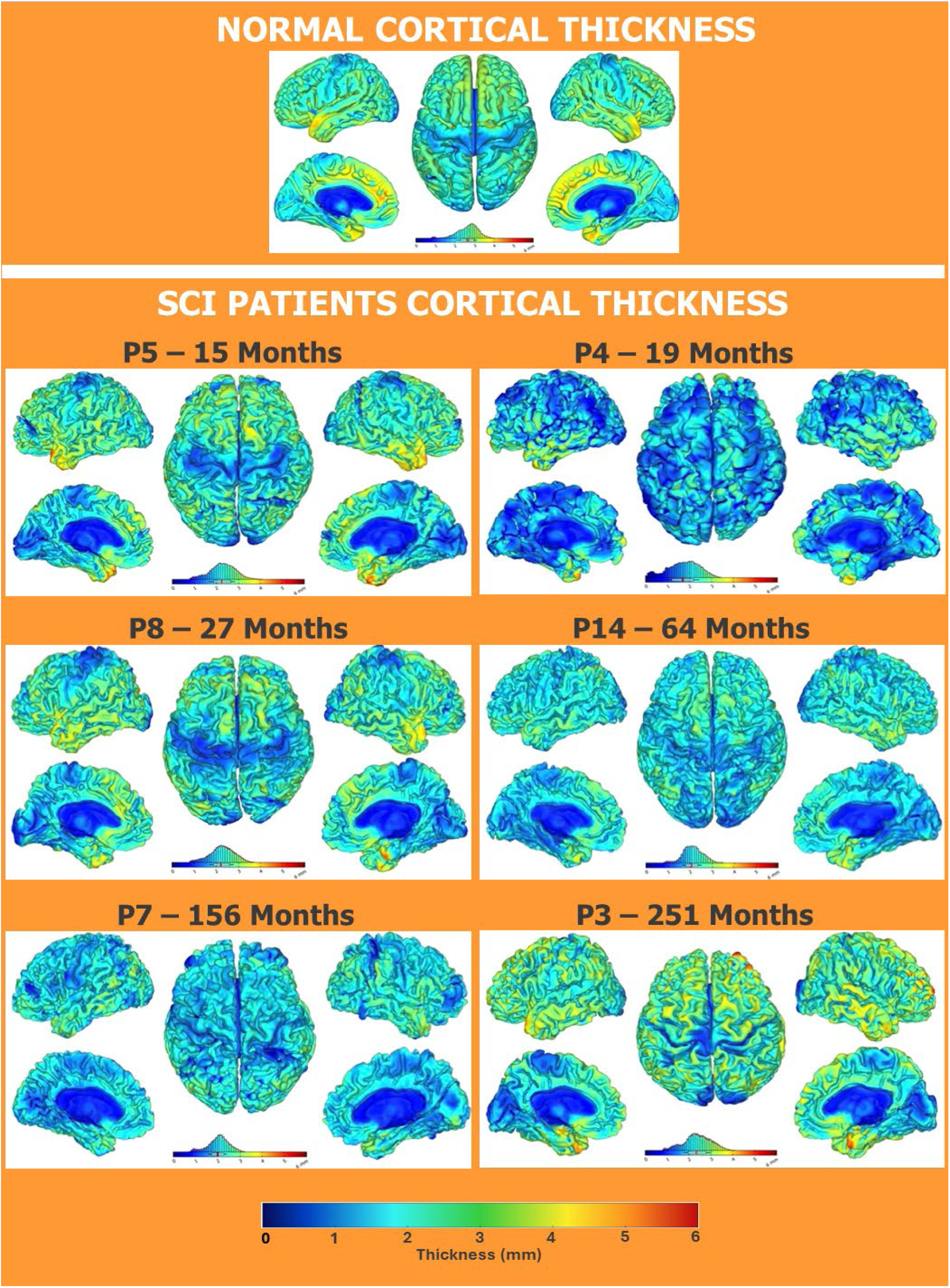
Comparison between normal cortical thickness distribution (top) and the equivalent images obtained for six SCI patients included in our study with lesion times ranging from 15-251 months: Patient **P5**, 15 months; Patient **P4**, 19 months; Patient **P8**, 27 months; Patient **P14**, 64 months; Patient **P7**, 156 months and Patient **P3**, 251 months. Dark blue color indicates a potential reduction in cortical thickness. As early as 15 months after the original SCI (P5), a reduction in cortical thickness can be identified in the frontal, parietal and even in multiple visual areas in the occipital lobe. With a little longer SCI lesion time of 19 months (P4), we can identify a very widespread reduction in cortical thickness, extending over much of the neocortex, and including the temporal lobe. Such a reduction in cortical thickness was very prominent all over the visual cortical areas of the occipital-parietal-temporal transition areas. Widespread reduction in cortical thickness was also observed in patients with longer SCI, but establishing a potential correlation between SCI time and spatial distribution of cortical atrophy was limited in our study. This happens due to technical limitations (see Methods) that made it impossible to quantify the extent of the original SCI in our patients. This may partially explain why the patient with the oldest SCI in our sample, 251 months (P3) did not necessarily exhibit the largest, or the more spatially widespread reductions in cortical thickness. This may be explained by the possibility that patient P3 had a much smaller SCI than patients with shorter SCI time. A colored scale bar displayed at the bottom of the figure denotes the thickness in mm; the colder the color, the thinner the cortical area.

To further illustrate this cortical atrophy effect, Figure 9 details the distribution of cortical thickness of patient P4 when compared to normal values for the 68 different cortical areas analyzed. Red dots in this bar graph depict reductions equal to or smaller than 2.5 standard deviations (SDs) from the normal average cortical thickness for a given cortical area. Meanwhile, purple dots depict reductions between 1-2.5 SD, and light blue dots indicate reduction or increase in cortical thickness on the order of 1 SD.

**Figure 9.**
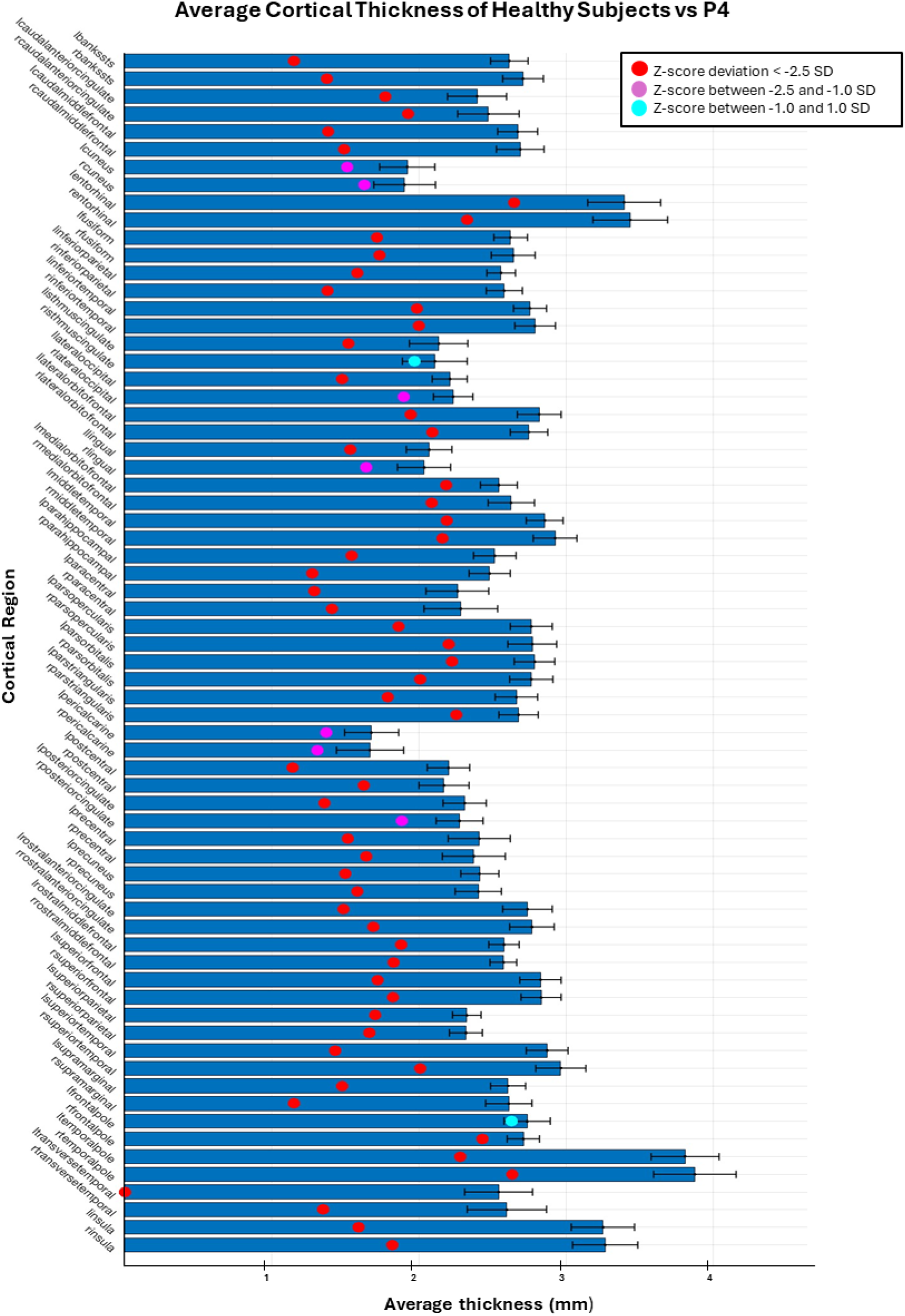
Distribution of cortical thickness of patient P4 when compared to normal values for 68 different cortical areas. Red dots depict reductions equal to or smaller than 2.5 standard deviations (SDs) from the normal average cortical thickness for a given cortical area. Purple dots depict reductions between 1-2.5 SD, and light blue dots indicate reductions or increases in cortical thickness in the order of 1 SD. The widespread distribution of red dots in this bar graph further confirms that an SCI, even after a relatively short period of time (19 months), induces a widely distributed reduction in cortical thickness across frontal, temporal, parietal, and occipital cortical areas.

The widespread spatial distribution of red dots in this bar graph further confirms that an SCI, even after a relatively short period of time (19 months), can induce a widely distributed reduction in cortical thickness. These include major reductions in multiple temporal, parietal and frontal loves, and again, an unexpected meaningful effect in some visual cortical areas in the occipital, parietal and temporal lobes.

The overall impact of the SCI in terms of cortical thickness for a sample of our cohort of SCI patients (n=16) can be appreciated by the data included in Tables 4 and 5. While Table 4 displays both the percentage of cortical thickness reduction and the absolute tissue loss in millimeters for the top 20 cortical areas prior to the initiation of the WANR protocol, Table 5 contains the same values for cortical lobes, averaged across both hemispheres, as well two highlighted gyri (pre and postcentral), one lobule (paracentral), and cortical regions of particular interest (left and right insula).

**Table 4.**
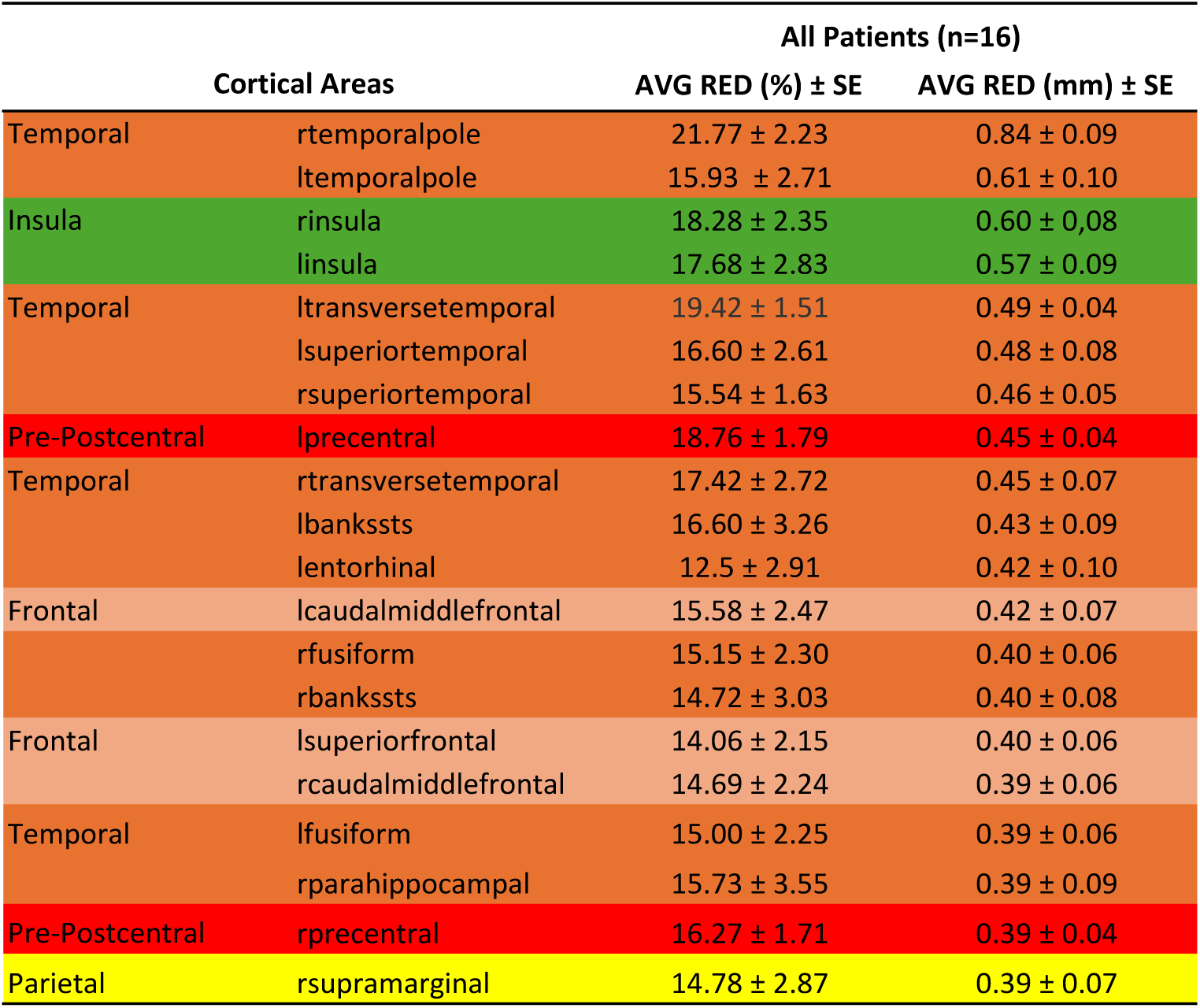
Top 20 cortical areas that exhibited the greatest reduction in cortical thickness (AVG±SE; measurements both in terms of percentage of reduction, when compared to values obtained from a pool of healthy subjects, and in mm, for a subset of 16 of the 18 patients enrolled in our clinical trial, prior to the onset of training with the WANR protocol. Areas are ranked by AVG RED (mm). Cortical areas in the temporal lobe areas are shown in brown; the insulas are shown in green; the combined pre and postcentral gyri are shown in light red; frontal areas in salmon; and parietal areas are shown in yellow. For the designation of each cortical area, the letter “l” depicts the left hemisphere, and “r” the right hemisphere.

**Table 5.**
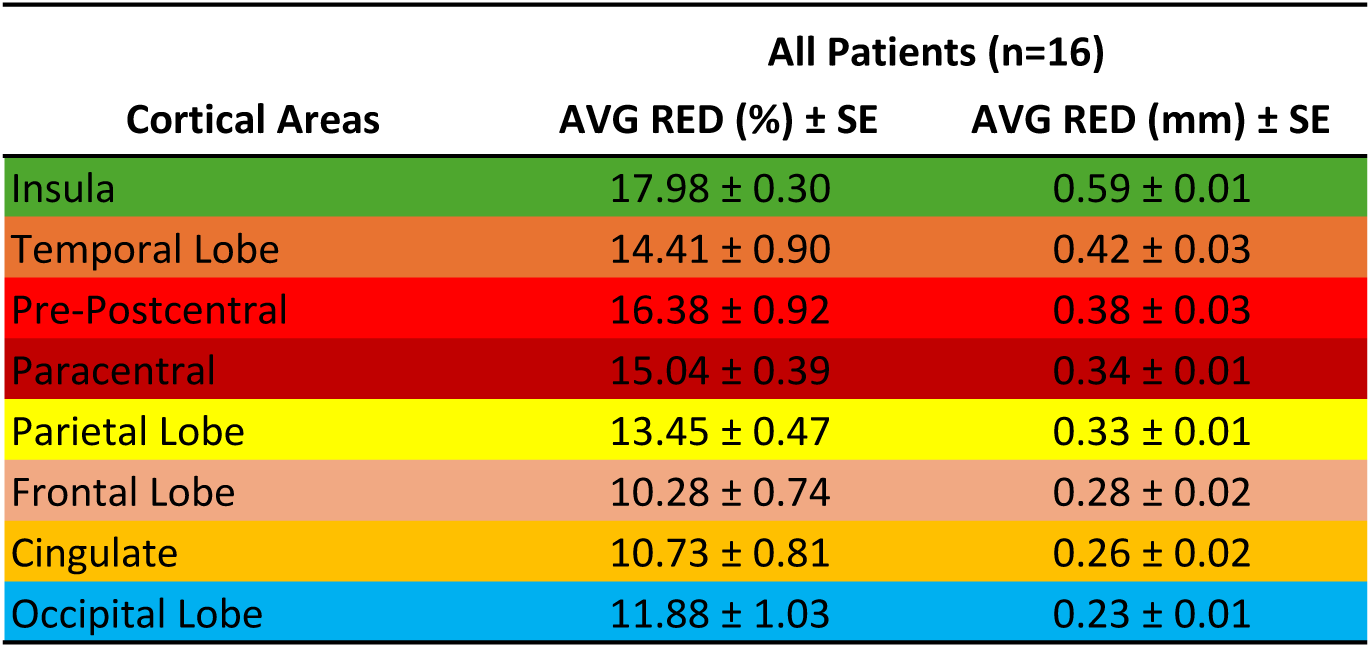
Global atrophy of the two brain hemispheres areas (AVG±SE; measurements both in terms of percentage of reduction, when compared to values obtained from a pool of healthy subjects, and in mm, for a subset of 16 of the 18 patients enrolled in our clinical trial, prior to the onset of training with the WANR protocol. Areas are ranked by AVG RED (mm). The insulas are shown in green; cortical areas in the temporal lobe areas are shown in brown; the combined pre and postcentral gyri are shown in light red; paracentral areas are shown in dark red; parietal areas are shown in yellow; frontal areas in salmon; cingulate lobe areas are shown in orange and occipital lobe are shown in blue.

| All Patients (n=16) |  |  |
| --- | --- | --- |
| Cortical Areas | AVG RED (%) ± SE | AVG RED (mm) ± SE |
| Insula | 17.98 ± 0.30 | 0.59 ± 0.01 |
| Temporal Lobe | 14.41 ± 0.90 | 0.42 ± 0.03 |
| Pre-Postcentral | 16.38 ± 0.92 | 0.38 ± 0.03 |
| Paracentral | 15.04 ± 0.39 | 0.34 ± 0.01 |
| Parietal Lobe | 13.45 ± 0.47 | 0.33 ± 0.01 |
| Frontal Lobe | 10.28 ± 0.74 | 0.28 ± 0.02 |
| Cingulate | 10.73 ± 0.81 | 0.26 ± 0.02 |
| Occipital Lobe | 11.88 ± 1.03 | 0.23 ± 0.01 |

Tables 4 and 5 are supplemented by Supplementary Table 1, which ranks all 68 cortical areas surveyed in our structural MRI data in terms of both percentage of cortical thickness reduction and absolute reduction in millimeters, based on data from seven patients from the NR group and nine from the original WANR group. All structural MRI data were collected prior to the onset of training.

When inspecting Tables 4 and Supplementary 1, one notices that, surprisingly, the right temporal pole leads the ranking, in terms of percentage of cortical thickness reduction, with 21.7±2.23%, corresponding to a very significant loss of 0.84±0.09mm in cortical tissue. Meanwhile, the left temporal pole, ranked in 2nd position, exhibited a loss of 16.0±2.7% or 0.6±0.1mm. Unexpectedly, the right and left insula occupied the 3^rd^ and 4^th^ position, with a loss of 18.3±2.4% or 0.60±±0.1mm (right) and 17.7±2.8% or 0.6±0.1mm (left) respectively.

The left transverse temporal gyrus or Heschl’s gyrus with 19.4±1.51% (0.49±0.04mm) occupied the 5^th^ position in the ranking. For comparison, the right transversal gyrus exhibited a loss of 17.4±2.7%, which meant a reduction in cortical thickness of 0.45±0.1mm. Other important reductions in cortical thickness in the temporal lobe were observed in the following locations: the banks of the left superior temporal sulcus (16.6±3.3% or 0.4±0.1mm), the left superior temporal gyrus (16.6±2.6% or 0.48±0.1mm), the right (15.7±3.6% or 0.4±0.1mm) and left (15.3±3.1% or 0.4±0.1mm) parahippocampal gyri, and the right (15.2±2.3% or 0.4±0.1mm) and left (15.0±2.3% or 0.4±0.1mm) fusiform gyri.

The left precentral gyrus, where the primary motor cortex is located, appeared only in 8th place in our ranking with a cortical thickness reduction of 18.7±1.8% (0.45±0.04mm), while the right precentral gyrus was positioned in the 19th place with a loss of 16.3±1.7% (0.4±0.04mm). Although one would have originally suspected that the paracentral lobules and the postcentral gyri would also have experienced some of the largest losses in cortical thickness in SCI patients, they did not make the top ten areas in our ranking, when the percentage of cortical tissue reduction was considered. While the right paracentral lobule occupied only the 26^th^ position in the ranking (15.4±2.5% or 0.35±0.06mm), the left paracentral lobule was placed in the 30^th^ position (14.7±2.5% or 0.33±0.06mm). Meanwhile, the left postcentral gyrus was ranked 24^th^, with a 16.2±2.4% (0.4±0.05mm) loss in cortical thickness, while the right postcentral gyrus experienced a 14.3±1.6% or 0.3±0.03 mm reduction in cortical thickness (38^th^ position).

Conversely, much more surprisingly was the high-ranking position of the right and left insula (3^rd^ and 4^th^), which experienced a loss of 18.3±2.4% or 0.60±±0.1mm (right) and 17.7±2.8% or 0.6±0.1mm (left) respectively.

The left pericalcarine cortical region exhibited the largest average loss in cortical thickness in the occipital lobes at 17.0±2.0% (0.3±0.03mm) reduction in cortical thickness overall. In addition, the left lingual gyrus (44^th^ place in the ranking by percentage) experienced a loss of 14.0±1.7% or 0.3±0.04mm.

Regarding the frontal lobe, the main reduction in cortical thickness was observed in the left and right caudal middle frontal gyri. While the former experienced a mean reduction of 15.6±2.5% (0.42±0.07mm), the latter underwent a loss of 14.7±2.2% or 0.4±0.06mm.

Other cortical areas in the parietal lobe also exhibited significant losses in thickness. For example, the right (14.8±2.9% or 0.40±0.1mm) and left (14.2±2.2% or 0.4±0.1mm) supramarginal gyrus, the right (14.2±2.2% or 0.3±0.1mm) and left (13.6±2.1% or 0.33±0.1mm) precuneus, the right inferior parietal (14.1±2.5% or 0.4±0.1mm) and the right superior parietal (14.0±1.6% or 0.3±0.04mm) gyri.

When we considered cortical thickness reduction as a function of the different lobes in both hemispheres (Table 5), minus the paracentral lobule and pre- and postcentral gyrus which were lumped together, we observed that the insula led the loss in cortical thickness (18.00±0.3% or 0.60±0.01mm), followed by the combination of the pre- and postcentral gyri (16.4±0.92% or 0.38±0.030), the paracentral lobules (15.04±0.4 or 0.34±0.01mm), the temporal lobes with an average of 14.4±0.9% (0.42±0.03mm), the parietal (minus the postcentral gyrus (13.5±0.5% or 0.33±0.01mm), the occipital (11.9±1.03% or 0.23±0.01mm) lobes, the cingulate with an average of 10.7±0.8% (0.3±0.02mm), the frontal lobe (minus the precentral gyrus) (10.3±0.74% or 0.3±0.02). No major differences were observed between left and right hemispheres when data from each of the cortical lobes were compared.

Once we characterized the main features of the cortical atrophy induced by SCI, we compared our findings with the normal distribution of cortical thickness obtained from a sample of healthy brains. Figure 10A-C illustrates the results of this comparison which proved to be statistically significant (*p<0.001*, *p<0.05* and *p<0.05*, respectively). As such, this analysis confirmed that multiple cortical, and even subcortical structures, were shown to be affected by SCI, leading to a significant reduction in their volumes when compared to normal healthy brains.

**Figure 10.**
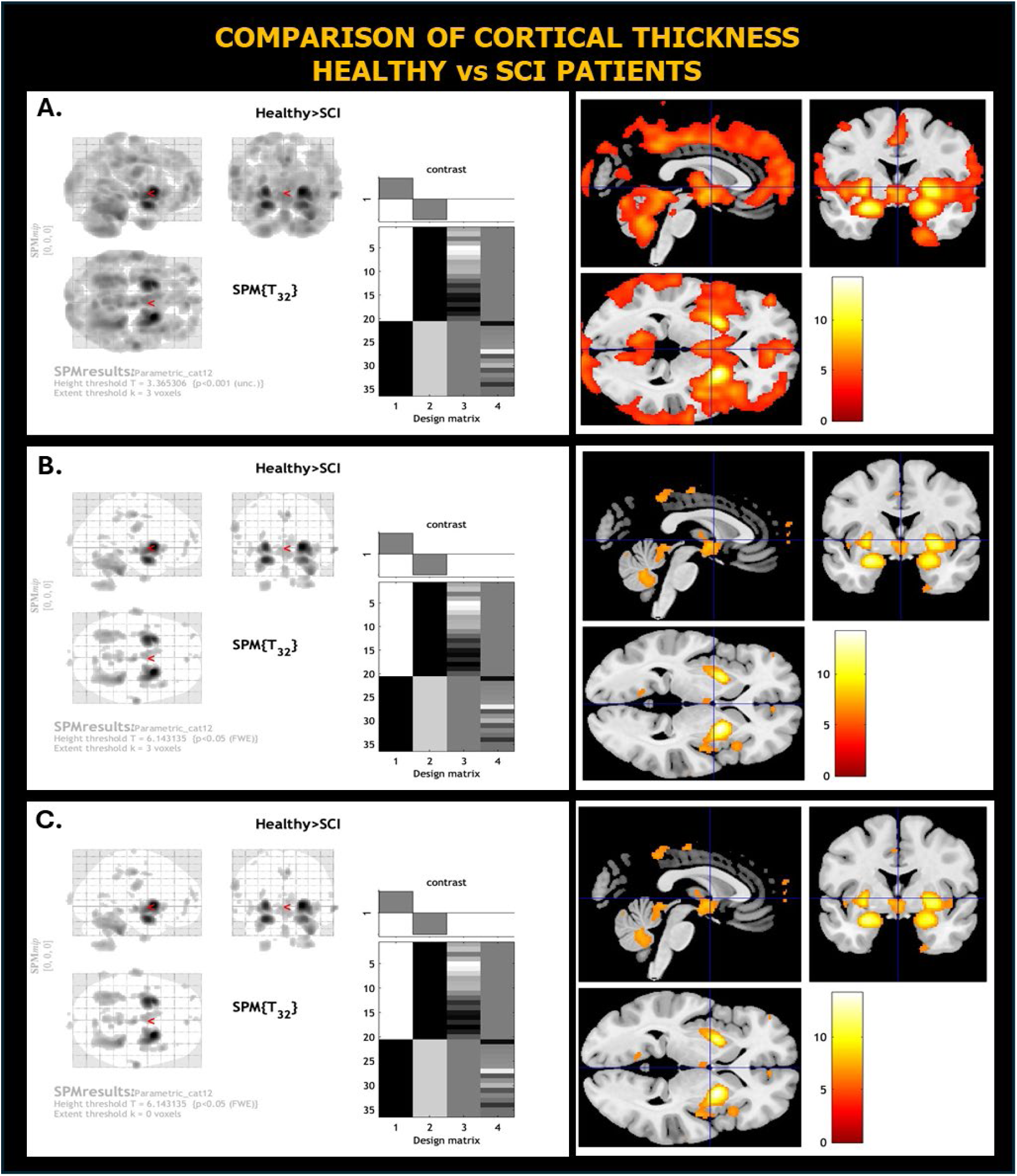
Statistical analysis of cortical thickness and subcortical structure volume differences between brains of healthy subjects and 18 patients included in this protocol, prior to the onset of training. (**A**) p<0.001, Height threshold T= 3.365306, Extent threshold k=3 voxels; (**B**) p<0.05, Height threshold T= 6.143135, Extent threshold k=3 voxels; (**C**) p<0.05, Height threshold T= 6.143135, Extent threshold k=0 voxels.

##### 3.3.1.2. Impact of WANR protocol in cortical thickness

The next step was to repeat the same analysis at the mid- (5 months) and final point (9 months) of the training protocol. As an example of our main findings, Figure 11 illustrates the major changes in cortical thickness observed in patient P4 after 9 months of training with the WANR protocol. By comparing the histogram of this figure with the one from Figure 9, one can see that a large number of cortical areas underwent a significant increase in cortical thickness after completion of the WANR protocol when compared to the onset of the clinical trial.

**Figure 11.**
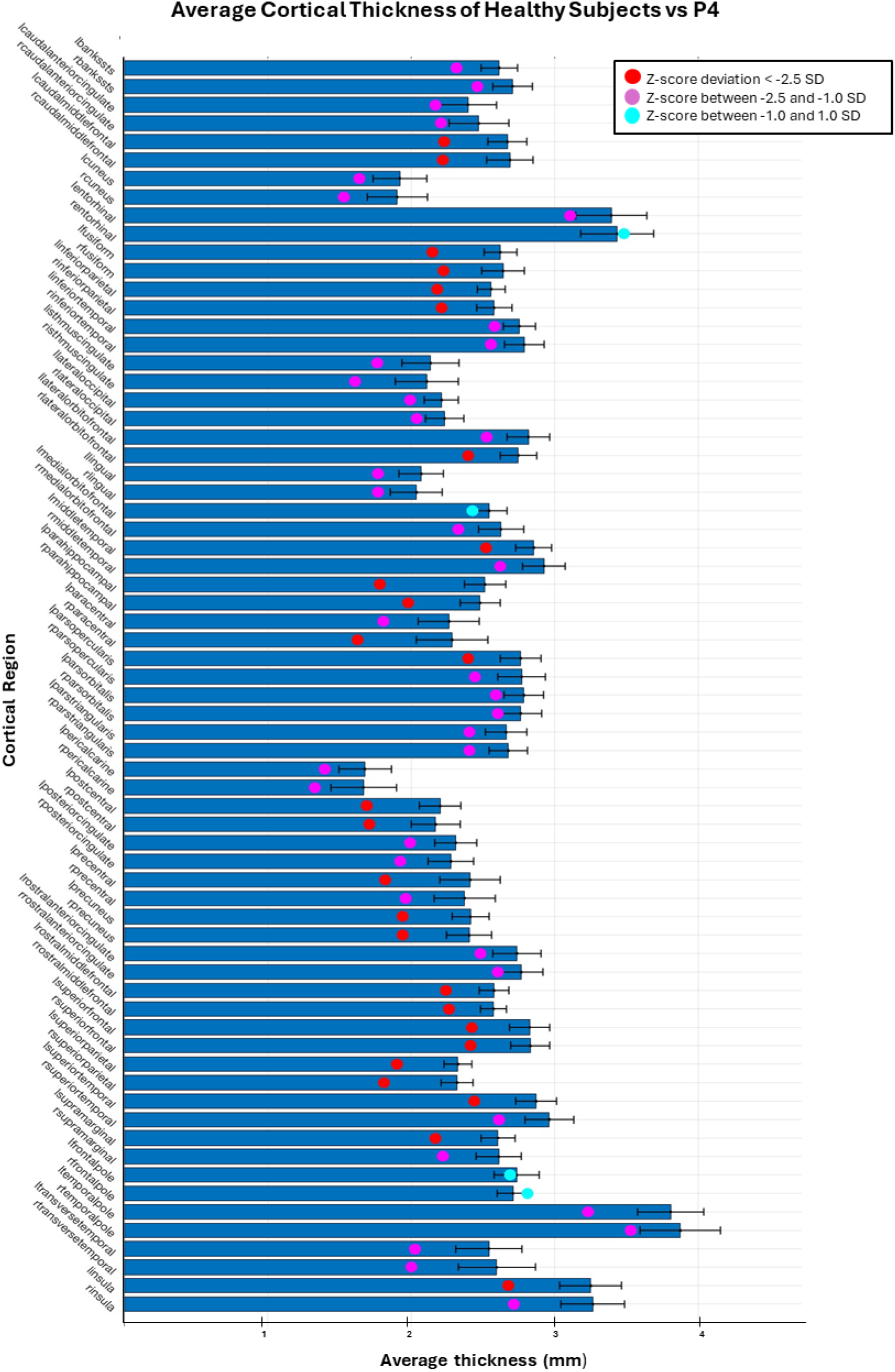
Distribution of cortical thickness (red, purple and light blue dots) of patient P4 after 9 months of training compared to normal values (dark blue bars) for 68 different cortical areas. Red dots depict reductions equal or smaller than 2.5 standard deviations (SDs) from the normal average cortical thickness for a given cortical area. Purple dots depict reductions between 1-2.5 SD, and light blue dots indicate reductions or increase in cortical thickness in the order of 1 SD. When compared to Figure 9 (pre-training), it is possible to see that a large number of cortical areas underwent a significant recovery in cortical thickness.

Figure 12 reveals in more detail the spatial evolution of the recovery in cortical thickness for patient P4 from the pre-training stage to 5 months, and then 9 months after onset of training, for both the left (Figure 12A) and the right (Figure 12B) hemispheres.

**Figure 12.**
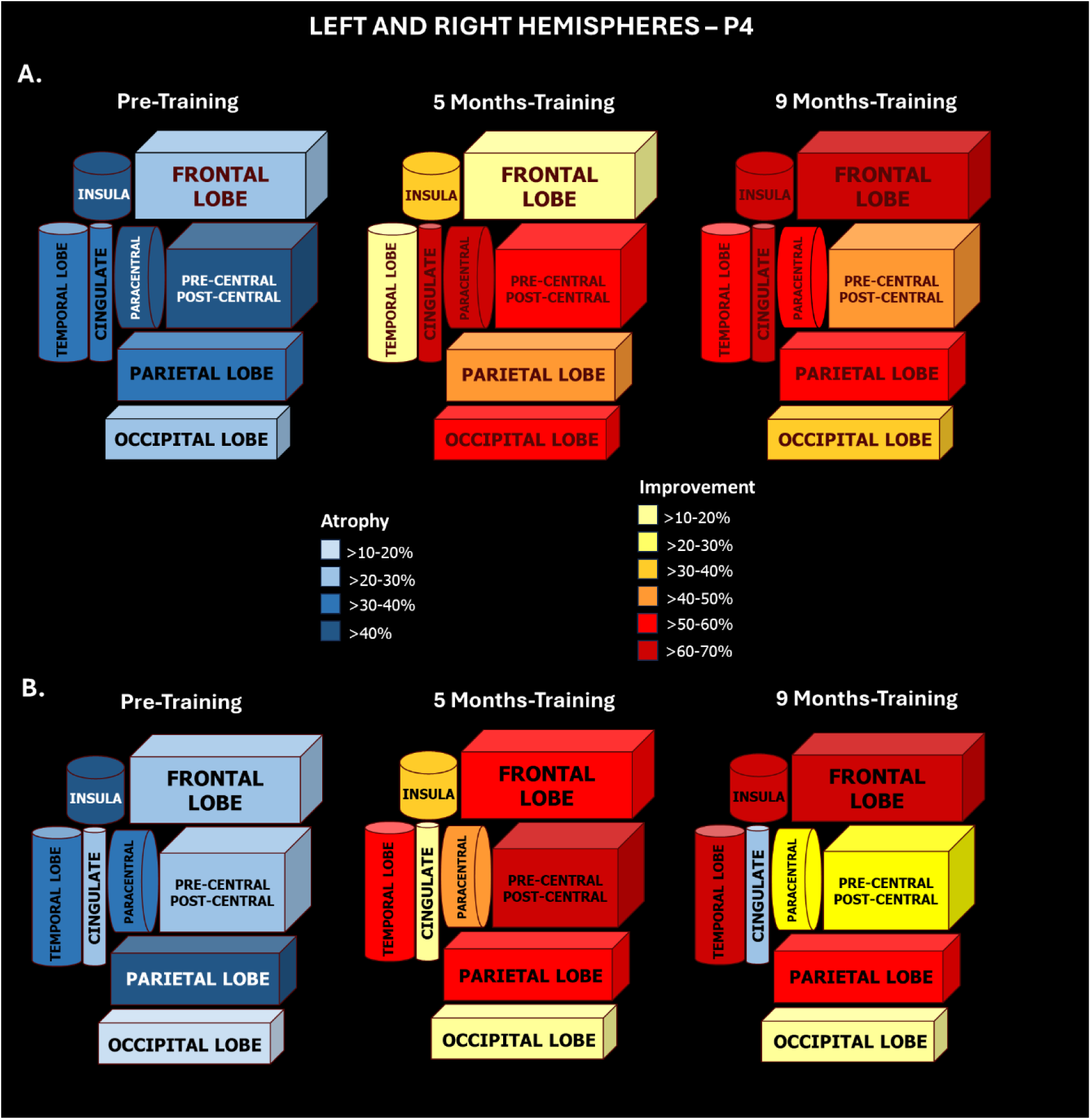
3D Graphic representation of spatial evolution of the recovery in cortical thickening for patient P4 in three different times: Pre-training, 5 months and 9 months, in (**A**) left and (**B**) right hemispheres. Color code for the percentage of cortical atrophy and improvement, when compared to normal healthy controls, follows the scale. (**A**) <u>Pre-training</u>: Greatest levels of cortical atrophy are in the insula, paracentral and pre-postcentral (>40%; darkest blue), followed by temporal, parietal and cingulate (>30-40%; dark blue). The smallest losses in thickness were in the frontal and occipital lobes (>20-30%; light blue). <u>5-months training</u>: The greatest improvement was in the cingulate and paracentral (>60-70%; dark red), followed by the occipital and pre-postcentral (>50-60%; light red), the parietal (>40-50%; dark orange), and the insula (>30-40%; light orange). The frontal and temporal lobes had the smallest improvements (>10-20%; light yellow). <u>9-months training</u>: The greatest improvements were in insula, frontal and cingulate (>60-70%; dark red), followed by paracentral, temporal and parietal (>50-60%; light red) and pre-postcentral gyri (>40-50%; dark orange). The occipital lobe had the lower recovery (>30-40%; light orange). **(B)** <u>Pre-training</u>: The greatest levels of cortical atrophy are in the insula and parietal (>40%; darkest blue), followed by paracentral and temporal (>30-40%; dark blue) and pre-postcentral, frontal and cingulate (>20-30%; light blue). The smallest loss occurred in the occipital lobe (>10-20%; lightest blue). <u>5-months training</u>: The greatest improvement was in pre-postcentral gyri (>60-70%; dark red), followed by the temporal, parietal and frontal lobes (>50-60%; light red), the paracentral lobule (>40-50%; dark orange), the insula (>30-40%; light orange). The smallest improvements were in occipital and cingulate (>10-20%; light yellow). <u>9-months training</u>: The greatest improvements were in temporal, frontal and insula (>60-70%; dark red), followed by the parietal lobe (>50-60%; light red). A smaller improvement occurred in the pre-postcentral and paracentral (>20-30%; dark yellow), and in the occipital lobe (>10-20%; light yellow). An atrophy occurred in the cingulate (>20-30%; light blue).

Despite suffering an SCI only 19 months before enrollment in our protocol, patient P4 underwent one of the most significant losses in cortical thickness of our experimental group. And yet, she experienced the best clinical recovery in terms of motor performance, moving from ASIA A to ASIA C after only 5 months of training, and reaching a WISCI of 13 at the end of training. The 3D graph in Figure 12A indicates that, at the onset of the trial, the worst level of cortical atrophy in the patient’s left hemisphere was located in the insula (a loss of 51.2%, or 1.7 mm in thickness), followed by the paracentral lobule (43.1% or 1.0 mm), and the pre- and postcentral gyri (42.6% or 1.0mm), and then the temporal (34.5% or 1.0mm), parietal lobe (36.5% or 0.9mm), and cingulate cortex (35.5% or 0.9mm). Conversely, the smallest losses in thickness occurred in the frontal (27.7% or 0.8mm) and occipital (24.6%, 0.50mm) lobes. Meanwhile, in the right hemisphere (Figure 12B) the larger reduction in cortical thickness was located primarily in the insula (44.3% or 1.5mm) and the parietal lobe (41.2% or 1.0mm), followed by the paracentral (38.3% or 0.9mm), the temporal (36.8% or 1.0mm), the precentral and postcentral gyri (28% or 0.6mm), the frontal lobe (25.3% or 0.7mm), and the cingulate (21.2% or 0.5mm). The smallest loss occurred in the occipital lobe (17.5% or 0.3mm).

At the end of 5 months of training, this patient had already exhibited a remarkable recovery in cortical thickness in both hemispheres. On the left, the percentage of recovery was larger in the cingulate cortex (69.2% or 0.6mm), and the paracentral lobule (67.3% or 0.7mm), followed by the occipital lobe (56% or 0.3mm), the precentral and postcentral gyri (55.3% or 0.5mm), the parietal lobe (47.6% or 0.5mm), and the insula (37% or 0.6mm). A smaller, but still very significant recovery was also measured in the frontal lobe (20% or 0.4mm), while the temporal lobe showed the smallest recovery in percentage of cortical thickness (12.5% or 0.3mm). Meanwhile, in the right hemisphere, the percentage recovery in cortical thickness was more intense in the precentral and postcentral gyri (65.3% or 0.4 mm) and temporal lobe (57.8% or 0.6mm). The parietal lobe (55.3% or 0.6mm), the frontal lobe (47.8% or 0.35mm), and the paracentral lobule (44% or 0.4mm) were the next in terms of percentage of recovery. Then came the insula (39.1% or 0.6mm), the occipital lobe (19% or 0.1mm), and the cingulate (13.7% or 0.3mm).

By the time P4 reached the 9^th^ month of training, the center of her recovery in the left hemisphere included the insula (65.3% or 1.1mm), the frontal lobe (62.9% or 0.5mm), the cingulate cortex (62.2% or 0.6mm), and the paracentral lobule (53.3% or 0.5mm). This was closely followed by the temporal lobe (51.1% or 0.6mm) and the parietal lobe (50.5% or 0.5mm). The precentral and postcentral gyri (42.6% or 0.4mm) and the occipital lobe (38.4% or 0.2mm) exhibited lower, but still important recoveries. Meanwhile, in the right hemisphere, the highest percentage of cortical thickness recovery was centered on the temporal lobe (68.5% or 0.7mm), the frontal (62.9% or 0.4 mm), insula (62% or 0.9mm) and parietal (52.1% or 0.6mm) lobes. A much smaller percentage of change was seen at the pre-postcentral (29.4% or 0.2mm), paracentral lobule (24.6% or 0.2mm), occipital lobe (10.6% or 0.1mm) and the cingulate (-27.3% or 0.2mm).

By comparing the spatial distributions at 5 and 9 months, as well as the left and right hemispheres, one can see that the recovery in cortical thickness in patient P4 was widely distributed over most of her cortex. This process unfolded through a very dynamic spatio-temporal process over the 9 months of training. During this period, progressive improvements in some regions (like the insula) could coexist with subsequent decreases in cortical thickness (like the precentral and postcentral gyri in this case). Very likely, this dynamic process reflected a complex mix of both a plastic recovery in cortical thickness in some cortical areas, compensated by reductions in others that continued to degenerate as a result of the original SCI.

Figure 13 displays the patterns of cortical thickness observed in the same six patients of Figure 8 after 9 months of training. Patients P8 and P5 exhibited a major response to the training protocol in terms of cortical thickness increase. In both patients, as well as in patient P4, who exhibited the best clinical motor recovery of all 14 patients, we noticed robust increases in cortical thickness in frontal, parietal, and temporal areas, but also all over the occipital pole. As in the case of the cortical thickness reduction effect caused by the original SCI, the recovery extended into a variety of visual cortical areas. Even in patients who suffered an SCI many years ago, like P14 (64 months), P7 (156 months), and P3 (251 months), we could identify some level of increase in cortical thickness. However, such a recovery became much more restricted, in terms of both magnitude and spatial distribution in patients whose SCI had occurred 11-20 years prior to training onset. Indeed, a more constrained spatial recovery in cortical thickness was also associated with poorer clinical motor outcomes. For instance, patients P5, P7 and P3 were not able to evolve from ASIA A to C and exhibited lower WISCI scores than patients P8, P4, and P14 who not only reached higher WISCI values, but also transitioned to ASIA C at the end of the protocol. A quantitative description of the recovery in cortical thickness observed for the entire WANR group, from 5 to 9 months of training, as well as a comparison with the data obtained from the control group is described in Tables 6-13. Initially, Tables 6 and 8 rank in order the top 20 cortical areas with the highest recovery in cortical thickness for the WANR group and the respective values of cortical thickness for those areas obtained from the NR group. Supplementary Tables 2 and 3 exhibited the values for all 68 cortical areas analyzed. Meanwhile, Tables 7 and 9 describe the improvement in cortical thickness for the cortical lobes, the paracentral lobule, the precentral and postcentral gyri, and the insula region.

**Figure 13.**
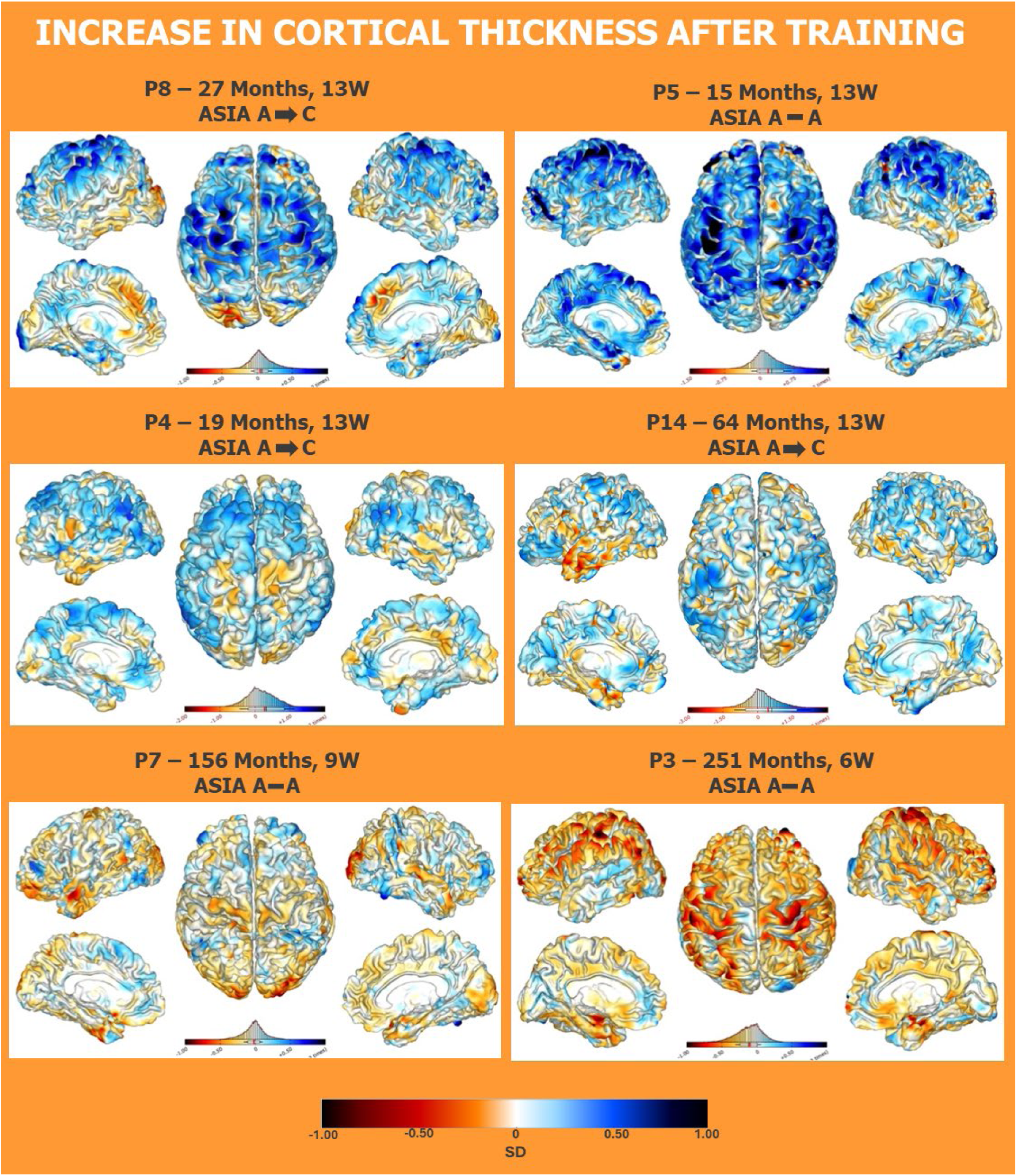
Structural MRIs of the same six patients shown in Figure 8 after 9 months of training. Colored scale bar displayed at the bottom of the figure denotes the values of standard deviation (from -1 to 1). In these plots, now the distinct tones of blue represent different levels of cortical thickness increase, with dark blue indicating the maximum increase, and light blue the smallest.

**Table 6.**
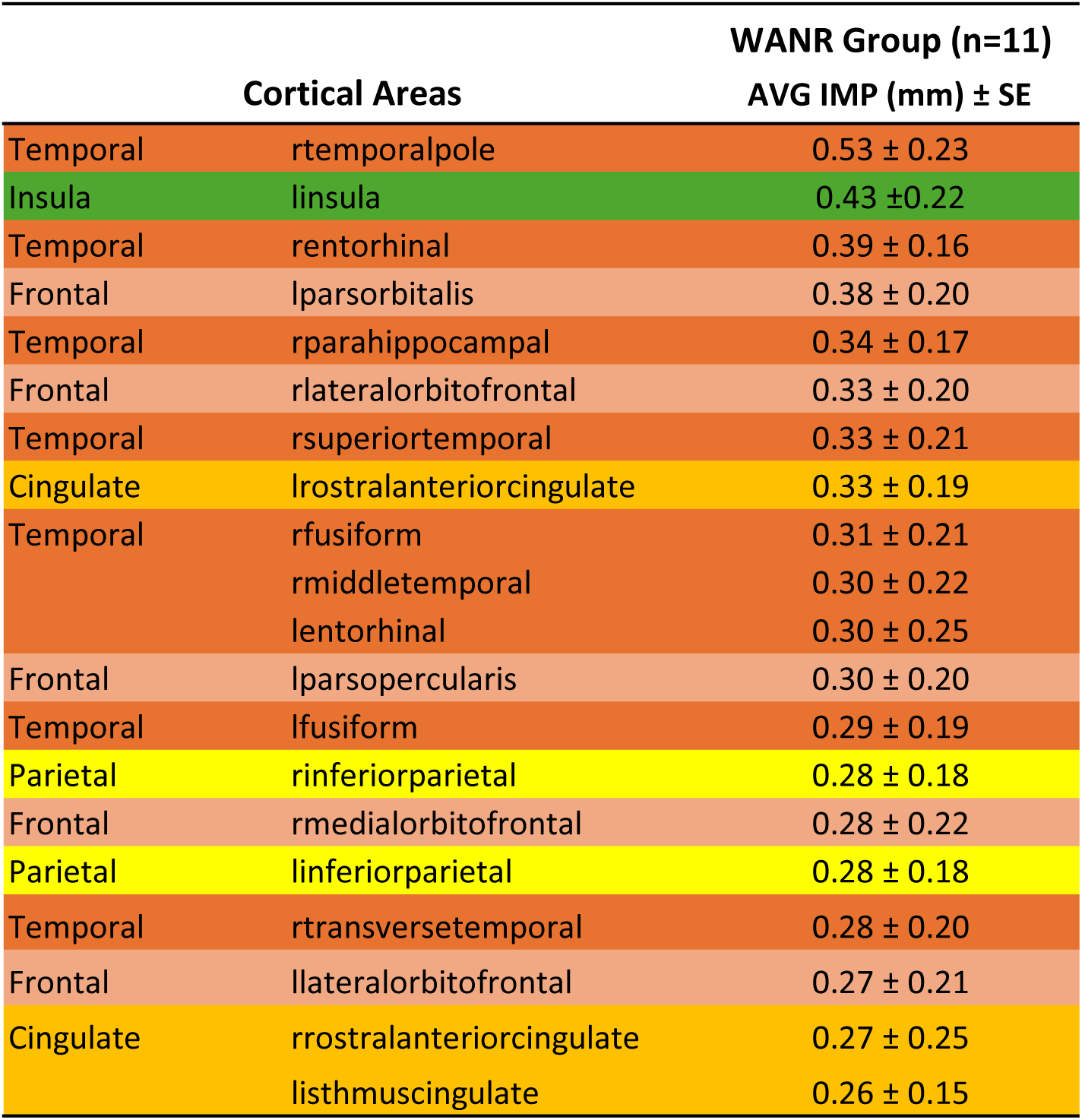
Top 20 cortical areas that exhibited the greatest improvement in cortical thickness (AVG±SE; measurements in mm) in 5 months for the WANR group (n=11). Cortical areas in the temporal lobe are shown in brown; the insulas are shown in green; frontal areas in salmon; cingulate lobe areas are shown in orange; and parietal areas are shown in yellow. For the designation of each cortical area, the letter “l” depicts left hemisphere, and “r” the right hemisphere.

**Table 7.**
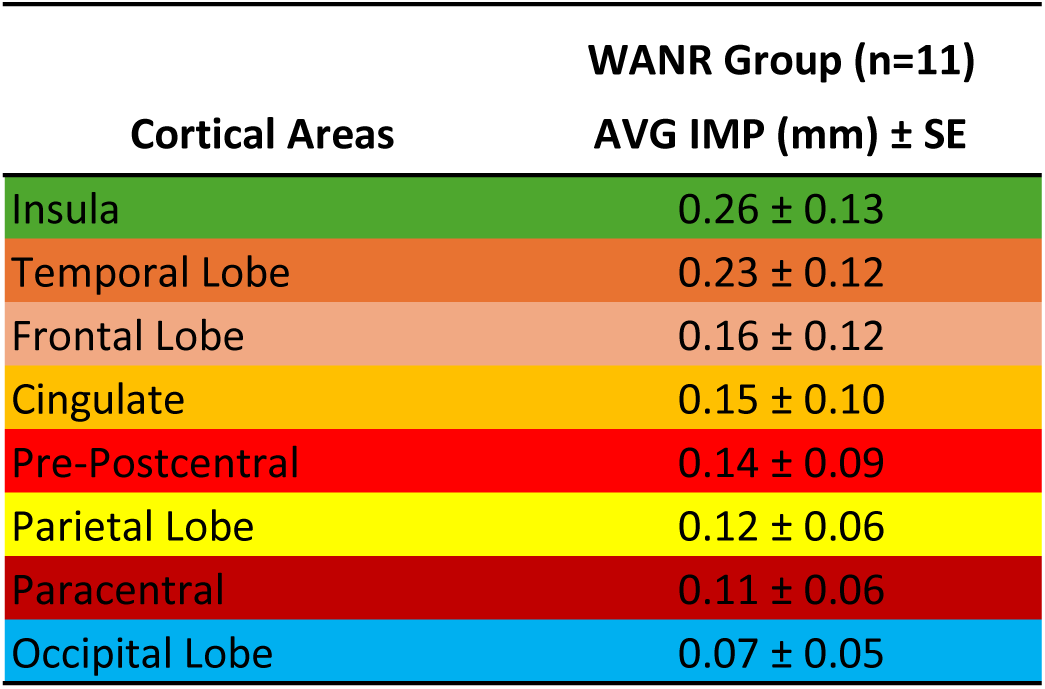
Global improvement of the two brain hemispheres areas in 5 months (AVG±SE; measurements in mm) for WANR group (n=11). The insulas are shown in green; cortical areas in the temporal lobe are shown in brown; frontal areas in salmon; cingulate lobe areas are shown in orange; the combined pre and postcentral gyri are shown in light red; parietal areas are shown in yellow; paracentral areas are shown in dark red; and occipital lobe areas are shown in blue.

|  |  | WANR Group (n=11) |
| --- | --- | --- |
| Cortical Areas |  | AVG IMP (mm) ± SE |
| Insula |  | 0.26 ± 0.13 |
| Temporal Lobe |  | 0.23 ± 0.12 |
| Frontal Lobe |  | 0.16 ± 0.12 |
| Cingulate |  | 0.15 ± 0.10 |
| Pre-Postcentral |  | 0.14 ± 0.09 |
| Parietal Lobe |  | 0.12 ± 0.06 |
| Paracentral |  | 0.11 ± 0.06 |
| Occipital Lobe |  | 0.07 ± 0.05 |

**Table 8.**
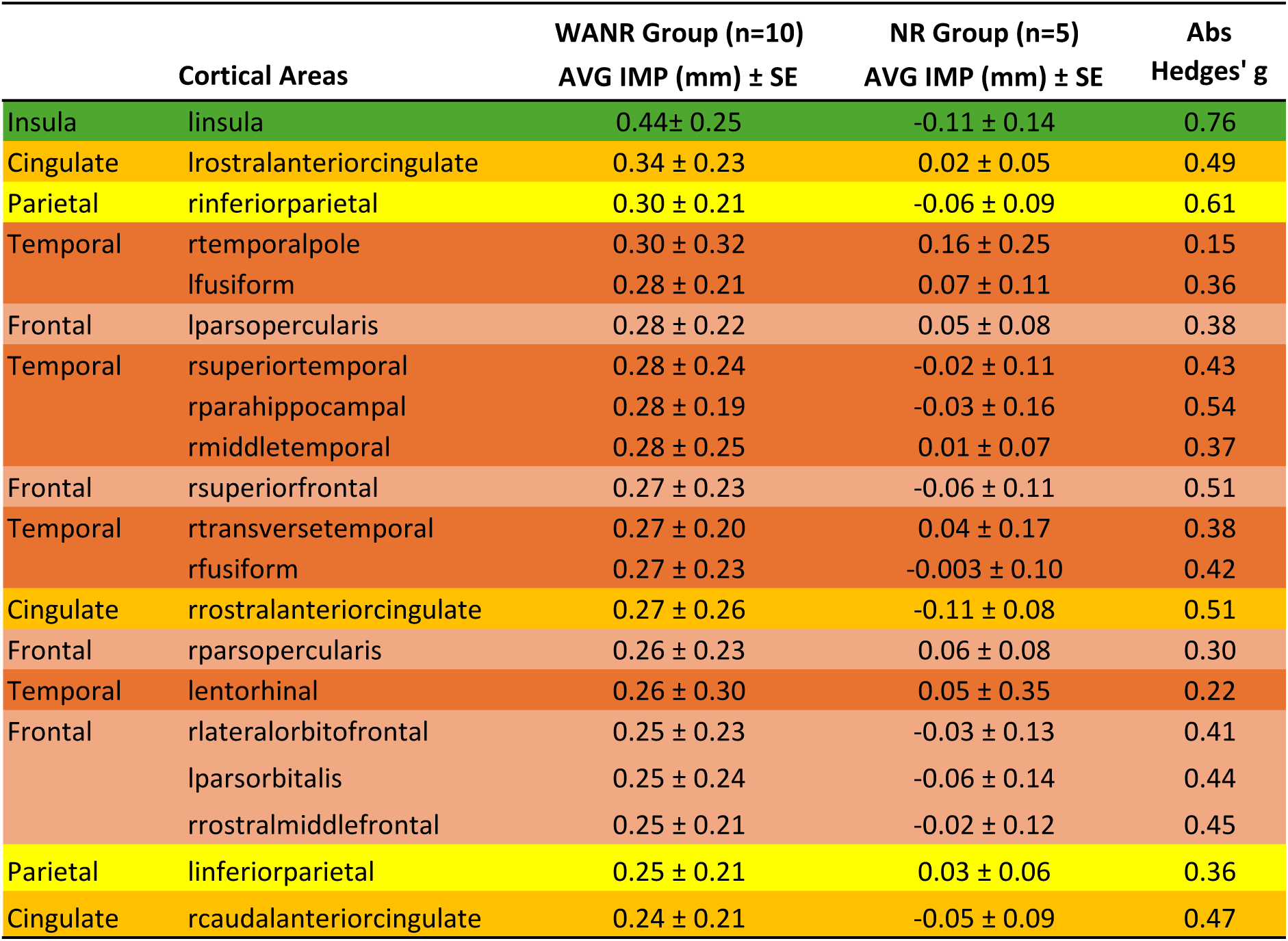
Top 20 cortical areas that exhibited the greatest improvement in cortical thickness in 9 months (AVG±SE; measurements in mm) for the WANR group (n=10), the values of the same areas for the NR group (n=5), and the Absolute Hedges’ g values for WANR x NR groups. Hedges’ g represents how many pooled standard deviations apart two group means are, with a small-sample bias correction. Its magnitude conveys the effect size (∼0.2 small, ∼0.5 medium, ∼0.8 large, ≥1.2 very large). Insula areas are shown in green; cingulate lobe areas are shown in orange; parietal areas are shown in yellow; temporal lobe areas are shown in brown; frontal lobe areas are shown in salmon. For the designation of each cortical area, the letter “l” depicts left hemisphere, and “r” the right hemisphere.

**Table 9.**
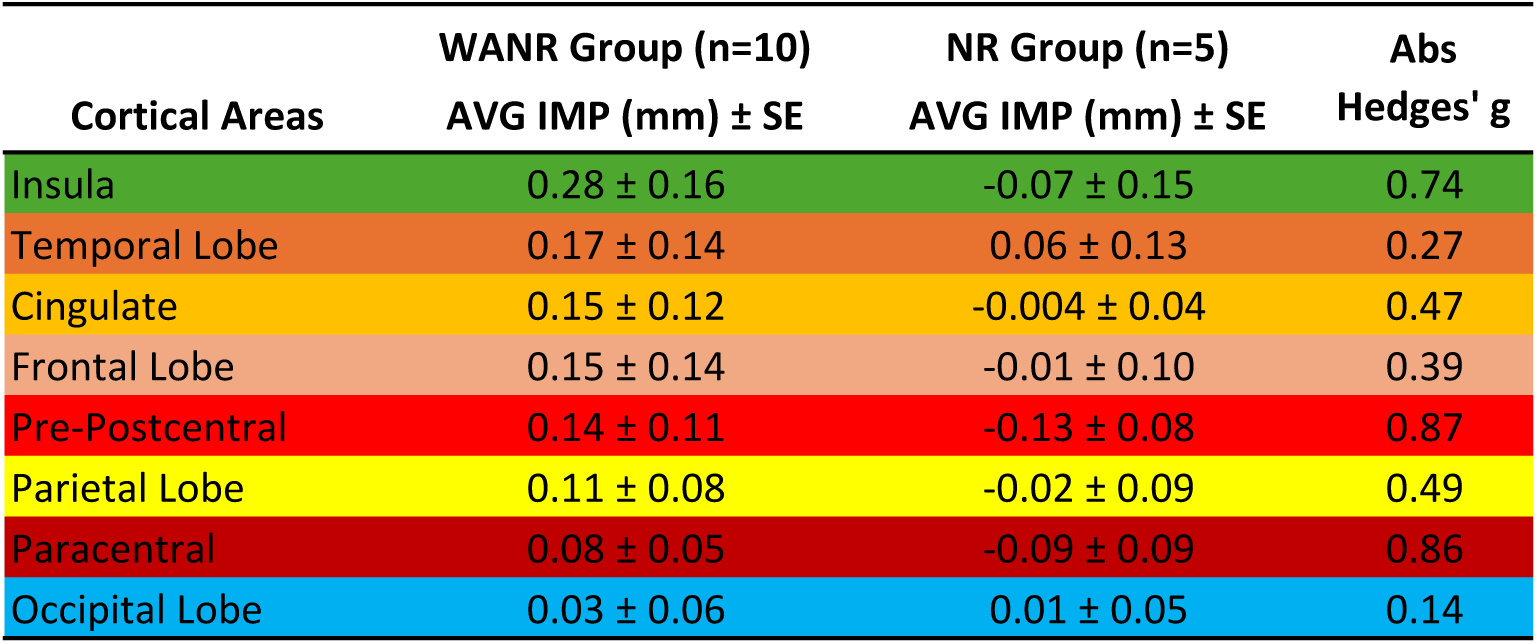
Global improvement of the two brain hemispheres areas for WANR and NR groups in 9 months (AVG±SE; measurements in mm). Comparison of the greatest improvement for WANR group (n=10 patients), the values of the same areas for NR group (n=5) and the Absolute Hedges’ g values for WANR x NR groups. Hedges’ g represents how many pooled standard deviations apart two group means are, with a small-sample bias correction. Its magnitude conveys the effect size (∼0.2 small, ∼0.5 medium, ∼0.8 large, ≥1.2 very large). Insula areas are shown in green; temporal lobe areas are shown in brown; cingulate lobe areas are shown in orange; frontal lobe areas are shown in salmon; the combined pre and postcentral gyri are shown in light red; parietal areas are shown in yellow; paracentral areas are shown in dark red; and occipital lobe areas are shown in blue.

| Cortical Areas | WANR Group (n=10)<br>AVG IMP (mm) ± SE | NR Group (n=5)<br>AVG IMP (mm) ± SE | Abs<br>Hedges' g |
| --- | --- | --- | --- |
| Insula | 0.28 ± 0.16 | -0.07 ± 0.15 | 0.74 |
| Temporal Lobe | 0.17 ± 0.14 | 0.06 ± 0.13 | 0.27 |
| Cingulate | 0.15 ± 0.12 | -0.004 ± 0.04 | 0.47 |
| Frontal Lobe | 0.15 ± 0.14 | -0.01 ± 0.10 | 0.39 |
| Pre-Postcentral | 0.14 ± 0.11 | -0.13 ± 0.08 | 0.87 |
| Parietal Lobe | 0.11 ± 0.08 | -0.02 ± 0.09 | 0.49 |
| Paracentral | 0.08 ± 0.05 | -0.09 ± 0.09 | 0.86 |
| Occipital Lobe | 0.03 ± 0.06 | 0.01 ± 0.05 | 0.14 |

Supplementary Tables 2 and 3 summarize the measurements of cortical thickness, obtained for the same 68 cortical areas, at both 5 and 9 months of training for 10 of the 14 patients of the WANR and NR groups.

By 5 months of training, patients of the WANR group exhibited a widespread increase in cortical thickness that was several times bigger than that of the NR group at 9 months. The highest increase in the WANR group was observed at the right temporal lobe (0.53±0.23mm) when compared to the value of the NR group (0.16±0.25mm). Moreover, while the left insula showed an increase of 0.43±0.22mm, the same structure displayed a reduction in thickness (-0.11±0.14mm) in the control group. By selecting an arbitrary threshold (0.20mm) to compare the two distributions, it was observed that 30 cortical areas out of 68 (or 44% of the total) of the WANR group exhibited an increase of cortical thickness equal or larger than this value. Meanwhile, only three cortical areas showed an increase of 0.2mm in the control group. Overall, the gain in cortical thickness of the WANR group at 5 months of training that was equal or above 0.2mm included: 11 cortical areas in the temporal lobe, nine in the frontal, four in the cingulate cortex, two in the parietal lobe, one in the occipital, one in the insula, and one in the left and one in the right precentral lobule.

Considering the entire WANR group at 9 months of training, a total of 28 cortical areas (41% of the total) still exhibited gains in cortical thickness above 0.2mm (Supplementary Table 3). The highest included the left insula (0.44±0.25mm), the left rostral anterior cingulate (0.34±0.23mm), the right inferior parietal gyrus (0.30±0.21mm), and the right temporal pole (0.30±0.32mm). The cortical areas with highest recovery included: nine in the temporal lobe, nine in the frontal, two in the parietal, four in the cingulate, one insula (left hemisphere), one in the occipital (right lingual gyrus), and two in the pre/post-central gyri.

Overall, by comparing the rankings of recovery in cortical thickness at both 5 and 9 months of training, one noticed that the two lists are almost identical in terms of their composition, i.e. they contain, with the exception of the right entorhinal, left lateral orbital frontal, and left isthmus cingulate that appear only in the 5 month ranking list, and right superior frontal gyrus, the right pars opercularis, and right caudal anterior cingulate that appear only in the 9 months ranking, the same group of 14 cortical areas appear in both rankings. What differs is the position which these 14 cortical areas occupy in the two rankings. In most cases the peak of the recovery in cortical thickness was already reached at 5 months. By 9 months, either these values remained stable or showed only small increases or decreases, suggesting that most of the recovery in cortical thickness was already in place by 5 months when the whole experimental group was considered together.

When one considered the average increase in cortical thickness of individual cortical lobes or lobules, at both 5 and 9 months (Tables 7 and 9), the insula showed the highest increase (0.26±0.13mm at 5 months and 0.28±0.16 at 9 months), followed by the temporal lobe at 5 months (0.23±0.12mm) and 9 months (0.17±0.14mm). Next, at 5 months, one finds the frontal lobe (0.16±0.12mm), and at 9 months the cingulate cortex (0.15±0.12mm). Then, the pre-postcentral gyri with increases of 0.14±0.09mm (5 months) and 0.14±0.11mm (9 months). The parietal lobe follows, showing increases at both at 5 (0.12±0.06mm) and 9 months (0.11±0.08mm). This is followed by the paracentral lobule (0.11±0.06mm at 5 months, and 0.08±0.05mm at 9 months. Finally, the occipital lobe showed increases at 5 (0.07±0.05mm) and 9 months (0.03±0.06mm). At 5 months, we also noticed some differences across the two hemispheres (Table 10). For example, while in the left hemisphere the insula experienced an increase of 0.43±0.22mm, the improvement on the right was four times smaller at 0.10±0.11mm. Conversely, the temporal lobe in the right hemisphere experienced an increase of 0.30±0.15mm, compared to 0.16±0.09mm in the left hemisphere. The occipital lobe also showed an increase twice as large in the right (0.10±0.08mm) versus the left hemisphere (0.05±0.03mm). The remaining lobes/lobule showed similar increases in cortical thickness across the hemispheres. At 9 months, the left insula maintained a cortical thickness that was four times greater when compared to the right homologous area.

**Table 10.** Improvement of the left **(A)** and right **(B)** hemispheres areas in 5 months (AVG±SE; measurements in mm) for WANR group (n=11). The insulas are shown in green; cortical areas in the temporal lobe are shown in brown; frontal areas in salmon; cingulate lobe areas are shown in orange; the combined pre and postcentral gyri are shown in light red; parietal areas are shown in yellow; paracentral areas are shown in dark red; and occipital lobe areas are shown in blue.

| A) |  | B) |  |
| --- | --- | --- | --- |
| LEFT Hemisphere |  | RIGHT Hemisphere |  |
| WANR Group (n=11) |  | WANR Group (n=11) |  |
| | AVG IMP (mm) ± SE | | AVG IMP (mm) ± SE |
| Insula | $0.43 \pm 0.22$ | Temporal Lobe | $0.30 \pm 0.15$ |
| Cingulate | $0.17 \pm 0.09$ | Frontal Lobe | $0.18 \pm 0.14$ |
| Temporal Lobe | $0.16 \pm 0.09$ | Pre-Postcentral | $0.14 \pm 0.10$ |
| Frontal Lobe | $0.15 \pm 0.10$ | Cingulate | $0.12 \pm 0.13$ |
| Pre-Postcentral | $0.15 \pm 0.09$ | Parietal Lobe | $0.12 \pm 0.07$ |
| Parietal Lobe | $0.12 \pm 0.06$ | Paracentral | $0.11 \pm 0.06$ |
| Paracentral | $0.10 \pm 0.07$ | Insula | $0.10 \pm 0.11$ |
| Occipital Lobe | $0.05 \pm 0.03$ | Occipital Lobe | $0.10 \pm 0.08$ |

##### 3.3.1.3. Comparison of WANR patients that transition or not to ASIA C

In addition to comparing the results of the entire WANR group with the NR data at 9 months, we also carried out a subdivision of the WANR group into two subgroups: one with patients that made the transition from ASIA A to ASIA C (n=4), and one for patients that, despite showing improvements in their WISCI, remained at the ASIA A status (n=6).

When comparing the two subgroups at 5 months of training, the largest increase in cortical thickness in the ASIA A-C subgroup was located in the right temporal pole (0.87±0.45mm) and in the left insula (0.72±0.46mm), while the same structures led the ranking in the ASIA A-A subgroup, albeit with smaller gains (respectively 0.25±0.18mm and 0.19±0.09mm) (Table 11). When cortical areas showing thickness gains equal to or higher than 0.2mm were considered, one could identify a total of 36 cortical areas (or 54% of the 68 cortical areas analyzed) in the ASIA A-C subgroup. These included 13 areas in the temporal lobe, nine in the frontal, five in the cingulate cortex, three in the parietal, two in the pre- and postcentral gyri, two in the occipital lobe, one in the insula and one in the paracentral lobule (see areas limited by the black arrow; Supplementary Table 4). Only seven cortical areas in the subgroup ASIA A-A displayed gains equal to or above 0.2mm: four areas in the temporal lobe, one in the frontal lobe, one in the cingulate cortex, and the left insula. As in other cases, the right temporal pole (0.25±0.18mm) and the left insula (0.19±0.09mm) led the ranking Supplementary Table 5.

**Table 11.** Top 20 cortical areas that exhibited the greatest improvements in 5 months (AVG±SE; measurements in mm) for ASIA A-C (n=5), the values of the same areas for ASIA A-A patients (n=6) and the Absolute Hedges’ g values for ASIA A-C x ASIA A-A. Both subgroups are part of the WANR group. Hedges’ g represents how many pooled standard deviations apart two group means are, with a small-sample bias correction. Its magnitude conveys the effect size (∼0.2 small, ∼0.5 medium, ∼0.8 large, ≥1.2 very large). Cortical areas in temporal lobe are shown in brown; insula areas are shown in green; cingulate lobe areas are shown in orange; frontal lobe areas are shown in salmon; and parietal lobe areas are shown in yellow. For the designation of each cortical area, the letter “l” depicts left hemisphere, and “r” the right hemisphere.

| Cortical Areas |  | ASIA A-C (n=5)<br>AVG IMP (mm) ± SE | ASIA A-A (n=6)<br>AVG IMP (mm) ± SE | Abs<br>Hedges' g |
| --- | --- | --- | --- | --- |
| Temporal | rtemporalpole | 0.87 ± 0.45 | 0.25 ± 0.18 | 0.77 |
| Insula | linsula | 0.72 ± 0.46 | 0.19 ± 0.09 | 0.69 |
| Temporal | rentorhinal | 0.64 ± 0.20 | 0.18 ± 0.22 | 0.86 |
| Cingulate | rrostralanteriorcingulate | 0.64 ± 0.53 | -0.04 ± 0.08 | 0.77 |
| Frontal | lparsorbitalis | 0.64 ± 0.43 | 0.16 ± 0.08 | 0.66 |
| Cingulate | lrostralanteriorcingulate | 0.62 ± 0.40 | 0.08 ± 0.07 | 0.82 |
| Temporal | rfusiform | 0.62 ± 0.44 | 0.04 ± 0.04 | 0.79 |
|  | rparahippocampal | 0.62 ± 0.33 | 0.10 ± 0.11 | 0.89 |
|  | rsuperiortemporal | 0.58 ± 0.45 | 0.12 ± 0.05 | 0.62 |
|  | rmiddletemporal | 0.56 ± 0.48 | 0.09 ± 0.04 | 0.61 |
| Parietal | rinferiorparietal | 0.56 ± 0.36 | 0.06 ± 0.06 | 0.82 |
| Frontal | rlateralorbitofrontal | 0.55 ± 0.45 | 0.15 ± 0.08 | 0.54 |
|  | rmedialorbitofrontal | 0.55 ± 0.46 | 0.06 ± 0.08 | 0.64 |
|  | lparsopercularis | 0.55 ± 0.44 | 0.09 ± 0.05 | 0.63 |
| Cingulate | rcaudalanteriorcingulate | 0.55 ± 0.48 | -0.17 ± 0.18 | 0.83 |
| Parietal | linferiorparietal | 0.53 ± 0.39 | 0.07 ± 0.02 | 0.72 |
| Temporal | rtransverse temporal | 0.52 ± 0.41 | 0.07 ± 0.10 | 0.65 |
|  | lfusiform | 0.52 ± 0.42 | 0.10 ± 0.06 | 0.60 |
| Frontal | rparsopercularis | 0.51 ± 0.44 | 0.03 ± 0.01 | 0.67 |
|  | rsuperiorfrontal | 0.50 ± 0.48 | 0.01 ± 0.07 | 0.63 |

Next, when we considered only the patients from the WANR group who made the transition from ASIA A-C, and compared them to the patients of the same group that remained as ASIA A, we noticed that, following a very significant increase from pre-training to 5 months, all cortical areas exhibited an even further increase in cortical thickness from 5 to 9 months of training. These increases were much larger than both the values obtained for the ASIA A-A subgroup of the experimental group and those of the control group. For instance, at 5 months of training, the insula led the gains in thickness with a much higher increase in the left hemisphere (0.72±0.46mm) than the right one (0.07±0.20mm) (Supplementary Table 6). By 9 months, the left insula had increased by 0.97±0.54mm when compared to the right one (0.34±0.19mm) (Supplementary Table 7).

Overall, at 9 months of training, the number of cortical areas of the ASIA A-C subgroup that showed cortical thickness gains above 0.2mm grew to 51 out of 68 cortical areas (or 75% of the analyzed sample) (Supplementary Table 8). These included the right temporal pole (1.04±0.65mm), the rostral anterior cingulate (0.92±0.54mm), the left entorhinal cortex (0.91±0.62mm), the right superior temporal gyrus (0.77±0.54mm). Even the right (0.64±0.4mm) and left (0.63±0.39mm) pre/post-central gyri showed very significant increases in the ASIA A-C subgroup. Overall, the distribution of the 51 cortical areas included: 16 cortical areas in the temporal lobe, 16 in the frontal lobe, five in the parietal, five in the cingulate, three in the pre- post-central gyri, the right and left insula, and two cortical areas (the right cuneus and the right lingual gyrus) in the occipital lobe. Apart from the insula, no significant differences were seen when we divided the cortical areas across the two hemispheres. For the subgroup ASIA A-A no cortical area reached the threshold of 0.2mm (Supplementary Table 9). All cortical areas seemed to remain very close to the values obtained in the control group.

The temporal lobe experienced the second largest improvement in cortical thickness at both 5 (0.38±0.24) and 9 months (0.53±0.28mm), followed by the cingulate cortex at 5 (0.32±0.21mm) and 9 months (0.40±0.25mm), and the frontal lobe which showed an increase of 0.30±0.26mm at 5 months to 0.47±0.28mm at 9 months. Meanwhile, the pre and postcentral gyri increased from 0.26±0.19mm (5 months) to 0.41±0.19 mm (9 months), and the parietal lobe increased from 0.23±0.13mm (5 months) to 0.31±0.15mm. Interestingly enough, the paracentral lobule exhibited much smaller recoveries at both 5 (0.15±0.10mm) and 9 months (0.21±0.06mm), while the occipital lobe showed an identical increase of 0.14±0.09mm at 5 months and 0.14±0.13mm at 9 months (Tables 12 and 14).

**Table 12.** Global improvement of the two brain hemispheres areas in 5 months (AVG±SE; measurements in mm) for ASIA A-C and ASIA A-A patients (both part of the WANR group). Comparison of the greatest improvement for ASIA A-C patients (n=5), the values of the same areas for ASIA A-A (n=6) and the Absolute Hedges’ g values for ASIA A-C x ASIA A-A groups. Hedges’ g represents how many pooled standard deviations apart two group means are, with a small-sample bias correction. Its magnitude conveys the effect size (∼0.2 small, ∼0.5 medium, ∼0.8 large, ≥1.2 very large). Insula areas are shown in green; temporal lobe areas are shown in brown; cingulate lobe areas are shown in orange; frontal lobe areas are shown in salmon; the combined pre and postcentral gyri are shown in light red; parietal areas are shown in yellow; paracentral areas are shown in dark red; and occipital lobe areas are shown in blue.

| Cortical Areas | ASIA A-C (n=5)<br>AVG IMP (mm) ± SE | ASIA A-A (n=6)<br>AVG IMP (mm) ± SE | Abs<br>Hedges' g |
| --- | --- | --- | --- |
| Insula | $0.40 \pm 0.28$ | $0.15 \pm 0.10$ | 0.49 |
| Temporal Lobe | $0.38 \pm 0.24$ | $0.11 \pm 0.07$ | 0.66 |
| Cingulate | $0.32 \pm 0.21$ | $0.002 \pm 0.03$ | 0.92 |
| Frontal Lobe | $0.30 \pm 0.26$ | $0.05 \pm 0.06$ | 0.57 |
| Pre-Postcentral | $0.26 \pm 0.19$ | $0.05 \pm 0.07$ | 0.62 |
| Parietal Lobe | $0.23 \pm 0.13$ | $0.03 \pm 0.02$ | 0.97 |
| Paracentral | $0.15 \pm 0.10$ | $0.07 \pm 0.07$ | 0.37 |
| Occipital Lobe | $0.14 \pm 0.09$ | $0.02 \pm 0.03$ | 0.77 |

**Table 13.** Top 20 cortical areas that exhibited the greatest improvement in 9 months (AVG±SE; measurements in mm) for ASIA A-C (n=4), the values of the same areas for ASIA A-A patients (n=6), and the and the Absolute Hedges’ g values for ASIA A-C x ASIA A-A groups. Both subgroups are part of the WANR group. Hedges’ g represents how many pooled standard deviations apart two group means are, with a small-sample bias correction. Its magnitude conveys the effect size (∼0.2 small, ∼0.5 medium, ∼0.8 large, ≥1.2 very large). Cortical areas in temporal lobe are shown in brown; insula areas are shown in green; cingulate lobe areas are shown in orange; frontal lobe areas are shown in salmon; and parietal lobe areas are shown in yellow. For the designation of each cortical area, the letter “l” depicts left hemisphere, and “r” the right hemisphere.

|  |  | ASIA A-C (n=4) | ASIA A-A (n=6) | Abs |
| --- | --- | --- | --- | --- |
|  | Cortical Areas | AVG IMP (mm) ± SE | AVG IMP (mm) ± SE | Hedges' g |
| Temporal | rtemporalpole | 1.04 ± 0.65 | -0.20 ± 0.10 | 1.39 |
| Insula | linsula | 0.97 ± 0.54 | 0.08 ± 0.08 | 1.17 |
| Cingulate | rrostralanteriorcingulate | 0.92 ± 0.54 | -0.16 ± 0.03 | 1.48 |
| Temporal | lensorhinal | 0.91 ± 0.62 | -0.18 ± 0.13 | 1.23 |
|  | rsuperiortemporal | 0.77 ± 0.54 | -0.05 ± 0.05 | 1.11 |
| Cingulate | lrostralanteriorcingulate | 0.76 ± 0.54 | 0.05 ± 0.06 | 0.95 |
| Frontal | rsuperiorfrontal | 0.75 ± 0.50 | -0.04 ± 0.06 | 1.13 |
| Temporal | rmiddletemporal | 0.75 ± 0.59 | -0.04 ± 0.03 | 0.97 |
|  | rfusiform | 0.75 ± 0.53 | -0.04 ± 0.03 | 1.08 |
|  | rparahippocampal | 0.74 ± 0.37 | -0.03 ± 0.06 | 1.50 |
| Parietal | rinferiorparietal | 0.74 ± 0.47 | 0.01 ± 0.04 | 1.13 |
| Frontal | llateralorbitofrontal | 0.74 ± 0.51 | -0.10 ± 0.07 | 1.18 |
|  | lparsopercularis | 0.70 ± 0.51 | 0.01 ± 0.03 | 1.00 |
| Temporal | lfusiform | 0.69 ± 0.47 | 0.01 ± 0.09 | 1.01 |
| Frontal | rlateralorbitofrontal | 0.68 ± 0.54 | -0.04 ± 0.08 | 0.97 |
|  | rparsopercularis | 0.68 ± 0.55 | -0.02 ± 0.04 | 0.94 |
|  | lparsorbitalis | 0.68 ± 0.56 | -0.04 ± 0.08 | 0.93 |
| Temporal | rtransversetemporal | 0.68 ± 0.45 | 0.01 ± 0.09 | 1.05 |
| Parietal | linferiorparietal | 0.67 ± 0.47 | -0.04 ± 0.05 | 1.10 |
| Frontal | rrostralmiddlefrontal | 0.67 ± 0.45 | -0.04 ± 0.08 | 1.13 |

**Table 14.** Global improvement of the two brain hemispheres areas in 9 months (AVG±SE; measurements in mm) for ASIA A-C and ASIA A-A patients (both part of the WANR group). Comparison of the greatest improvement for ASIA A-C patients (n=4), the values of the same areas for ASIA A-A (n=6), and the Absolute Hedges’ g values for ASIA A-C x ASIA A-A groups. Hedges’ g represents how many pooled standard deviations apart two group means are, with a small-sample bias correction. Its magnitude conveys the effect size (∼0.2 small, ∼0.5 medium, ∼0.8 large, ≥1.2 very large). Insula areas are shown in green; temporal lobe areas are shown in brown; frontal lobe areas are shown in salmon; the combined pre and postcentral gyri are shown in light red; cingulate lobe areas are shown in orange; parietal areas are shown in yellow; paracentral areas are shown in dark red; and occipital lobe areas are shown in blue.

|  | ASIA A-C (n=4) | ASIA A-A (n=6) | Abs |
| --- | --- | --- | --- |
| Cortical Areas | AVG IMP (mm) ± SE | AVG IMP (mm) ± SE | Hedges' g |
| Insula | 0.65 ± 0.29 | 0.03 ± 0.09 | 1.42 |
| Temporal Lobe | 0.53 ± 0.28 | -0.07 ± 0.04 | 1.55 |
| Frontal Lobe | 0.47 ± 0.28 | -0.07 ± 0.04 | 1.35 |
| Pre-Postcentral | 0.41 ± 0.19 | -0.03 ± 0.06 | 1.51 |
| Cingulate | 0.40 ± 0.25 | -0.01 ± 0.05 | 1.15 |
| Parietal Lobe | 0.31 ± 0.15 | -0.03 ± 0.05 | 1.50 |
| Paracentral | 0.21 ± 0.06 | -0.01 ± 0.06 | 1.43 |
| Occipital Lobe | 0.14 ± 0.13 | -0.04 ± 0.03 | 1.00 |

Conversely, in the NR group (n=5), at 9 months (Table 9), we observed a very small increase in cortical thickness (e.g. in the temporal lobe 0.06±0.13mm), no meaningful increase (e.g. occipital cortex, 0.01±0.05mm), and even cortical thickness reductions. The most significant of the latter were the pre- and postcentral gyri (-0.13±0.08mm). No major differences between hemispheres were observed in the control group. Such negligible changes were virtually identical to the pre-training measures, including the fact that some lobes tended to show a further small reduction in cortical thickness from 5 to 9 months of training.

##### 3.3.1.4. Comparison of Healthy x SCI groups

Next, we performed the same statistical analysis employed to compare our initial cohort of SCI patients to healthy controls in order to compare the distribution of cortical thickness from the same healthy patients against the results obtained for patients after the end of the 9-month training with the WANR (Figure 14A, for the group descriptions, see section 2.7.1 in Materials and Methods). Although the two distributions were still statistically different, it was clear that the training with the WANR reduced the differences in cortical thickness between the two groups. To further test whether this hypothesis was truly consistent, we added to the present experimental data set of 14 WANR-CN patients similar structural MRI data obtained from six WANR-BR patients, who had trained with the WANR protocol in a previous study that lasted for 28 months (see [14, 20]). Once this data set was added, the difference between the two distributions (healthy vs treated SCI patients) was further reduced (Figure 14B). As seen in Figure 14C, the two distributions became even closer, suggesting that the longer the training with the WANR protocol, the higher the chances of reversing the widespread reduction in cortical thickness observed following a SCI.

**Figure 14.**
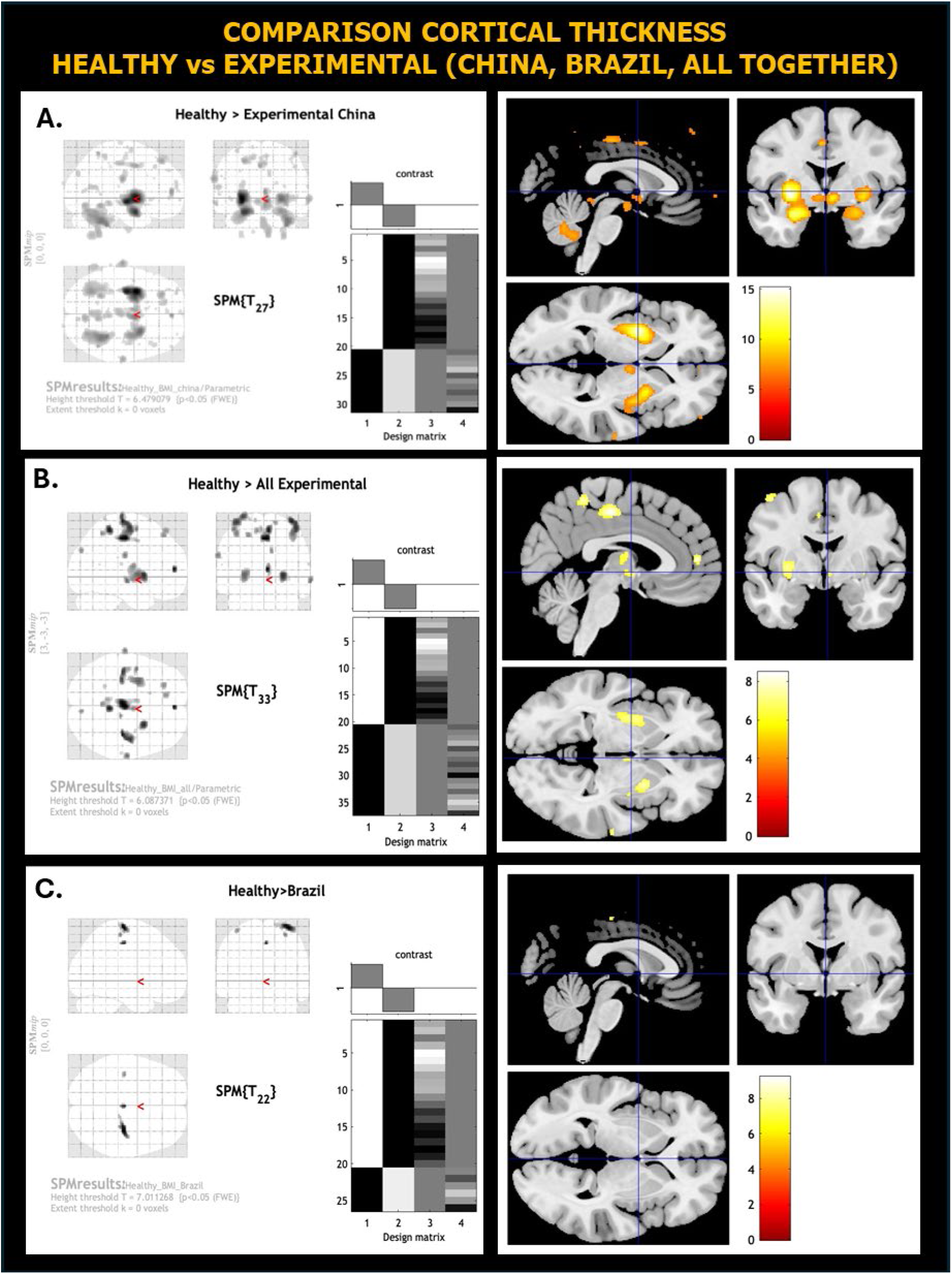
(**A**) Statistical analysis of cortical thickness and subcortical structure volumes differences between brains of 20 healthy subjects (data from OpenfMRI database - Washington University 120) and 14 patients at the end of the 9-month training with the WANR (WANR-CN). The two distributions were still statistically different (p<0.05, Height threshold T=6.479079, Extent threshold k= 0 voxels); it was clear that the training with the WANR reduced the differences in cortical thickness between the two groups. **(B)** To further test whether this hypothesis was truly consistent, we added to the present experimental data set of 14 patients with similar structural MRI data obtained from six other patients (WANR-BR) who had trained with the WANR protocol in a previous study in Brazil, that lasted for 28 months. Once this data set was added, the difference between the two distributions (healthy vs treated SCI patients) was further reduced (p<0.05, Height threshold T=6.087371, Extent threshold k= 0 voxels). (**C**) As a final check, we then compared the healthy distribution of cortical thickness only with the data from the six WANR-BR patients who had trained for a longer period of time. As can be seen, the two distributions became even closer, suggesting that the longer the training with the WANR protocol, the higher the chances of reversing the widespread reduction in cortical thickness observed following a SCI (p<0.05, Height threshold T=7.011268, Extent threshold k= 0 voxels).

##### 3.3.1.4a. Pairwise cortical thickness correlations

In order to further characterize the global effect of the spinal cord injury and the training with the WANR protocol on cortical structure, we calculated the S correlations between the thickness of 68 cortical areas in five different groups: a pool of control healthy patients (n=20), a sample of the spinal cord injury patients (n=23) (SCI patients), a group of patients after 5 months (n=14) and 9 months (n=12) of training with the WANR protocol (WANR-CN), and finally a group of WANR-BR patients (n=6). For the group descriptions, see section 2.7.1 in Materials and Methods). For each group, we selected the 150 more significant correlations between pairs of cortical areas to create direct graphs that represent the global spatial distribution of cortico-cortical thickness correlations. Figures 15-19 exhibit the graph for each of the five groups. To further quantify and characterize the spatial distribution of such correlations we divided them into four types:

1. **Bilateral homologous** (between the same cortical areas across both cortical hemispheres).
2. **Bilateral heterologous**, between different cortical areas in the right and left hemispheres. This group was further divided into two subgroups: one (called Same) in which the correlation was between cortical areas in both hemispheres that belonged to the same lobe (e.g. left and right parietal lobe), or a second subgroup (Different), a pair of cortical areas that belonged to different lobes.
3. **Ipsilateral short-range,** involving a pair of cortical areas belonging to the same lobe in the same hemisphere.
4. **Ipsilateral long-range,** involving a pair of cortical areas belonging to distinct lobes in the same hemisphere.

**Figure 15.**
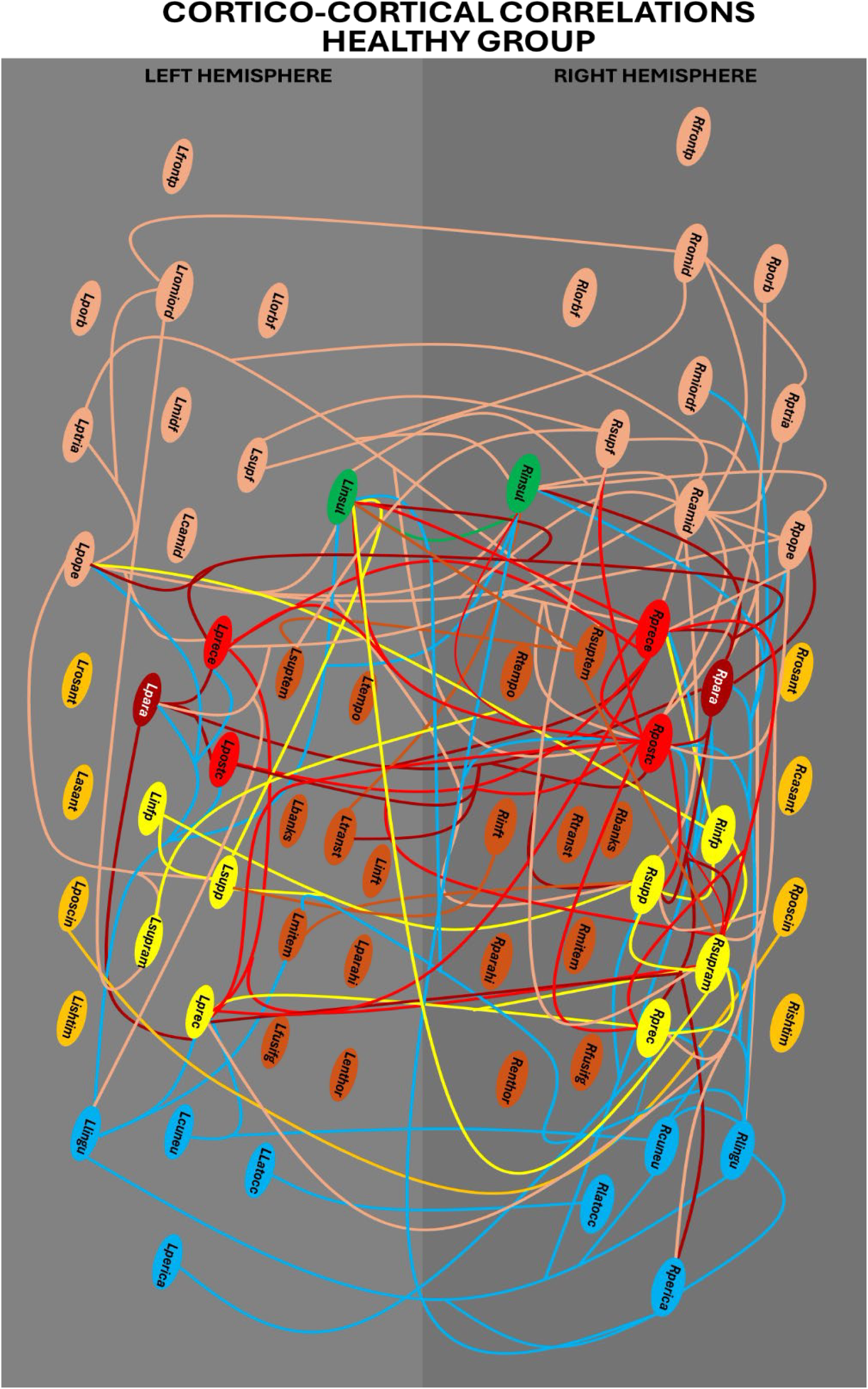
Graphical representation of the global spatial distributions of 150 more significant correlations between pairs of cortical areas of healthy individuals. The left hemisphere is represented in light grey (on the left) and the right hemisphere is represented in dark grey (on the right). Oval structures represent the cortical areas studied, which are color coded as follows: In **dark red** is the **paracentral** area for each hemisphere: Rpara/Lpara (paracentral). In bright **red** are the two **pre-postcentral** areas: Rpostc/Lpostc (postcentral), Rprece/Lprece (precentral). In **brown** are nine **temporal** lobe areas: Rbanks/Lbanks (bankssts), Renthor/Lenthor (enthorrinal), Rfusig/Lfusig (fusiform), Rinft/Linft (inferiortemporal), Rmitem/Lmitem (middletemporal), Rparahi/Lparahi (parahippocampal), Rsuptem/Lsuptem (superiortemporal), Rtempo/Ltempo (temporalpole), Rtranst/Ltranst (transversetemporal). In **orange** are the four **cingulate** areas: Rcasant/Lcasant (caudalanteriorcingulate), Risthim (isthmuscingulate), Rposcin/Lposcin (posteriorcingulate), Rrosant/Lrosant (rostralanteriorcingulate). In **salmon** are the eight **frontal** lobe areas: Rcamid/Lcamid (caudalmiddlefrontal), Rlorbf/ Llorbf (lateralorbitofrontal), Rmiordf/Lmiordf (medialorbitofrontal), Rpope/Lpope (parsopercularis). Rporb/Lporb (parsorbitalis), Rptria/Lptria (parstriangularis), Rromid/Lromid (rostralmiddlefrontal), Rsupf/Lsupf (superiorfrontal), Rfrontp/Lfrontp (frontalpole). In **yellow** are the four **parietal** lobe areas: Rinfp/Linfp (inferiorparietal), Rprec/Lprec (precuneus), Rsupp/Lsupp (superiorparietal), Rsupram/Lsupram (supramarginal). In **blue** are the four **occipital** lobe areas: Rcuneu/Lcuneu (cuneus), Rlatocc/Llatocc (lateraloccipital), Rlingu/Llingu (lingual), Rperica/Lperica (pericalcarine). In **green** is the **insula**: Rinsul/Linsul.

Detailed analysis of each of the directed graphs, as well comparisons across the five groups analyzed, revealed interesting dynamic features in the spatial distribution of these cortico-cortical correlations. Starting with Figure 15, one notices that, in healthy subjects, there are many cortical hubs distributed across all lobes of both cerebral hemispheres. This somewhat spatially diffuse distribution of cortico-cortical correlation hubs defines a core formed by the strong correlations (red lines) among the precentral and postcentral gyri and the paracentral lobule in both the left and right cerebral hemispheres. Not only are these structures heavily correlated with one another, but they also exhibit strong correlations with other cortical hubs. For instance, the pre- and postcentral gyri shared correlations with hubs in the parietal (left and right superior parietal lobe, the right supramarginal gyrus, and the right and left precuneus) and frontal lobe (right superior frontal gyrus). Meanwhile, the thickness of the paracentral lobules in both hemispheres is highly correlated with the right and left insula, the right and left pars opercularis, and the right and left precuneus (parietal lobe). A correlation between the right paracentral lobule and the right pericalcarine region is also noteworthy.

In healthy subjects, several areas in the frontal lobe also defined important correlation hubs, like the right caudal middle frontal gyrus, the right superior frontal gyrus, the right and left pars opercularis, and the right and left middle frontal gyrus. In the parietal lobe, the right and left superior parietal gyrus, the right and left supramarginal gyrus, the right and left precuneus, and the right inferior parietal gyrus are the main hubs. In the occipital lobes, both the right and left lingual gyri thickness are significantly correlated with cortical areas in the frontal, parietal, temporal (left middle temporal gyrus), and even the left and right insulas. The thickness of the right and left insulas are also highly correlated with cortical areas in all cortical lobes.

However, it is noticeable in healthy subjects that the widespread distributions of hubs and their diffuse correlations, observed using the top 150 correlations, somewhat spared most of the areas in the temporal lobes, perhaps with the exception of the right superior temporal gyrus. The same relative sparing can be observed when the cingulate gyrus is considered. Overall, using the classification of Table 15, in healthy subjects 17 correlations belonged to the bilateral homologous group, 52 to the bilateral heterologous (21 of the Same variety, and 31 in the Different category), 33 to the short-range ipsilateral, and 48 to the long-range ipsilateral variety.

**Table 15.** Summary of the spatial distribution of cortico-cortical thickness correlations obtained for all the five groups: healthy, SCI, WANR-CN 5 months, WANR-CN 9 months and WANR-BR 28 months.

|  | **Healthy** | **SCI** | **WANR-CN**<br>**5 months** | **WANR-CN**<br>**9 months** | **WANR-BR**<br>**28 months** |
| --- | --- | --- | --- | --- | --- |
| Bilateral homologous | 17 | 18 | 7 | 11 | 3 |
| Ipsilateral short | 33 | 30 | 23 | 27 | 22 |
| Bilateral heterologous | 52 | 60 | 53 | 60 | 70 |
| Same lobe | 21 | 23 | 17 | 20 | 17 |
| Different lobe | 31 | 37 | 36 | 40 | 53 |
| Ipsilateral long | 48 | 42 | 67 | 52 | 55 |

Comparing Figure 15 to Figure 16, which represents the direct graph describing the spatial distribution of thickness correlations in a pool of spinal cord injury patients, one notices a striking contrast, in terms of both the distribution of cortical hubs and their main correlations. For starters, Figure 15 reveals the central core of hubs and correlations is now located in the temporal lobes, replacing the main cluster formed by the pre/postcentral gyri and paracentral lobule observed in healthy patients. In SCI patients, this latter cluster preserved correlations primarily only among themselves but lost most of the correlations with other cortical areas located in different lobes.

**Figure 16.**
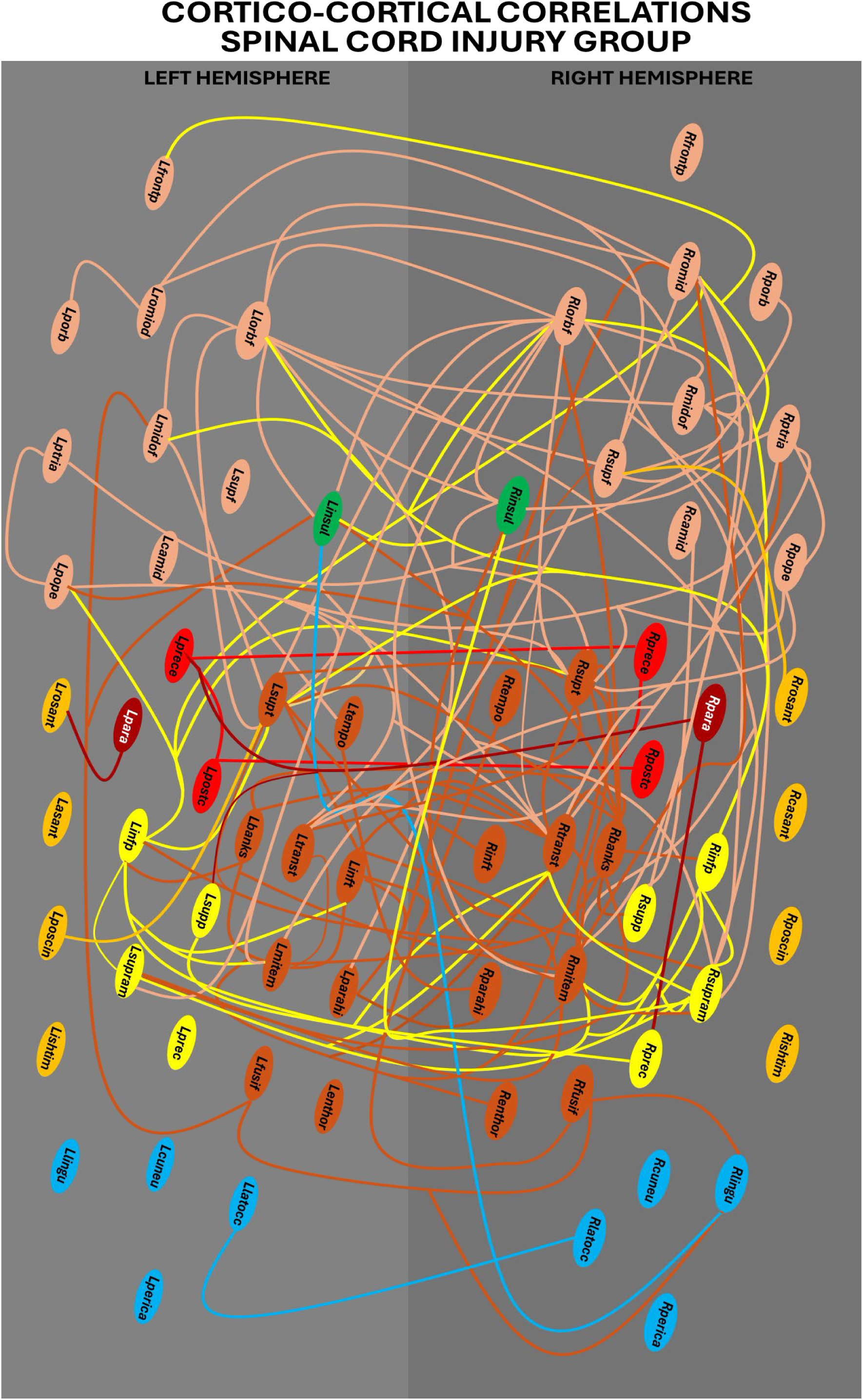
Graphical representation of the global spatial distributions of 150 more significant correlations between pairs of cortical areas for the SCI group. The left hemisphere is represented in light grey (on the left) and the right hemisphere is represented in dark grey (on the right). Oval structures represent the cortical areas studied, which are color coded as follows: In **dark red** is the **Paracentral** area for each hemisphere: Rpara/Lpara (paracentral). In bright **red** are the two **pre-postcentral** areas: Rpostc/Lpostc (postcentral), Rprece/Lprece (precentral). In **brown** are nine **temporal** lobe areas: Rbanks/Lbanks (bankssts), Renthor/Lenthor (enthorrinal), Rfusig/Lfusig (fusiform), Rinft/Linft (inferiortemporal), Rmitem/Lmitem (middletemporal), Rparahi/Lparahi (parahippocampal), Rsuptem/Lsuptem (superiortemporal), Rtempo/Ltempo (temporalpole), Rtranst/Ltranst (transversetemporal). In **orange** are the four **cingulate** areas: Rcasant/Lcasant (caudalanteriorcingulate), Risthim (isthmuscingulate), Rposcin/Lposcin (posteriorcingulate), Rrosant/Lrosant (rostralanteriorcingulate). In **salmon** are the eight **frontal** lobe areas: Rcamid/Lcamid (caudalmiddlefrontal), Rlorbf/ Llorbf (lateralorbitofrontal), Rmiordf/Lmiordf (medialorbitofrontal), Rpope/Lpope (parsopercularis). Rporb/Lporb (parsorbitalis), Rptria/Lptria (parstriangularis), Rromid/Lromid (rostralmiddlefrontal), Rsupf/Lsupf (superiorfrontal), Rfrontp/Lfrontp (frontalpole). In **yellow** are the four **parietal** lobe areas: Rinfp/Linfp (inferiorparietal), Rprec/Lprec (precuneus), Rsupp/Lsupp (superiorparietal), Rsupram/Lsupram (supramarginal). In **blue** are the four **occipital** lobe areas: Rcuneu/Lcuneu (cuneus), Rlatocc/Llatocc (lateraloccipital), Rlingu/Llingu (lingual), Rperica/Lperica (pericalcarine). In **green** is the **insula**: Rinsul/Linsul.

In the SCI group, except for the right and left temporal pole, almost every temporal cortical area analyzed was heavily correlated, not only among themselves, but also with cortical regions all over both cerebral hemispheres. These long-range correlations of temporal cortical area thickness included primarily frontal, parietal, and the insulas, and only a single structure in the occipital lobe (the right lingual gyrus). The main temporal hubs included, in both hemispheres, the banks of the superior temporal gyrus, superior temporal gyri, the transverse gyrus, the middle temporal gyrus, the parahippocampal gyrus, and to a lesser degree the fusiform gyrus. In the frontal lobe, the main hubs moved a bit more rostrally, since now they included primarily the right and left lateral orbital gyri, the right, and to a lesser degree, the left rostral middle frontal gyrus, the middle orbitofrontal gyri in both hemispheres, and the right superior frontal gyrus. In the parietal lobes, the main hubs were now restricted to the inferior parietal and the supramarginal gyri in both hemispheres. As in healthy subjects, the thickness of both the left and right insulas were heavily correlated with many cortical areas, but in SCI patients they did not include the pre/postcentral gyri and the paracentral lobule. Unlike in healthy subjects, no correlation cortical hub was identified in the occipital lobes, considering the top 150 cortico-cortical correlations. The same was true for the cingulate gyrus. Overall, 18 correlations involved bilateral homologous cortical areas, 60 bilateral heterologous (23 of the Same variety and 37 of the Different), 30 correlations of the short-range ipsilateral, and 42 long-range ipsilateral.

At 5 months of training with the WANR (Figure 17), we noticed already significant changes in the global spatial distribution of cortical thickness correlations when compared to the SCI group. First, there was a change in the pattern of correlations involving temporal cortical areas, which now included a greater number of long-range ipsilateral cortico-cortical correlations (67 in this group and 42 in the SCI group and 48 in the Healthy group) with the frontal and parietal lobes, but also with right and left postcentral gyri, the right and left paracentral lobule and the left precentral gyrus.

**Figure 17.**
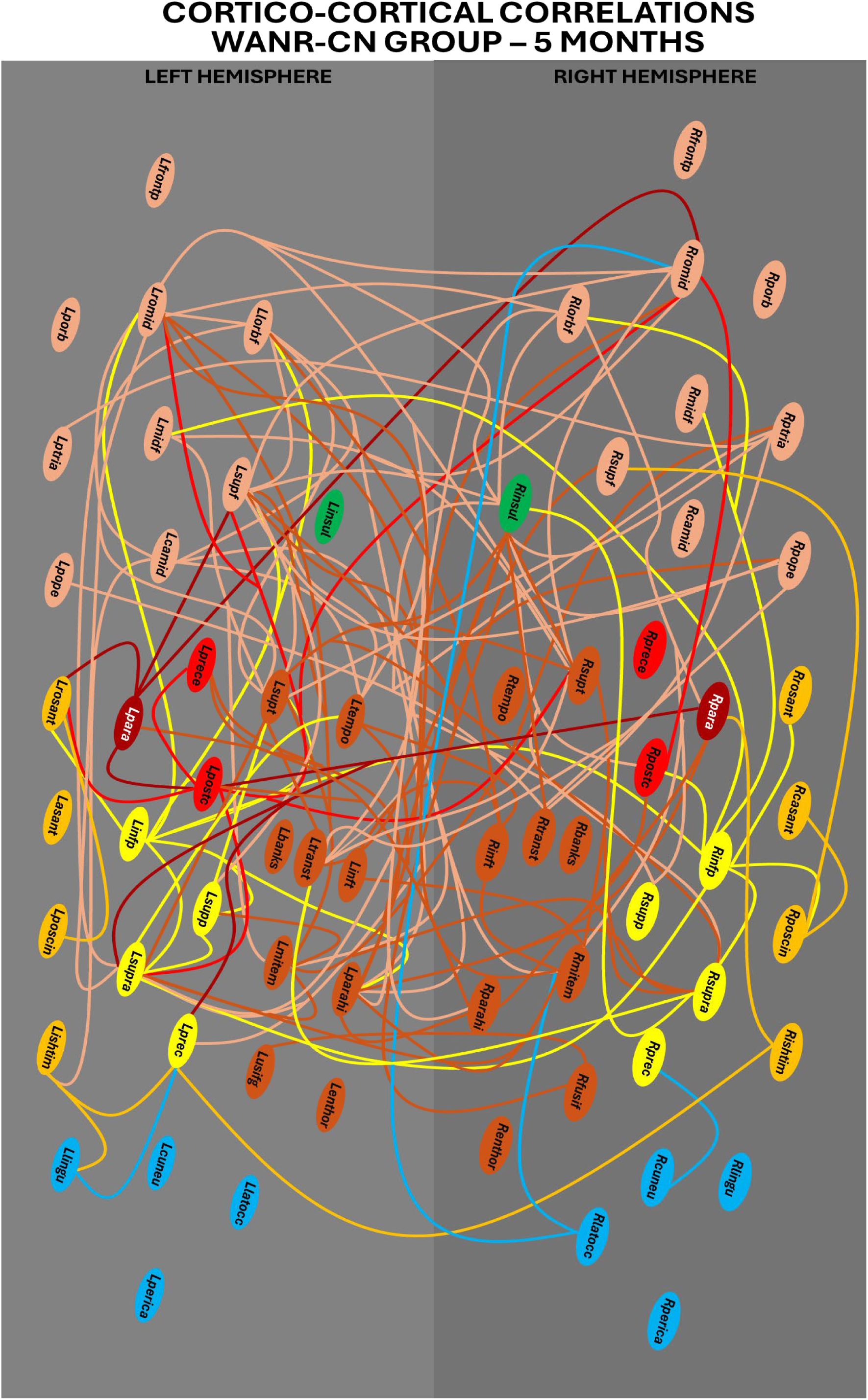
Graphical representation of the global spatial distributions of 150 more significant correlations between pairs of cortical areas for the WANR-CN group after 5 months of training. The left hemisphere is represented in light grey (on the left) and the right hemisphere is represented in dark grey (on the right). Oval structures represent the cortical areas studied, which are color coded as follows: In **dark red** is the **paracentral** area for each hemisphere: Rpara/Lpara (paracentral). In bright **red** are the two **pre-postcentral** areas: Rpostc/Lpostc (postcentral), Rprece/Lprece (precentral). In **brown** are nine **temporal** lobe areas: Rbanks/Lbanks (bankssts), Renthor/Lenthor (enthorrinal), Rfusig/Lfusig (fusiform), Rinft/Linft (inferiortemporal), Rmitem/Lmitem (middletemporal), Rparahi/Lparahi (parahippocampal), Rsuptem/Lsuptem (superiortemporal), Rtempo/Ltempo (temporalpole), Rtranst/Ltranst (transversetemporal). In **orange** are the four **cingulate** areas: Rcasant/Lcasant (caudalanteriorcingulate), Risthim (isthmuscingulate), Rposcin/Lposcin (posteriorcingulate), Rrosant/Lrosant (rostralanteriorcingulate). In **salmon** are the eight **frontal** lobe areas: Rcamid/Lcamid (caudalmiddlefrontal), Rlorbf/ Llorbf (lateralorbitofrontal), Rmiordf/Lmiordf (medialorbitofrontal), Rpope/Lpope (parsopercularis). Rporb/Lporb (parsorbitalis), Rptria/Lptria (parstriangularis), Rromid/Lromid (rostralmiddlefrontal), Rsupf/Lsupf (superiorfrontal), Rfrontp/Lfrontp (frontalpole). In **yellow** are the four **parietal** lobe areas: Rinfp/Linfp (inferiorparietal), Rprec/Lprec (precuneus), Rsupp/Lsupp (superiorparietal), Rsupram/Lsupram (supramarginal). In **blue** are the four **occipital** lobe areas: Rcuneu/Lcuneu (cuneus), Rlatocc/Llatocc (lateraloccipital), Rlingu/Llingu (lingual), Rperica/Lperica (pericalcarine). In **green** is the **insula**: Rinsul/Linsul.

Interestingly, the left and right paracentral lobules also recovered some of their long-range correlations with structures in the parietal and frontal lobes and among themselves, a clear distinction when compared to the SCI group. In the parietal lobe, the main hubs were the inferior parietal and the supramarginal gyrus in both hemispheres, and the left superior parietal gyrus. In the frontal lobe, both the rostral middle frontal gyrus and the lateral orbitofrontal gyrus in both hemispheres remained as the main hubs, together with the left superior frontal gyrus, and the right pars triangularis, right pars opercularis, and left caudal middle frontal lobe and middle orbitofrontal gyrus. The right insula remained a major hub, but not the left one.

In clear contrast to both the SCI and the Healthy groups, at 5 months of training many cortical areas located in the cingulate gyrus showed high thickness correlations among themselves and with other cortical areas, including in the opposite hemisphere. These included the right and left posterior and isthmus cingulate, the right caudal anterior cingulate and the left rostral anterior cingulate. Likewise, correlations involving areas in the occipital lobe also reappeared after 5 months of training. The main contributor was the right lateral occipital area and the left lingual gyrus. Overall, at 5 months of training, seven correlations included bilateral homologous, 53 bilateral heterologous (17 Same, and 36 Different), 23 short-range ipsilateral, and 67 long-range ipsilateral. Clearly, a more spatially distributed pattern of cortico-cortical correlations was evident after this initial period of training.

At the end of the clinical trial, at 9 months of training, the overall graph describing the spatial distribution of cortico-cortical thickness correlations changed significantly (Figure 18). For starters, the preeminence of correlations involving the pre/postcentral gyri and the paracentral lobule, observed in healthy subjects, but absent in the SCI group, was clearly present in this group, involving not only correlations among themselves, but also with other cortical structures. Likewise, a larger number of correlations involved different structures of the cingulate gyrus, including long-range ipsi- and bilateral correlations, both of the S and D varieties. These originated in the left and right posterior cingulate, the right rostral anterior cingulate, and the left isthmus. Also noticeable is the increase in correlations involving cortical hubs in the occipital lobe, with the highest contributions from the left lingual gyrus, and the right lateral occipital area.

**Figure 18.**
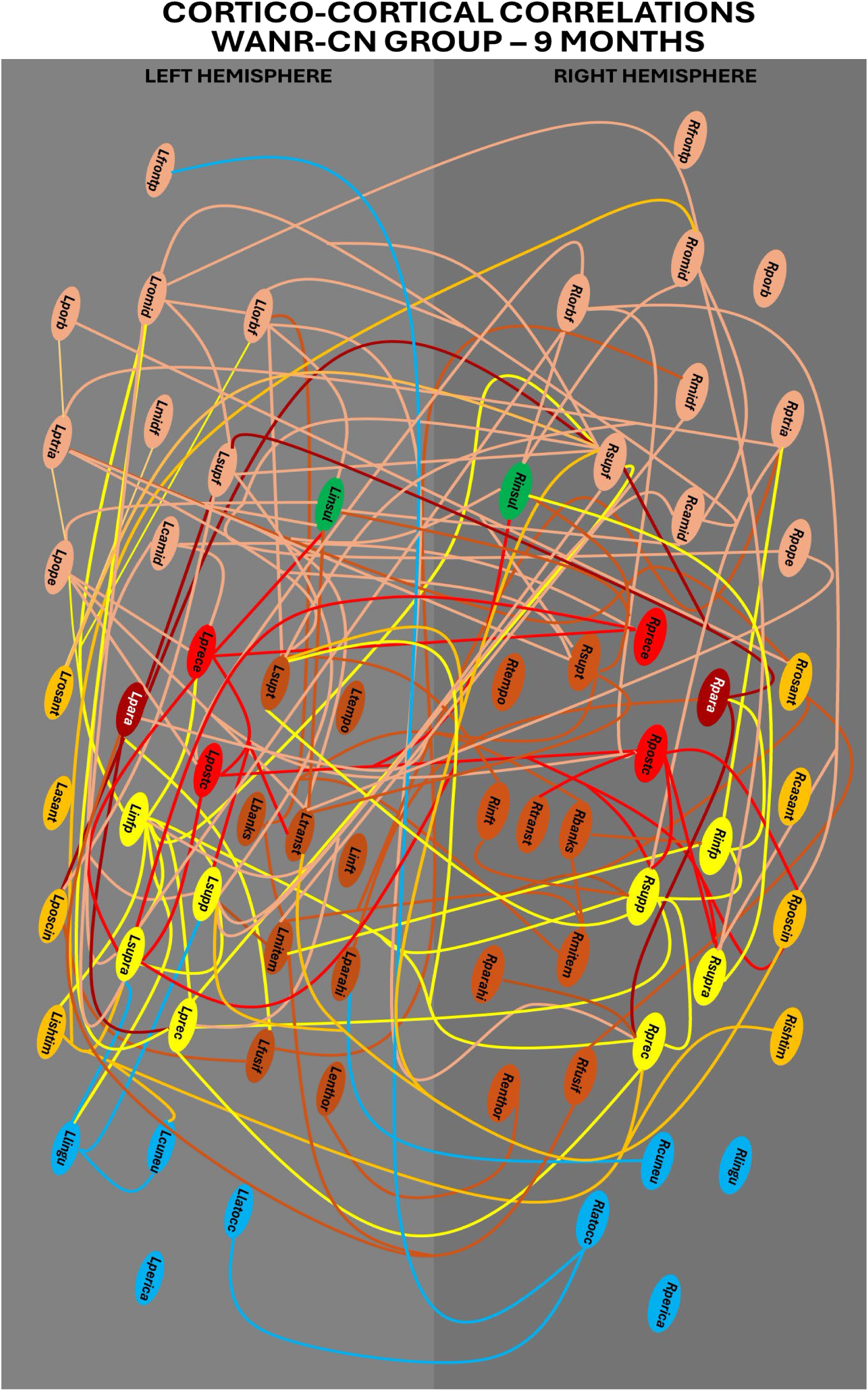
Graphical representation of the global spatial distributions of 150 more significant correlations between pairs of cortical areas for the WANR-CN group after 9 months of training. The left hemisphere is represented in light grey (on the left) and the right hemisphere is represented in dark grey (on the right). Oval structures represent the cortical areas studied, which are color coded as follows: In **dark red** is the **paracentral** area for each hemisphere: Rpara/Lpara (paracentral). In bright **red** are the two **pre-postcentral** areas: Rpostc/Lpostc (postcentral), Rprece/Lprece (precentral). In **brown** are nine **temporal** lobe areas: Rbanks/Lbanks (bankssts), Renthor/Lenthor (enthorrinal), Rfusig/Lfusig (fusiform), Rinft/Linft (inferiortemporal), Rmitem/Lmitem (middletemporal), Rparahi/Lparahi (parahippocampal), Rsuptem/Lsuptem (superiortemporal), Rtempo/Ltempo (temporalpole), Rtranst/Ltranst (transversetemporal). In **orange** are the four **cingulate** areas: Rcasant/Lcasant (caudalanteriorcingulate), Risthim (isthmuscingulate), Rposcin/Lposcin (posteriorcingulate), Rrosant/Lrosant (rostralanteriorcingulate). In **salmon** are the eight **frontal** lobe areas: Rcamid/Lcamid (caudalmiddlefrontal), Rlorbf/ Llorbf (lateralorbitofrontal), Rmiordf/Lmiordf (medialorbitofrontal), Rpope/Lpope (parsopercularis). Rporb/Lporb (parsorbitalis), Rptria/Lptria (parstriangularis), Rromid/Lromid (rostralmiddlefrontal), Rsupf/Lsupf (superiorfrontal), Rfrontp/Lfrontp (frontalpole). In **yellow** are the four **parietal** lobe areas: Rinfp/Linfp (inferiorparietal), Rprec/Lprec (precuneus), Rsupp/Lsupp (superiorparietal), Rsupram/Lsupram (supramarginal). In **blue** are the four **occipital** lobe areas: Rcuneu/Lcuneu (cuneus), Rlatocc/Llatocc (lateraloccipital), Rlingu/Llingu (lingual), Rperica/Lperica (pericalcarine). In **green** is the **insula**: Rinsul/Linsul.

In the parietal lobe, in addition to the inferior, superior lobes and supramarginal gyri in both hemispheres, both the left and right precuneus appeared as important correlation hubs. The right insula continued to be a major hub, while the left insula returned to be a major one, in contrast to the graph from the 5-months group. Regarding the frontal lobe, the bilateral superior frontal, orbitofrontal, rostral and caudal middle frontal gyri, and pars triangularis and opercularis defined the main frontal hub network. Conversely, although temporal cortical areas continued to provide an important contribution to the thickness correlation graph at 9 months of training, they were somewhat reduced, in terms of their extent and density, when compared to the SCI and the 5-months groups. After 9 months of training, the main temporal hubs include the superior and middle temporal gyri in both hemispheres, and the left transverse gyrus. Overall, in the 9-months group, 11 correlations were of the bilateral homologous type, 60 bilateral heterologous (20 S, and 40 D), 27 short-range ipsilateral, and 52 long-distance ipsilateral.

For the purpose of comparison, we also included in our analysis the same type of correlational data obtained from a different group of six patients enrolled in a previous study where subjects trained with the WANR for 28 months (WANR-BR group). The graph summarizing the cortico-cortical thickness correlations can be seen in Figure 19.

**Figure 19.**
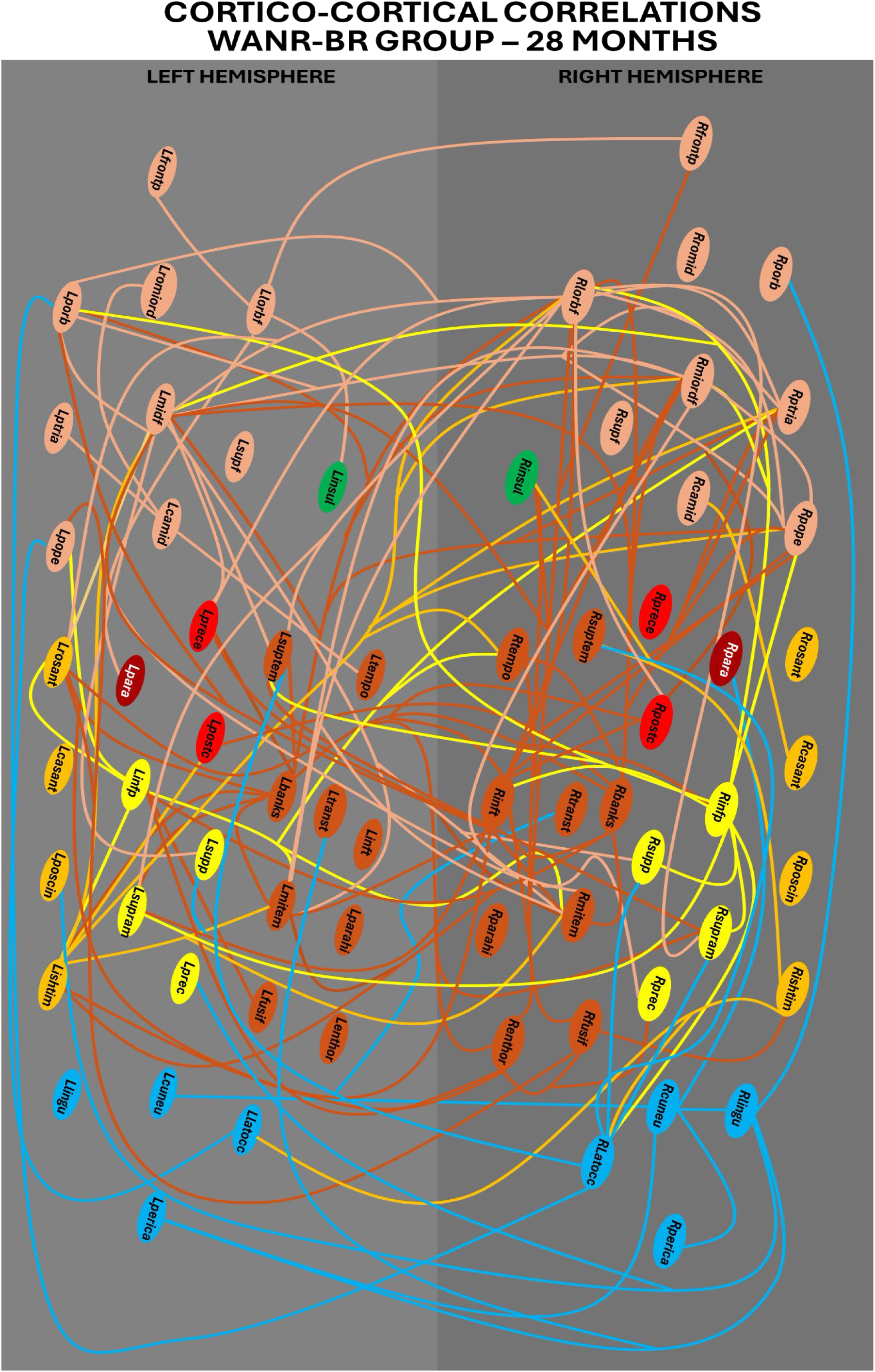
Graphical representation of the global spatial distributions of 150 more significant correlations between pairs of cortical areas for the WANR-BR group after 28 months of training. The left hemisphere is represented in light grey (on the left) and the right hemisphere is represented in dark grey (on the right). Oval structures represent the cortical areas studied, which are color coded as follows: In **dark red** is the **paracentral** area for each hemisphere: Rpara/Lpara (paracentral). In bright **red** are the two **pre-postcentral** areas: Rpostc/Lpostc (postcentral), Rprece/Lprece (precentral). In **brown** are nine **temporal** lobe areas: Rbanks/Lbanks (bankssts), Renthor/Lenthor (enthorrinal), Rfusig/Lfusig (fusiform), Rinft/Linft (inferiortemporal), Rmitem/Lmitem (middletemporal), Rparahi/Lparahi (parahippocampal), Rsuptem/Lsuptem (superiortemporal), Rtempo/Ltempo (temporalpole), Rtranst/Ltranst (transversetemporal). In **orange** are the four **cingulate** areas: Rcasant/Lcasant (caudalanteriorcingulate), Risthim (isthmuscingulate), Rposcin/Lposcin (posteriorcingulate), Rrosant/Lrosant (rostralanteriorcingulate). In **salmon** are the eight **frontal** lobe areas: Rcamid/Lcamid (caudalmiddlefrontal), Rlorbf/ Llorbf (lateralorbitofrontal), Rmiordf/Lmiordf (medialorbitofrontal), Rpope/Lpope (parsopercularis). Rporb/Lporb (parsorbitalis), Rptria/Lptria (parstriangularis), Rromid/Lromid (rostralmiddlefrontal), Rsupf/Lsupf (superiorfrontal), Rfrontp/Lfrontp (frontalpole). In **yellow** are the four **parietal** lobe areas: Rinfp/Linfp (inferiorparietal), Rprec/Lprec (precuneus), Rsupp/Lsupp (superiorparietal), Rsupram/Lsupram (supramarginal). In **blue** are the four **occipital** lobe areas: Rcuneu/Lcuneu (cuneus), Rlatocc/Llatocc (lateraloccipital), Rlingu/Llingu (lingual), Rperica/Lperica (pericalcarine). In **green** is the **insula**: Rinsul/Linsul.

**Figure 20.**
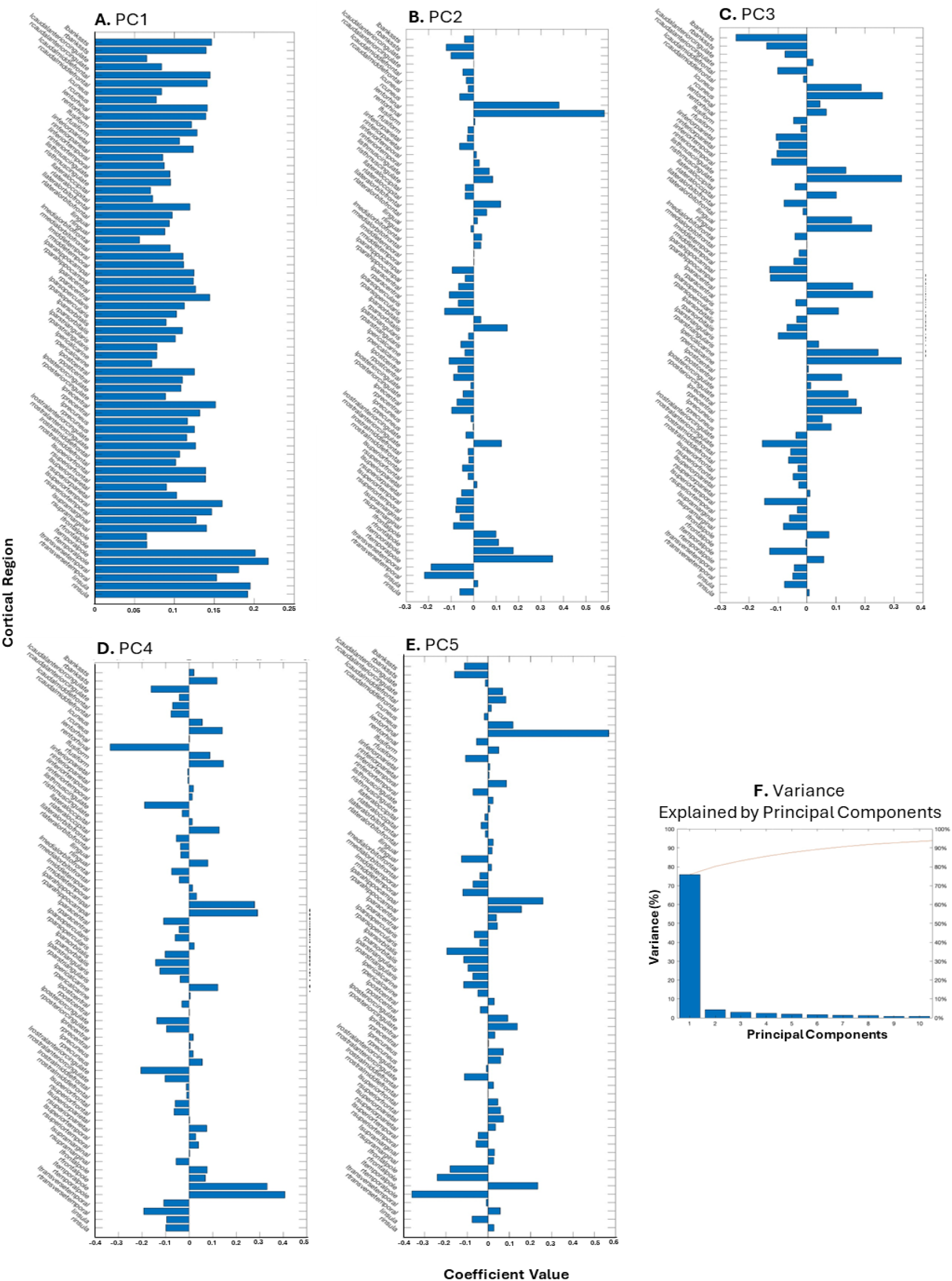
(**A-E**) Coefficient values for PC1 to PC5: Contribution, or weight, of each of 68 cortical areas for each of the first 5 principal components derived from this analysis. As expected, PC1 was formed by a weighted sum of all the 68 cortical areas. As one moved to higher principal components, the contribution of specific cortical areas was highlighted, either through higher or lower PC weights. (**F**) Variance explained by PCs: PC1 alone accounted for more than 75% of the entire variance of cortical thickness observed across the different conditions included in the original database. A final score for a given PC was calculated by the sum of the products of the cortical thickness of each cortical area by the weight assigned to that area in a given PC.

In general terms, we can see a further expansion of the spatial distribution of the cortico-cortical correlations, given that now, in addition to many long-range correlations involving areas in the temporal, frontal and parietal lobes, a much larger contribution involving many cortical areas located in the occipital lobe are clearly present. These included primarily the right lateral occipital area, the right cuneus, and the right lingual gyrus, but also the left pericalcarine region, a pattern that resembles the one observed in healthy subjects. Also relevant is the significant contribution of cortical hubs in the cingulate gyrus, like the right and left isthmus, and the left rostral anterior cingulate. Interestingly, long-range correlations, both ipsilateral and bilateral, responded for the majority of those involving occipital and cingulate hubs. The same can be said for the widespread correlations established by a large number of temporal hubs, like the right and left banks of the superior temporal gyrus, the right inferior and middle temporal lobes, the left superior temporal lobe, and even the right entorhinal and fusiform gyri. This pattern of temporal lobe cortical thickness correlations represented a further expansion of the widespread spatial distribution seen for the same lobe after 9 months of training in the present study. Again, this is in clear contrast to the pattern observed in SCI patients, which included a much larger contribution of correlations among temporal cortical areas in the same and contralateral hemisphere.

Long-range correlations, widely spread in cortical space, also characterized the contributions of hubs in the parietal and frontal lobes. In the case of the former, the right and left superior and inferior lobes, and the right and left supramarginal gyri were the main contributors. Regarding the frontal lobe, despite the great similarity with the pattern observed in the 9-months group of the present study, one notices a further rostral movement of the spatial distribution of hubs contributing to both short and long-range correlations. Similarly, to the 9-months group, the main frontal hubs in both hemispheres included the lateral and medial orbitofrontal gyri, and the pars opercularis and triangularis. In addition, the left pars orbitalis and caudal middle frontal lobe were also important contributors. The more rostral shift was defined by the inclusion of both the right and left frontal poles for the first time in the list of relevant hubs contributing to the distribution of cortico-cortical thickness correlations.

Going in the opposite direction, not only is a reduction seen in the contribution of both the right and left insulas, but also in the correlations among the pre- and postcentral gyri and the paracentral lobule among themselves, as seen in the 9-month WANR-CN group. Instead, these structures exhibited very significant correlations with frontal, temporal, parietal and even occipital cortical areas, illustrating, once more, the broader spatial distribution of these cortico-cortical thickness correlations after 28 months of training with the WANR (WANR-BR group). Confirming the qualitative of cortical thickness observed across the different conditions included in the original database. Thus, using three principal components, we were able to represent close to 80% of the total variance of the data set, indicating that our strategy to reduce 68 dimensions (one for each cortical area analyzed) to a lower dimension space was very successful. This confirms our previous results with pairwise cortico-cortical thickness correlations by showing that such structural changes, as a result of both SCI and training with the WANR protocol, were highly correlated across the entire spatial domain of the neocortex.

As higher components were considered, the contribution of specific cortical areas, or groups of them, became apparent, either through higher or lower individual PC weights. A final score for each PC, for each of the patients involved in the analysis, was then calculated by the sum of the products of the cortical thickness of each cortical area by the weight given to that area in a given PC.

To compare the different groups combined in this analysis, we next built a 3D graph in which each axis represents the final score of each of the first three PCs for each patient, in each of the three periods of training (0, 9 and 28 months) included in this analysis (Figure 21). In this 3D PC space, red dots represent the initial global state of cortical thickness of the 11 SCI patients of the present study, prior to start of training. Green dots represent the cortical thickness score for the same 11 patients after 9 months of training. Magenta dots depict the 3D PC score for the six patients from a separate study who trained for 28 months with the WANR protocol. Finally, blue dots represent the 3D PC scores for healthy control subjects. Once plotted in this 3D space, it is possible to visualize a trend for the patients’ PC scores to move from the leftmost section of the graph (training onset) towards the rightmost part of the graph, where the cluster of PC scores obtained for healthy subjects’ cortical thickness are located, as a function of training time. In other words, as the time of training increased from 0 (red dots) to 9 months (green dots) for the same set of patients, one can see a clear displacement of the PC scores to the right. By adding six patients who trained for 28 months, one can see that the magenta dots, which represent the global cortical thickness state of these patients’ brains, are located further to the right, and much closer to the cluster formed by the PC scores of healthy subjects’ brains.

**Figure 21.**
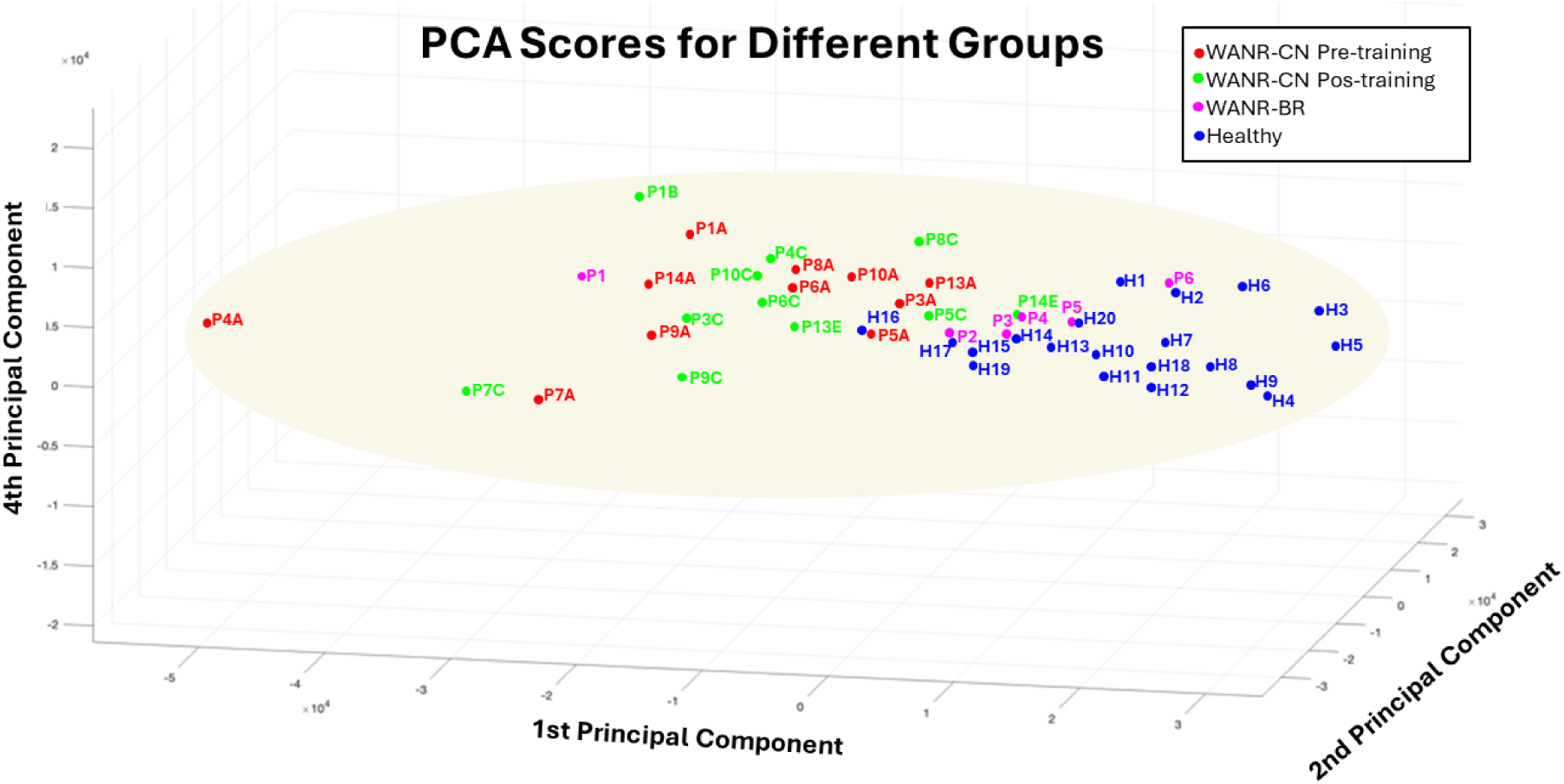
3D graph in which each axis represents the final score of each of the first 3 PCs for each patient, in each of the three periods of training (0, 9 and 28 months). PC1 ranges from -5 to 3; PC2 ranges from -3 to 3; PC4 ranges from -2 to 2 (x10^4^). In this 3D PC space, red dots represent the initial global state of cortical thickness of the 11 WANR-CN SCI patients of the present study, prior to start of training. Green dots represent the cortical thickness score for the same 11 patients after 9 months of training. Magenta dots depict the 3D PC score for the six WANR-BR patients from a separate study who trained for 28 months with the WANR protocol. Finally, blue dots represent the 3D PC scores for healthy control subjects. Once plotted in this 3D space, it is possible to visualize a trend for the patients’ PC scores, which are based on a global measurement of cortical thickness, to move from the leftmost section of the graph (training onset) towards the rightmost part of the graph, where the PC scores obtained for healthy subject’s cortical thickens are located, as a function of training time. In other words, as the time of training increased from 0 (red dots) to 9 months (green dots) for the same set of patients, one can see a clear displacement of the PC scores to the right. By adding six patients who trained for 28 months to the analysis, one can see that the magenta dots, which represent the global cortical thickness state of these patients’ brains, is located further to the right, and much closer to the cluster formed by the PC scores of healthy subjects’ brains.

To further quantify this “global cortical thickness change” in PC space over the 9 months of training, we calculated the vectoral distance between the red and green dot pairs for each of the 11 WANR-CN patients of the present study included in this analysis. These distances are shown in Figure 22. Interestingly, the patient with the best clinical recovery (P4) exhibited the longest displacement in 3D PC space. Likewise, other patients with good clinical recovery, like P8 and P14, also exhibited very prominent trajectories in PC space. Conversely, patients with longer lesion times that did not achieve a great deal of motor recovery had very short trajectories in this PC space (e.g. P7 and P3).

**Figure 22.**
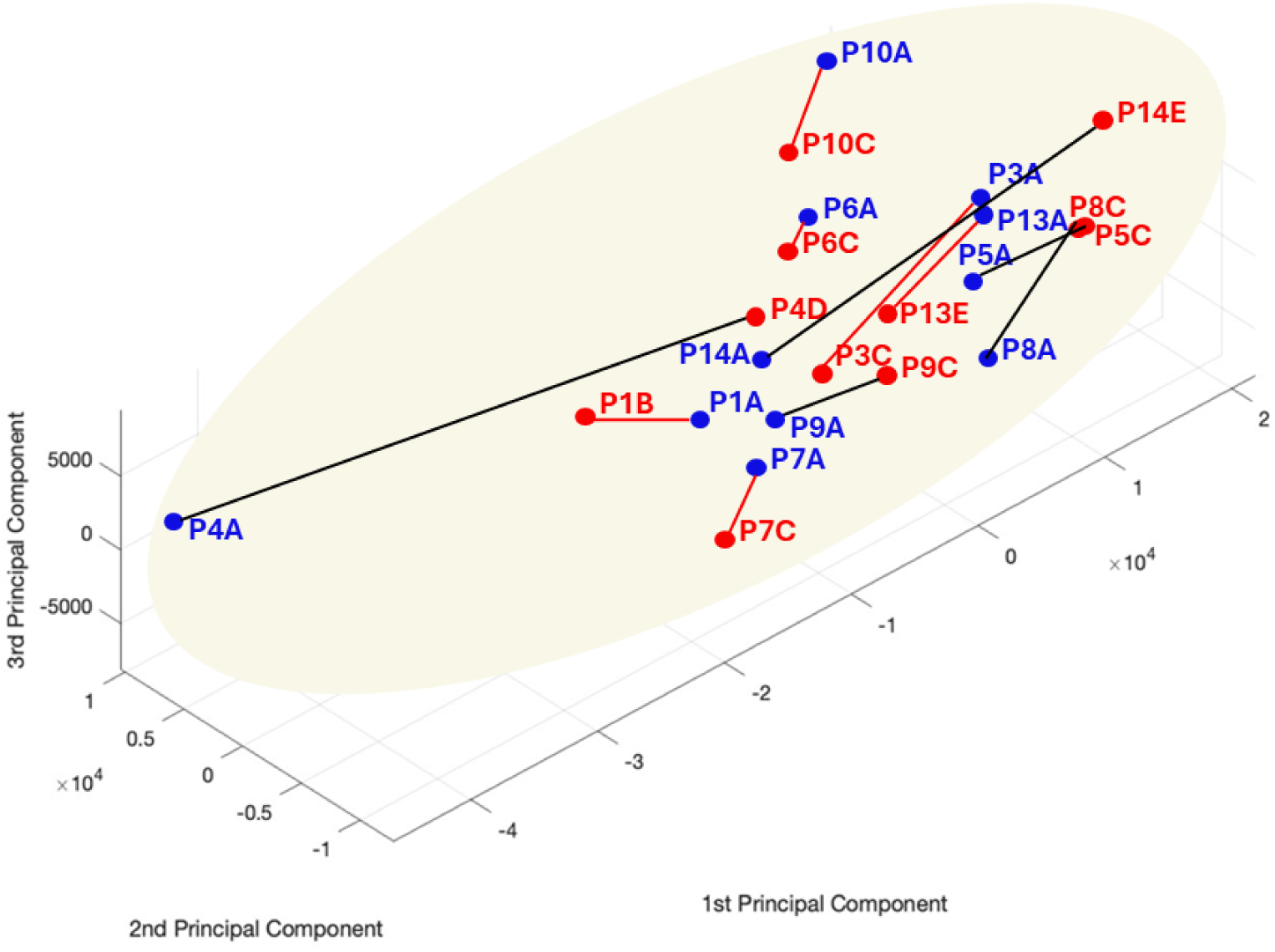
Global cortical thickness changes over 9 months of training in principal component (PC) space. To further quantify this “global cortical thickness change” in PC space over the period of training, we calculated the vectorial distance between the red and green dot pairs for each of the 11 WANR-CN patients included in this analysis.

#### 3.1.1. Functional MRI Results

##### 3.1.1.1. Comparison of Healthy x SCI groups

###### 3.1.1.1.a. Seed-to-Voxel analysis

In order to characterize the functional connectivity of our SCI patients’ brains and compare it to healthy subjects and then those subjected to training with the WANR protocol, we carried out a 2nd level seed-to-voxel analysis for all groups. Chosen seeds were the bilateral precentral gyrus and the sensorimotor network.

In the seed-to-voxel analysis (Fig. 23), SCI patients showed significantly increased functional connectivity between the precentral gyrus and several other brain regions when compared to healthy controls. The most prominent clusters were located in the lateral occipital cortex, occipital fusiform gyrus, precuneus, cingulate gyrus, superior frontal gyrus and cerebellum, with peak uncorrected p-values reaching < 0.000001 and FWE-corrected cluster-level significance in most findings.

**Figure 23.**
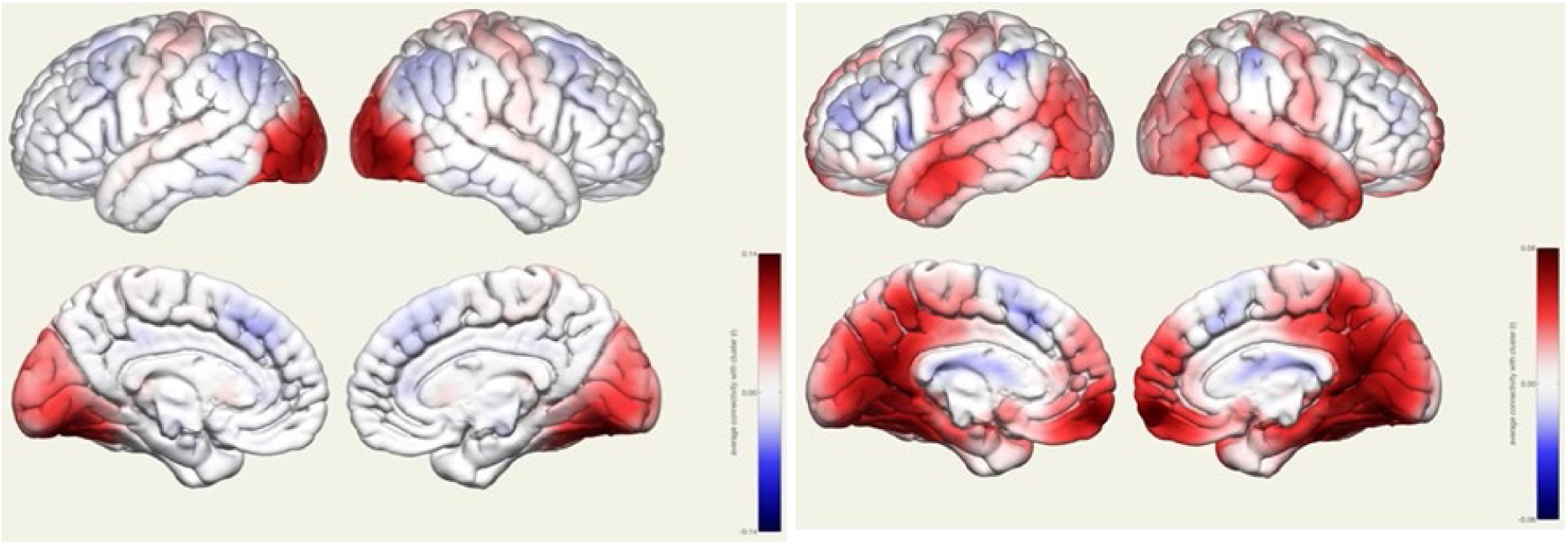
Graphic representation of the results of seed-to-voxel analysis. **SCI** patients showed significantly increased functional connectivity between the **precentral gyrus** and several other brain regions when compared to healthy controls. The most prominent clusters were located in the **lateral occipital cortex, occipital fusiform gyrus, precuneus, cingulate gyrus, superior frontal gyrus and cerebellum**, with peak uncorrected p-values reaching < 0.000001 and FWE-corrected cluster-level significance in most findings. Similarly, the **sensorimotor network** seed revealed enhanced connectivity in SCI patients toward frontal pole, bilateral **posterior parietal**, **motor-associative**, and **visual processing regions**, including the **precuneus**, **supplementary motor area** and **right inferior temporal/fusiform gyrus**, again with strong statistical significance. Red labelling indicates high functional connectivity and Blue low functional connectivity.

Similarly, the sensorimotor network seed revealed enhanced connectivity in SCI patients toward the frontal pole, bilateral posterior parietal, motor-associative, and visual processing regions, including the precuneus, supplementary motor area and right inferior temporal/fusiform gyrus, again with strong statistical significance.

In the WANR > Healthy contrast group, increased connectivity from the precentral gyrus was observed in clusters involving the lingual gyrus, lateral occipital cortex, precuneus, cuneus, occipital pole, and bilateral cerebellum and posterior cingulate, as well as prefrontal, and parietal regions (Fig. 24). The sensorimotor network seed also showed significantly stronger connectivity in the BMI group with the bilateral parietal and occipital cortices, including large clusters at the lingual gyrus (bilateral), intracalcarine cortex, lateral occipital cortex and cerebellum.

**Figure 24.**
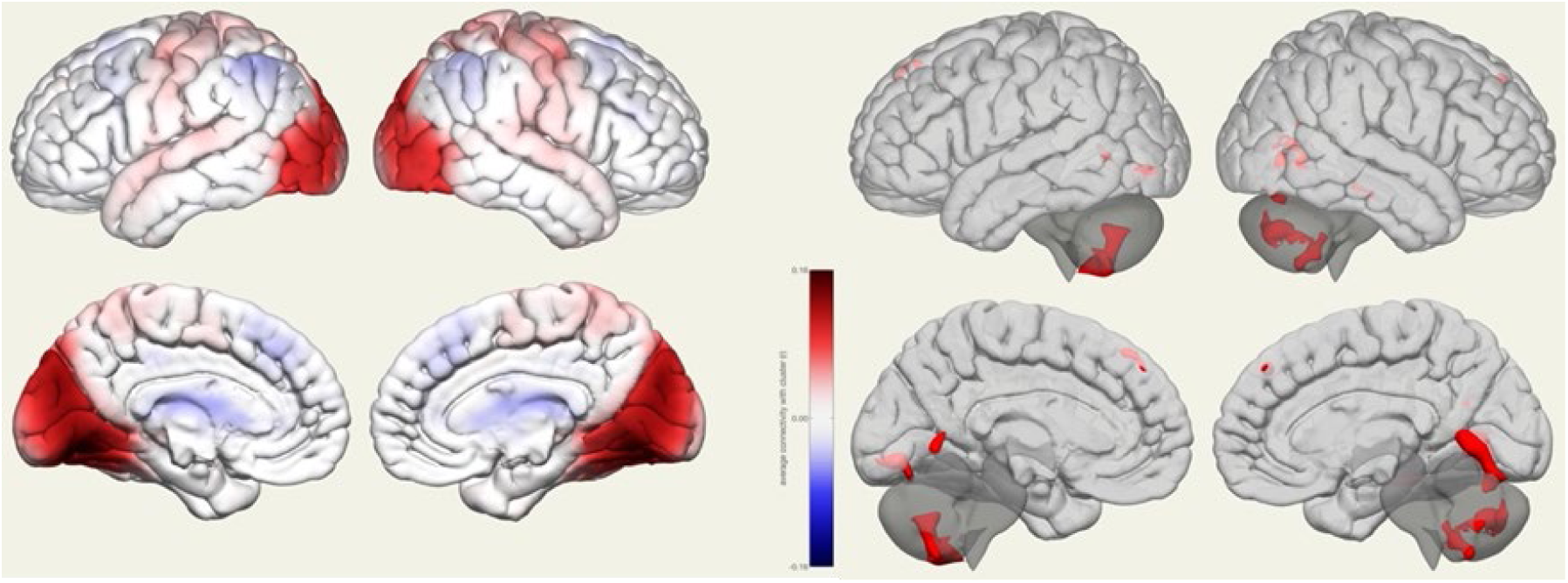
Graphic representation of the results of seed-to-voxel analysis. In the **WANR > Healthy** contrast, increased connectivity from the **precentral gyrus** was observed in clusters involving the lingual gyrus, lateral occipital cortex, precuneus, cuneus, occipital pole and **bilateral cerebellum and posterior cingulate**, as well as prefrontal and parietal regions. The **sensorimotor network** seed also showed significantly stronger connectivity in the WANR group with bilateral **parietal** and **occipital** cortices, including large clusters at the lingual gyrus (bilateral), intracalcarine cortex, lateral occipital cortex and cerebellum. Red labeling indicates high functional connectivity and Blue low functional connectivity.

###### 3.1.1.1.b. ROI-to-ROI analysis

***Healthy versus SCI patients***: A ROI-to-ROI analysis was also conducted using contrasts between SCI, WANR and Healthy Individuals. The comparison between SCI and WANR individuals did not reach significant levels, but the comparison between healthy controls and both groups resulted in several significant differences.

***Connectivity***. This analysis revealed that the most significant hyperconnectivity regions in healthy individuals included the cerebello-cortical and subcortical connections (Fig. 25A). These suggest stronger cerebello-subcortical interactions in healthy individuals. Conversely, in SCI patients, the most significant hyperconnectivity included increased connections of the hippocampus-insula and visual cortex with prefrontal and subcortical regions. These may indicate compensatory reorganization involving limbic, sensory-insular, and visual networks in SCI patients (Fig. 25B).

**Figure 25.**
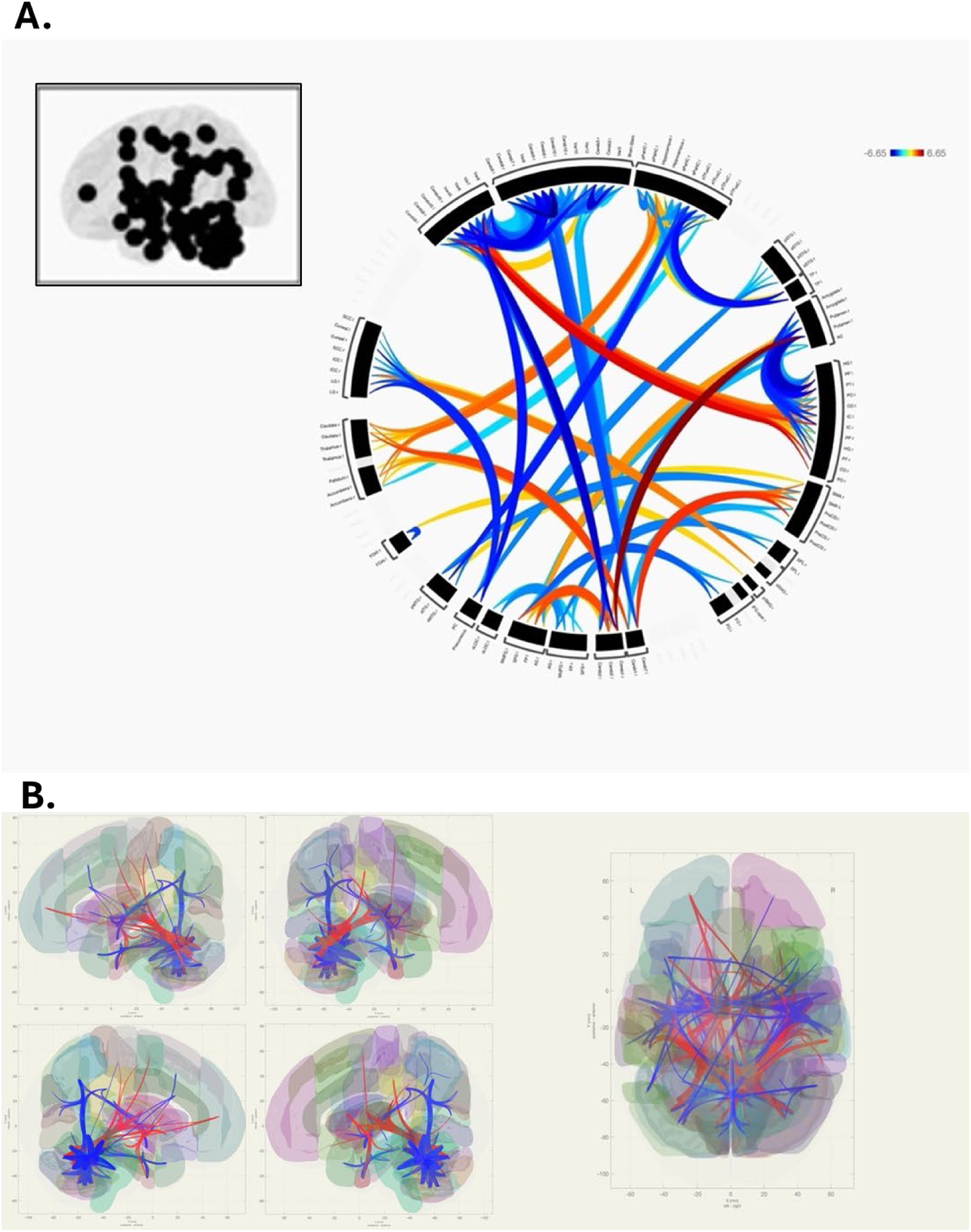
Connectivity of Healthy X SCI patients. Hyperconnected regions in Healthy subjects shown in warm (orange-red) colors and hyperconnected regions in SCI patients shown in cold colors (blue).

Graph-theory analysis revealed no significant group differences considering Global Efficiency, Local Efficiency, Betweenness Centrality, Closeness Centrality, Eccentricity and Clustering Coefficient. However, significant differences were observed in Eigenvector Centrality, Cost, Average Path Length and Degree, as follows below:

***Eigenvector Centrality***. Eigenvector centrality measures how well-connected a node is to other well-connected nodes in the network. It assigns higher importance to nodes that are connected to other central nodes. A brain region with high eigenvector centrality is not only highly connected, but it is particularly connected to other major hubs, indicating its great influence in both information flow and a pivotal role in global network integration. Conversely, decreases in eigenvector may reflect loss of influence of a region within the network (e.g., due to injury or degeneration). In our analysis (Fig. 26A; Tables 16 and 17), Healthy individuals exhibited significantly higher eigenvector centrality in primary visual, somatosensory, and motor cortices, suggesting preserved network integrity in these sensory-motor hubs. In contrast, SCI patients showed higher centrality in temporal regions, cerebellum, and basal ganglia, possibly reflecting compensatory neuroplasticity or network reorganization. Regions such as the posterior cingulate cortex and precuneus, key parts of the Default Mode Network, also appeared more central in SCI, which may relate to increased internal attention or altered baseline network activity after spinal cord injury. The cerebellar and vermis activations point to increased reliance on subcortical/midline structures, a finding that is also consistent with motor system reorganization.

**Figure 26.**
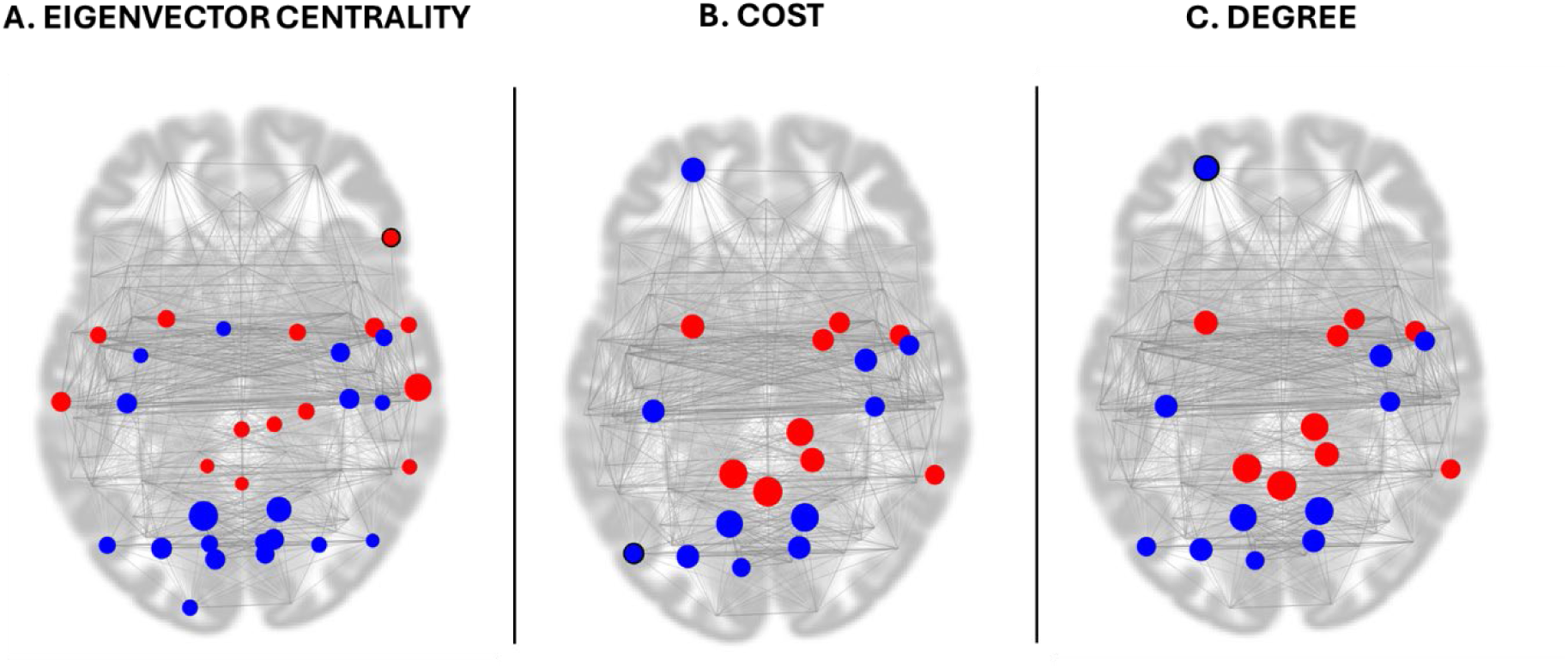
Diagram showing the results for SCI>Healthy subjects (in red) and Healthy>SCI (in blue) for **A.** Eigenvector Centrality, **B.** Cost and **C.** Degree.

**Table 16.** Eigenvector centrality in Healthy>SCI (positive beta).

| **ROI (Full Name)** | **Beta** | **FDR-corrected p-value** | **Functional Interpretation** |
| --- | --- | --- | --- |
| Lingual Gyrus Left (LG l) | 1.36 | 0.000190 | Visual processing – shape recognition |
| Lingual Gyrus Right (LG r) | 1.26 | 0.000698 | Bilateral visual integration |
| Intracalcarine Cortex Right (ICC r) | 1.03 | 0.008322 | Primary visual cortex (V1) |
| Occipital Fusiform Gyrus Left (OFusG l) | 1.13 | 0.008322 | Object and face recognition |
| Postcentral Gyrus Left (PostCG l) | 1.11 | 0.008322 | Primary somatosensory cortex |
| Postcentral Gyrus Right (PostCG r) | 1.05 | 0.009623 | Somatosensory cortex (contralateral limb) |
| Precentral Gyrus Right (PreCG r) | 1.03 | 0.014673 | Primary motor cortex |
| Cuneal Cortex Left (Cuneal l) | 0.92 | 0.015463 | Visual attention |
| Cuneal Cortex Right (Cuneal r) | 0.93 | 0.020167 | Visual integration |
| Central Opercular Cortex Right (CO r) | 0.73 | 0.020167 | Orofacial sensorimotor processing |
| Intracalcarine Cortex Left (ICC l) | 0.91 | 0.020167 | Primary visual cortex |
| Supracalcarine Cortex Right (SCC r) | 0.75 | 0.020167 | Visual association cortex |
| Inferior Lateral Occipital Cortex Left (iLOC l) | 1.02 | 0.020167 | Visual shape/motion detection |
| Supplementary Motor Area Left (SMA l) | 0.62 | 0.020167 | Motor planning |
| Occipital Pole Left (OP l) | 0.75 | 0.024858 | Primary visual pole |
| Parietal Operculum Right (PO r) | 0.80 | 0.025779 | Somatosensory integration |
| Occipital Fusiform Gyrus Right (OFusG r) | 0.90 | 0.024858 | Object recognition |
| Precentral Gyrus Left (PreCG l) | 0.79 | 0.027370 | Motor cortex (left hemisphere) |

**Table 17.**
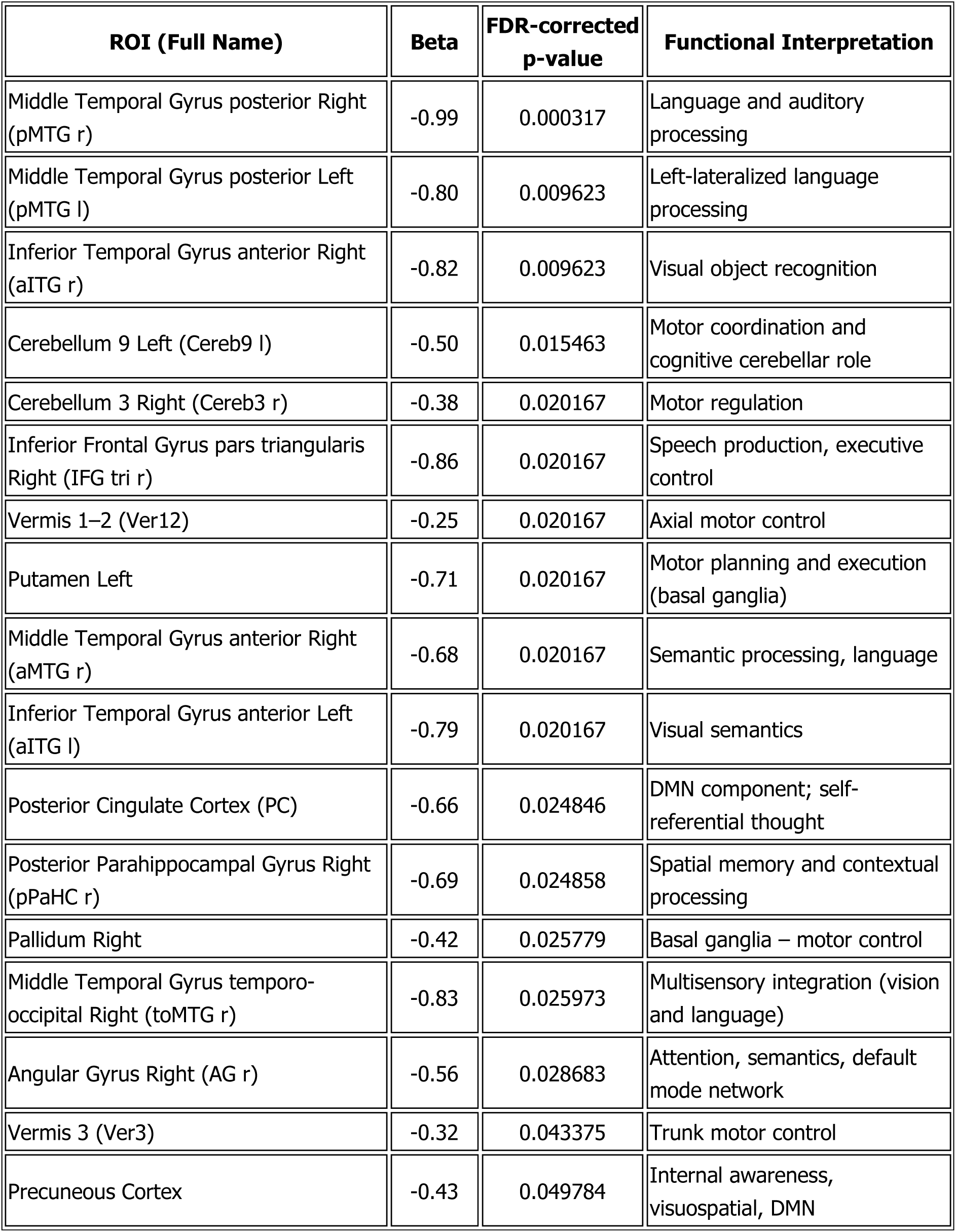
Eigenvector centrality SCI>Healthy (negative beta).

| ROI (Full Name) | Beta | FDR-corrected p-value | Functional Interpretation |
| --- | --- | --- | --- |
| Middle Temporal Gyrus posterior Right (pMTG r) | -0.99 | 0.000317 | Language and auditory processing |
| Middle Temporal Gyrus posterior Left (pMTG l) | -0.80 | 0.009623 | Left-lateralized language processing |
| Inferior Temporal Gyrus anterior Right (aITG r) | -0.82 | 0.009623 | Visual object recognition |
| Cerebellum 9 Left (Cereb9 l) | -0.50 | 0.015463 | Motor coordination and cognitive cerebellar role |
| Cerebellum 3 Right (Cereb3 r) | -0.38 | 0.020167 | Motor regulation |
| Inferior Frontal Gyrus pars triangularis Right (IFG tri r) | -0.86 | 0.020167 | Speech production, executive control |
| Vermis 1–2 (Ver12) | -0.25 | 0.020167 | Axial motor control |
| Putamen Left | -0.71 | 0.020167 | Motor planning and execution (basal ganglia) |
| Middle Temporal Gyrus anterior Right (aMTG r) | -0.68 | 0.020167 | Semantic processing, language |
| Inferior Temporal Gyrus anterior Left (aITG l) | -0.79 | 0.020167 | Visual semantics |
| Posterior Cingulate Cortex (PC) | -0.66 | 0.024846 | DMN component; self-referential thought |
| Posterior Parahippocampal Gyrus Right (pPaHC r) | -0.69 | 0.024858 | Spatial memory and contextual processing |
| Pallidum Right | -0.42 | 0.025779 | Basal ganglia – motor control |
| Middle Temporal Gyrus temporo-occipital Right (toMTG r) | -0.83 | 0.025973 | Multisensory integration (vision and language) |
| Angular Gyrus Right (AG r) | -0.56 | 0.028683 | Attention, semantics, default mode network |
| Vermis 3 (Ver3) | -0.32 | 0.043375 | Trunk motor control |
| Precuneous Cortex | -0.43 | 0.049784 | Internal awareness, visuospatial, DMN |

Cost. Cost refers to the sum of the connection weights a node has. Essentially, it reflects how costly it is to maintain connectivity for that region, based on connection strength or density. Higher cost can suggest greater investment in communication, possibly indicating more active or integrative regions, while lower cost may signal network pruning or underuse, especially in the case of diseases or post-injury states. In our analysis (Fig. 26B; Tables 18 and 19), healthy individuals exhibited significantly higher Cost values in several cortical regions, including the bilateral Lingual Gyrus, Occipital Fusiform Gyrus (left), Intracalcarine Cortex (right), Postcentral Gyrus (bilaterally), and Precentral Gyrus (right), all areas associated with primary sensory, motor, and visual processing. Additionally, higher Cost was observed in the Frontal Pole (left) and Central Opercular Cortex (right), suggesting increased global connectivity in frontal and opercular regions among healthy subjects.

**Table 18.** Cost in Healthy> SCI (positive beta).

| ROI (Full Name) | Beta | T | dof | p-unc | p-FDR |
| --- | --- | --- | --- | --- | --- |
| Lingual Gyrus Right | 0.08 | 4.14 | 31 | 0.000245 | 0.008089 |
| Lingual Gyrus Left | 0.09 | 3.84 | 31 | 0.000568 | 0.014991 |
| Frontal Pole Left | 0.06 | 3.62 | 31 | 0.001047 | 0.019734 |
| Occipital Fusiform Gyrus Left | 0.08 | 3.46 | 31 | 0.001576 | 0.023628 |
| Postcentral Gyrus Left | 0.08 | 3.45 | 31 | 0.001632 | 0.023628 |
| Intracalcarine Cortex Right | 0.06 | 3.29 | 31 | 0.002471 | 0.029652 |
| Precentral Gyrus Right | 0.08 | 3.14 | 31 | 0.003654 | 0.040197 |
| Central Opercular Cortex Right | 0.05 | 3.10 | 31 | 0.004055 | 0.041173 |
| Postcentral Gyrus Right | 0.07 | 2.90 | 31 | 0.006757 | 0.049550 |

**Table 19.** Cost in SCI> Healthy (negative beta).

| ROI (Full Name) | Beta | T | dof | p-unc | p-FDR |
| --- | --- | --- | --- | --- | --- |
| Cerebellum 9 Left | -0.08 | -4.55 | 31 | 0.000078 | 0.007149 |
| Vermis 9 | -0.07 | -4.43 | 31 | 0.000108 | 0.007149 |
| Cerebellum 3 Right | -0.06 | -4.20 | 31 | 0.000210 | 0.008089 |
| Cerebellum 4/5 Right | -0.07 | -3.64 | 31 | 0.000983 | 0.019734 |
| Putamen Left | -0.07 | -3.42 | 31 | 0.001790 | 0.023628 |
| Inferior Temporal Gyrus,<br>anterior division Right | -0.06 | -3.03 | 31 | 0.004876 | 0.043027 |
| Pallidum Right | -0.05 | -3.03 | 31 | 0.004889 | 0.043027 |
| Middle Temporal Gyrus,<br>temporooccipital Right | -0.07 | -2.94 | 31 | 0.006142 | 0.047781 |
| Putamen Right | -0.05 | -2.94 | 31 | 0.006154 | 0.047781 |

Conversely, SCI patients demonstrated greater Cost values in subcortical and cerebellar regions, notably the Putamen (bilateral), Pallidum (right), Cerebellar lobules 3, 4-5 (right), Cerebellar lobule 9 (left), and Vermis 9, as well as in temporal regions such as the Inferior Temporal Gyrus (right) and Temporo-occipital Middle Temporal Gyrus (right). These findings may reflect a compensatory shift in network topology toward subcortical and cerebellar hubs in SCI patients.

***Average Path Length***. The average path length is the mean shortest distance between a node and all other nodes in the network. Shorter average path lengths reflect efficient communication and rapid information transfer across the network. On the other hand, an increase in path length suggests inefficient or disrupted communication, often seen in neurodegeneration or network disintegration.

When comparing the Average Path Length between groups, no brain regions exhibited significantly greater values in the Healthy group relative to SCI. However, the left Postcentral Gyrus showed a significantly increased Average Path Length in SCI patients compared to healthy controls (β = –0.24, T = –4.01, p-FDR = 0.047; Table 20). This suggests that, in individuals with spinal cord injury, this somatosensory region is less efficiently integrated within the overall network, requiring longer paths for communication with other brain areas. Such disruption may reflect impaired information transfer and compromised network organization in sensorimotor processing.

**Table 20.** Average Path Length in SCI> Healthy (negative beta).

| ROI (Full Name) | Beta | T | dof | p-unc | p-FDR |
| --- | --- | --- | --- | --- | --- |
| Postcentral Gyrus Left | -0.24 | -4.01 | 31 | 0.000357 | 0.047151 |

Degree. Degree is the number of direct connections (edges) that a node has to other nodes (Fig. 26C; Tables 21 and 22). A high degree implies that a region is directly communicating with many others, potentially acting as a local hub. A decrease in degree may reflect reduced local connectivity, possibly due to loss of axonal projections or functional disconnection.

**Table 21.** Degree Healthy>SCI.

| ROI (Full Name) | $\beta$ | T | dof | p-unc | p-FDR |
| --- | --- | --- | --- | --- | --- |
| Lingual Gyrus Right | 10.99 | 4.14 | 31 | 0.00025 | 0.00809 |
| Lingual Gyrus Left | 11.25 | 3.84 | 31 | 0.00057 | 0.01499 |
| Frontal Pole Left | 7.86 | 3.62 | 31 | 0.00105 | 0.01973 |
| Occipital Fusiform Gyrus Left | 10.45 | 3.46 | 31 | 0.00158 | 0.02363 |
| Postcentral Gyrus Left | 10.58 | 3.45 | 31 | 0.00163 | 0.02363 |
| Intracalcarine Cortex Right | 8.34 | 3.29 | 31 | 0.00247 | 0.02965 |
| Precentral Gyrus Right | 10.39 | 3.14 | 31 | 0.00365 | 0.04020 |
| Central Opercular Cortex Right | 6.50 | 3.10 | 31 | 0.00406 | 0.04117 |
| Postcentral Gyrus Right | 9.28 | 2.90 | 31 | 0.00676 | 0.04955 |

**Table 22.** Degree SCI>Healthy.

| ROI (Full Name) | $\beta$ | T | dof | p-unc | p-FDR |
| --- | --- | --- | --- | --- | --- |
| Cerebellum 9 Left | -9.88 | -4.55 | 31 | 0.00008 | 0.00715 |
| Vermis 9 | -9.09 | -4.43 | 31 | 0.00011 | 0.00715 |
| Cerebellum 3 Right | -7.77 | -4.20 | 31 | 0.00021 | 0.00809 |
| Cerebellum 4/5 Right | -9.32 | -3.64 | 31 | 0.00098 | 0.01973 |
| Putamen Left | -9.09 | -3.42 | 31 | 0.00179 | 0.02363 |
| Inferior Temporal Gyrus,<br>anterior division Right | -8.28 | -3.03 | 31 | 0.00488 | 0.04303 |
| Pallidum Right | -6.37 | -3.03 | 31 | 0.00489 | 0.04303 |
| Middle Temporal Gyrus,<br>temporooccipital Right | -8.98 | -2.94 | 31 | 0.00614 | 0.04778 |
| Putamen Right | -6.60 | -2.94 | 31 | 0.00615 | 0.04778 |

Group-level analysis of the Degree centrality metric also revealed significant differences between Healthy participants and individuals with spinal cord injury. Specifically, several regions exhibited a significantly higher Degree in the Healthy group, indicating greater overall connectivity. These regions included the Lingual Gyrus (bilaterally), Postcentral Gyrus (bilaterally), Precentral Gyrus (Right), Occipital Fusiform Gyrus (Left), Intracalcarine Cortex (Right), Frontal Pole (Left), and Central Opercular Cortex (Right). All these regions survived false discovery rate (FDR) correction with p-FDR < 0.05, suggesting robust group-level effects.

The SCI group, however, showed higher Degree in subcortical and cerebellar structures, such as the Cerebellum 9 (Left), Vermis 9, Cerebellum 3 and 4/5 (Right), Putamen (bilaterally), Pallidum (Right), and temporal regions including the Inferior Temporal Gyrus (anterior division, Right) and Middle Temporal Gyrus (temporooccipital part, Right). These findings suggest a potential compensatory or reorganization pattern in SCI, with increased connectivity in deep and posterior regions.

###### Healthy vs WANR patients

***Connectivity***. In the comparison between WANR and Healthy subjects, several brain regions demonstrated significant differences in connectivity strength (Fig. 27). Notably, WANR subjects exhibited increased connectivity between multiple cerebellar and vermal regions. Among the most robust effects:

- Pallidum (Left) showed significantly higher connectivity with the Vermis 3 in WANR subjects (T = 7.03, p-FDR = 0.000024).
- Enhanced cerebellar-cerebellar connectivity was also evident, such as between Cerebellum Crus II (Left) and Cerebellum 9 (Right) (T = 6.45), and Cerebellum 8 (Left) with Cerebellum 9 (Left) (T = 6.42).
- Connectivity within vermal regions, including Vermis 7 and Vermis 8, and between Cerebellum 9 and Vermis 8, also showed increased strength in the WANR group.

**Figure 27.**
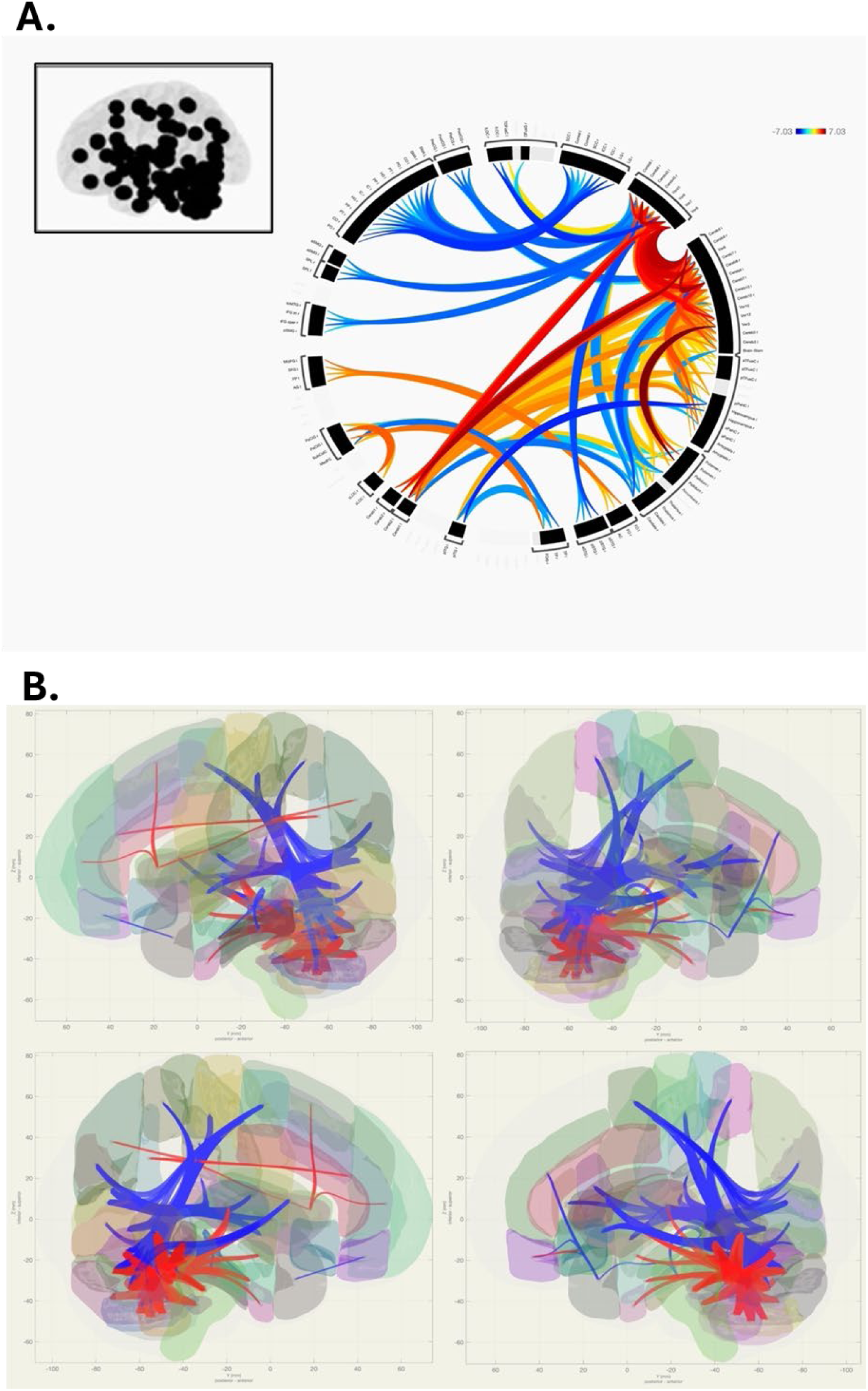
Connectivity between WANR>Healthy areas in warm (yellow, orange and red) colors and Healthy>WANR areas in cold colors (blue).

These results suggest that WANR-related changes may involve stronger cerebellar and midline (vermis) integration, which could reflect adaptive or compensatory mechanisms in motor, balance, or cognitive control networks.

Graph-theory analysis for WANR x Healthy groups revealed no significant group differences in Local Efficiency, Betweeness Centrality, Closeness Centrality, Eccentricity, Average Path Length and Clustering Coefficient, but showed significant differences in Global Efficiency, Eigenvector Centrality, Cost, and Degree, as follows:

***Global Efficiency***. Global Efficiency is a graph theory metric that reflects how efficiently information is exchanged across the entire brain network. It is inversely related to the average shortest path length between all pairs of nodes; higher values indicate more efficient communication across the network.

In the comparison of global efficiency between WANR patients and healthy controls, (Fig. 28A; Table 23) two cerebellar regions showed significantly greater global efficiency in the WANR group after FDR correction. These regions include the left Cerebellum 9 and the right Cerebellum 4/5, suggesting that information is transferred more efficiently across these areas in WANR compared to healthy subjects.

**Figure 28:**
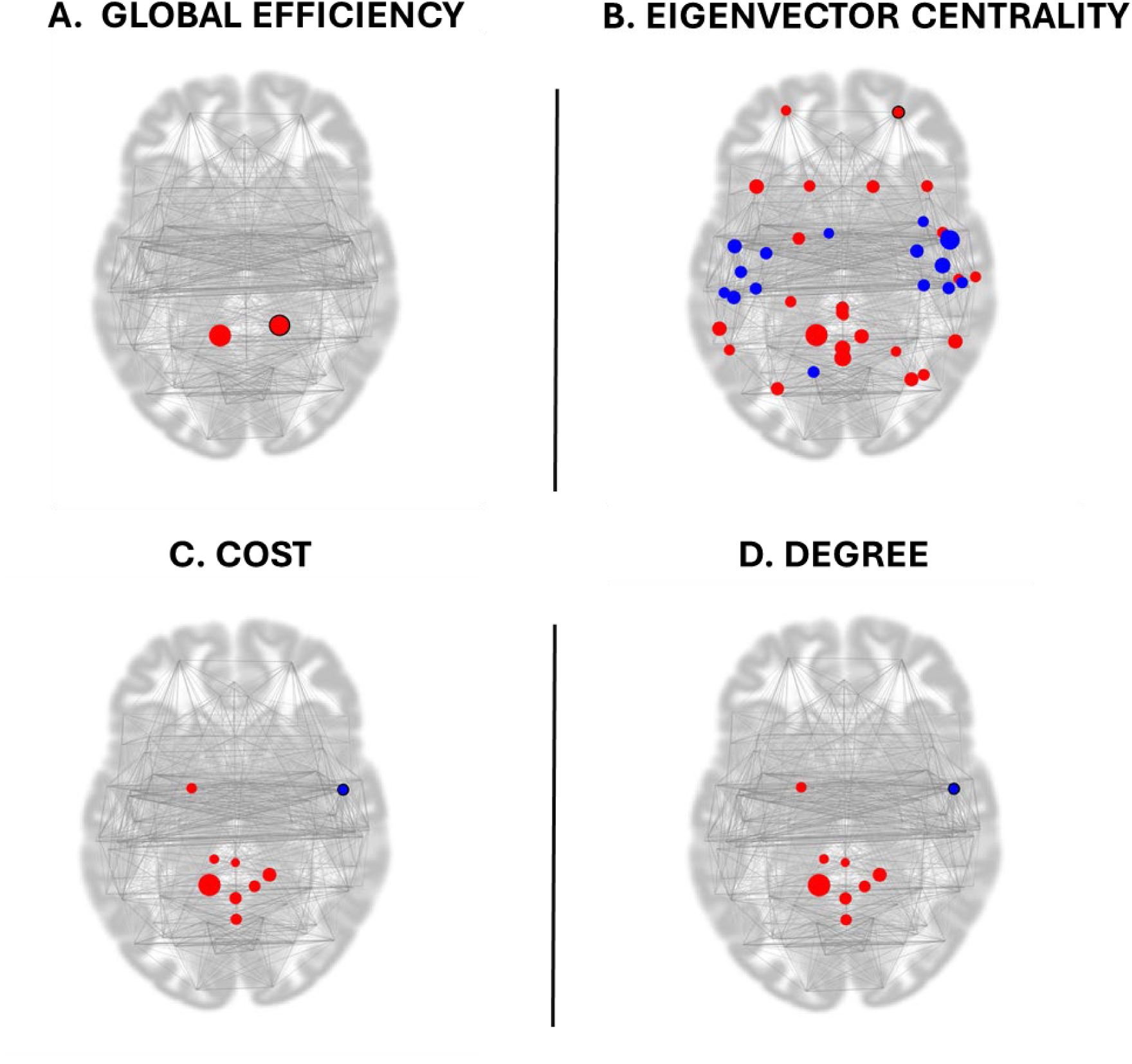
Global Efficiency in Cerebellar networks of WANR patients have significantly higher values than Healthy subjects. Eigenvector Centrality, Cost and Degree WANR>Healthy (red) and Healthy>WANR (blue).

**Table 23.** Global efficiency WANR>Healthy.

| ROI (Full Name) | Beta | T | dof | p-unc | p-FDR | MNI Coordinates (x, y, z) |
| --- | --- | --- | --- | --- | --- | --- |
| Cerebellum 9 Left | 0.13 | 4.39 | 26 | 0.000166 | 0.021926 | (-11, -49, -46) |
| Cerebellum 4/5 Right | 0.07 | 4.02 | 26 | 0.000445 | 0.029343 | (16, -44, -19) |

***Eigenvector Centrality*.** In the comparison between WANR and Healthy subjects (Fig. 28B; Tables 24 and 25), several regions showed significantly altered Eigenvector Centrality, reflecting differences in the influence of these nodes within the overall functional network. Regions with significantly higher centrality in WANR subjects included bilateral Cerebellum 9, Precuneous Cortex, Vermis 9, Middle Frontal Gyrus (left), Angular Gyrus (right), Supramarginal Gyrus (posterior left), Cerebellum Crus II (bilaterally), and the posterior division of the Cingulate Gyrus. Conversely, regions with higher centrality in Healthy controls included the Central Opercular Cortex (bilateral), Heschl’s Gyrus (right), Parietal Operculum Cortex (left), and Precentral Gyrus (right). These results indicate a distributed pattern of network reorganization involving both cortical and cerebellar areas.

**Table 24:** Eigenvector centrality WANR>Healthy regions.

| ROI | $\beta$ (Beta) | T | p-unc | p-FDR |
| --- | --- | --- | --- | --- |
| Cerebellum 9 Left | 1.08 | 5.60 | 0.000007 | 0.000909 |
| Precuneous Cortex | 0.76 | 4.26 | 0.000239 | 0.010499 |
| Vermis 9 | 0.70 | 3.90 | 0.000609 | 0.016082 |
| Middle Frontal Gyrus Left | 0.87 | 3.70 | 0.001007 | 0.017293 |
| Angular Gyrus Right | 0.91 | 3.66 | 0.001141 | 0.017293 |
| Cerebellum 9 Right | 0.90 | 3.64 | 0.001181 | 0.017293 |
| Supramarginal Gyrus (posterior) Left | 0.98 | 3.63 | 0.001220 | 0.017293 |
| Cerebellum Crus II Right | 0.88 | 3.54 | 0.001538 | 0.018459 |
| Cerebellum Crus II Left | 0.93 | 3.32 | 0.002671 | 0.025179 |
| Cingulate Gyrus Posterior | 0.72 | 3.28 | 0.002923 | 0.025292 |

**Table 25:** Eigenvector centrality Healthy > WANR regions.

| ROI | $\beta$ (Beta) | T | p-unc | p-FDR |
| --- | --- | --- | --- | --- |
| Central Opercular Cortex Right | -1.17 | -4.96 | 0.000037 | 0.002462 |
| Heschl's Gyrus Right | -1.14 | -4.00 | 0.000461 | 0.015229 |
| Central Opercular Cortex Left | -0.93 | -3.60 | 0.001310 | 0.017293 |
| Parietal Operculum Cortex Left | -0.88 | -3.45 | 0.001904 | 0.020941 |
| Precentral Gyrus Right | -1.14 | -3.42 | 0.002085 | 0.021167 |

Cost. Cost analysis comparing WANR subjects with healthy controls (Fig. 28C; Tables 25 and 26) revealed that several brain regions exhibited significantly higher values in the WANR group. The most prominent findings were observed in the Cerebellum, particularly in the Cerebellum 9 (left and right), Cerebellum 4/5 Right, and Cerebellum 3 Left, suggesting enhanced connectivity density in these areas. Additionally, Vermis regions (Vermis 1/2, 8, and 9) and the Pallidum Left also showed increased Cost in WANR subjects. In contrast, only the Central Opercular Cortex Right displayed significantly higher Cost in healthy controls compared to WANR participants.

**Table 26:** Cost WANR>Healthy.

| ROI | $\beta$ (Beta) | T | p-unc | p-FDR |
| --- | --- | --- | --- | --- |
| Cerebellum 9 Left | 0.14 | 8.15 | 0.000000 | 0.000002 |
| Cerebellum 4/5 Right | 0.10 | 5.08 | 0.000027 | 0.001814 |
| Vermis 9 | 0.09 | 4.48 | 0.000131 | 0.005780 |
| Cerebellum 9 Right | 0.10 | 4.29 | 0.000216 | 0.007124 |
| Vermis 8 | 0.10 | 4.14 | 0.000321 | 0.008478 |
| Pallidum Left | 0.04 | 3.84 | 0.000716 | 0.013507 |
| Cerebellum 3 Left | 0.05 | 3.55 | 0.001507 | 0.024866 |
| Vermis 1/2 | 0.04 | 3.26 | 0.003076 | 0.045112 |

Degree. Analysis of the Degree metric (Fig. 28D; Tables 27 and 28) revealed significantly higher values in BMI participants in multiple regions, predominantly within the cerebellar and vermal structures. Notably, Cerebellum 9 (left and right) and Cerebellum 4/5 Right showed the strongest effects, alongside Vermis 8, 9, and 1/2, and Cerebellum 3 Left. The Pallidum Left also presented increased Degree in BMI individuals. Conversely, the Central Opercular Cortex Right displayed significantly higher Degree in healthy participants.

**Table 27:**
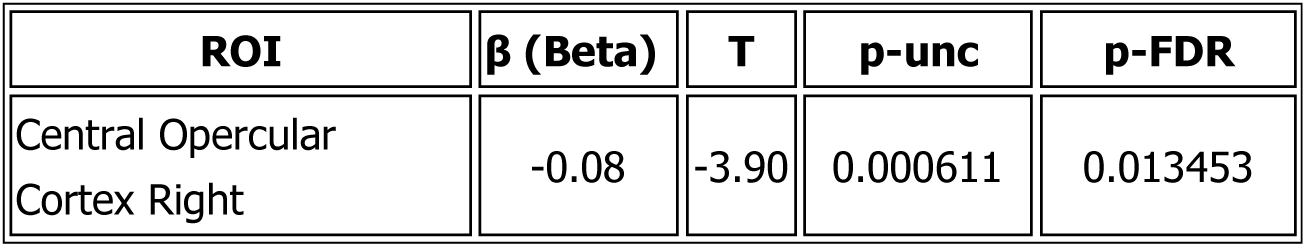
Cost Healthy>WANR.

**Table 28.** Degree WANR>Healthy.

| ROI | $\beta$ (Beta) | T | p-unc | p-FDR |
| --- | --- | --- | --- | --- |
| Cerebellum 9 Left | 18.30 | 8.15 | <0.000001 | 0.000002 |
| Cerebellum 4/5 Right | 13.17 | 5.08 | 0.000027 | 0.001814 |
| Vermis 9 | 11.52 | 4.48 | 0.000131 | 0.005780 |
| Cerebellum 9 Right | 13.23 | 4.29 | 0.000216 | 0.007124 |
| Vermis 8 | 13.13 | 4.14 | 0.000321 | 0.008478 |
| Pallidum Left | 4.82 | 3.84 | 0.000716 | 0.013507 |
| Cerebellum 3 Left | 6.00 | 3.55 | 0.001507 | 0.024866 |
| Vermis 1/2 | 5.70 | 3.26 | 0.003076 | 0.045112 |

**Table 29.** Degree Healthy>BMI.

| ROI | $\beta$ (Beta) | T | p-unc | p-FDR |
| --- | --- | --- | --- | --- |
| Central Opercular Cortex Right | -10.10 | -3.90 | 0.000611 | 0.013453 |

###### 3.1.1.1.c. Principal Component Analysis of Functional MRI data

Given the usefulness and clarity provided by applying PCA to the entirety of our structural MRI data, we employed the same approach to the functional MRI data collected from the SCI patients analyzed in the present clinical trial, before and after training. To build these PC spaces (Figure 29), we considered data representing a total of 877 traditional functional metrics extracted from the functional MRI data for all SCI patients. A little over 50% of the total variance of the fMRI data was accounted for by the first five principal components, while the first 10 PCs accounted for about 73% of the total variance. In this 3D PC space, blue dots and labels represent patients at the onset of training, while red dots and labels depict the same patients after 9 months of training. Arrows connect each patient’s initial and final location in these 3D PC spaces in order to depict the distances covered in fMRI PC space during training. Interesting enough, an even clearer illustration of the effect of the WANR training was obtained in this analysis since the blue dots tended to be located in the outer space of an imaginary sphere in this PC space, while red dots were somewhat clustered in an inner location of this sphere. Thus, patients who underwent very significant clinical recoveries, like P4, P14, P6, P8, and P13, exhibited arrows that originated in outer blue dots (beginning of training) and ended up in more inner red dots (9 months of training). Patients with no significant clinical recovery, like P7, tended to have blue and red dots close to each other.

**Figure 29.**
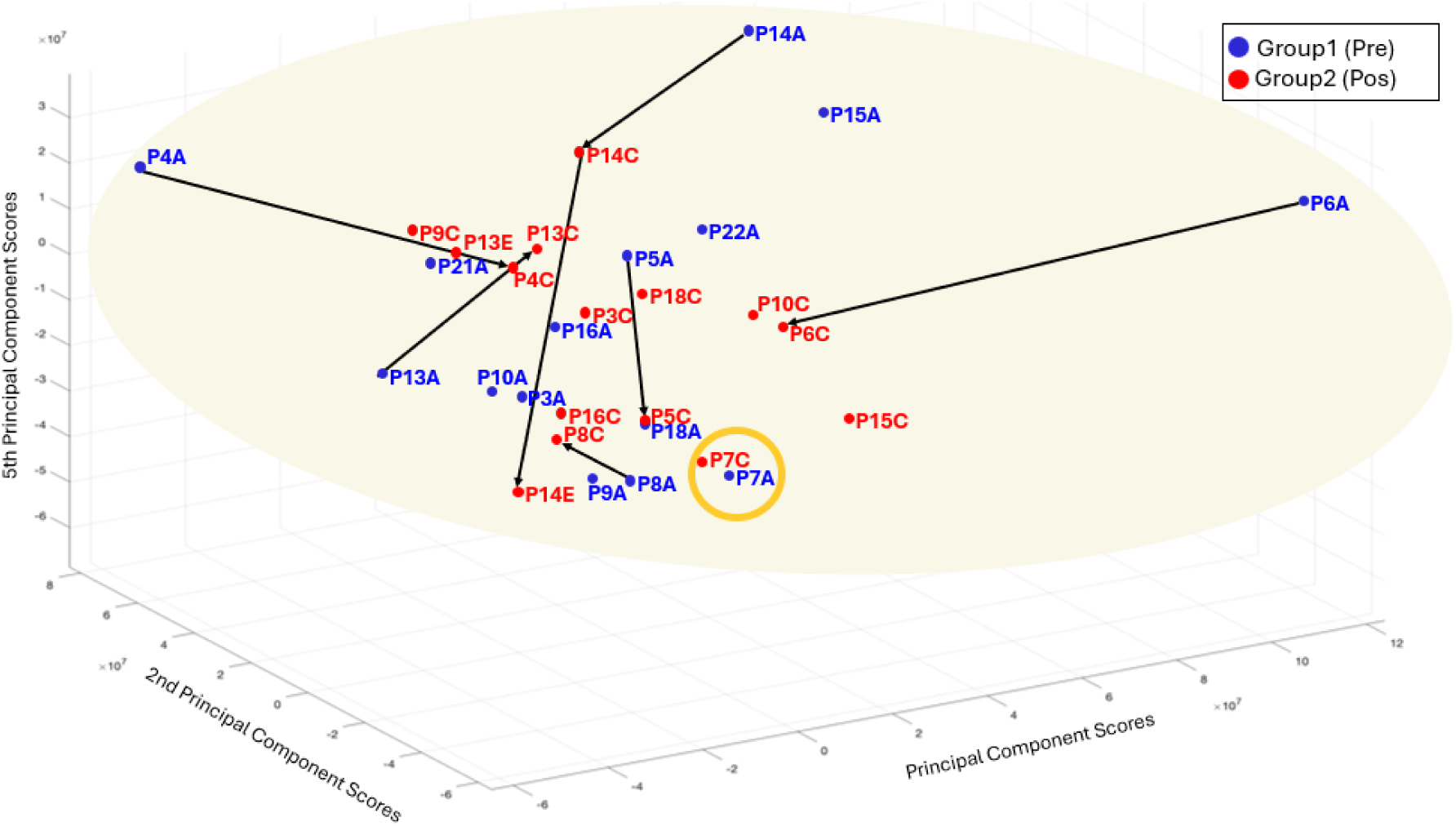
3D PCA space derived from functional MRI data. PC scores range is -6 to 12; 2^nd^ PC scores range is -6 to 8; 5^th^ PC scores range is -6 to 3 (x10^7^). Blue dots and labels represent patients at the onset of training, while red dots and labels depict the same patients after 9 months of training. Arrows connect each patient’s initial and final location in these 3D PC spaces in order to depict the distances covered in this fMRI PC space during training. Patients who underwent very significant clinical recoveries, like P4, P14, P6, P8, and P13 exhibited longer arrows, i.e. travelled more in the PC space, from the beginning to the end of training. Patients with no significant clinical recovery, like P7 (orange circle), tended to have blue and red dots close to each other, and hence shorter arrows.

Next, we analyzed potential changes in cortico-cortical functional connectivity as a result of training. Figure 30A displays a few pairwise cortico-cortical connections that were enhanced after 9 months of training. They included, for instance: (1) a connection between the right postcentral and the lingual gyrus (Fig. 30A); (2) the right Hershl and the precentral gyrus (Fig. 30B); (3) the right posterior inferior temporal gyrus and the right cuneo and lateral occipital cortex (Fig. 30C); (4) the left lingual and left postcentral gyri (Fig. 30D); (5) the right lingual and left postcentral and reduction with the cerebellar vermis (Fig. 30E); (6) the increase between the left lateral occipital cortex and the cuneus and the occipital pole (Fig. 30F); (7) the left par opercularis and the cuneus and the lingual gyrus (Fig. 30G); and (8) the cerebellar vermis and left precentral gyrus and right superior temporal gyrus (Fig. 30H).

**Figure 30.**
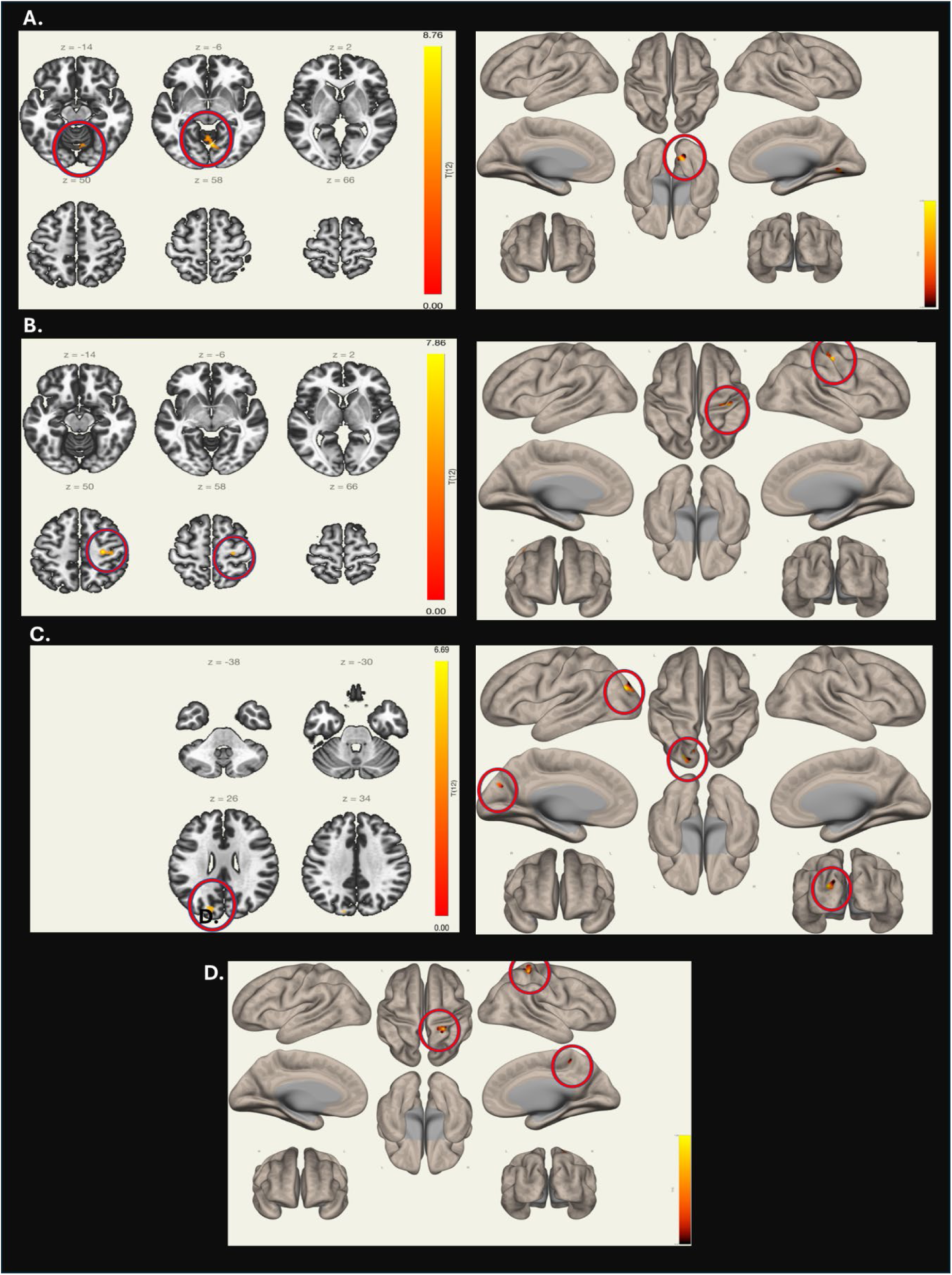
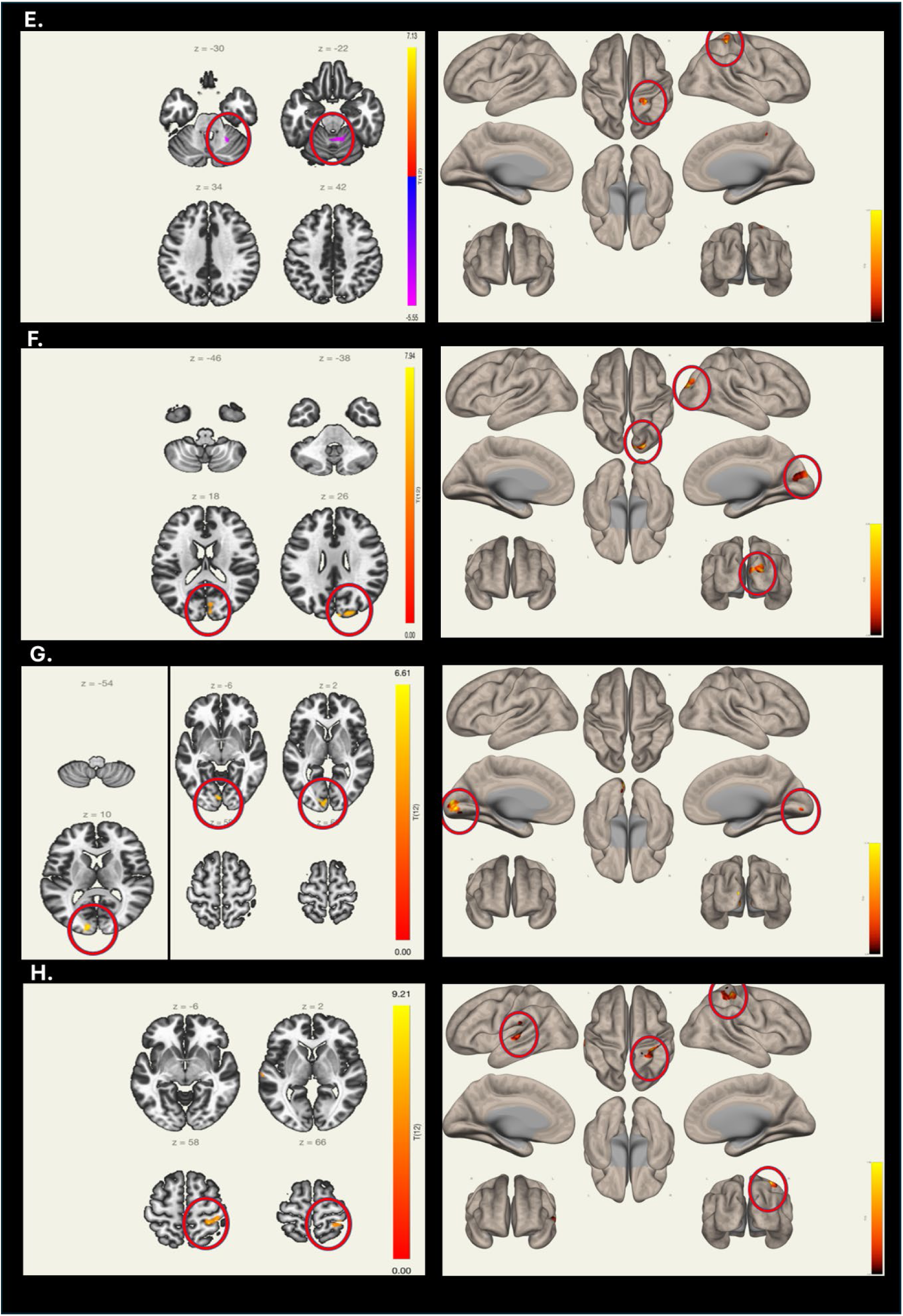
Changes in cortico-cortical connectivity as a result of training. (**A**) Connection between the right postcentral and the lingual giry, (**B**) the right Hershl and the precentral gyri, (**C**) right posterior inferior temporal gyrus and the right cuneo and lateral occipital cortex, (**D**) the left lingual and left postcentral gyri. (**E**) right lingual and left postcentral and reduction with the cerebellar vermis, (**F**) increase between the left lateral occipital cortex and the cuneus and the occipital pole, (**G**) left par opercularis and the cuneus and the lingual gyrus, and (**H**) the cerebellar vermis and left precentral gyrus and right superior temporal gyrus.

Figure 31 summarizes this enhancement in cortico-cortical connectivity highlighting that most of it involved visual cortical areas. This matches nicely with our structural MRI data that highlighted that both a great deal of the reduction in cortical thickness following a SCI and the recovery following training with the WANR protocol involved visual cortical areas located in the occipital lobe, but also in frontal, parietal and temporal regions.

**Figure 31.**
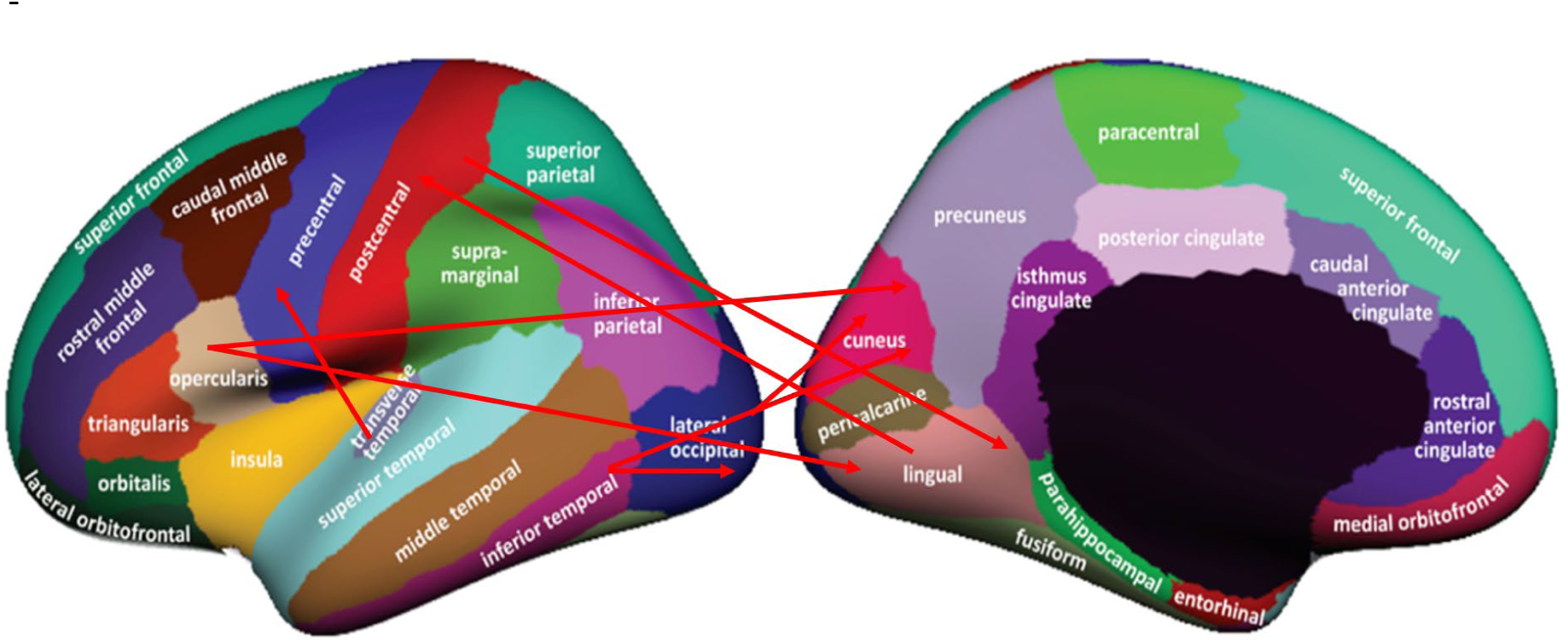
Summary of enhancement in cortico-cortical connectivity highlighting that most of this enhancement in connectivity involved visual cortical areas.

### 3.2. ERP Analysis

Visual ERPs, collected from both a group of healthy controls (n=21) and SCI patients (n=17) were compared (Figure 32). Red traces represent a signal from the control group, while blue trace depicts the equivalent recording obtained for the SCI group. For each electrode location, a yellow bar depicts a recording period interval in which a significant difference was observed between the control and the SCI group, based on an independent sample t-test.

**Figure 32.**
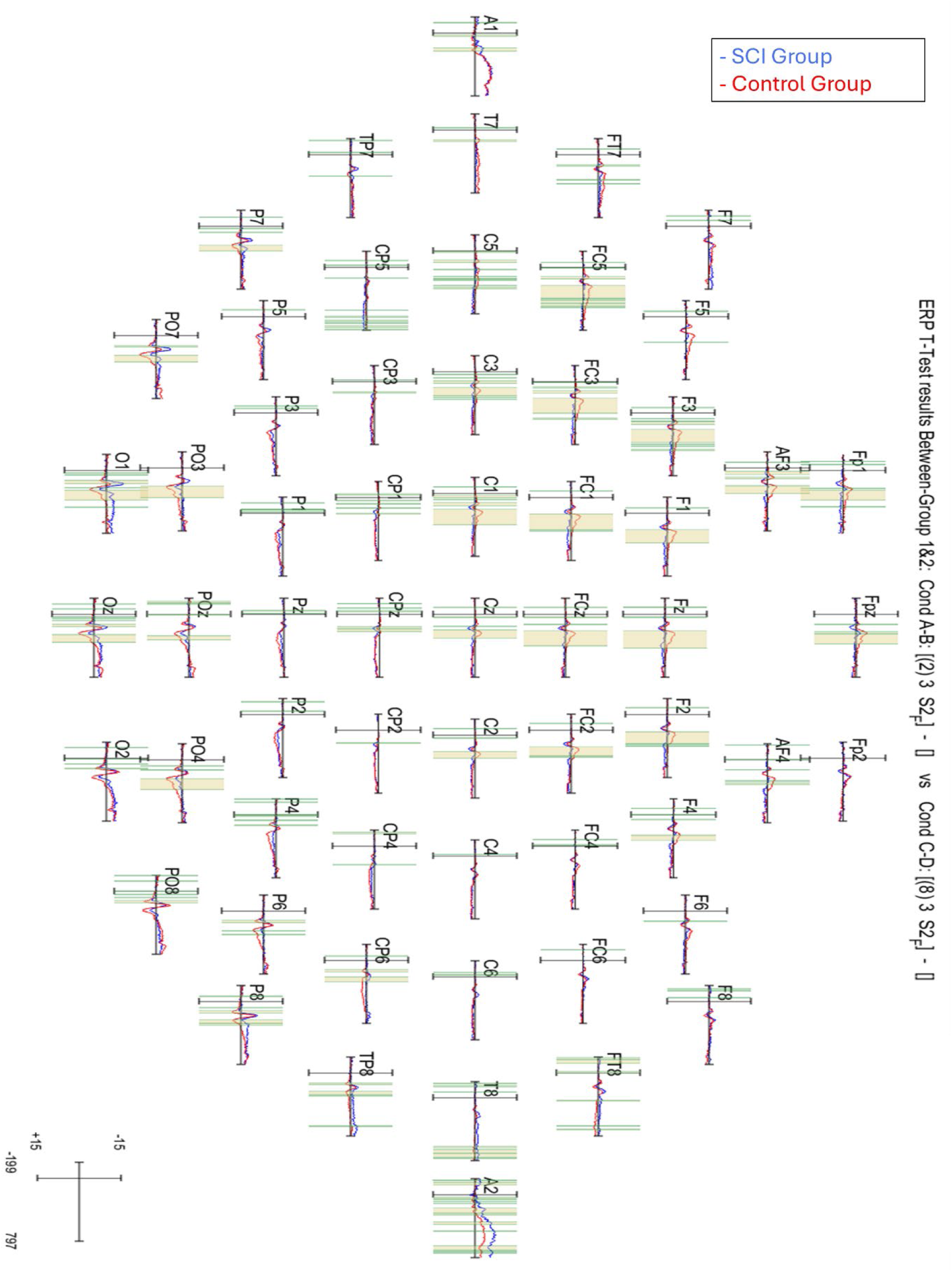
Visual ERPs collected from both a group of healthy controls (n=21, red traces) and SCI patients (n=17, blue traces). For each electrode location, a yellow bar depicts a recording period interval in which a significant difference was observed between the control and the SCI group, based on independent sample t-test.

Table 30 displays the significant differences observed between the two groups when considering the amplitude and latency of P1, the latency of P2, and N2 average magnitude and latency. Thus, while in the control group the P1 average amplitude during the S2 presentation was 2.11±3.15 μV, in the SCI group it was reduced to 0.56±0.53μV (t= 2.209, p<0.05). Average P1 latency in the control was 124.66±10.58 ms, while in the SCI group it was 111.44±8.55 ms (t = 4.16, p<0.001). Regarding the P2 latency during S1, in the control group it was 164.08±14.7ms, while for the SCI group it was 186.22±12.64 (t= -4.908, p <0.001). For N2 average amplitude and latency, the values were the following: -1.13±1.65μV for the control group vs 0.23±0.88 μV for the SCI group (t= -3.059, p<0.01); N2 control group latency = 247.89±16.84ms versus 283.52±16.51ms for SCI group (t= -6.542, p<0.001).

**Table 30.** Significant differences observed between a group of healthy control and SCI patients when considering the amplitude and latency of P1, the latency of P2, and N2 average magnitude and latency.

|  |  | Stimulate | Control group | SCI group | t | p |
| --- | --- | --- | --- | --- | --- | --- |
| P1 | Amplitude | S2 | $2.11 \pm 3.15 \text{mV}$ | $0.56 \pm 0.53 \text{mV}$ | 2.209 | < 0.05 |
| | Latency | S2 | $124.66 \pm 10.58$ | $111.44 \pm 8.55$ | 4.164 | < 0.001 |
| P2 | Latency | S1 | $164.08 \pm 14.7$ | $186.22 \pm 12.64$ | -4.91 | < 0.001 |
| N2 | Amplitude | B12 | $-1.13 \pm 1.65 \text{mV}$ | $0.23 \pm 0.88 \text{mV}$ | -3.06 | < 0.01 |
| | Latency | B12 | $247.89 \pm 16.84$ | $283.52 \pm 16.51$ | - 6.54 | < 0.001 |

We also tested whether we could detect changes in particular electrodes during different phases of the visual task. This analysis revealed that the frontal P1 amplitude during S2 stimulation was significantly different between the two groups. Repeated measurement analysis of variance was performed on the average amplitude of FZ, PZ, FC3, and FC4 under S2 stimulation, and the main effect of the electrodes was significant (F ₍₃, ₃₆₎=4.566, P<0.05), η²= 0.113). The average amplitude of PZ is significantly different from FZ (p<0.05) and FZ (p<0.05). The main effect of the group was significant, and the control group was significantly higher than the SCI group (p<0.05).

Repeated measurement analysis of variance was performed on the average amplitude of FZ, PZ, CP3, and CP4 under the stimulation produced by images of buildings, and the main effect of the electrodes was significant (F ₍₃, ₃₆₎=3.860, P<0.05), η²= 0.097). There was also a significant difference in the average amplitude between FZ and CP3 (p<0.01) and CP4 (p<0.01). The main effect of the group is not significant, and the interaction effect is not significant.

For a subgroup of seven SCI patients who underwent training with the WANR, we were able to measure visual ERPs at three different moments of the protocol: starting point (V0), mid-point (V1, 3-6 months after onset of training), and end point (V2, 9 months after training onset). Longitudinal analysis revealed that from V0 to V2, in four out of the seven patients in which the midpoint ERP measurement (V1) was made (between 5-6 months after onset of training), a very large increase in visually evoked ERP magnitude was observed across most of the electrodes, with particular emphasis on the frontal and parieto-occipital regions. For instance, patient P14 (V1=5 months) exhibited a 7-fold increase, patient P7 (V1=5 months) a 5-fold increase, P6 (V1= 6 months) a 10-fold increase, and patient P8 (V1= 6months) a 6- fold increase (See Figure 33). In three patients who were measured between 3-5 months into training with the WANR, much smaller enhancements in ERP were detected. By the end of the protocol (V2=9 months), these increases in ERP magnitude had reversed somewhat, becoming much smaller than observed at V1, but somewhat higher than V0 and even the control group (Figure 33C).

**Figure 33.**
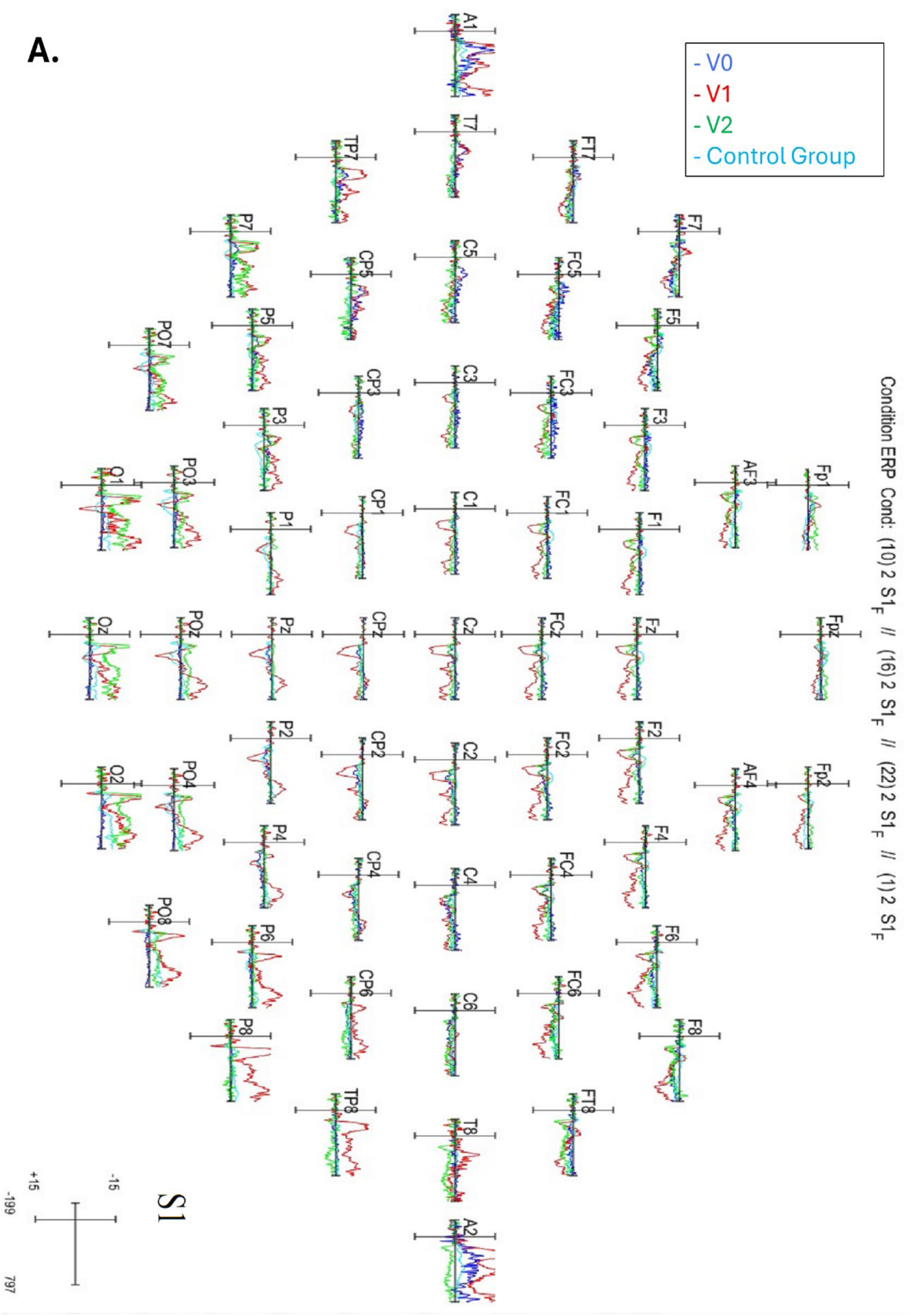
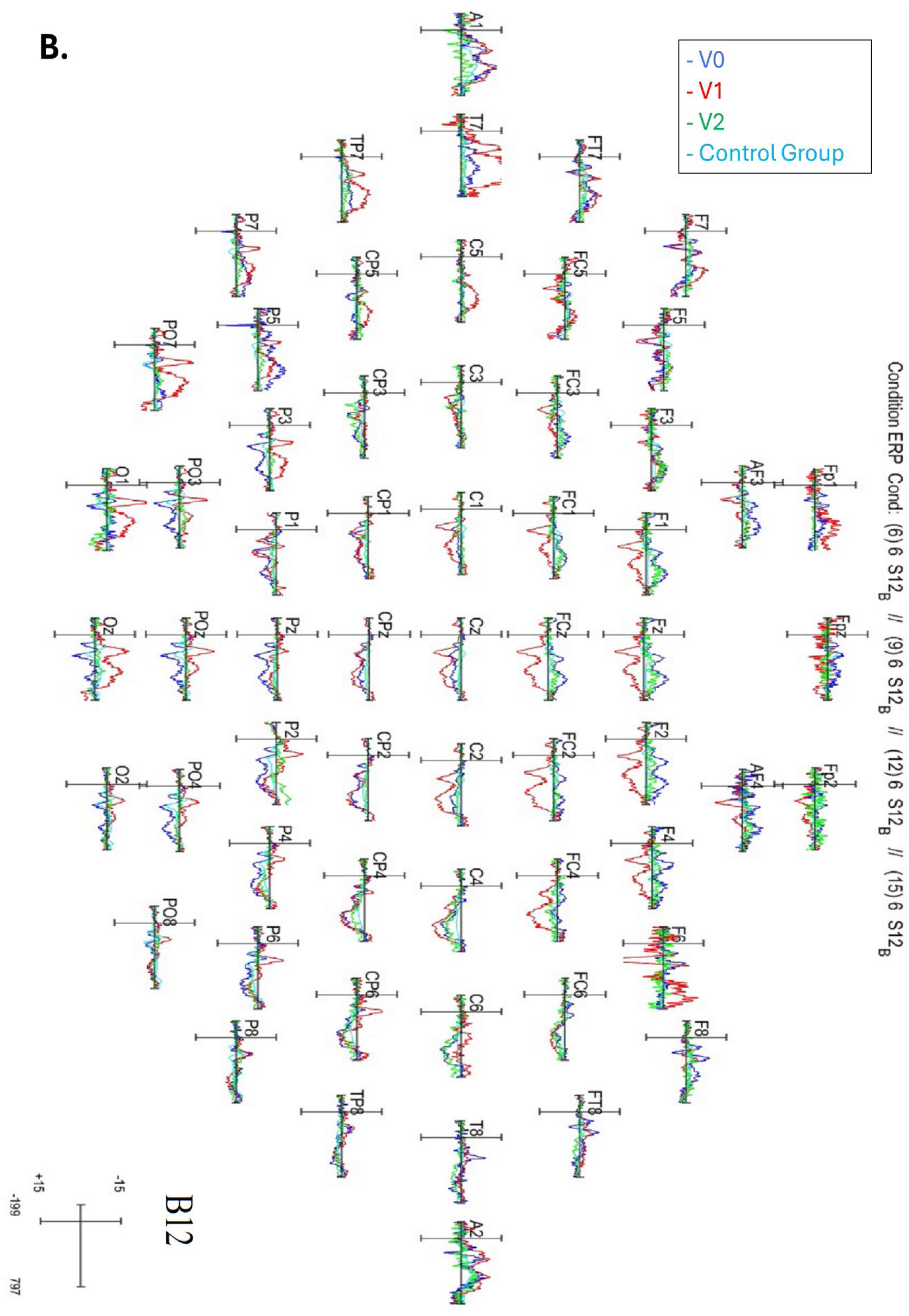
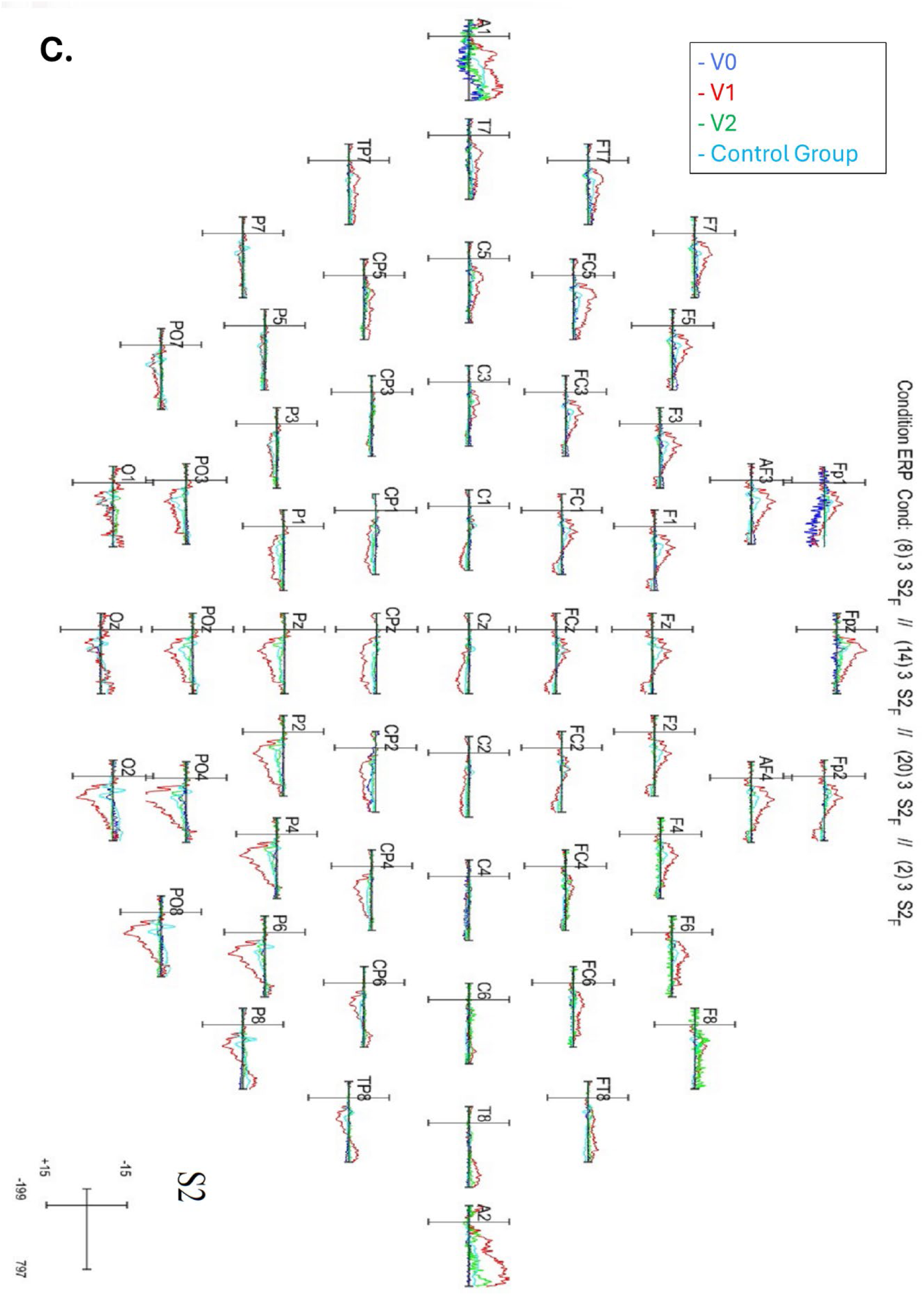
Longitudinal ERP analysis from V0 (pre-training; blue lines), to V1 (5-6 months; red lines), to V2 (9 months; green lines) for three patients: P6 (**A**), P14 (**B**) and P8 (**C**). Blue lines denote the amplitude for Control group. V0 – V1 – V2 followed a nonlinear changing pattern. At V1, a very large increase in visually evoked ERP magnitude was observed across most of the electrodes, particularly in frontal and parieto-occipital regions. From V0 to V1, the amplitude for P6 (V1= 6 months) exhibited a 10-fold increase, P14 (V1=5 months) a 7-fold increase, and P8 (V1= 6months) a 6-fold increase. At 9 months (V2), these increases in ERP magnitude had reversed somewhat, becoming much smaller than observed at V1 but somewhat higher than V0 and even the control group.

These results suggest that increases in ERP magnitude are dependent on training time with the WANR protocol. In other words, when the V1 measurements were made between 5-6 months after training onset, one could observe very high increases in ERP magnitude over most of the brain, while at 3 months such changes were much smaller and barely noticeable. Thus, when compared to the baseline, after 5-6 months, the amplitude of ERP components, particularly in the frontal lobe and in the visual cortical areas of the parietal and occipital lobes, increased exponentially. These findings suggest that the initial stages of clinical recovery could manifest themselves around 5-6 months of training and that the overall activity of visual cortical areas – among other frontal and parietal regions – may provide a good indicator of the magnitude of this clinical recovery. The first assumption was confirmed by the fact that significant motor recovery was already noticed in some of our patients by 5 months of training, as measured by the analysis of parameters like LEMS and WISCI. Moreover, both structural and functional MRI analysis revealed that the initial stages of reversal of the reduction in cortical thickness observed in visual cortical following an SCI began to be noticed within 5 months of training with the WANR protocol.

In order to visualize more easily the longitudinal evolution of visual ERP measurement for all seven patients throughout the 9 months of training, we once again applied PCA to all the ERP parameters measures from V0 to V2. Figure 34 displays the PCA space built using the first three principal components which together accounted for about 80% of the total variance. Inspection of Figure 34 indicates that at V0 (training onset, blue dots) all patients are tightly clustered at the center of the 3D PCA space. At V1 (red dots), the patients are distributed much farther from the center of the PC space. These included the patients with the largest increases in ERP magnitude, like P7, P8, P6, and P14. Two of these patients, P8 and P14, exhibited very signific clinical motor recoveries as early as V1. By the end of training, (V2, green dots), the patient’s ERPs tended to return to the center of the PCA space (e.g. P8, P6, P14) or move away from the red dots (P13, P9). By uniting the three points (V0, V1, V2), one could track the evolution of each of the patient’s ERP data in PCA space, as we did for the structural and functional MRI data.

**Figure 34.**
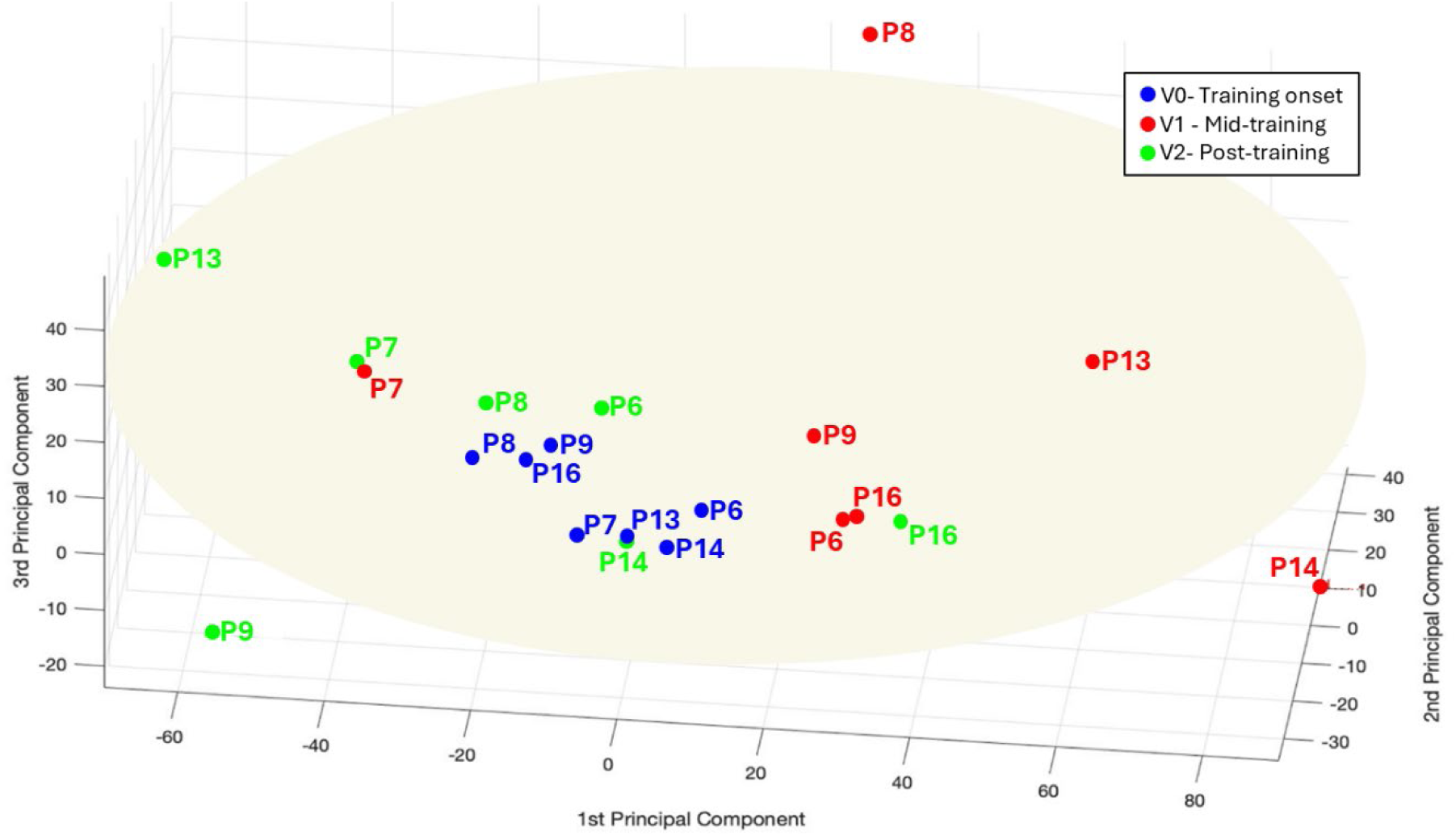
PCA space built using the first 3 principal components derived for the 24 ERP parameters obtained. V0 (training onset, blue dots), V1 (mid training, red dots), V2 (end of training, green dots).

As a way to synthetize our overall findings and initiate a preliminary search for potential indicators of clinical outcome derived from the different brain measurements obtained in this study, we created a correlation matrix that included a series of clinical parameters (i.e. lesion time, age, LEMS, delta ASIA, Final WISCI) and all the patients’ PC scores, obtained from structural and functional MRI imaging, and ERP measurements obtained at the onset (V0), mid-point (V1), and end of training (V2).

Other preliminary correlations between clinical parameters and brain imaging parameters were obtained. For example, Lesion Time was positively correlated with the patients’ score for structural MRI PC3 at V0 (r = 0.92), inversely correlated with MRI PC1 for both V0 (r= -0.87) and V1 (r= -0.97), and positively correlated with MRIPC4 (r = 0.99) and PC5 (r= 0.84) at V2. Lesion time was also inversely correlated with the fourth principal component of structural MRI derived at the end of training (r= 0.85; Fig. 35). Interestingly, this PC included high positive weights for the right and left temporal poles and right and left para-hippocampal areas, and high negative weight for the entorhinal cortex. Further analysis revealed that the patient’s LEMS was inversely correlated with each patient’s score for the structural MRI PC5 at V1 (r= - 0.93), and PC2 at V2 (r = -0.82). In addition, Delta Asia was negatively correlated with patients’ score of structural MRI PC5 at time V0 (r = -0.88) and at time V1 (r = - 0.88). These latter correlations raise the hypothesis that structural MRI changes, measured at the onset or mid-point of the training, could eventually be used for predicting the final clinical outcome of the SCI patients. Further support for this hypothesis was found by the identification of a negative correlation between Final WISCI and the same patients’ score of structural MRI PC5 at V1.

**Figure 35.**
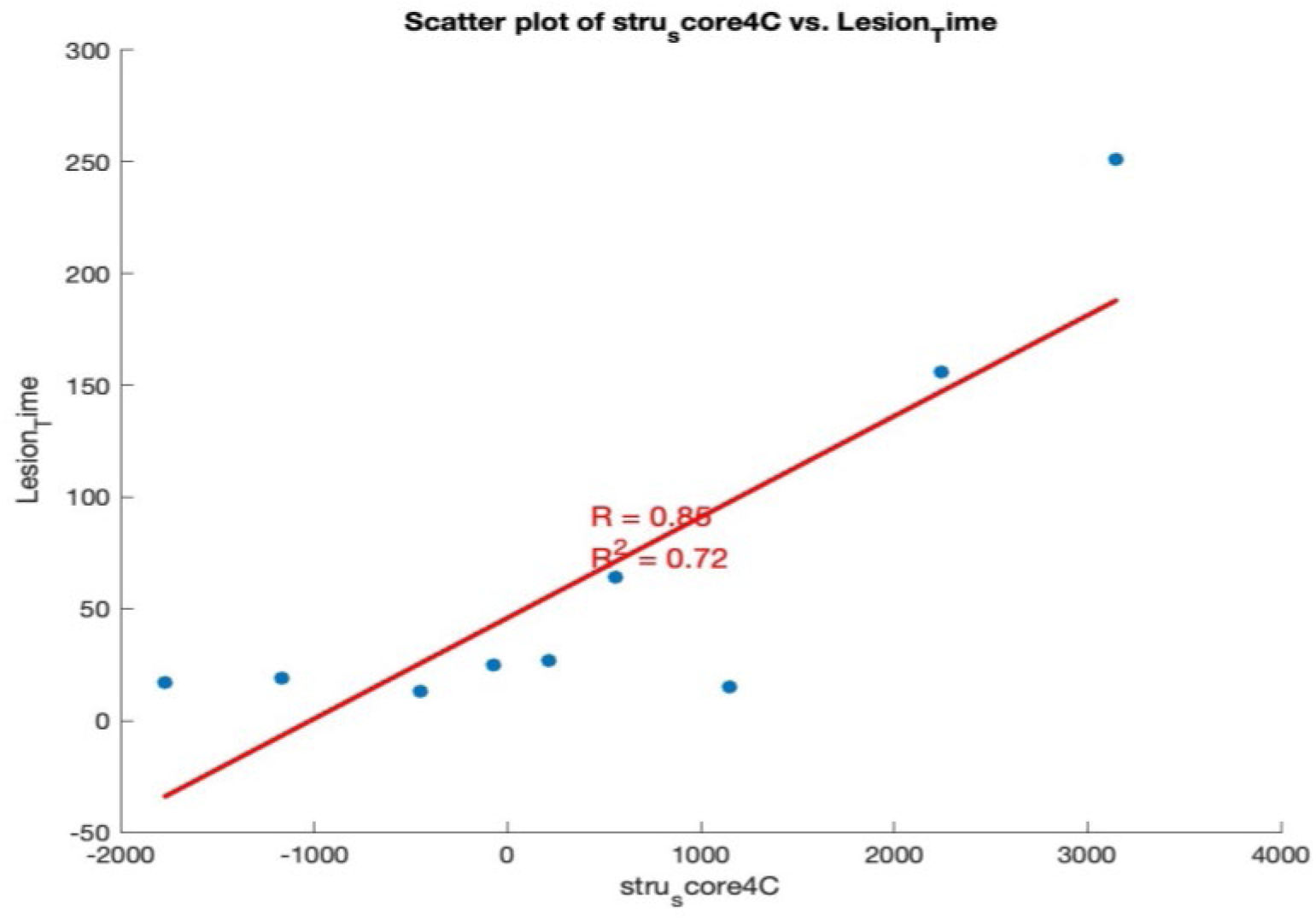
– Correlation between lesion time and PC4 for nine SCI patients.

Similarly interesting correlations were observed when PC scores derived from functional MRI data were considered. For example, patient age, Lesion time, and Final WISCI were also correlated with score of FMRI PC2 at V0 (r = -0.88), Score FMRI PC3 at V2, and Score 3 FMRI at time V0. Finally, scores for different ERP PCs were highly correlated with patient age (e.g., positively correlated with scores for PC2 and PC4 at V0, and negatively correlated with score PC3 at V0). LEMS was negatively correlated with the score for ERP PC1 at V0 and positively correlated with scores for ERP PC1 at V1. Final WISCI was also positively correlated with score ERP PC5 at both V0 and V1, while Delta Asia was negatively correlated with score ERP PC1 at V0 and positively correlated with score ERP PC2 at V1. Interestingly, patients’ scores for FMRI PC were also highly correlated to the scores for ERP PCs. For instance, FMRI PC2 was positively correlated to scores for ERP PC3 at both V0 and V1, showing a great degree of consistency over a period of 5 months. Score FMRI PC1 at V0 was correlated to both ERP PC4 and ERP PC5 at V2. Finally, score FMRI PC2 and PC3 at V0 were highly negatively correlated score for ERP PC 2 at V0 and ERP PC5 at V2.

## DISCUSSION

### Improved Motor Function

In this randomized, controlled, assessor-blind, two-group, single-center trial, we demonstrated that ASIA A paraplegics, suffering from severe spinal cord lesions, ranging from 13 to 251 months, can experience a very significant motor recovery when exposed to a modified version of the WANR protocol, a non-invasive neurorehabilitation training procedure based on the combination of an EEG-based BMI, plus visual feedback, delivered through immersive virtual reality, and robotic locomotion training. Such a partial motor recovery, which was manifested by a significant improvement in voluntary motor control below the level of the SCI lesion, as measured by the LEMS assessments, resulted in a substantial functional improvement in autonomous locomotion, as measured by the WISCI assessment. Such paired neurological and functional recovery was detected as early as 5 months of training and continued to evolve until the end of training (9 months). As a result of this partial motor recovery, at the end of 9 months training, 50% of the 14 patients subjected to the WANR protocol, either partially (n=2) or throughout 9 months of training (n=12), evolved from ASIA A to an ASIA C status, or from complete to partial paraplegia. While the LEMS values and the rate of ASIA upgrading was somewhat similar to the one obtained in a previous study in which the WANR protocol was applied for 12 months [14], the values of functional recovery observed in the present study, as measured by WISCI, were higher in some patients when compared to our previous studies employing the WANR [14, 18, 20]. Thus, while here three patients reached a WISCI of 13 and eight reached a value of 9, no patient reached a WISCI above 9 in our previous study, despite the significant neurological improvements observed. This difference could be the result of a higher number of training sessions per week here (four sessions), when compared to our previous studies (two sessions).

In the present study, one of our patients (P4) became virtually autonomous after 9 months of training, needing only a simple walker and no weight support to walk by herself even on open terrains, like sidewalks. Moreover, another patient (P11, WISCI = 9, ASIA A to ASIA C in 9 months), with a 20-year SCI, was able to walk with crutches, despite two decades confined to bed or a wheelchair. Although she did not reach the same level as P4, patient P11’s functional recovery was also very impressive. Patients P8 and P5 also exhibited important functional recoveries. Curiously, the functional improvement in patient P5 occurred even though she was not able to make a transition to ASIA C.

In contrast to our previous study, no significant changes in light and crude touch, or in pinprick sensations (a second primary outcome) were documented in the present study. One of the potential reasons for this discrepancy is that in the present study the WANR was applied without the implementation of a haptic feedback system, coupled with the immersive virtual reality system, that provided continuous tactile feedback to patients in our previous studies [14]. Another possible cause is that the assessment of tactile and pain prick sensation is much more susceptible to subjective judgments by both patients and the clinical team. Given that the two studies were carried out by different clinical teams of different nationalities and training backgrounds, some differences in evaluation criteria may be expected and could have contributed somewhat to these differences in tactile sensation assessments.

### Longitudinal Structural and Functional Brain Imaging reveals extensive cortical plasticity post-SCI and during and after WANR training

A few structural and functional imaging studies of SCI patient cohorts have appeared in the literature [32–38], although longitudinal studies like ours are much rarer. By using this approach, we were able to characterize the impact of both the SCI and the WANR on both the brain structure and function of the same pool of SCI patients and compare the findings with a pool of healthy subjects. Initial assessment of our pool of SCI patients through brain imaging and visual ERP measurement revealed the occurrence of a widespread reduction in cortical thickness that was present even in patients with 13- to 19-month spinal cord injuries. The degree of cortical atrophy was larger in cortical areas surrounding the primary motor and somatosensory cortices, which are supposed to suffer the brunt of cell death and atrophy following an SCI [35, 37]. Accordingly, we observed major cortical thickness atrophy in multiple temporal cortical areas, such as right and left temporal poles, superior temporal gyri, and transversal temporal gyrus. In addition, a high level of atrophy was documented in the insulas. All these cortical areas suffered reductions in thickness that were superior or equal to the left precentral gyrus where the primary motor cortex is located. Other frontal and parietal cortical areas also exhibited reductions in cortical thickness similar to the left and right precentral gyri. Somewhat smaller reductions were also observed in visual areas of the parietal and occipital lobes. Statistical analysis (Figure 10) indicated that all these structural cortical changes, and other volume reductions in subcortical areas, contribute to making the brain of SCI patients significantly distinct from those of healthy subjects.

Although some structural MRI findings similar to ours have been reported before in the literature [38], the present study provided a higher degree of spatial detail by analyzing up to 68 cortical areas individually. Given the high number of variables analyzed, we introduce a classic multivariate analysis technique, principal component analysis, as well as pairwise correlation analysis, to better characterize the impact of the SCI and the WANR on our patients. Both methods confirmed the widespread spatial spread of cortical atrophy experienced by our cohort of SCI patients. In the case of PCA, this was clearly illustrated by the fact that more than 75% of the variance of our structural MRI data could be accounted by the first principal component, which was formed by a weighted sum of all 68 cortical areas included in the analysis.

The potential functional significance of such widespread structural atrophy was measured through the employment of functional MRI and visual ERPs. Functional MRI revealed the occurrence of an extensive pattern of functional reorganization, characterized by altered connectivity between cortical, subcortical, and cerebellar regions. Thus, SCI patients exhibited increased functional connectivity between the hippocampus and posterior insular cortex, as well as enhanced coupling between visual cortices and frontal or subcortical areas. These findings are consistent with compensatory mechanisms aimed at maintaining body awareness, integrating residual somatosensory input, and modulating emotional and interoceptive processes — functions often disrupted by SCI-induced deafferentation [33, 39]. Notably in this process, the involvement of the posterior insula, associated with enteroception and pain, may also reflect maladaptive plasticity [40] linked to neuropathic phenomena common in chronic SCI [41, 42].

Graph-theoretical metrics derived from functional MRI further supported most of our structural cortical findings and our interpretation of their potential impact. Previously these metrics have been used to examine network reorganization after SCI by other authors [43, 44]. Indeed, our SCI patients showed reduced Cost and Degree in primary sensorimotor and visual cortices (e.g., Precentral/Postcentral Gyri, Lingual Gyrus), suggesting decreased global participation of these areas in network communication. In related spinal pathology models (e.g. cervical spondylotic myelopathy), decreased nodal centralities in postcentral, putamen, and lingual gyrus, and increased centralities in cerebellar regions, suggested as compensatory recruitment, have been documented [45, 46]. Conversely, increased Cost and Degree in cerebellar (lobules 3, 4–5, 9), subcortical (Putamen, Pallidum), and temporal regions may represent compensatory recruitment of alternate motor planning and integrative hubs. Moreover, the elevated Average Path Length in the left Postcentral Gyrus indicates less efficient communication between this somatosensory hub and the rest of the brain, possibly underlying proprioceptive and tactile deficits. Overall, these findings align with prior evidence of plasticity in SCI [47, 48], while extending them to indicate a potential reallocation of network resources away from sensorimotor cortex toward, other cortical areas, and cerebellar and subcortical circuits. This could reflect both compensatory and maladaptive processes, with potential implications for neuromodulatory interventions and rehabilitation targeting spared or reorganized networks, like the cortical areas in the temporal, frontal, parietal and occipital lobes that are either near the primary motor and somatosensory cortices, or highly interconnected to these cortical areas. In that sense, the functional MRI data matched our structural analysis very well, including, for instance, the dense pattern of cortico-cortical thickness correlations between multiple temporal cortical areas in SCI patients when compared to a pool of healthy controls. Thus, while in healthy subjects the precentral and postcentral gyri, as well as the paracentral lobule, are fundamental hubs in terms of exhibiting high cortico-cortical thickness correlations among themselves and with other cortical structures in the frontal, temporal, parietal, and even occipital lobes, after an SCI the pattern of these cortico-cortical thickness correlations contracted somewhat to center primarily in multiple temporal areas, but also in regions located in the frontal and parietal lobes. This shift included a dramatic reduction in correlations between the pre/postcentral gyri and paracentral lobule, as well as a virtual elimination of hubs located in the visual cortex. Basically, this shift in correlations seems to encapsulate both the set of cortical areas that suffered major reductions in cortical thickness at the same time as a result of the SCI, like multiple temporal and frontal cortical areas, as well as those that were involved in the process of functional adaptation/plasticity, as measured by functional MRI that followed the SCI. As such, this analysis helped visualize and quantify the broad spatial spread of structural cortical changes triggered by an SCI. Indeed, to our knowledge, the present study is the first to show such a vastly distributed impact on cortical structural and functional dynamics following an SCI. In this context, it is important to highlight that further support for the somewhat unexpected involvement of visual cortical areas in the process of SCI-driven cortical atrophy was obtained by analysis of visual ERP. This analysis revealed both a statically significant reduction in magnitude and an equally highly significant increase of latency of visually evoked potentials in multiple cortical regions in SCI patients.

### Growing support for the hypothesis that SCI induces cognitive deficits by accelerating the normal process of brain aging

The widespread spatial and functional brain changes observed in the present study offer further structural and functional support for a growing literature that indicates that, in addition to the well-known motor, tactile, and visceral deficits, an estimated 40-64% of SCI patients exhibit extensive cognitive deficits (for reviews see [49–52]. For example, Molina et al. described deficits in attention, processing speed learning and memory, executive functions and recognition, in the subacute and chronic phases of patients with traumatic SCI. Elevated rates of depression have also been described in these patients [50]. Indeed, when compared to healthy subjects, SCI patients exhibit a 13 times higher risk of developing some type of cognitive deficit [53]. Initially some authors attributed this high prevalence of cognitive deficits to the fact that a large percentage of SCIs occur as a result of a concomitant traumatic brain injury (TBI) [54–57]. However, recent studies have indicated that even in the absence of TBI, the risk of cognitive impairment in SCI patients is elevated and higher than in the population without an SCI [58]. In addition, recent country-wide epidemiological studies, performed mainly in Asia, have indicated that SCI patients also have a higher risk for developing dementia [50, 59], Parkinson’s [60], Alzheimer’s disease [61, 62], and even multiple sclerosis [63].

Altogether, these findings have led some authors to propose that an SCI triggers an acceleration of the normal process of brain aging [49, 64]. In this context, it is important to mention that the cortical areas that we identified in the present study as the ones which suffer the most significant losses in thickness following an SCI – like superior temporal, entorhinal and parahippocampal gyrus in the temporal lobe, and several cortical regions of the frontal lobe - are virtually identical to the ones that undergo atrophy both during the normal process of brain aging [65], as well as those resulting from neurodegenerative disorders, such as Parkinson’s [66, 67] and Alzheimer’s disease [68].

According to Mattson and Arumugam [69], the process of natural brain aging can be fully described by a list of 10 hallmarks: (1) mitochondrial dysfunction, (2) intracellular accumulation of oxidatively damaged proteins, nuclei acids, and lipids, (3) dysregulated energy metabolism, (4) impaired cellular waste disposal mechanisms (autophagy-lysosome and proteasome functionality), (5) impaired adaptive stress response signaling, (6) compromised DNA repair, (7) aberrant neuronal network activity, (8) dysregulated neuronal Ca2+ handling, stem cell exhaustion, and (10) inflammation (Figure 36).

**Figure 36.**
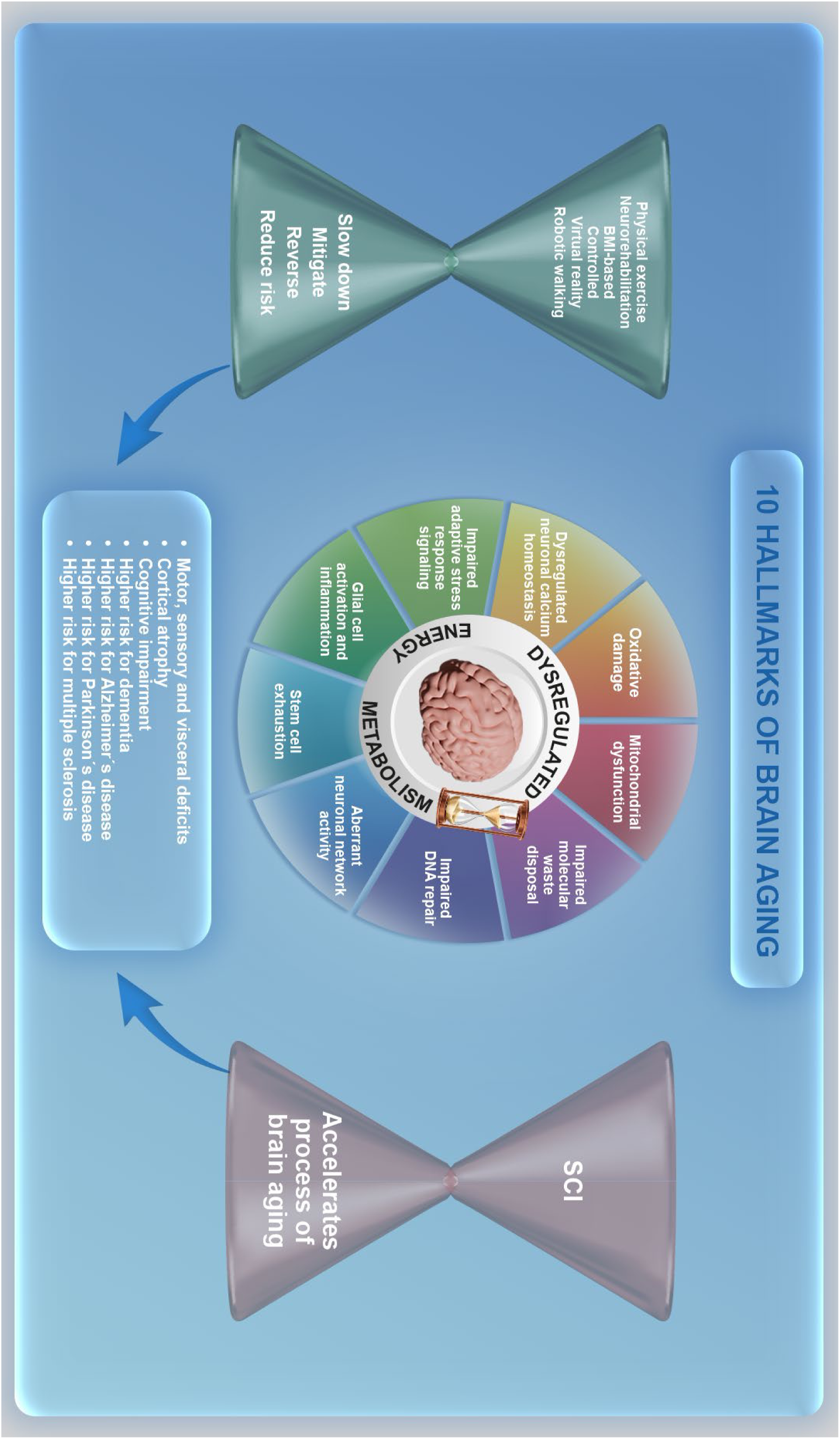
Ten hallmarks of brain aging according to Matson and Arumugam [69]. All these components seem to be triggered by an SCI and most of them seem to be reversed by exercise and BMI training.

According to the same authors, the mechanisms underlying the physiopathology of neurodegenerative disorders like PD and AD may include the exacerbation or acceleration of the molecular and physiological hallmarks of brain aging [69]. Interestingly, a detailed literature search indicates that the same type of reasoning may apply to the chronic effects of an SCI. As shown in Figure 36, most if not all hallmarks of aging listed above seem to be associated, directly or indirectly, with the consequences of an SCI, as shown in both animal and clinical studies (for a review see [39]. Thus, experimental evidence indicates that a severe SCI triggers not only a local, but also a chronic brain inflammatory reaction, due to the production in the spinal cord of chemokines, such as CCL2 and CCL3, that can be transported by axons all the way to brain regions, such as the thalamus, the cortex, the hippocampal CA3 and the dentate gyrus (DG) [50]. Once there, these chemokines activate microglia [50], which release a series of cytokines (e.g. Interleukin 1B and Il-6, and tumor necrosis factor alpha TNF-alpha) that trigger an inflammatory reaction that culminates with a chronic reduction in hippocampal neurogenesis in murine experimental models of SCI [34, 70]; for a review see [71]. Such a reduction in hippocampal neurogenesis has been suggested to play a key role in the genesis of cognitive deficits, depression, and even dementia [72]. For a review see [71]. Eventually, cortical atrophy will result as the final structural consequence of this widespread inflammatory process. During this process, studies in animals have shown that SCI impairs mitochondrial function [73, 74], dysregulating energy metabolism and leading to the production of oxidative damage through the excessive production and accumulation of reactive oxygen species (ROS) [73, 75], the latter due to the impairment of mechanisms of cellular waste disposal. SCI also contributes to impaired DNA repair [76], aberrant neuronal and neural network activity [33], due to an increase in neuronal excitability and reduction in GABA-mediated inhibition [77], and impaired stress response [75]. Therefore, extensive experimental evidence supports the hypothesis that SCI may accelerate, in part or fully, each one of the 10 hallmarks of brain aging proposed by Mattson and Arumugam [69].

Put together, the extensive animal experimental literature on the biochemistry and molecular mechanisms involved in the physiopathology of SCI, particularly the triggering of widespread brain inflammation, plus the demonstration of a high prevalence of cognitive deficits in SCI patients, and the epidemiological evidence for high risk for neurodegenerative disorders, and the characterization of extensive cortical atrophy by brain imaging provide very strong multidimensional support for the hypothesis that SCI indeed triggers an acceleration of the natural mechanisms of brain aging. Clearly, much more research will be needed to categorically demonstrate the merit of this theory. Certainly, if this proves to be the case, the impact of such a demonstration on future neurorehabilitation strategies will be enormous (see discussion below). But even now, these preliminary findings clearly warrant that, going forward, several new variables should be monitored in future clinical studies of SCI rehabilitations protocols. First and foremost, comprehensive cognitive assessments should hereby become routine in the longitudinal evaluation of SCI patients, as well as part of the protocol to measure the clinical impact of any neurorehabilitation strategy or new neurorehabilitation technology.

### The impact of the WANR on cortical atrophy and function

Concurrent longitudinal analysis of brain imaging via structural and functional MRI revealed that training with the modified version of the WANR protocol for 9 months was correlated to: (1) a partial reversal of the widespread reduction in cortical thickness induced by an SCI. This recovery was initially detected at 5 months and peaked at 9 months; (2) The recovery in cortical thickness included the vast majority of the areas that suffered atrophy as a consequence of SCI, but also some other regions, such as different subdivisions of the cingulate gyrus; (3) Such a recovery was maximal in the subgroup of patients who made the transition from ASIA A to C, reaching values of 0.9-1.0 mm in the right temporal pole, left insular, right rostral anterior cingulate cortex, and left entorhinal cortex; (4) a clear change in the spatial distribution of the most important hubs of cortico-cortical thickness correlations. This global cortical pattern of reorganization began its expansion at around 5 months, with the increase in cortical thickness of temporal and cingulate areas and the insula, and a resurgence of significant cortico-cortical thickness correlations between the pre/postcentral gyri and paracentral lobule, and the increase in frontal, cingulate, parietal and occipital hubs. This process was further expanded at 9 months. Analysis of an additional sample of patients who trained with the WANR for 28 months showed a pattern of cortico-cortical thickness correlations that was very similar to healthy subjects; (5) A change in cortico-cortical connectivity, particularly involving connections between the precentral and the superior temporal gyri and between the postcentral gyrus and the visual cortical areas of the occipital lobe; and (6) functional cortical reorganization manifested in multiple circuits. Regarding this latter finding, when the same SCI individuals were re-scanned after undergoing WANR training, allowing us to perform a within-subject assessment of rehabilitation-induced plasticity, trained participants showed increased functional connectivity strength and graph-theoretical indices within cerebellar and vermal regions (e.g., cerebellum 9, Vermis 9), when compared to healthy subjects. This result is consistent with an enhanced integration of motor-coordinative networks. Moreover, an elevated Cost and Degree in these regions support the occurrence of such a plastic reorganization and may reflect the important role played by the cerebellum in refining sensorimotor control, timing, and feedback learning — all critical for performance of our BMI-based protocol. Increased Global Efficiency in the left Cerebellum 9 and right Cerebellum 4–5 also indicated more direct and integrated communication within motor circuits following BMI-based training. Meanwhile, the analysis of Eigenvector Centrality revealed heightened centrality in executive and integrative regions (precuneus, angular gyrus, and the middle frontal gyrus), alongside reduced centrality in sensorimotor hubs (e.g., Heschl’s and the precentral gyri). Overall, these changes may result from a global functional shift from impaired cortical motor loops toward more distributed, possibly compensatory, cortical networks involved in planning, cognition, and proprioceptive feedback. Taken together, these observations suggest that chronic use of a BMI may facilitate targeted neuroplasticity, enhancing the role of subcortical and posterior cortical areas within the broader motor control network. Such a plastic reorganization could support behavioral improvements in movement precision, coordination, and motor imagery in SCI patients undergoing BMI rehabilitation. Indeed, our functional MRI findings in the WANR group are very similar to a study in which training with a motor imaging-based brain-computer interface protocol led to a significant improvement in upper extremity movements in stroke patients [71]. This raises the hypothesis that the cortical and subcortical “pathways” of plastic reorganization, following training with neurorehabilitation protocols based on motor imaging-based BMIs/BCIs, may be independent of the underlying cause of motor impairment.

Consistent with the imaging findings, which showed recovery of thickness and enhanced connectivity of visual cortical areas, we also detected a very significant increase in visual ERPs at around 3 months, which reached a maximum around 5-6 months and was reduced, back to control levels, by 9 months of training. The fact that the WANR protocol relies heavily on the use of immersive virtual reality with strong and salient visual feedback may explain these findings, at least partially. In addition, in animal and clinical studies, moderate levels of aerobic exercise, similar to the type of robotic walking that also is part of the WANR, have also been shown to enhance visual cortical plasticity and neuronal tuning in the visual cortex [78, 79], as well as increase visually evoked potentials [80]. Although results in human studies are somewhat controversial (see [81] for a review), our findings serve as an incentive to further clarify the contribution of exercise and/or neurorehabilitation approaches, involving moderate to intense physical activity, on visual cortical processing and plasticity in future human clinical studies.

The non-linear nature of the process of cortical thickness recovery was further highlighted by the dynamic spatiotemporal pattern observed by the detailed analysis of the cortical structure modifications observed in individual SCI patients included in our study. For instance, during patient P4’s training, the one with the best function recovery of our pool of patients, we observed that multiple pathways were apparently recruited during the 9 months process of cortical thickness recovery. This dynamic process included both enhancements and decreases in cortical thickness happening in distinct cortical structures at different moments of the training. This suggests that the process of cortical plasticity in SCI patients during the WANR application unfolded through very complex spatiotemporal dynamics, involving plastic compensatory mechanisms throughout the cortex and the rest of the brain. Such a process may be guided by, among other factors, the tight energetic budget in which the brain has to operate normally [82].

The inclusion in our analysis of another group of six ASIA A from a previous study, who trained for a much longer period (28 months) with the WANR, showed that time of training is a very important factor in defining the full potential of our protocol to induce a meaningful clinical recovery [14, 18, 20]. In fact, after 28 months of training, all of the patients that remained enrolled in a previous version of the WAP made a transition to ASIA C. MRI brain analysis in six of these patients revealed a very significant recovery in the levels of cortical atrophy. This was clearly documented by plotting the data from the present study and the former in a single PCA space which showed that, as the time of training with the WANR increased, the cortical thickness of SCI patients approached that of the brains of healthy subjects. PCA was also very useful in highlighting the global impact of the WANR training on functional MRI and ERP features of our SCI patients’ brains. In fact, these observations imply that PCA, as well other multivariate statistical techniques, should be incorporated in the analysis of multidimensional data obtained during the implementation of neurorehabilitation protocols to include, not only brain imaging and EEG and ERP parameters, but also clinical variables of interest and even cognitive assessment data. Indeed, the fact that some preliminary correlations were found between some principal components extracted from our different data sets (structural and functional MRI, and ERPs) with important clinical variables, such as age, lesion time, initial and final ASIA score, and WISCI, just to mention a few, warrant looking for potential global structural and functional brain variables that could serve as potential estimates of clinical prognosis in terms of both neurological and functional outcomes. For that hypothesis to be tested properly, however, much larger cohorts of SCI patients would have to be engaged in random, controlled clinical trials in which longitudinal structural and functional MRI is combined with tactile, auditory, and visual ERPs, and broad cognitive testing.

In summary, the use of multiple methods to assess changes in brain structure and function revealed crucial insights into how cortical and subcortical networks reorganize following spinal cord injury and how this reorganization can be modulated through BMI-based interventions. The consistent involvement of cerebellar, vermal, and basal ganglia structures — across both SCI and BMI states — highlights their central role in sensorimotor compensation and plasticity. Moreover, the partial recovery of global efficiency and centrality metrics in executive and integrative hubs suggests that targeting these networks through rehabilitation or neuromodulation may promote functional gains. Therefore, future studies should focus on longitudinal assessments, multimodal integration (e.g., EEG-fMRI, DTI), and interindividual variability in BMI response. Understanding which patterns of network reorganization predict recovery could enable personalized neurorehabilitation strategies, enhancing both motor performance and quality of life in SCI populations. These observations reinforce the importance of targeting reorganized or preserved networks in neuromodulatory interventions. For SCI patients, enhancing cerebellar and temporal, frontal and parietal connectivity may offer new avenues for motor rehabilitation. In BMI users, optimizing multimodal feedback integration and reinforcing cerebellar-subcortical loops may further improve control and function.

Overall, our findings using this longitudinal approach offered valuable insights into the neurobiological underpinnings of clinical recovery and brain adaptation, with potential to guide precision interventions and personalized neurorehabilitation strategies.

### A potential new multidimensional mechanism for induction of clinical recovery by a BMI-based neurorehabilitation protocol

In the present study, we showed that both cortical atrophy and its reversal by training with a non-invasive BMI-based neurorehabilitation protocol is not limited to S1 and M1, but instead it occurs in a highly distributed way all over the neocortex and even in subcortical structures. Previous longitudinal MRI studies have indicated that this process is dependent on motor cortex [35], cerebellum and spinal cord reactivation, triggered by the rehabilitation exercise [36]. For its turn, reactivation of motor cortex in experimental animal models of SCI has been shown to trigger a cascade of structural rearrangement that includes the activation of otherwise dormant axons and the sprouting of axonal collaterals, increase in dendritic trees, increases in synaptogenesis, and increase in glia, which would account for the recovery in cortical thickness (for reviews see [48]. Therefore, it seems proper to speculate that all these processes of structural remodeling likely took place in our SCI patients during the 9 months of training with the WANR.

At this point, it is important to highlight that exercise alone has been shown repeatedly to be beneficial, particularly in aging populations, but also in patients suffering from neurodegenerative diseases, like Parkinson’s [67], in terms of improving learning and memory as well as executive functions, and mitigating or even reversing, the occurrence of age-related brain atrophy – in particular, cortical atrophy – that leads to cognitive decline in this age group [83–89]. Exercise has also shown benefits in SCI patients [90, 91]. In fact, one revision has indicated that at least 80% of brain gray matter can be potentially modified by exercise [92]. Consistent with this appraisal, individual or collective exercise has also been shown to partially reverse cortical atrophy, likely by facilitating synaptic plasticity in the hippocampus, a crucial region for memory and spatial navigation, through the production of growth factors, as well as facilitation of their biochemical cascades, both in the body periphery and the brain [83, 86]. Exercise also seems to have a neuroprotective contribution that likely delays the onset of neurodegenerative disorders, such as PD, AD, or even reduces the cognitive decline caused by these diseases after the appearance of clinical symptoms [84].

Although the original motivation for the introduction of the BMI approach by Chapin and Nicolelis was to create a new experimental paradigm to investigate the neurophysiological properties of large populations of neurons in behaving animals [10, 93–95], soon after these pioneer studies in rodents and monkeys, it became apparent that BMIs held a vast clinical potential to restore the mobility and other neurological functions lost to neurological disorders, such as spinal cord injury [11, 12]. Indeed, BMIs or one of its potential applications, brain-computer interfaces, have been tested in patients suffering from a broad range of neurological disorders, such as stroke [71, 96–101], Parkinson’s disease [102, 103], epilepsy (see [104] for a study in an epilepsy animal model), paraplegia and quadriplegia [14, 105], and locked-in syndrome [106, 107], to mention just a few. Clinical improvement has been documented in some of these applications [71, 96–99, 101]. To this day, however, the basic mechanisms that allow training with BMIs to achieve such clinical recovery, particularly in SCI patients, remain elusive.

The WANR combines a non-invasive, EEG-based BMI control paradigm, with virtual reality and robotic-assisted locomotion. Virtual reality and robotic-assisted technology, have been used in the past, alone or combined, in neurorehabilitation protocols [14, 108–111]. Partial neurological and functional recovery have been reported with these approaches [14, 17, 71]. Moreover, cortical atrophy in both PD and schizophrenic patients can be partially reversed by group exercise [88]. Based on the data generated in this and our previous studies [14, 18, 20], we have now constructed a potential working hypothesis for how the WANR may induce both significant neurologic and functional recovery. Prior studies have suggested that, for any level of clinical recovery to be triggered after an SCI lesion, there is a need for reactivation of the motor cortex to occur [35]. Moreover, several groups have also indicated that reactivation of a recently discovered muscle-brain crosstalk, in which the skeletal musculature, working as an endocrine organ, is capable of influencing brain plasticity through the release of a series of molecules, collectively known as myokines, in the bloodstream that can also play a fundamental role in the process of neurological recovery [112–114]. In this context, different forms of neurorehabilitation strategies are believed to trigger a cascade of molecular, biochemical, structural and physiological neural plastic adaptations which can eventually lead to neurologic and clinical recovery.

Figure 37 describes our hypothesis on how the WANR protocol may induce neurological recovery. This hypothesis is based on the WAP’s 13-year experience with BMI-based neurorehabilitation studies in SCI, combined with novel insights produced by other groups during the same time span. In general, we propose that our non-invasive BMI component of our protocol, which is based on motor imagery and EEG recordings, may operate through two distinct key physiological processes that unfold in different time scales, but are integrated seamlessly during the period of neurorehabilitation training. The first one, which we call “the fast neurophysiological loop”, provides the fundamental neurophysiological pre-requisite of continuous strong motor cortex re-activation in SCI patients to trigger cortical and subcortical neural plasticity. This is achieved by requiring that patients imagine the sort of body movements they intend to perform, while using an EEG-based BMI or BCI to control either a virtual reality avatar of themselves, or some mechanical device, such as a robotic-assisted locomotion device, or an autonomous lower limb exoskeleton, such as employed in our previous studies [14, 18, 20].

**Figure 37.**
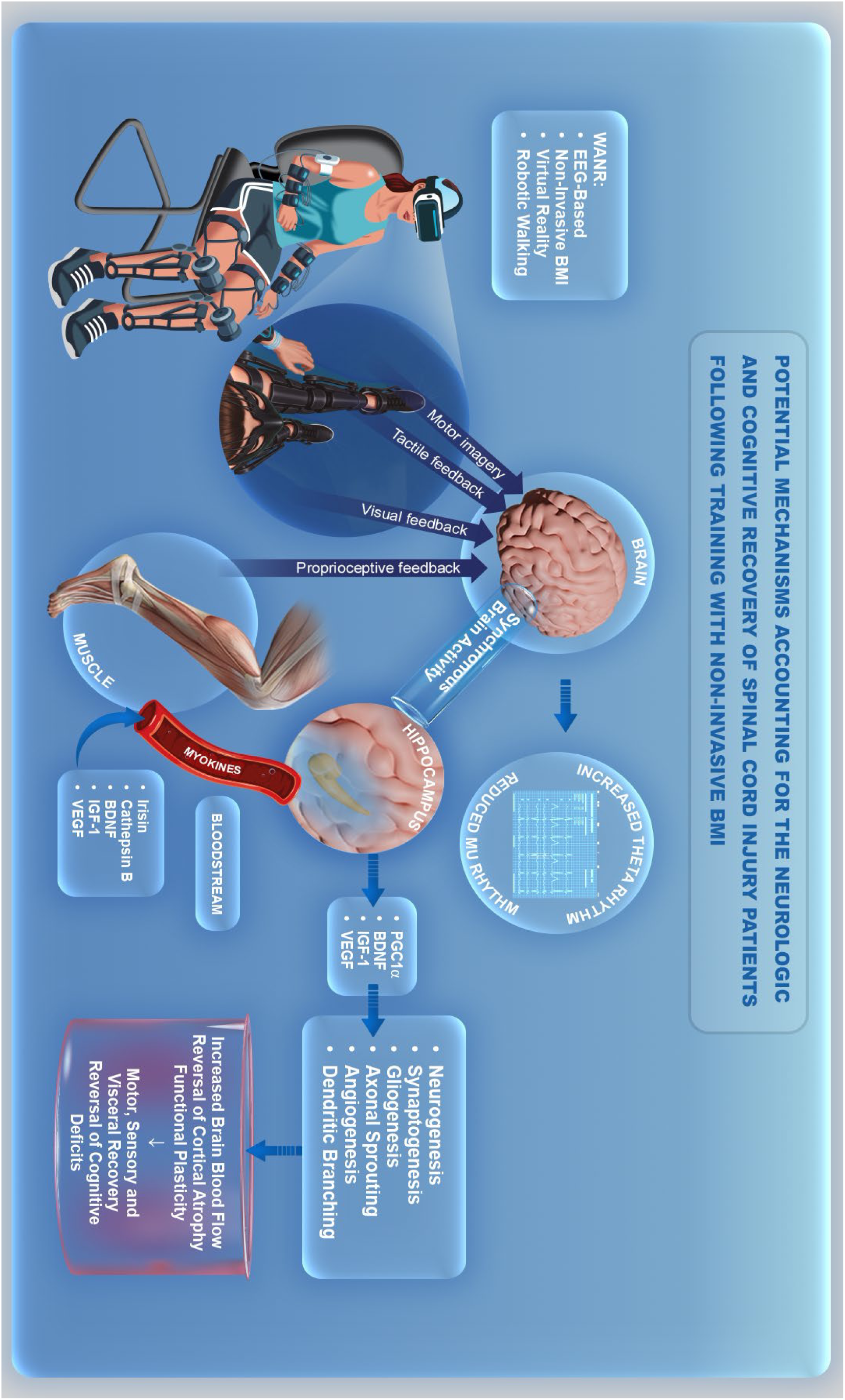
Hypothesis on how the WANR protocol works. This hypothesis is based on the WAP’s 13-year experience with BMI-based neurorehabilitation studies in SCI, combined with novel insights produced by other groups during the same time span.

By providing rich visuals, like in the present study, or visual-tactile feedback - as in previous studies - to these patients’ brains, and generating some small degree of proprioceptive feedback, during BMI-controlled assisted robotic/exoskeleton walking, our protocol allows multimodal reinforcement signals to reach the same motor cortical neurons that are recruited to drive the BMI. These include regular upper cortical motor neurons that survived the retrograde Wallerian degeneration caused by the SCI (for a review see [39], or even subsets of specialized cells, like the cortical mirror neuron network, spread across frontal and parietal cortical areas, which have been shown to be activated by virtual reality feedback in BMI experiments involving monkeys [115]. This spatiotemporal convergence of visual, tactile and proprioceptive signals into the pools of M1, S1 and V1 cortical neurons engaged in BMI control could thereafter trigger a typical Hebbian learning process, not only locally in pools of primary motor and somatosensory cortical areas, but also spread to include populations of neurons located in other cortical areas like, for instance, a variety of frontal, temporal, parietal and even visual cortical areas, that are either connected to M1/S1 or are co-activated by the multimodal sensory feedback generated by training with the WANR. In the particular case of the present study, the use of visual feedback could explain the parallel increase in cortical thickness and ERP magnitude observed in the visual cortical areas of SCI patients who exhibited clinical improvements. In addition, part of this effect in visual areas could be generated by the resumption of body movements and assisted locomotion which have been shown, both in animals and human subjects, to activate neurons in visual cortical areas and even alter their tuning properties [80, 116–119].

Reactivation of the primary motor cortex by this fast component could also enhance the production of highly synchronized theta waves that reach the hippocampus, an area fundamental for sustaining a variety of plastic adaptations, such as synaptogenesis and neurogenesis [120–122]. Many human and animal studies have shown a significant generation of theta waves in the hippocampus during physical activity, such as walking, swimming, or running [123–127]. Such synchronous hippocampal theta activity is believed to play a fundamental role in the maintenance of hippocampal functions, such spatial mapping, learning, and the encoding and retrieving of memories (for a review see [128]. Moreover, synchronous activation of hippocampal neurons by theta waves has been proposed as a key facilitator of the process of adult neurogenesis in the hippocampus [129]. Such a generation of new neurons during adulthood is believed to be important for preventing age-dependent, and neurodegenerative diseases-induced cognitive decline (see [130] for a detailed discussion). It may also be involved in the mechanisms of neurological recovery triggered by neurorehabilitation therapy, such as the WANR. Consistent with this notion, previous spectral analysis of EEG recordings in one of our previous studies has revealed intense power in the theta frequency range in motor cortex while SCI patients engaged in motor imagery during application of the WANR protocol (see Figure 5 of [14].

According to our hypothesis “a second neurorehabilitation loop”, triggered by long-term training with the WANR, is determined by a much slower mechanism than the neurophysiological one. That would be likely engaged when continuous training with the WANR for several weeks or months allows SCI patients to begin to exhibit voluntary limb muscle contractions, and subsequently gain in muscle mass, either through assisted robotic walking, or due to the initial stages of clinical recovery. Once this training stage is reached, lower limb muscles would resume/enhance their endocrine function and begin once again to secrete a series of myokines in the blood stream, like irisin and cathepsin, which can reach the brain after crossing the brain-blood vessel barrier [113, 114]; for a review see [112]. Upon reaching the brain, and in particular the hippocampus, these myokines could facilitate the local production of nerve growth factors, like brain-derived nerve factor (BDNF), insulin-like growth factor (IGF-1) and vascular endothelial growth factor (VEGF), as well PGC1∝, a transcription cofactor that plays a fundamental role in regulating mitochondrial function, and maintaining energy metabolism that is essential for keeping neurons functioning properly [113, 114]; for a review see [112, 131, 132].

Such a cocktail of growth factors would then be responsible for promoting a variety of plastic effects at the hippocampal level, but likely also in other cortical and subcortical structures, including neurogenesis and glycogenesis, axonal sprouting and dendritic branching, and angiogenesis and synaptogenesis [112, 133]. As a whole, these structural adaptations would underlie the functional cortical plastic recovery, e.g. functional compensation by cortical areas adjacent to M1/S1 as well as functional reorganization of the long-range connectivity between cortical and subcortical structures (like the cerebellum and basal ganglia), from which the clinical improvement observed in this study is likely to have emerged. It is important to mention that muscles also secrete BDNF and IGF-1 in the bloodstream [113, 131], but whether these muscle-produced growth factors act on the brain directly remains to be demonstrated categorically [113]. In addition, according to our hypothesis, synchronous hippocampal theta wave activity, generated during motor imagery, could also lead to the production of the same nerve growth factors in the hippocampus.

Our hypothesis for how the WANR exerts its clinical effects is further supported by a series of studies that purport that rehabilitation training – primarily physical exercise - after SCI in experimental animals seems to contribute to reversing most of the so-called hallmarks of regular brain aging. For instance, in addition to leading to the production of the above-mentioned growth factors [134], SCI rehabilitation: (1) reduces the levels of pro-inflammatory cytokines Il-1ß and Il-17 in the central nervous system, (2) increases the levels of anti-inflammatory cytokines IL-10 and TGF-Beta [135–139], (3) promotes synthesis of proteins related to cortical plasticity [140], (4) enhances the density and function of mitochondria, (5) improves the mitochondrial regulation of intracellular Calcium++ [141, 142], and (6) reduces inflammation by reducing activation of M1 and increasing activation of M2 microglia [143–145].

Essentially, all the biochemical, physiological and structural features triggered by rehabilitation training further support the theory that SCIs accelerate the regular process of brain aging, by suggesting that long-term training with non-invasive neurorehabilitation protocols, particularly the ones that potentiate the already known beneficial effects of body exercise, through the combination of virtual reality and BMI operation of robotic-assisted locomotion, may even be used to delay or even mitigate the cognitive decline that accompanies the normal process of brain aging. As far as we can say, this is the first time in which the use of BMI for retarding the process of regular brain aging has been proposed in the literature. Further studies will be needed to demonstrate the validity of this proposal, but the preliminary evidence available seems to indicate that this should be the next logical step for the field of BMI-based neurorehabilitation to follow.

## CONCLUSIONS

Overall, the findings of the present study clearly demonstrate that clinically relevant motor recovery can be achieved, in a matter of a few months, in ASIA A paraplegics, through the use of a non-invasive BMI-based protocol combined with visual feedback and robotic locomotion. Since the WANR is totally defined by a combination of non-invasive methods, its application could start almost immediately after the occurrence of an SCI, even while patients are still hospitalized, or during their acute recovery phase. To stimulate leg muscles, generated in the actual protocol by the use of robotic walking, the simple addition of a non-invasive functional electrical stimulation to our current protocol would suffice to produce the level of sustained muscular activity that is likely to generate the mix of myokines needed to aid the neurorehabilitation process. By starting the application of such a protocol early, it is likely that the process of widespread cortical atrophy that follows the SCI could be delayed or even blocked. By the same token, although the WANR has until now been primarily used in ASIA A patients, the present results suggest that it could be routinely implemented for ASIA C patients, as early as possible after the original SCI. Very likely, these patients could therefore attain even greater levels of both neurological and functional recovery following the use of the WANR. As such, the present findings, as well as our decade long experience with the use of non-invasive neurorehabilitation approaches for SCI patients, clearly imply that a considerable degree of locomotion autonomy can be achieved without the need for more invasive chronic implants, either in the brain and/or the spinal cord, which impose all sorts of unnecessary risks for such patients. This is particularly true, for example, when one takes into account that a variety of non-invasive techniques for spinal cord electrical or electromagnetic stimulations are readily available and could be used before considering surgical implantation. Therefore, at the very least, non-invasive approaches should have precedence in defining the initial neurorehabilitation approach to paraplegic patients soon after the SCI takes place. After a few months of training, the results in terms of neurological, functional and cognitive improvement could be evaluated to decide whether a much more invasive approach would be needed. Such evaluations, whenever possible, should be paralleled by longitudinal brain imaging analysis and other physiological assessments, such as tactile, auditory, and visual ERP measurements. In addition, longitudinal measures of myokines, chemokines, cytokines, and growth factors, like BDNF, VEGF, and IGF-1 in the blood and cerebrospinal fluid should be added to the neurorehabilitation protocol to evaluate the degree of recovery and clinical prognosis.

Based on the more than 12 years of experience applying the WANR to a total of 26 ASIA A patients in two different hospital settings (Brazil and China), we have reached a consensus that, to be considered as a reliable and practical clinical tool, BMI-based neurorehabilitation protocols should fulfill four straightforward conditions, summarized by the acronym SEAS: Safe, Efficacious, Affordable, and Scalable. Overall, these four fundamental principles offer a clear set of criteria to guide the choice between either a non-invasive or an invasive BMI protocol for any type of neurological patient. Indeed, setting objective and evidence-based criteria like these are even more pertinent today, given that academic groups [21, 146–149], as well as private companies (e.g. Neuralink), are pushing very strongly for the exclusive use of only invasive chronic brain and/or spinal cord implants to treat SCI patients. Since its initial inception more than 25 years ago, invasive BMIs, based on chronic brain implants, have proved to be the best neurophysiological approach to investigate the neurophysiological properties of large populations of individual neurons in experimental animals [94, 95]. However, once this approach is translated into human subjects, a large series of other critical factors and considerations should be taken into account before recommending a more invasive procedure. In particular, instead of only considering engineering and computation aspects, such as the higher fidelity and resolution for fine motor control derived from intracranial neuronal recordings, the decision of what approach to use for a given neurological patient must prioritize primarily the maximization of clinical benefit, safety, while minimizing cost and unnecessary risk-taking. Those are easily summarized by our SEAS criteria. Thus, besides the well-known risks associated with invasive neurosurgical procedures (e.g. bleeding, failure to fully penetrate the cortex, post-surgical infection, implant rejection and failure, to name just a few), while small in the case of chronic cortical implants, are far from negligible. For instance, biocompatibility issues related to cortical implants have not yet been fully resolved [25]. This means that cortical implants could stop working in a matter of a few months or even a few weeks in extreme cases. In addition, implantable BMIs are currently expensive and cannot hope to achieve the type of scale-up required to treat the tens of millions of patients who are currently suffering from devastating levels of body paralysis worldwide, as a consequence of an SCI or other neurological motor disorders. Therefore, based on the more than a quarter of century of experience that some of us have acquired with both invasive and non-invasive BMIs, we conclude that non-invasive BMIs currently fulfill the SEAS criteria much better than their invasive counterparts. Put in other words, non-invasive BMI-based protocols are safe, pose no potential side effects; they are efficacious in addressing the objective and life-improving goals of neurorehabilitation for SCI patients; they are highly affordable; and, finally, they can be easily scaled up to cover millions of patients worldwide.

The arguments disclosed in the previous paragraph are further corroborated by the observation that up to now all clinical results involving invasive BMI protocols for treating SCI patients have been limited to either single case reports or, in a small minority of studies, very small patient cohorts or a single case study [17, 149]. In addition to being restricted to demonstration cases and proof-of-concept studies, these preliminary clinical reports also suffer from the fact that some may present misleading conclusions, since they target primarily ASIA C SCI patients who have retained significant levels of lower limb motor control or have been subjected to extensive neurorehabilitation training prior to receiving a cortical and/or spinal cord implant [17, 21, 149]. Moreover, the clinical outcomes reported in ASIA C patients are not substantially better than those obtained with non-invasive approaches and don’t warrant the use of such invasive procedures. In other words, although chronic brain implants are certainly more technologically sophisticated and provide higher spatial resolution neurophysiological data, they have yet to produce a vastly superior clinical outcome in SCI patients that can justify their well-known pitfalls which, unfortunately, seem to be glossed over by their enthusiastic proponents in both academic and private sector settings. This is especially concerning in this particular field, i.e. BMI research, given that today more than ever, potential serious conflicts of interest exist between the academic and the private sectors.

These obvious shortcomings highlight a variety of other challenging issues which are rarely brought forward, such as the difficulties in recruiting paraplegic SCI patients to receive invasive brain implants. At least in our decade-long clinical experience dealing with such patients, it became quite clear that they are generally not very amenable to undergoing invasive procedures. Risk of further clinical complications, the high cost of the invasive protocols, and the fact that cortical implants are not capable of lasting for very long periods of time, all contribute to the patients’ reluctance to undergo these procedures. In fact, in our experience, all paraplegic patients we interviewed clearly indicated their preference for receiving non-invasive therapy instead. At this point, it is important to highlight another important factor that nowadays seems to drive the push for promoting invasive BMIs both in the scientific literature and the market place, i.e. the economic motivation of startup companies in the BMI field, their investors, and their partners in academia, to bet huge amounts of funds on costly technologies that could lead to larger company valuations and, in theory, larger commercial revenues in the future. Yet, neither of these predictions are likely to materialize in the near future primarily because, under the current state-of-the-art, chronic cortical implants do not last long enough and cannot be used to efficiently scale up to a large population of patients, due to their prohibitively high costs for both private and public health systems. Therefore, speculative financial interests alone may explain why, despite the fact that more affordable BMI approaches show significant and useful clinical outcomes, more private funding in the neurotech sector has been allocated to startup companies that aim at developing and utilizing more sophisticated invasive technologies. Undoubtedly, these factors, which are rarely discussed in the scientific literature, bias the debate on whether invasive BMIs offer the best medical compromise for SCI patients in terms of clinical benefits, safety, long-term stability, and affordability. Based on our present and past results, we propose that, at least for paraplegic ASIA A patients, the non-invasive BMI-based approach offers the best compromise in all the key categories considered to be fundamental for providing patients with the best possible clinical outcome.

In addition to the results that deal specifically with SCI, the finding that training with the WANR over a period of several months can induce a significant reversal of the loss in cortical thickness raises two very important hypotheses for future dissemination of this and other non-invasive BMI-based neurorehabilitation protocols. The first one proposes that the WANR, or a slight modification of its original design, could be employed in the process of preventing, or even mitigating and neurorehabilitating, the clinical impacts of a large variety of other brain disorders, notably the neurodegenerative ones, which are characterized by significant levels of cortical atrophy, particularly the frontal and temporal lobes, which lead to major cognitive deficits. This seems like a logical next step, given that reduction in cortical thickness has been associated with the clinical manifestations of a broad range of neurological disorders, like PD and AD, and even some psychiatric disorders. Although further clinical studies would be required to demonstrate the merit of this proposition, the simple prospect that a very safe, affordable, and easy to implement neurorehabilitation protocol may be effective in a variety of other brain disorders in addition to SCI warrants further testing.

Perhaps even more audacious, but no less logical, is the notion that the WANR, or a slightly modified version of it, could also be used, in conjunction with different forms of exercise, to enhance the neuroprotective effects of the latter activity in delaying or even preventing the occurrence of the devastating cognitive decline observed during the normal process of brain aging. This hypothesis is based on the assumption that the combination of natural physical exercise, virtual reality training with rich visuo-tactile feedback, and a motor imaging, BMI-based protocol could in theory delay or even mitigate most of the ten hallmarks of normal brain aging, preventing, among other things, cortical atrophy and, as a fundamental clinical consequence, reducing the occurrence of a multitude of cognitive deficits that emerge as we age.

## Data Availability

All data produced in the present study are available upon reasonable request to the authors.

## AUTHOR CONTRIBUTIONS

Project Design: Guo-Guang Zhao, Yi Tang, Jie Lu, Biao Chen, Peng-Hu Wei, Wei-Qun Song, Hang Wu, Eduardo Alho and Miguel A.L. Nicolelis.

Protocol Design: Eduardo Alho, Adriana Ragoni and Miguel Nicolelis.

Collection and analysis of MRI data: Yi Shan, Jing Li, Eduardo Alho and Miguel Nicolelis.

BMI training and ERP data collection: Yan-Feng Yang, Yuan-Yuan Zhang, Ping Wu.

Clinical Rehabilitation training and Assessments: Lin Liu, Lin Zhu, Chen-Xi Sun, Gui-Xiang Shan.

Manuscript writing and correction: Eduardo Alho, Adriana Ragoni, and Miguel A.L. Nicolelis.

## Acknowledgements

We thank Neiva Paraschiva (AASDAP, Associação Alberto Santos Dumont para Apoio à Pesquisa) and Susan Halkiotis (Nicolelis Institute for Advanced Brain Studies) for their work, help and support for this study. We thank Solaiman Shokur for his scientific contribution at the beginning of this study, and Patrick Braga for the design of figures 7, 36 and 37. Finally, we want to thank the patients for their commitment, and trust in our clinical team and the objectives of this research.

