## Supplementary Tables for "TRAINING WITH A NON-INVASIVE BRAIN-MACHINE INTERFACE, COMBINED WITH VIRTUAL REALITY AND ASSISTED ROBOTIC LOCOMOTION, INDUCES SIGNIFICANT MOTOR RECOVERY AND PARTIAL REVERSAL OF WIDESPREAD CORTICAL ATROPHY IN ASIA A PARAPLEGICS: A RANDOMIZED, CONTROLLED, SINGLE-CENTER TRIAL"

Supplementary Tables – 9

|  |  | All Patients (n=16) |  |
|---|---|---|---|
| Cortical Areas |  | AVG RED (%) ± SE | AVG RED (mm) ± SE |
| Temporal | rtemporalpole | 21.77 ± 2.23 | 0.84 ± 0.09 |
|  | ltemporalpole | 15.93 ± 2.71 | 0.61 ± 0.10 |
| Insula | rinsula | 18.28 ± 2.35 | 0.60 ± 0.08 |
|  | linsula | 17.68 ± 2.83 | 0.57 ± 0.09 |
| Temporal | ltransversetemporal | 19.42 ± 1.51 | 0.49 ± 0.04 |
|  | lsuperiortemporal | 16.60 ± 2.61 | 0.48 ± 0.08 |
|  | rsuperiortemporal | 15.54 ± 1.63 | 0.46 ± 0.05 |
| Pre-Postcentral | lprecentral | 18.76 ± 1.79 | 0.4 5± 0.04 |
| Temporal | rtransversetemporal | 17.42 ± 2.72 | 0.45 ± 0.07 |
|  | lbankssts | 16.60 ± 3.26 | 0.43 ± 0.09 |
|  | lentorhinal | 12.5 ± 2.91 | 0.42 ± 0.10 |
| Frontal | lcaudalmiddlefrontal | 15.58 ± 2.47 | 0.42 ± 0.07 |
|  | rfusiform | 15.15 ± 2.30 | 0.40 ± 0.06 |
|  | rbankssts | 14.72 ± 3.03 | 0.40 ± 0.08 |
| Frontal | lsuperiorfrontal | 14.06 ± 2.15 | 0.40 ± 0.06 |
|  | rcaudalmiddlefrontal | 14.69 ± 2.24 | 0.39 ± 0.06 |
| Temporal | lfusiform | 15.00 ± 2.25 | 0.39 ± 0.06 |
|  | rparahippocampal | 15.73 ± 3.55 | 0.39 ± 0.09 |
| Pre-Postcentral | rprecentral | 16.27 ± 1.71 | 0.39 ± 0.04 |
| Parietal | rsupramarginal | 14.78 ± 2.87 | 0.39 ± 0.07 |
| Temporal | lparahippocampal | 15.28 ± 3.09 | 0.38 ± 0.08 |
| Parietal | lsupramarginal | 14.20 ± 2.22 | 0.37 ± 0.06 |
|  | rinferiorparietal | 14.12 ± 2.46 | 0.36 ± 0.06 |
| Pre-Postcentral | lpostcentral | 16.23 ± 2.40 | 0.36 ± 0.05 |
| Frontal | rsuperiorfrontal | 12.60 ± 2.14 | 0.36 ± 0.06 |
| Paracentral | rparacentral | 15.43 ± 2.50 | 0.35 ± 0.06 |
| Temporal | rmiddletemporal | 12.00 ± 1.44 | 0.35 ± 0.04 |
| Parietal | rprecuneus | 14.16 ± 2.21 | 0.34 ± 0.05 |
| Temporal | lmiddletemporal | 11.72 ± 1.65 | 0.33 ± 0.05 |
| Paracentral | lparacentral | 14.65 ± 2.48 | 0.33 ± 0.06 |
| Frontal | rparsopercularis | 11.87 ± 1.36 | 0.33 ± 0.04 |
| Parietal | lprecuneus | 13.56 ± 2.10 | 0.33 ± 0.05 |
|  | rsuperiorparietal | 13.96 ± 1.59 | 0.32 ± 0.04 |
| Frontal | lparsopercularis | 11.67 ± 1.85 | 0.32 ± 0.05 |
| Temporal | rentorhinal | 9.38 ± 4.43 | 0.32 ± 0.15 |
| Cingulate | lrostralanteriorcingulate | 11.61 ± 2.68 | 0.32 ± 0.07 |
| Frontal | llateralorbitofrontal | 10.98 ± 1.95 | 0.31 ± 0.05 |
| Pre-postcentral | rpostcentral | 14.25 ± 1.61 | 0.31 ± 0.03 |
| Cingulate | rrostralanteriorcingulate | 11.07 ± 2.78 | 0.31 ± 0.08 |
| Frontal | rrostralmiddlefrontal | 11.89 ± 1.86 | 0.31 ± 0.05 |
| Cingulate | lposteriorcingulate | 13.24 ± 2.29 | 0.31 ± 0.05 |
| Parietal | linferiorparietal | 11.89 ± 1.96 | 0.30 ± 0.05 |
| Frontal | lrostralmiddlefrontal | 11.68 ± 1.68 | 0.30 ± 0.04 |
| Occipital | llingual | 13.96 ± 1.71 | 0.29 ± 0.04 |
|  | lpericalcarine | 16.96 ± 1.96 | 0.28 ± 0.03 |
| Frontal | lparsorbitalis | 10.03 ± 1.31 | 0.28 ± 0.04 |
| Cingulate | risthmuscingulate | 13.22 ± 2.80 | 0.28 ± 0.06 |
| Frontal | rparsorbitalis | 9.61 ± 2.60 | 0.27 ± 0.07 |
|  | lparstriangularis | 9.70 ± 1.95 | 0.26 ± 0.05 |
| Parietal | lsuperiorparietal | 10.90 ± 1.65 | 0.25 ± 0.04 |
| Cingulate | rposteriorcingulate | 10.95 ± 2.00 | 0.25 ± 0.05 |
| Occipital | rpericalcarine | 14.35 ± 2.18 | 0.24 ± 0.04 |
| Frontal | rlateralorbitofrontal | 8.54 ± 1.73 | 0.23 ± 0.05 |
| Occipital | rlingual | 11.35 ± 2.01 | 0.23 ± 0.04 |
| Cingulate | listhmuscingulate | 10.75 ± 2.41 | 0.23 ± 0.05 |
| Frontal | rparstriangularis | 8.26 ± 1.05 | 0.22 ± 0.03 |
| Temporal | linferiortemporal | 7.86 ± 1.95 | 0.22 ± 0.05 |
| Cingulate | rcaudalanteriorcingulate | 8.59 ± 1.88 | 0.21 ± 0.05 |
| Occipital | lcuneus | 11.03 ± 1.77 | 0.21 ± 0.03 |
|  | rlateraloccipital | 9.33 ± 1.57 | 0.21 ± 0.03 |
|  | llateraloccipital | 9.09 ± 1.82 | 0.20 ± 0.04 |
| Frontal | rfrontalpole | 7.19 ± 1.89 | 0.19 ± 0.05 |
|  | lfrontalpole | 6.79 ± 1.45 | 0.19 ± 0.04 |
| Temporal | rinferiortemporal | 6.66 ± 2.19 | 0.19 ± 0.06 |
| Occipital | rcuneus | 9.01 ± 2.43 | 0.17 ± 0.05 |
| Frontal | rmedialorbitofrontal | 6.39 ± 1.84 | 0.17 ± 0.05 |
| Cingulate | lcaudalanteriorcingulate | 6.40 ± 2.31 | 0.15 ± 0.06 |
| Frontal | lmedialorbitofrontal | 3.57 ± 1.54 | 0.09 ± 0.04 |

**Supplementary Table 1.** This table supplements Tables 4 and 5. It ranks the level of reduction of all the 68 cortical areas surveyed in structural MRI for a subset of 16 of the 18 patients enrolled in our clinical trial (WANR group=9; NR group=7), prior to the onset of the training. Values are the AVG±SE; measurements both in terms of percentage of reduction, when compared to values obtained from a pool of healthy subjects, and in mm. Areas are ranked by AVG RED (mm). The insulas are shown in green; cortical areas in the temporal lobe areas are shown in brown; the combined pre and postcentral gyri areas are shown in light red; paracentral areas are shown in dark red; parietal areas are shown in yellow; frontal areas in salmon; cingulate lobe areas are shown in orange and occipital lobe areas are shown in blue. For the designation of each cortical area, the letter "l" depicts the left hemisphere, and "r" the right hemisphere.

| | Cortical Areas | WANR Group (n=11) AVG IMP (mm) ± SE |
|---|---|---|
| Temporal | rtemporalpole | 0.53 ± 0.23 |
| Insula | linsula | 0.43 ±0.22 |
| Temporal | rentorhinal | 0.39 ± 0.16 |
| Frontal | lparsorbitalis | 0.38 ± 0.20 |
| Temporal | rparahippocampal | 0.34 ± 0.17 |
| Frontal | rlateralorbitofrontal | 0.33 ± 0.20 |
| Temporal | rsuperiortemporal | 0.33 ± 0.21 |
| Cingulate | lrostralanteriorcingulate | 0.33 ± 0.19 |
| Temporal | rfusiform | 0.31 ± 0.21 |
| | rmiddletemporal | 0.30 ± 0.22 |
| | lentorhinal | 0.30 ± 0.25 |
| Frontal | lparsopercularis | 0.30 ± 0.20 |
| Temporal | lfusiform | 0.29 ± 0.19 |
| Parietal | rinferiorparietal | 0.28 ± 0.18 |
| Frontal | rmedialorbitofrontal | 0.28 ± 0.22 |
| Parietal | linferiorparietal | 0.28 ± 0.18 |
| Temporal | rtransversetemporal | 0.28 ± 0.20 |
| Frontal | llateralorbitofrontal | 0.27 ± 0.21 |
| Cingulate | rrostralanteriorcingulate | 0.27 ± 0.25 |
| | listhmuscingulate | 0.26 ± 0.15 |
| Frontal | rparsopercularis | 0.24 ± 0.20 |
| Temporal | lmiddletemporal | 0.24 ± 0.22 |
| Pre-Postcentral | lprecentral | 0.23 ± 0.16 |
| Frontal | rsuperiorfrontal | 0.23 ± 0.22 |
| Occipital | rlingual | 0.23 ± 0.15 |
| Frontal | rparstriangularis | 0.20 ± 0.20 |
| Pre-Postcentral | rprecentral | 0.19 ± 0.17 |
| Frontal | rrostralmiddlefrontal | 0.19 ± 0.19 |
| Temporal | ltemporalpole | 0.19 ± 0.09 |
| Cingulate | rcaudalanteriorcingulate | 0.16 ± 0.25 |
| Temporal | rbankssts | 0.15 ± 0.11 |
| Parietal | rsupramarginal | 0.13 ± 0.10 |
| Temporal | lsuperiortemporal | 0.11 ± 0.05 |
| | rinferiortemporal | 0.11 ± 0.05 |
| Paracentral | rparacentral | 0.11 ± 0.06 |
| Frontal | lmedialorbitofrontal | 0.11 ± 0.05 |
| Paracentral | lparacentral | 0.10 ± 0.07 |
| Occipital | rcuneus | 0.10 ± 0.15 |
| Cingulate | lposteriorcingulate | 0.10 ± 0.07 |
| Temporal | lbankssts | 0.10 ± 0.10 |
| Insula | rinsula | 0.10 ± 0.11 |
| Frontal | rparsorbitalis | 0.10 ± 0.11 |
| | lsuperiorfrontal | 0.09 ± 0.07 |
| Cingulate | risthmuscingulate | 0.09 ± 0.05 |
| Temporal | lparahippocampal | 0.09 ± 0.05 |
| Parietal | lsupramarginal | 0.08 ± 0.05 |
| Frontal | lfrontalpole | 0.08 ± 0.26 |
| | lparstriangularis | 0.08 ± 0.05 |
| Temporal | linferiortemporal | 0.08 ± 0.05 |
| Parietal | lprecuneus | 0.08 ± 0.04 |
| Occipital | llingual | 0.08 ± 0.03 |
| Pre-Postcentral | rpostcentral | 0.08 ± 0.06 |
| Frontal | lcaudalmiddlefrontal | 0.08 ± 0.09 |
| Pre-Postcentral | lpostcentral | 0.07 ± 0.07 |
| Occipital | rlateraloccipital | 0.06 ± 0.04 |
| Frontal | rcaudalmiddlefrontal | 0.06 ± 0.08 |
| Occipital | llateraloccipital | 0.05 ± 0.06 |
| Parietal | rprecuneus | 0.05 ± 0.04 |
| Occipital | lcuneus | 0.04 ± 0.03 |
| Parietal | lsuperiorparietal | 0.04 ± 0.02 |
| Occipital | lpericalcarine | 0.03 ± 0.04 |
| Temporal | ltransversetemporal | 0.03 ± 0.08 |
| Cingulate | lcaudalanteriorcingulate | 0.02 ± 0.08 |
| Parietal | rsuperiorparietal | 0.01 ± 0.05 |
| Occipital | rpericalcarine | -0.003 ± 0.03 |
| Frontal | lrostralmiddlefrontal | -0.003 ± 0.05 |
| Cingulate | rposteriorcingulate | -0.03 ± 0.05 |
| Frontal | rfrontalpole | -0.06 ± 0.05 |

**Supplementary Table 2.** This table supplements Tables 6 and 7. It ranks the level of improvement of all the 68 cortical areas surveyed in structural MRI for the WANR group (n=11) after 5 months of training. Values are AVG±SE; measurements mm. The insulas are shown in green; cortical areas in the temporal lobe areas are shown in brown; the combined pre and postcentral gyri areas are shown in light red; paracentral areas are shown in dark red; parietal areas are shown in yellow; frontal areas in salmon; cingulate lobe areas are shown in orange and occipital lobe areas are shown in blue. For the designation of each cortical area, the letter "l" depicts the left hemisphere, and "r" the right hemisphere.

| | Cortical Areas | WANR Group (n=10) AVG IMP (mm) ± SE | NR Group (n=5) AVG IMP (mm) ± SE | Abs Hedges' g |
|---|---|---|---|---|
| Insula | linsula | 0.44± 0.25 | -0.11 ± 0.14 | 0.76 |
| Cingulate | lrostralanteriorcingulate | 0.34 ± 0.23 | 0.02 ± 0.05 | 0.49 |
| Parietal | rinferiorparietal | 0.30 ± 0.21 | -0.06 ± 0.09 | 0.61 |
| Temporal | rtemporalpole | 0.30 ± 0.32 | 0.16 ± 0.25 | 0.15 |
| | lfusiform | 0.28 ± 0.21 | 0.07 ± 0.11 | 0.36 |
| Frontal | lparsopercularis | 0.28 ± 0.22 | 0.05 ± 0.08 | 0.38 |
| Temporal | rsuperiortemporal | 0.28 ± 0.24 | -0.02 ± 0.11 | 0.43 |
| | rparahippocampal | 0.28 ± 0.19 | -0.03 ± 0.16 | 0.54 |
| | rmiddletemporal | 0.28 ± 0.25 | 0.01 ± 0.07 | 0.37 |
| Frontal | rsuperiorfrontal | 0.27 ± 0.23 | -0.06 ± 0.11 | 0.51 |
| Temporal | rtransversetemporal | 0.27 ± 0.20 | 0.04 ± 0.17 | 0.38 |
| | rfusiform | 0.27 ± 0.23 | -0.003 ± 0.10 | 0.42 |
| Cingulate | rrostralanteriorcingulate | 0.27 ± 0.26 | -0.11 ± 0.08 | 0.51 |
| Frontal | rparsopercularis | 0.26 ± 0.23 | 0.06 ± 0.08 | 0.30 |
| Temporal | lentorhinal | 0.26 ± 0.30 | 0.05 ± 0.35 | 0.22 |
| Frontal | rlateralorbitofrontal | 0.25 ± 0.23 | -0.03 ± 0.13 | 0.41 |
| | lparsorbitalis | 0.25 ± 0.24 | -0.06 ± 0.14 | 0.44 |
| | rrostralmiddlefrontal | 0.25 ± 0.21 | -0.02 ± 0.12 | 0.45 |
| Parietal | linferiorparietal | 0.25 ± 0.21 | 0.03 ± 0.06 | 0.36 |
| Cingulate | rcaudalanteriorcingulate | 0.24 ± 0.21 | -0.05 ± 0.09 | 0.47 |
| Frontal | llateralorbitofrontal | 0.23 ± 0.24 | -0.01 ± 0.15 | 0.35 |
| Pre-Postcentral | rprecentral | 0.23 ± 0.19 | -0.15 ± 0.06 | 0.72 |
| Temporal | lmiddletemporal | 0.23 ± 0.24 | 0.11 ± 0.09 | 0.18 |
| Frontal | rparstriangularis | 0.22 ± 0.23 | 0.01 ± 0.10 | 0.32 |
| Pre-Postcentral | lprecentral | 0.21 ± 0.19 | -0.14 ± 0.10 | 0.66 |
| Cingulate | listhmuscingulate | 0.20 ± 0.17 | 0.02 ± 0.10 | 0.38 |
| Occipital | rlingual | 0.17 ± 0.17 | -0.01 ± 0.07 | 0.37 |
| Frontal | rmedialorbitofrontal | 0.17 ± 0.24 | 0.04 ± 0.08 | 0.20 |
| | rparsorbitalis | 0.15 ± 0.07 | -0.04 ± 0.18 | 0.63 |
| Insula | rinsula | 0.13 ± 0.11 | -0.03 ± 0.16 | 0.41 |
| Parietal | rsupramarginal | 0.12 ± 0.11 | -0.03 ± 0.10 | 0.43 |
| Temporal | ltemporalpole | 0.12 ± 0.16 | -0.16 ± 0.19 | 0.55 |
| Frontal | lfrontalpole | 0.11 ± 0.27 | 0.04 ± 0.08 | 0.10 |
| Temporal | rbankssts | 0.11 ± 0.12 | 0.01 ± 0.07 | 0.29 |
| | lsuperiortemporal | 0.11 ± 0.10 | 0.02 ± 0.11 | 0.26 |
| Cingulate | lposteriorcingulate | 0.10 ± 0.08 | -0.002 ± 0.06 | 0.46 |
| Frontal | lcaudalmiddlefrontal | 0.10 ± 0.10 | -0.08 ± 0.09 | 0.63 |
| Occipital | rcuneus | 0.09 ± 0.16 | 0.02 ± 0.07 | 0.16 |
| Pre-Postcentral | lpostcentral | 0.09 ± 0.07 | -0.11 ± 0.09 | 0.84 |
| Frontal | rcaudalmiddlefrontal | 0.09 ± 0.09 | -0.09 ± 0.08 | 0.68 |
| Paracentral | lparacentral | 0.08 ± 0.06 | -0.08 ± 0.06 | 0.84 |
| Temporal | rentorhinal | 0.08 ± 0.16 | 0.18 ± 0.37 | 0.15 |
| Frontal | lsuperiorfrontal | 0.08 ± 0.08 | -0.04 ± 0.08 | 0.48 |
| Paracentral | rparacentral | 0.07 ± 0.06 | -0.10 ± 0.13 | 0.72 |
| Temporal | lparahippocampal | 0.07 ± 0.07 | 0.11 ± 0.21 | 0.13 |
| Parietal | lsupramarginal | 0.06 ± 0.08 | -0.02 ± 0.10 | 0.33 |
| Temporal | lbankssts | 0.06 ± 0.13 | 0.13 ± 0.14 | 0.17 |
| Occipital | lpericalcarine | 0.06 ± 0.04 | 0.03 ± 0.04 | 0.23 |
| Parietal | rprecuneus | 0.06 ± 0.05 | -0.05 ± 0.11 | 0.56 |
| Parietal | lprecuneus | 0.04 ± 0.06 | 0.02 ± 0.06 | 0.12 |
| Pre-Postcentral | rpostcentral | 0.04 ± 0.05 | -0.12 ± 0.08 | 0.96 |
| Cingulate | lcaudalanteriorcingulate | 0.04 ± 0.07 | -0.01 ± 0.05 | 0.26 |
| Temporal | ltransversetemporal | 0.03 ± 0.03 | 0.06 ± 0.10 | 0.17 |
| Frontal | lrostralmiddlefrontal | 0.03 ± 0.07 | -0.05 ± 0.10 | 0.34 |
| Temporal | linferiortemporal | 0.03 ± 0.07 | 0.15 ± 0.09 | 0.52 |
| Cingulate | rposteriorcingulate | 0.03 ± 0.06 | 0.03 ± 0.10 | 0.01 |
| Parietal | rsuperiorparietal | 0.02 ± 0.05 | -0.01 ± 0.10 | 0.17 |
| Occipital | llingual | 0.02 ± 0.04 | 0.03 ± 0.07 | 0.12 |
| | llateraloccipital | 0.02 ± 0.06 | 0.002 ± 0.05 | 0.06 |
| Frontal | lparstriangularis | 0.01 ± 0.07 | 0.03 ± 0.08 | 0.08 |
| Temporal | rinferiortemporal | 0.01 ± 0.08 | 0.11 ± 0.09 | 0.38 |
| Frontal | lmedialorbitofrontal | 0.01 ± 0.05 | 0.04 ± 0.09 | 0.20 |
| Occipital | rlateraloccipital | 0.0002 ± 0.04 | 0.06 ± 0.08 | 0.34 |
| Parietal | lsuperiorparietal | -0.003 ± 0.05 | -0.03 ± 0.08 | 0.13 |
| Occipital | lcuneus | -0.01 ± 0.04 | 0.02 ± 0.03 | 0.21 |
| Cingulate | risthmuscingulate | -0.01 ± 0.05 | 0.08 ± 0.06 | 0.55 |
| Occipital | rpericalcarine | -0.07 ± 0.05 | -0.07 ± 0.05 | 0.04 |
| Frontal | rfrontalpole | -0.11 ± 0.12 | 0.03 ± 0.08 | 0.42 |

**Supplementary Table 3.** This table supplements Tables 8 and 9. It ranks the level of improvement of all the 68 cortical areas surveyed in structural MRI (values are AVG±SE; measurements mm) for the WANR group (n=11) after 9 months of training, the values of the same areas for the NR group (n=5), and the Absolute Hedges' g values for WANR x NR groups. Hedges' g represents how many pooled standard deviations apart two group means are, with a small-sample bias correction. Its magnitude conveys the effect size (~0.2 small, ~0.5 medium, ~0.8 large, ≥1.2 very large). The insulas are shown in green; cortical areas in the temporal lobe areas are shown in brown; the combined pre and postcentral gyri areas are shown in light red; paracentral areas are shown in dark red; parietal areas are shown in yellow; frontal areas in salmon; cingulate lobe areas are shown in orange and occipital lobe areas are shown in blue. For the designation of each cortical area, the letter "l" depicts the left hemisphere, and "r" the right hemisphere.

| Cortical Areas | | ASIA A-C (n=5)<br>AVG IMP (mm) ± SE |
|---|---|---|
| Temporal | rtemporalpole | 0.87 ± 0.45 |
| Insula | linsula | 0.72 ± 0.46 |
| Temporal | rentorhinal | 0.64 ± 0.20 |
| Cingulate | rrostralanteriorcingulate | 0.64 ± 0.53 |
| Frontal | lparsorbitalis | 0.64 ± 0.43 |
| Cingulate | lrostralanteriorcingulate | 0.62 ± 0.40 |
| Temporal | rfusiform | 0.62 ± 0.44 |
| | rparahippocampal | 0.62 ± 0.33 |
| | rsuperiortemporal | 0.58 ± 0.45 |
| | rmiddletemporal | 0.56 ± 0.48 |
| Parietal | rinferiorparietal | 0.56 ± 0.36 |
| Frontal | rlateralorbitofrontal | 0.55 ± 0.45 |
| | rmedialorbitofrontal | 0.55 ± 0.46 |
| | lparsopercularis | 0.55 ± 0.44 |
| Cingulate | rcaudalanteriorcingulate | 0.55 ± 0.48 |
| Parietal | linferiorparietal | 0.53 ± 0.39 |
| Temporal | rtransversetemporal | 0.52 ± 0.41 |
| | lfusiform | 0.52 ± 0.42 |
| Frontal | rparsopercularis | 0.51 ± 0.44 |
| | rsuperiorfrontal | 0.50 ± 0.48 |
| Temporal | lmiddletemporal | 0.48 ± 0.48 |
| Frontal | llateralorbitofrontal | 0.47 ± 0.47 |
| Temporal | lentorhinal | 0.46 ± 0.56 |
| Frontal | rparstriangularis | 0.46 ± 0.43 |
| | rrostralmiddlefrontal | 0.45 ± 0.41 |
| Pre-Postcentral | lprecentral | 0.43 ± 0.35 |
| | rprecentral | 0.41 ± 0.35 |
| Occipital | rlingual | 0.38 ± 0.34 |
| Cingulate | listhmuscingulate | 0.37 ± 0.30 |
| Occipital | rcuneus | 0.33 ± 0.30 |
| Temporal | ltemporalpole | 0.29 ± 0.16 |
| Parietal | rsupramarginal | 0.25 ± 0.22 |
| Temporal | lbankssts | 0.21 ± 0.22 |
| | rbankssts | 0.19 ± 0.22 |
| Cingulate | lposteriorcingulate | 0.18 ± 0.14 |
| Paracentral | lparacentral | 0.17 ± 0.13 |
| Temporal | lsuperiortemporal | 0.15 ± 0.10 |
| Cingulate | lcaudalanteriorcingulate | 0.14 ± 0.10 |
| Frontal | rcaudalmiddlefrontal | 0.13 ± 0.16 |
| | lsuperiorfrontal | 0.13 ± 0.13 |
| Paracentral | rparacentral | 0.13 ± 0.09 |
| Parietal | rprecuneus | 0.12 ± 0.07 |
| Occipital | llateraloccipital | 0.12 ± 0.12 |
| Temporal | lparahippocampal | 0.12 ± 0.08 |
| Occipital | llingual | 0.12 ± 0.03 |
| Frontal | lparstriangularis | 0.12 ± 0.10 |
| Pre-Postcentral | lpostcentral | 0.11 ± 0.13 |
| Temporal | rinferiortemporal | 0.11 ± 0.09 |
| Frontal | lmedialorbitofrontal | 0.11 ± 0.06 |
| Parietal | lprecuneus | 0.11 ± 0.08 |
| | lsupramarginal | 0.11 ± 0.11 |
| Frontal | lfrontalpole | 0.11 ± 0.58 |
| Pre-Postcentral | rpostcentral | 0.08 ± 0.09 |
| Parietal | rsuperiorparietal | 0.08 ± 0.04 |
| | lsuperiorparietal | 0.08 ± 0.02 |
| Frontal | lcaudalmiddlefrontal | 0.08 ± 0.20 |
| Insula | rinsula | 0.07 ± 0.20 |
| Occiptal | lpericalcarine | 0.06 ± 0.05 |
| | rlateraloccipital | 0.06 ± 0.03 |
| | lcuneus | 0.06 ± 0.05 |
| Cingulate | rposteriorcingulate | 0.06 ± 0.06 |
| Frontal | rparsorbitalis | 0.06 ± 0.17 |
| Cingulate | risthmuscingulate | 0.03 ± 0.06 |
| Temporal | linferiortemporal | 0.02 ± 0.07 |
| Frontal | lrostralmiddlefrontal | 0.002 ± 0.08 |
| | rfrontalpole | -0.002 ± 0.06 |
| Occipital | rpericalcarine | -0.01 ± 0.04 |
| Temporal | ltransversetemporal | -0.13 ± 0.15 |

**Supplementary Table 4.** This table supplements Tables 11 and 12. It ranks the level of improvement of all the 68 cortical areas surveyed in structural MRI (values are AVG±SE; measurements mm) for the subgroup of the WANR patients that made a transition from ASIA A to ASIA C (n=5), after 5 months of training. A total of 36 cortical areas (54% of the 68 cortical areas analyzed) have values ≥ 0.2mm (limited by the black arrow): 13 areas in the temporal lobe, nine in the frontal, five in the cingulate cortex, three in the parietal, two in the pre- and postcentral gyri, two in the occipital lobe, one in the insula and one in the paracentral lobule. The insulas are shown in green; cortical areas in the temporal lobe areas are shown in brown; the combined pre and postcentral gyri areas are shown in light red; paracentral areas are shown in dark red; parietal areas are shown in yellow; frontal areas in salmon; cingulate lobe areas are shown in orange and occipital lobe areas are shown in blue. For the designation of each cortical area, the letter "l" depicts the left hemisphere, and "r" the right hemisphere.

| Cortical Areas | | ASIA A-A (n=6) AVG IMP (mm) ± SE |
|---|---|---|
| Temporal | rtemporalpole | 0.25 ± 0.18 |
| Insula | linsula | 0.19 ± 0.09 |
| Temporal | rentorhinal | 0.18 ± 0.22 |
| | lentorhinal | 0.17 ± 0.13 |
| Frontal | lparsorbitalis | 0.16 ± 0.08 |
| Temporal | ltransversetemporal | 0.16 ± 0.06 |
| Cingulate | listhmuscingulate | 0.16 ± 0.12 |
| Frontal | rlateralorbitofrontal | 0.15 ± 0.08 |
| Cingulate | risthmuscingulate | 0.14 ± 0.07 |
| Frontal | rparsorbitalis | 0.13 ± 0.16 |
| Temporal | linferiortemporal | 0.13 ± 0.06 |
| | rsuperiortemporal | 0.12 ± 0.05 |
| Insula | rinsula | 0.12 ± 0.12 |
| Temporal | rinferiortemporal | 0.11 ± 0.06 |
| Frontal | llateralorbitofrontal | 0.11 ± 0.06 |
| Temporal | rbankssts | 0.11 ± 0.10 |
| Frontal | lmedialorbitofrontal | 0.10 ± 0.08 |
| Temporal | lfusiform | 0.10 ± 0.06 |
| | rparahippocampal | 0.10 ± 0.11 |
| Paracentral | rparacentral | 0.10 ± 0.10 |
| Temporal | ltemporalpole | 0.10 ± 0.09 |
| Occipital | rlingual | 0.09 ± 0.07 |
| Frontal | lparsopercularis | 0.09 ± 0.05 |
| Temporal | lsuperiortemporal | 0.09 ± 0.05 |
| | rmiddletemporal | 0.09 ± 0.04 |
| Cingulate | lrostralanteriorcingulate | 0.08 ± 0.07 |
| Pre-Postcentral | lprecentral | 0.07 ± 0.05 |
| Parietal | linferiorparietal | 0.07 ± 0.02 |
| Frontal | lcaudalmiddlefrontal | 0.07 ± 0.07 |
| Temporal | rtransversetemporal | 0.07 ± 0.10 |
| Pre-Postcentral | rpostcentral | 0.07 ± 0.09 |
| Frontal | lsuperiorfrontal | 0.06 ± 0.07 |
| Occipital | rlateraloccipital | 0.06 ± 0.06 |
| Parietal | lsupramarginal | 0.06 ± 0.03 |
| Frontal | lfrontalpole | 0.06 ± 0.14 |
| | rmedialorbitofrontal | 0.06 ± 0.08 |
| Parietal | rinferiorparietal | 0.06 ± 0.06 |
| Temporal | lparahippocampal | 0.06 ± 0.08 |
| Frontal | lparstriangularis | 0.05 ± 0.06 |
| Parietal | lprecuneus | 0.05 ± 0.03 |
| Temporal | lmiddletemporal | 0.05 ± 0.05 |
| Paracentral | lparacentral | 0.05 ± 0.06 |
| Temporal | rfusiform | 0.04 ± 0.04 |
| Occipital | llingual | 0.04 ± 0.04 |
| Cingulate | lposteriorcingulate | 0.03 ± 0.05 |
| Pre-Postcentral | lpostcentral | 0.03 ± 0.05 |
| Frontal | rparsopercularis | 0.03 ± 0.01 |
| Occipital | lcuneus | 0.02 ± 0.05 |
| Parietal | rsupramarginal | 0.02 ± 0.04 |
| Pre-Postcentral | rprecentral | 0.02 ± 0.08 |
| Occipital | lpericalcarine | 0.01 ± 0.06 |
| Frontal | rsuperiorfrontal | 0.01 ± 0.07 |
| Occipital | rpericalcarine | 0.004 ± 0.05 |
| Temporal | lbankssts | 0.003 ± 0.04 |
| Parietal | lsuperiorparietal | -0.001 ± 0.03 |
| Occipital | llateraloccipital | -0.003 ± 0.03 |
| Frontal | rcaudalmiddlefrontal | -0.004 ± 0.05 |
| | rparstriangularis | -0.01 ± 0.04 |
| | lrostralmiddlefrontal | -0.01 ± 0.06 |
| Parietal | rprecuneus | -0.01 ± 0.04 |
| Frontal | rrostralmiddlefrontal | -0.03 ± 0.07 |
| Cingulate | rrostralanteriorcingulate | -0.04 ± 0.08 |
| Parietal | rsuperiorparietal | -0.05 ± 0.07 |
| Cingulate | lcaudalanteriorcingulate | -0.08 ± 0.12 |
| Occipital | rcuneus | -0.09 ± 0.06 |
| Cingulate | rposteriorcingulate | -0.10 ± 0.08 |
| Frontal | rfrontalpole | -0.11 ± 0.07 |
| Cingulate | rcaudalanteriorcingulate | -0.17 ± 0.18 |

**Supplementary Table 5.** This table supplements Tables 11 and 12. It ranks the level of improvement of all the 68 cortical areas surveyed in structural MRI (values are AVG±SE; measurements mm) for the subgroup of the WANR patients that continued as ASIA A (n=6), after 5 months of training. Only seven cortical areas displayed gains ≥ 0.2mm (limited by the black arrow): four areas in the temporal lobe, one in the frontal lobe, one in the cingulate cortex, and the left insula. The insulas are shown in green; cortical areas in the temporal lobe areas are shown in brown; the combined pre and postcentral gyri areas are shown in light red; paracentral areas are shown in dark red; parietal areas are shown in yellow; frontal areas in salmon; cingulate lobe areas are shown in orange and occipital lobe areas are shown in blue. For the designation of each cortical area, the letter "l" depicts the left hemisphere, and "r" the right hemisphere.

**A)**

| LEFT Hemisphere | ASIA A-C (n=5) AVG IMP (mm) ± SE | ASIA A-A (n=6) AVG IMP (mm) ± SE | Abs Hedges' g |
|---|---|---|---|
| Insula | 0.72 ± 0.46 | 0.19 ± 0.09 | 0.69 |
| Cingulate | 0.33 ± 0.17 | 0.05 ± 0.04 | 0.96 |
| Pre-Postcentral | 0.27 ± 0.19 | 0.05 ± 0.05 | 0.65 |
| Frontal Lobe | 0.24 ± 0.21 | 0.08 ± 0.05 | 0.46 |
| Temporal Lobe | 0.24 ± 0.18 | 0.09 ± 0.06 | 0.44 |
| Parietal Lobe | 0.21 ± 0.12 | 0.05 ± 0.02 | 0.84 |
| Paracentral | 0.17 ± 0.13 | 0.05 ± 0.06 | 0.52 |
| Occipital Lobe | 0.09 ± 0.05 | 0.02 ± 0.02 | 0.72 |

**B)**

| RIGHT Hemisphere | ASIA A-C (n=5) AVG IMP (mm) ± SE | ASIA A-A (n=6) AVG IMP (mm) ± SE | Abs Hedges' g |
|---|---|---|---|
| Temporal Lobe | 0.53 ± 0.30 | 0.12 ± 0.09 | 0.79 |
| Frontal Lobe | 0.36 ± 0.30 | 0.02 ± 0.06 | 0.65 |
| Cingulate | 0.32 ± 0.26 | -0.04 ± 0.05 | 0.82 |
| Parietal Lobe | 0.25 ± 0.14 | 0.00 ± 0.03 | 1.06 |
| Pre-Postcentral | 0.25 ± 0.19 | 0.04 ± 0.09 | 0.57 |
| Occipital Lobe | 0.19 ± 0.17 | 0.02 ± 0.03 | 0.60 |
| Paracentral | 0.13 ± 0.09 | 0.10 ± 0.10 | 0.13 |
| Insula | 0.07 ± 0.20 | 0.12 ± 0.12 | 0.11 |

**Supplementary Table 6.** Comparison of improvement in left and right hemispheres for ASIA A-C (n=5) and ASIA A-A (n=6) subgroups after 5 months of training with the WANR protocol. Values (AVG±SE; measurements mm) are ranked by the level of improvement for the ASIA A-C subgroup. Absolute Hedges' g column displays the values for A-C x A-A subgroups. Hedges' g represents how many pooled standard deviations apart two group means are, with a small-sample bias correction. Its magnitude conveys the effect size (~0.2 small, ~0.5 medium, ~0.8 large, ≥1.2 very large). The insulas are shown in green; cortical areas in the temporal lobe areas are shown in brown; the combined pre and postcentral gyri areas are shown in light red; paracentral areas are shown in dark red; parietal areas are shown in yellow; frontal areas in salmon; cingulate lobe areas are shown in orange and occipital lobe areas are shown in blue. For the designation of each cortical area, the letter "l" depicts the left hemisphere, and "r" the right hemisphere.

A)

| LEFT Hemisphere | ASIA A-C (n=4) AVG IMP (mm) ± SE | ASIA A-A (n=6) AVG IMP (mm) ± SE | Abs Hedges' g |
|---|---|---|---|
| Insula | 0.97 ± 0.54 | 0.08 ± 0.08 | 1.17 |
| Pre-Postcentral | 0.44 ± 0.19 | -0.04 ± 0.06 | 1.65 |
| Temporal Lobe | 0.42 ± 0.21 | -0.06 ± 0.05 | 1.52 |
| Cingulate | 0.40 ± 0.24 | 0.02 ± 0.05 | 1.11 |
| Frontal Lobe | 0.39 ± 0.23 | -0.06 ± 0.05 | 1.35 |
| Parietal Lobe | 0.29 ± 0.14 | -0.05 ± 0.05 | 1.50 |
| Paracentral | 0.23 ± 0.10 | -0.01 ± 0.06 | 1.34 |
| Occipital Lobe | 0.08 ± 0.05 | -0.02 ± 0.02 | 1.16 |

B)

| RIGHT Hemisphere | ASIA A-C (n=4) AVG IMP (mm) ± SE | ASIA A-A (n=6) AVG IMP (mm) ± SE | Abs Hedges' g |
|---|---|---|---|
| Temporal Lobe | 0.64 ± 0.34 | -0.07 ± 0.03 | 1.53 |
| Frontal Lobe | 0.54 ± 0.33 | -0.07 ± 0.04 | 1.33 |
| Cingulate | 0.39 ± 0.27 | -0.04 ± 0.06 | 1.13 |
| Pre-Postcentral | 0.38 ± 0.20 | -0.02 ± 0.07 | 1.31 |
| Insula | 0.34 ± 0.19 | -0.02 ± 0.12 | 0.97 |
| Parietal Lobe | 0.33 ± 0.15 | -0.01 ± 0.05 | 1.46 |
| Occipital Lobe | 0.20 ± 0.24 | -0.06 ± 0.04 | 0.79 |
| Paracentral | 0.19 ± 0.04 | -0.01 ± 0.08 | 1.05 |

**Supplementary Table 7.** Comparison of improvement in left and right hemispheres for ASIA A-C (n=4) and ASIA A-A (n=6) subgroups after 9 months of training with the WANR protocol. Values (AVG±SE; measurements mm) are ranked by the level of improvement for the ASIA A-C subgroup. Absolute Hedges' g column displays the values for A-C x A-A subgroups. Hedges' g represents how many pooled standard deviations apart two group means are, with a small-sample bias correction. Its magnitude conveys the effect size (~0.2 small, ~0.5 medium, ~0.8 large, ≥1.2 very large). The insulas are shown in green; cortical areas in the temporal lobe areas are shown in brown; the combined pre and postcentral gyri areas are shown in light red; paracentral areas are shown in dark red; parietal areas are shown in yellow; frontal areas in salmon; cingulate lobe areas are shown in orange and occipital lobe areas are shown in blue. For the designation of each cortical area, the letter "l" depicts the left hemisphere, and "r" the right hemisphere.

| | Cortical Areas | ASIA A-C (n=4)<br>AVG IMP (mm) ± SE |
|---|---|---|
| Temporal | rtemporalpole | 1.04 ± 0.65 |
| Insula | linsula | 0.97 ± 0.54 |
| Cingulate | rrostralanteriorcingulate | 0.92 ± 0.54 |
| Temporal | lentorhinal | 0.91 ± 0.62 |
| | rsuperiortemporal | 0.77 ± 0.54 |
| Cingulate | lrostralanteriorcingulate | 0.76 ± 0.54 |
| Frontal | rsuperiorfrontal | 0.75 ± 0.50 |
| Temporal | rmiddletemporal | 0.75 ± 0.59 |
| | rfusiform | 0.75 ± 0.53 |
| | rparahippocampal | 0.74 ± 0.37 |
| Parietal | rinferiorparietal | 0.74 ± 0.47 |
| Frontal | llateralorbitofrontal | 0.74 ± 0.51 |
| Frontal | lparsopercularis | 0.70 ± 0.51 |
| Temporal | lfusiform | 0.69 ± 0.47 |
| Frontal | rlateralorbitofrontal | 0.68 ± 0.54 |
| | rparsopercularis | 0.68 ± 0.55 |
| | lparsorbitalis | 0.68 ± 0.56 |
| Temporal | rtransversetemporal | 0.68 ± 0.45 |
| Parietal | linferiorparietal | 0.67 ± 0.47 |
| Frontal | rrostralmiddlefrontal | 0.67 ± 0.45 |
| Temporal | lmiddletemporal | 0.67 ± 0.57 |
| Frontal | rparstriangularis | 0.66 ± 0.55 |
| | rmedialorbitofrontal | 0.65 ± 0.55 |
| Pre-Postcentral | rprecentral | 0.64 ± 0.40 |
| | lprecentral | 0.63 ± 0.39 |
| Cingulate | rcaudalanteriorcingulate | 0.61 ± 0.50 |
| Frontal | lfrontalpole | 0.57 ± 0.63 |
| Temporal | ltemporalpole | 0.53 ± 0.24 |
| | rentorhinal | 0.53 ± 0.24 |
| Cingulate | listhmuscingulate | 0.52 ± 0.39 |
| Occipital | rlingual | 0.44 ± 0.43 |
| | rcuneus | 0.36 ± 0.39 |
| Insula | rinsula | 0.34 ± 0.19 |
| Parietal | rsupramarginal | 0.34 ± 0.24 |
| Frontal | rcaudalmiddlefrontal | 0.31 ± 0.15 |
| Temporal | lsuperiortemporal | 0.31 ± 0.23 |
| Frontal | rparsorbitalis | 0.30 ± 0.11 |
| | lcaudalmiddlefrontal | 0.28 ± 0.19 |
| Temporal | lbankssts | 0.28 ± 0.30 |
| | rbankssts | 0.27 ± 0.27 |
| Pre-Postcentral | lpostcentral | 0.24 ± 0.11 |
| Cingulate | lposteriorcingulate | 0.24 ± 0.14 |
| Paracentral | lparacentral | 0.23 ± 0.10 |
| Parietal | lsupramarginal | 0.21 ± 0.16 |
| Temporal | lparahippocampal | 0.19 ± 0.03 |
| Parietal | lprecuneus | 0.19 ± 0.08 |
| Temporal | rinferiortemporal | 0.19 ± 0.13 |
| Frontal | lsuperiorfrontal | 0.19 ± 0.18 |
| Paracentral | rparacentral | 0.19 ± 0.04 |
| Frontal | lparstriangularis | 0.18 ± 0.15 |
| | rfrontalpole | 0.17 ± 0.19 |
| Parietal | rprecuneus | 0.15 ± 0.08 |
| Temporal | linferiortemporal | 0.15 ± 0.15 |
| Occipital | llateraloccipital | 0.15 ± 0.13 |
| Frontal | lrostralmiddlefrontal | 0.14 ± 0.12 |
| Pre-Postcentral | rpostcentral | 0.12 ± 0.03 |
| Parietal | rsuperiorparietal | 0.10 ± 0.02 |
| Cingulate | lcaudalanteriorcingulate | 0.09 ± 0.15 |
| Cingulate | rposteriorcingulate | 0.08 ± 0.06 |
| Occipital | llingual | 0.08 ± 0.06 |
| Parietal | lsuperiorparietal | 0.08 ± 0.05 |
| Occipital | lcuneus | 0.08 ± 0.03 |
| Frontal | lmedialorbitofrontal | 0.07 ± 0.07 |
| Occipital | rlateraloccipital | 0.05 ± 0.06 |
| Temporal | ltransversetemporal | 0.04 ± 0.05 |
| Occipital | lpericalcarine | 0.03 ± 0.05 |
| Occipital | rpericalcarine | -0.03 ± 0.10 |
| Cingulate | risthmuscingulate | -0.05 ± 0.11 |

**Supplementary Table 8.** Rank of the 68 cortical areas surveyed in structural MRI (values are AVG±SE; measurements mm) for the subgroup of the WANR patients that made a transition from ASIA A to ASIA C (n=4), after 9 months of training. A total of 51 cortical areas (75% of the 68 cortical areas analyzed) have values ≥ 0.2mm (limited by the black arrow). The insulas are shown in green; cortical areas in the temporal lobe areas are shown in brown; the combined pre and postcentral gyri areas are shown in light red; paracentral areas are shown in dark red; parietal areas are shown in yellow; frontal areas in salmon; cingulate lobe areas are shown in orange and occipital lobe areas are shown in blue. For the designation of each cortical area, the letter "l" depicts the left hemisphere, and "r" the right hemisphere.

| | Cortical Areas | ASIA A-A(n=6) AVG IMP (mm) ± SE |
|---|---|---|
| Insula | linsula | 0.08 ± 0.08 |
| Occipital | lpericalcarine | 0.08 ± 0.06 |
| Frontal | rparsorbitalis | 0.06 ± 0.06 |
| Cingulate | lrostralanteriorcingulate | 0.05 ± 0.06 |
| Temporal | ltransversetemporal | 0.03 ± 0.04 |
| Cingulate | lposteriorcingulate | 0.02 ± 0.09 |
| Temporal | lfusiform | 0.01 ± 0.09 |
| Parietal | rinferiorparietal | 0.01 ± 0.04 |
| Frontal | lsuperiorfrontal | 0.01 ± 0.07 |
| Cingulate | risthmuscingulate | 0.01 ± 0.06 |
| Temporal | rtransversetemporal | 0.01 ± 0.09 |
| Frontal | lparsopercularis | 0.01 ± 0.03 |
| Cingulate | lcaudalanteriorcingulate | 0.01 ± 0.06 |
| Temporal | rbankssts | 0.002 ± 0.10 |
| Parietal | rprecuneus | 0.0005 ± 0.06 |
| Cingulate | rcaudalanteriorcingulate | -0.004 ± 0.07 |
| Occipital | rlingual | -0.01 ± 0.03 |
| Cingulate | rposteriorcingulate | -0.01 ± 0.10 |
| Paracentral | rparacentral | -0.01 ± 0.08 |
| Pre-Postcentral | rpostcentral | -0.01 ± 0.07 |
| | lpostcentral | -0.01 ± 0.08 |
| Cingulate | listhmuscingulate | -0.01 ± 0.03 |
| Temporal | lparahippocampal | -0.01 ± 0.10 |
| Paracentral | lparacentral | -0.01 ± 0.06 |
| Insula | rinsula | -0.02 ± 0.12 |
| Frontal | lcaudalmiddlefrontal | -0.02 ± 0.07 |
| | rparsopercularis | -0.02 ± 0.04 |
| Parietal | rsuperiorparietal | -0.02 ± 0.07 |
| Occipital | llingual | -0.03 ± 0.05 |
| Temporal | lsuperiortemporal | -0.03 ± 0.03 |
| Occipital | rlateraloccipital | -0.03 ± 0.06 |
| Parietal | rsupramarginal | -0.03 ± 0.05 |
| Temporal | rparahippocampal | -0.03 ± 0.06 |
| Parietal | lsupramarginal | -0.03 ± 0.05 |
| Frontal | rrostralmiddlefrontal | -0.04 ± 0.08 |
| Temporal | rmiddletemporal | -0.04 ± 0.03 |
| Frontal | lrostralmiddlefrontal | -0.04 ± 0.09 |
| | lmedialorbitofrontal | -0.04 ± 0.06 |
| Pre-Postcentral | rprecentral | -0.04 ± 0.06 |
| Temporal | rfusiform | -0.04 ± 0.03 |
| Frontal | rlateralorbitofrontal | -0.04 ± 0.08 |
| | lparsorbitalis | -0.04 ± 0.08 |
| Parietal | linferiorparietal | -0.04 ± 0.05 |
| Frontal | rsuperiorfrontal | -0.04 ± 0.06 |
| Temporal | rsuperiortemporal | -0.05 ± 0.05 |
| | linferiortemporal | -0.05 ± 0.05 |
| Parietal | lprecuneus | -0.06 ± 0.06 |
| | lsuperiorparietal | -0.06 ± 0.08 |
| Frontal | rcaudalmiddlefrontal | -0.07 ± 0.05 |
| Temporal | lmiddletemporal | -0.07 ± 0.04 |
| Occipital | lcuneus | -0.07 ± 0.06 |
| Pre-Postcentral | lprecentral | -0.07 ± 0.05 |
| Occipital | llateraloccipital | -0.07 ± 0.04 |
| Frontal | rparstriangularis | -0.07 ± 0.04 |
| Temporal | lbankssts | -0.08 ± 0.06 |
| Occipital | rcuneus | -0.09 ± 0.05 |
| Frontal | lparstriangularis | -0.10 ± 0.03 |
| Temporal | rinferiortemporal | -0.10 ± 0.08 |
| Frontal | llateralorbitofrontal | -0.10 ± 0.07 |
| Occipital | rpericalcarine | -0.11 ± 0.05 |
| Frontal | rmedialorbitofrontal | -0.15 ± 0.07 |
| Cingulate | rrostralanteriorcingulate | -0.16 ± 0.03 |
| Temporal | ltemporalpole | -0.16 ± 0.11 |
| | lentorhinal | -0.18 ± 0.13 |
| Frontal | lfrontalpole | -0.20 ± 0.10 |
| Temporal | rtemporalpole | -0.20 ± 0.10 |
| | rentorhinal | -0.21 ± 0.09 |
| Frontal | rfrontalpole | -0.30 ± 0.10 |

**Supplementary Table 9.** Rank of the 68 cortical areas surveyed in structural MRI (values are AVG±SE; measurements mm) for the subgroup of the WANR patients that stayed in their original ASIA A status (n=6), after 9 months of training. By 9 months, no cortical area reached the threshold of 0.2mm. The insulas are shown in green; cortical areas in the temporal lobe areas are shown in brown; the combined pre and postcentral gyri areas are shown in light red; paracentral areas are shown in dark red; parietal areas are shown in yellow; frontal areas in salmon; cingulate lobe areas are shown in orange and occipital lobe areas are shown in blue. For the designation of each cortical area, the letter "l" depicts the left hemisphere, and "r" the right hemisphere.
